# Genomic Divergence Between Matched Primary and Metastatic Tumors Across Cancer Types: A Pan-Cancer Analysis of 5,692 Samples

**DOI:** 10.1101/2025.08.22.25334215

**Authors:** Yakup Ergun

## Abstract

**Introduction:** Metastasis represents the leading cause of cancer-related mortality and is characterized by complex biological processes such as genomic instability, immune evasion, and therapy resistance. While metastatic tumors often retain the truncal drivers of their primary counterparts, the extent and nature of additional somatic alterations acquired during progression remain incompletely defined across cancer types.

**Methods:** A comprehensive pan-cancer analysis was conducted using targeted sequencing data from 2,846 patients with matched primary and metastatic tumors (totaling 5,692 samples) obtained from the AACR Project GENIE v18.0 cohort. Harmonized variant calls were used to compare mutation burden, fraction of genome altered (FGA), gene-level mutation frequencies, copy number alterations (CNA), and structural variants (SV) between compartments. Statistical comparisons were adjusted for multiple testing using the Benjamini-Hochberg method.

**Results:** Metastatic tumors exhibited a significantly higher median mutation count (6 vs. 5; p < 0.001) and FGA (0.186 vs. 0.140; p < 0.001) compared to matched primary tumors. This increase was most prominent in non-small cell lung, breast, colorectal, pancreatic, and prostate cancers. Eleven genes, including KDM5A, CDKN2A, MYC, ESR1, and AR, were significantly enriched in metastases, suggesting mechanisms such as cell cycle deregulation, therapy-induced selection, and chromatin remodeling. Notably, ESR1 alterations were enriched in breast cancer metastases, consistent with endocrine therapy resistance, while AR alterations were markedly more frequent in metastatic prostate cancer. CNA analysis revealed recurrent amplifications (MYC, ERBB2, CCND1) and deletions (CDKN2A, PTEN, RB1) in metastatic tumors. Structural variants involving genes linked to DNA damage response and epigenetic regulation were also more prevalent in the metastatic setting.

**Conclusions:** In this large-scale matched cohort of 2,846 patients and 5,692 tumor samples, metastatic tumors exhibited increased mutation burden and widespread genomic instability. Although treatment data were not available to directly associate resistance-related alterations with specific therapies, the observed patterns suggest that these acquired changes reflect context-dependent selection for survival and proliferative advantage in advanced disease, rather than the emergence of novel metastasis-specific driver events.

## Introduction

Metastasis accounts for the majority of cancer-related mortality and represents the least tractable phase of disease biology, where dissemination, colonization, immune evasion, and therapy resistance converge within a dynamic evolutionary process [1].

Extensive genomic profiling has demonstrated that metastatic tumors generally preserve the truncal drivers of their primaries, while simultaneously acquiring additional alterations that confer fitness advantages in distant microenvironments. Large-scale whole-genome analyses of metastatic tumors have revealed high rates of whole-genome duplication, frequent biallelic inactivation of tumor-suppressor genes, and a predominance of clonal driver mutations. Collectively, these features suggest strong selective pressures [2]. By contrast, pan-cancer resources anchored in primary disease indicate that catastrophic structural events, including chromothripsis, breakage–fusion–bridge cycles, extrachromosomal DNA amplification, and copy-number–driven evolutionary trajectories, are frequent and often arise early in tumorigenesis, foreshadowing the genomic complexity observed at metastatic presentation [3,4].

Efforts to delineate the divergence between primary and metastatic tumors have primarily focused on linking these differences to underlying biological mechanisms and clinical consequences. Evidence indicates that the timing of systemic spread varies across cancer lineages and patients, ranging from early dissemination with parallel evolution to late metastatic spread after extensive primary tumor evolution [5–7]. Structural archetypes, including whole-genome duplication, complex amplification signatures, and chromothripsis, are consistently associated with aggressive biological behavior and adverse outcomes, emphasizing genome-scale instability as a unifying axis of metastatic fitness [4]. Furthermore, immune evasion is not confined to end-stage disease. Uniformly processed whole-genome sequencing cohorts across thousands of tumors have demonstrated that a considerable proportion of patients harbor genetically encoded immune-evasion events—most commonly HLA class I loss of heterozygosity—with broadly similar rates in both primary and metastatic settings, suggesting early acquisition of immune escape mechanisms [8]. The prevalence of such events scales with mutational burden in a non-linear manner (for example, focal HLA-I LOH peaking at approximately 10–20 mutations/Mb), underscoring that the immunogenic impact of mutations is influenced by their clonality and functional relevance in addition to their quantity [8].

These findings hold important implications for biomarker development and clinical practice. Tumor mutational burden (TMB) correlates with response to immune checkpoint inhibitors in select contexts and supports agnostic indications (for example, ≥10 mutations/Mb). However, its predictive value varies substantially across cancer types and cohorts, and may be confounded by spatiotemporal heterogeneity and prior therapeutic exposures [9–11,14]. Distinct mutational signatures attributable to specific treatments, such as platinum or temozolomide, can inflate TMB estimates without corresponding increases in immunogenic neoepitopes, thereby complicating interpretation at clinically applied thresholds [12,14]. Beyond immunotherapy, metastatic progression is frequently accompanied by the selective enrichment of actionable and resistance-associated alterations, including ESR1 in endocrine-treated breast cancer, AR in prostate cancer, and EGFR T790M in patients with EGFR TKI– treated non–small cell lung cancer, highlighting the importance of context-aware interpretation of variant function and clonality [2,13].

Although targeted sequencing has enabled precision oncology at scale, variability in assay design and analytical pipelines has historically limited the ability to make cross-study comparisons and to capture the full spectrum of genomic alterations, including structural variants and copy-number–associated scars. Recent harmonized analyses integrating primary and metastatic whole-genome sequencing now provide an opportunity for unbiased comparisons across compartments and cancer lineages [15].

Within this framework, a pan-cancer analysis of paired primary and metastatic tumor samples was conducted using comprehensive high-depth targeted sequencing data. The objective was to evaluate whether consistent and lineage-dependent differences in mutation burden are observed between primary and metastatic tumors and to identify gene-level alterations that demonstrate clinically meaningful shifts in frequency across compartments.

## Materials and Methods

### Data Source and Cohort Assembly

De-identified clinicogenomic data were obtained from the AACR Project GENIE consortium (version 18.0-public) through cBioPortal. This release comprises 211,526 patients and 250,018 tumor samples contributed by multiple international cancer centers, following standardized de-identification, harmonization, and quality control. Clinical annotations (including age at sequencing, sex, race/ethnicity, cancer type, and sample source) and harmonized somatic variant calls were used as provided.

For intra-patient primary–metastasis comparisons, only patients with exactly two sequenced tumor samples were included. Pairs were retained if one sample originated from a primary tumor and the other from a metastasis. Cases with missing or discordant site annotations or incomplete clinical data were excluded. This process yielded a matched cohort of 2,846 patients, each contributing one primary and one metastatic tumor sample (5,692 samples in total). For cancer type–specific and subtype analyses, rare histologies with insufficient representation were excluded to ensure stability of estimates; consequently, denominators at the subtype level did not always sum to the overall cohort total.

### Sequencing Assays

Samples were profiled using institution-specific targeted next-generation sequencing (NGS) assays recorded in the Sequence_Assay_ID metadata. The most frequent platforms were MSK-IMPACT (multiple panel versions, e.g., 341/410/468/505) and the DFCI OncoPanel (v2/v3/v3.1), with additional contributions from other institutional panels. No re-alignment or re-calling was performed. Analyses relied on harmonized variant calls distributed by GENIE to preserve cross-institution comparability despite assay heterogeneity.

### Variable Definitions and Data Extraction

Baseline variables (age at sequencing, sex, race/ethnicity, cancer type, and sample source) were obtained from the GENIE clinical file.

Mutation count, used as the mutation burden metric, was defined as the total number of non-synonymous somatic SNVs/indels per sample. Synonymous variants and known germline polymorphisms were excluded. Descriptive statistics (minimum, maximum, mean, standard deviation [SD], median, interquartile range [IQR], and mean absolute deviation [MAD]) were calculated separately for primary and metastatic tumors.

### Post-Sequencing Survival Derivation

Because overall survival timestamps were not uniformly available, survival was derived from date-of-birth (DOB)–anchored intervals. Age at death was calculated as (DOB to date-of-death interval)/365. For censored cases, age at last contact was calculated as (DOB to last-contact interval)/365. Age at sequencing was extracted directly from the clinical file. Post-sequencing survival was defined as the difference between age at death (or last contact) and age at sequencing. Survival times were expressed in months for reporting. Distributions were summarized separately for patients whose primary versus metastatic samples served as the index.

### Mutation Burden Analysis

Mutation counts from paired primary and metastatic tumors were compared in the overall cohort and across the 15 most represented cancer types. For each setting, mean±SD and median (Q1–Q3) were reported. Because counts were right-skewed and paired in structure, comparisons were made using two-sided Wilcoxon signed-rank tests. False discovery rate (FDR) correction was applied across cancer-type strata using the Benjamini–Hochberg procedure. Raw p-values and adjusted q-values were reported, with q < 0.05 considered statistically significant.

### Fraction of Genome Altered (FGA) Analysis

The fraction of genome altered (FGA) was defined as the proportion of the genome affected by copy number alterations (CNA), using values provided in the clinical metadata. FGA was analyzed as a continuous variable. Paired comparisons between primary and metastatic tumors were performed using Wilcoxon signed-rank tests, with summary statistics (mean±SD, median, range) reported for each group.

### Copy Number Alteration (CNA) Analysis

Gene-level CNA data were extracted from the discrete CNA matrix, coded as: −2 (deep deletion), −1 (shallow deletion), 0 (neutral), 1 (low-level gain), or 2 (amplification). For this analysis, only deep deletions (−2) and high-level amplifications (2) were considered. Frequencies were compared between primary and metastatic tumors using Fisher’s exact tests, with FDR-adjusted q-values.

### Structural Variant (SV) Analysis

Structural variants (SVs) were obtained from the harmonized annotation file. Binary indicators (present/absent) were generated per gene and sample. Only genes with at least five SV-positive cases in either group were analyzed. Comparisons were performed using Fisher’s exact tests with FDR adjustment. Genes with q < 0.05 were reported. Results were summarized pan-cancer and supplemented with subtype-level analyses.

### Gene-Level Alteration Frequency Analysis

Differences in gene-level mutation frequencies were assessed both pan-cancer and within cancer types. Genes with fewer than five events in either group were excluded. Fisher’s exact tests were used, with FDR correction. Genes with q < 0.05 were considered significantly differentially mutated.

### Statistical Software and Reporting

Analyses were conducted using R (v4.3.x) and Python (v3.11). Descriptive statistics included mean±SD and median (Q1–Q3) for continuous variables, and counts (%) for categorical variables. Comparative tests included Wilcoxon signed-rank tests for paired continuous variables and Fisher’s exact tests for categorical comparisons. Multiple testing was controlled using the Benjamini–Hochberg method, with both p-values and q-values reported. Statistical significance was defined as q < 0.05 unless otherwise specified.

## Results

### Patient Cohort and Baseline Characteristics

A total of 2,846 patients with 5,692 matched primary and metastatic tumor samples were included. The median age at sequencing was 63 years (interquartile range [IQR], 53–71), and 53.3% of patients were female. The majority self-identified as White (77.2%) and non-Hispanic/non-Latino (86.6%). The most common tumor types were non-small cell lung cancer (NSCLC, 24.7%), breast cancer (14.5%), and colorectal cancer (10.9%) (Table 1).

**Table 1.** Baseline Patient Characteristics.

| Characteristic | N | Percent | Median (Q1–Q3) |
| --- | --- | --- | --- |
| All patients | 2846 | 100% |  |
| Age at sequencing (years) |  |  | 63 (53-71) |
| Sex |  |  |  |
| Female | 1517 | 53.3% |  |
| Male | 1262 | 44.3% |  |
| Ethnicity |  |  |  |
| Non-Spanish/non-Hispanic | 2466 | 86.6% |  |
| Spanish/Hispanic | 146 | 5.1% |  |
| Race |  |  |  |
| White | 2198 | 77.2% |  |
| Asian | 216 | 7.6% |  |
| Black | 170 | 6.0% |  |
| Other | 74 | 2.6% |  |
| Native American | 3 | 0.1% |  |
| Pacific Islander | 1 | 0.0% |  |
| Age at death (years) |  |  | 65 (55-73) |

Metastatic tumors exhibited a significantly higher median mutation count compared with primary tumors (median 6.0 vs. 5.0; mean ± SD, 9.8 ± 17.9 vs. 8.6 ± 17.5; p < 0.001). No significant enrichment was detected for primary tumors. Survival following sequencing was shorter for patients with metastatic biopsies (median 17.1 months) compared with those with primary biopsies (median 27.7 months; p < 0.001), representing an approximate 11-month difference (Table 2).

**Table 2.** Comparison of Genomic Metrics Between Primary and Metastatic Tumors.

| Variable | Primary – Mean ± SD | Metastasis – Mean ± SD | Primary – Median (Range) | Metastasis – Median (Range) | p-value |
| --- | --- | --- | --- | --- | --- |
| Mutation count | 8.6 ± 17.5 | 9.8 ± 17.9 | 5.0 (1.0–510.0) | 6.0 (1.0–433.0) | <0.01 |
| Fraction genome altered | 0.189 ± 0.186 | 0.234 ± 0.215 | 0.140 (0.00–0.97) | 0.186 (0.00–1.00) | <0.01 |

### Genomic Alterations

The most frequently altered genes across the entire cohort were TP53 (57.2% in metastatic vs. 52.7% in primary), KRAS (22.6% vs. 22.0%), PIK3CA (13.1% vs. 13.0%), APC (9.6% vs. 9.2%), and ARID1A (9.6% vs. 9.3%). The 20 most common alterations are displayed in Figure 1, with full gene-level frequencies provided in Table S20.

**Figure 1.**
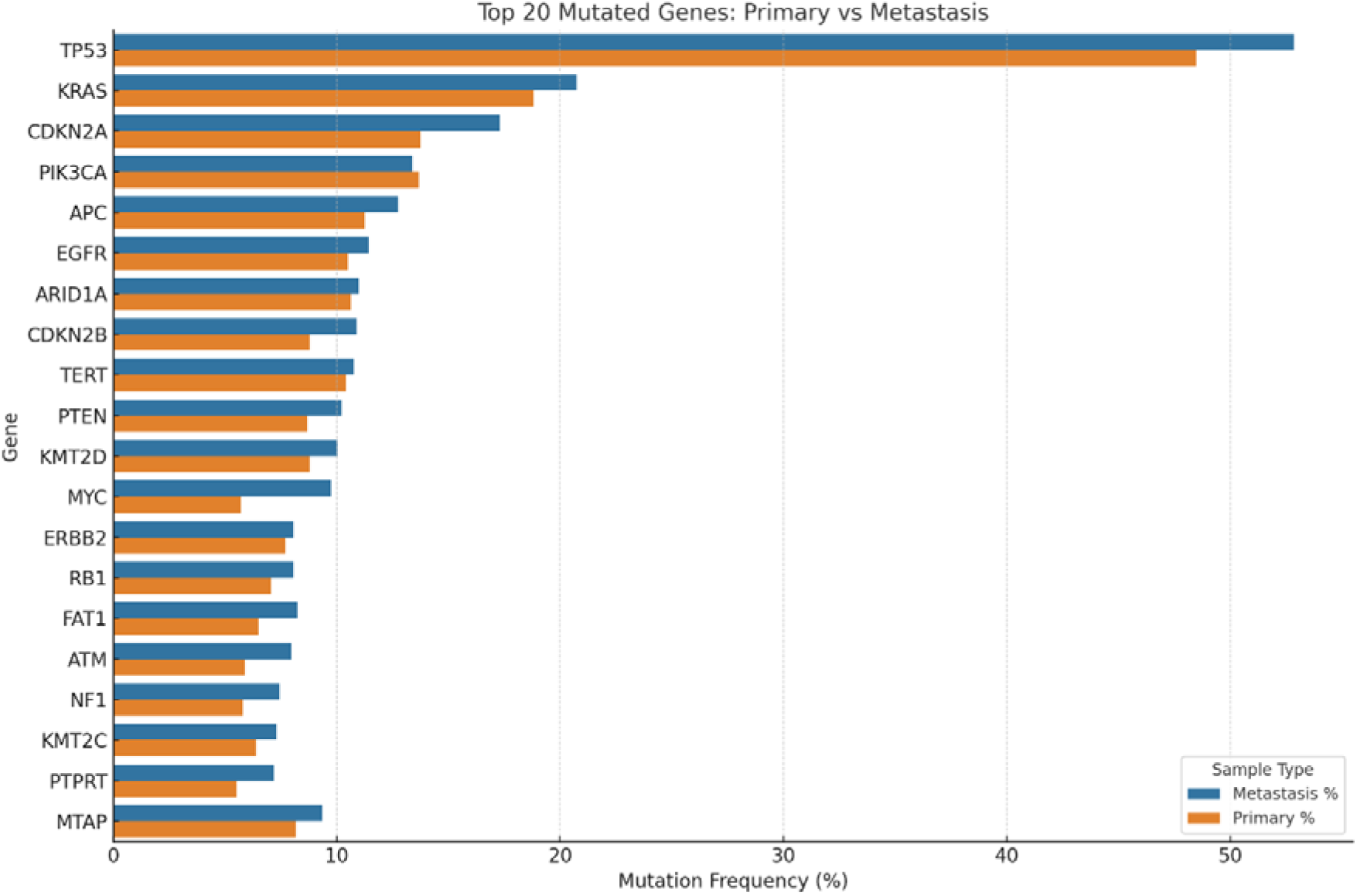
Distribution of the Most Frequently Mutated Genes in Primary and Metastatic Tumors.

Statistical comparison identified 11 genes significantly enriched in metastases (q < 0.05): KDM5A (15.8% vs. 10.4%), CDKN2A (14.7% vs. 10.3%), MYC (9.5% vs. 6.1%), ESR1 (5.2% vs. 3.1%), RAD21 (5.1% vs. 3.2%), AR (4.5% vs. 2.7%), RECQL4 (4.2% vs. 2.4%), ASXL1 (4.2% vs. 2.6%), AGO2 (4.1% vs. 2.5%), BCL2 (3.2% vs. 1.4%), and NFKB2 (2.8% vs. 1.6%) (Figure 2).

**Figure 2.**
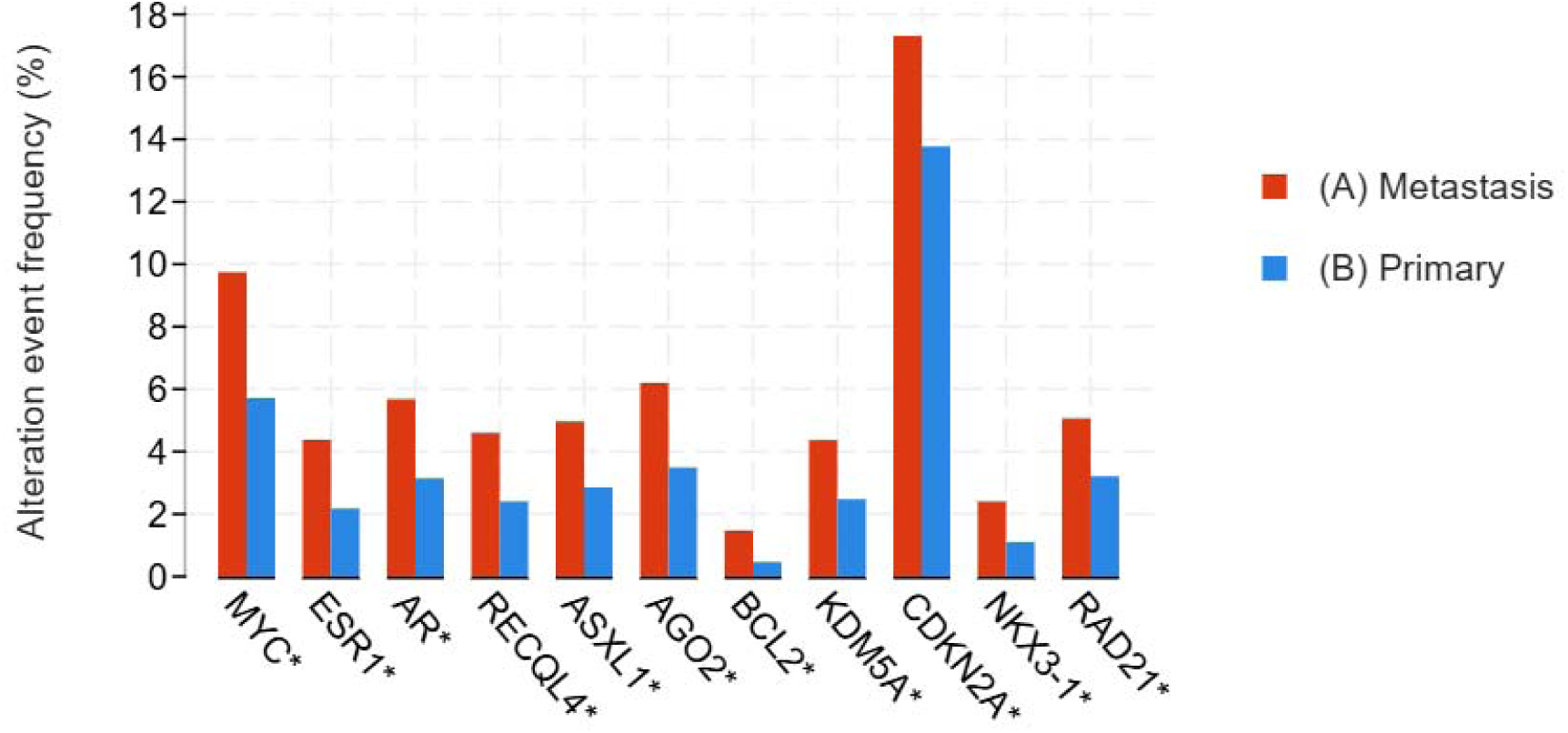
Significantly Altered Genes Between Primary and Metastatic Tumors in the Entire Cohort.

### Fraction of Genome Altered (FGA)

The median FGA was significantly greater in metastatic compared with primary tumors (0.13 vs. 0.09; p < 0.001), consistent across cancer types (Table 2), indicating increased genomic instability in metastatic lesions.

### Copy Number Alterations (CNA)

Metastatic tumors displayed frequent amplifications of MYC, CCND1, ERBB2, FGFR1, and MDM2, as well as deletions involving CDKN2A, PTEN, and RB1. These patterns highlight proliferative signaling advantages and loss of tumor suppressor function as common features of metastasis.

### Structural Variants (SV)

Structural variant analysis revealed several alterations more prevalent in metastases, including PAX8 (42.9% vs. 33.3%), SDC4 (33.3% vs. 25.0%), PTK2 (3.8% vs. 0.7%), and ABCC4 (9.1% vs. 6.7%). Additional enriched events involved IKZF3, EWSR1, STAT6, PRKDC, FOXO1, FOXA1, ATRX, and FGFR1. Many of these genes are implicated in DNA damage response and chromatin regulation, consistent with molecular adaptations favoring metastatic progression.

### Tumor Type Distribution and Subtype-Level Mutation Burden

The cohort comprised 2,846 patients across 15 tumor types (Figure 3). The most frequent malignancies were NSCLC (n = 702, 24.7%), breast cancer (n = 412, 14.5%), and colorectal cancer (n = 311, 10.9%), followed by bladder (n = 261, 9.2%), prostate (n = 223, 7.8%), pancreatic (n = 160, 5.6%), esophagogastric (n = 141, 5.0%), endometrial (n = 113, 4.0%), soft tissue sarcoma (n = 110, 3.9%), ovarian (n = 97, 3.4%), melanoma (n = 74, 2.6%), glioma (n = 65, 2.3%), renal cell carcinoma (n = 63, 2.2%), gastrointestinal stromal tumor (GIST, n = 53, 1.9%), and hepatobiliary tumors (n = 51, 1.8%).

**Figure 3.**
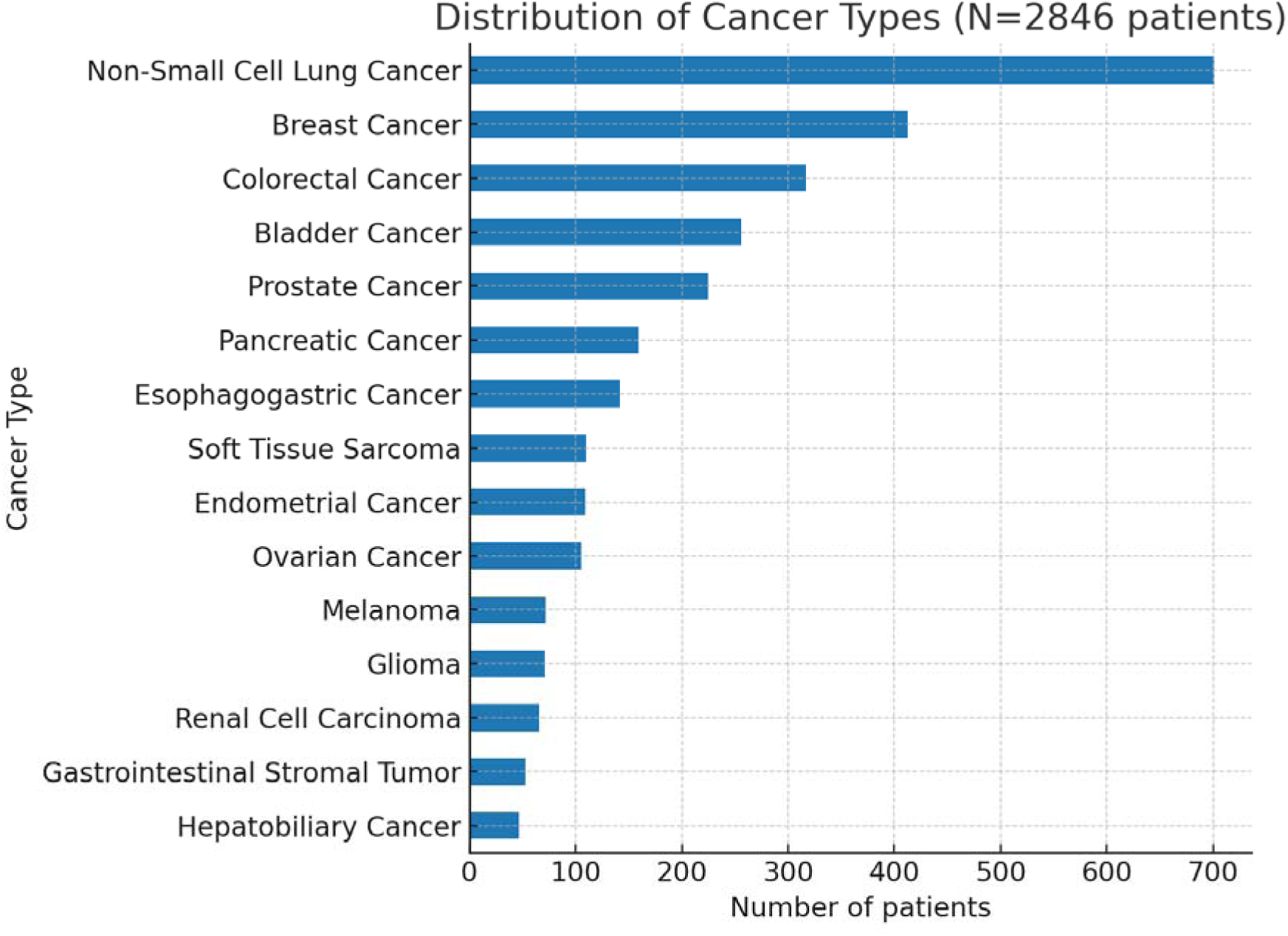
Cancer Type Distribution Across the Study Cohort (N=2846).

After exclusion of rare histologies, subtype-level analyses were conducted in 2,621 patients. Mutation burden was significantly higher in metastases for several tumor types (Table 3, Figure 4). Enrichment was observed in NSCLC (mean 10.5 vs. 8.8; median 8.0 vs. 6.0; q = 0.0023), breast cancer (mean 7.0 vs. 5.1; median 5.0 vs. 4.0; q = 0.00001), colorectal cancer (mean 11.7 vs. 10.9; median 8.0 vs. 7.0; q = 0.0023), pancreatic cancer (mean 6.5 vs. 4.7; median 5.0 vs. 4.0; q = 0.0023), and prostate cancer (mean 6.0 vs. 3.8; median 3.0 in both; q = 0.0114).

**Figure 4.**
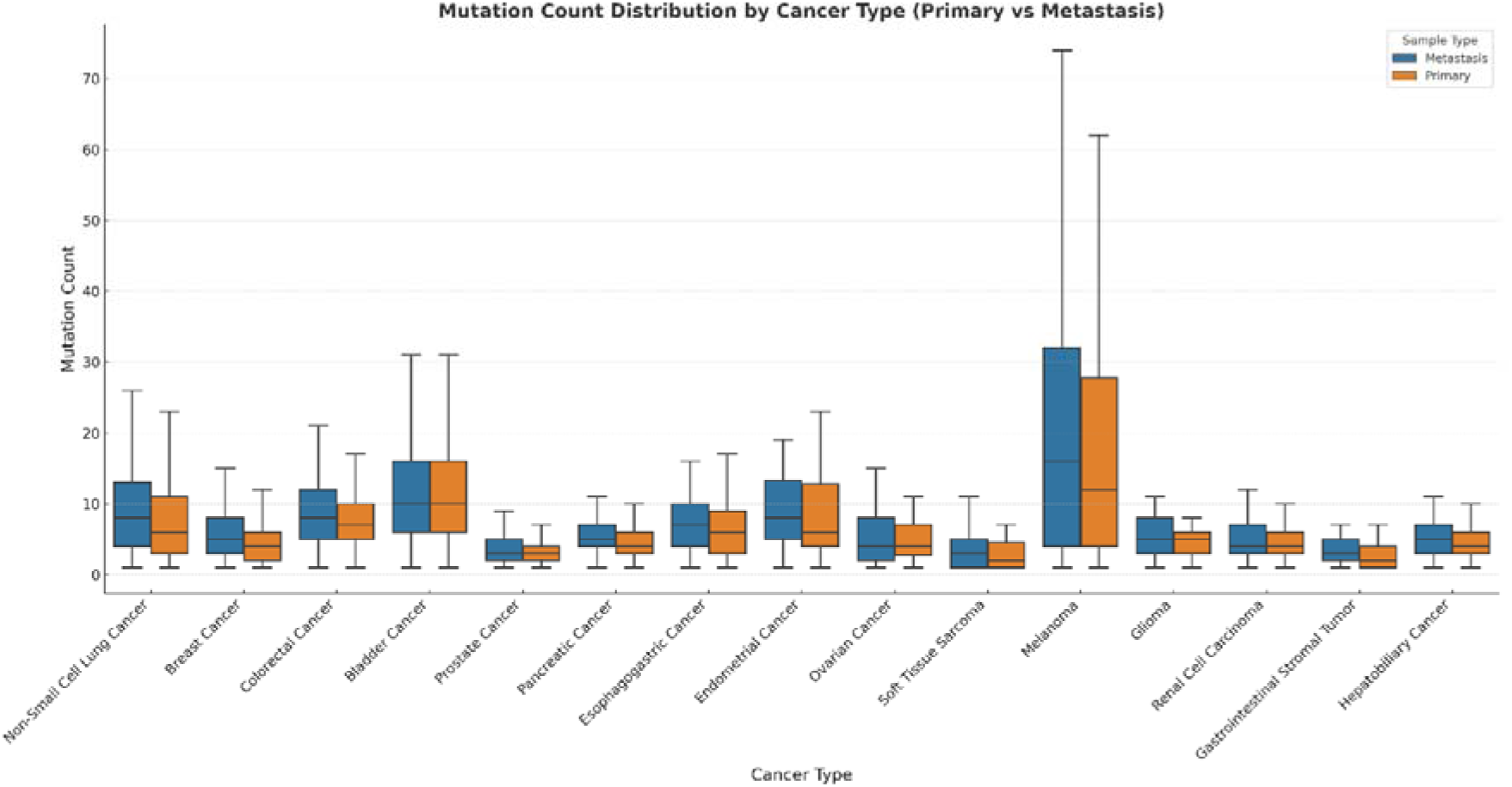
Mutation Count Distribution Across Primary and Metastatic Tumors by Cancer Type.

**Table 3.**
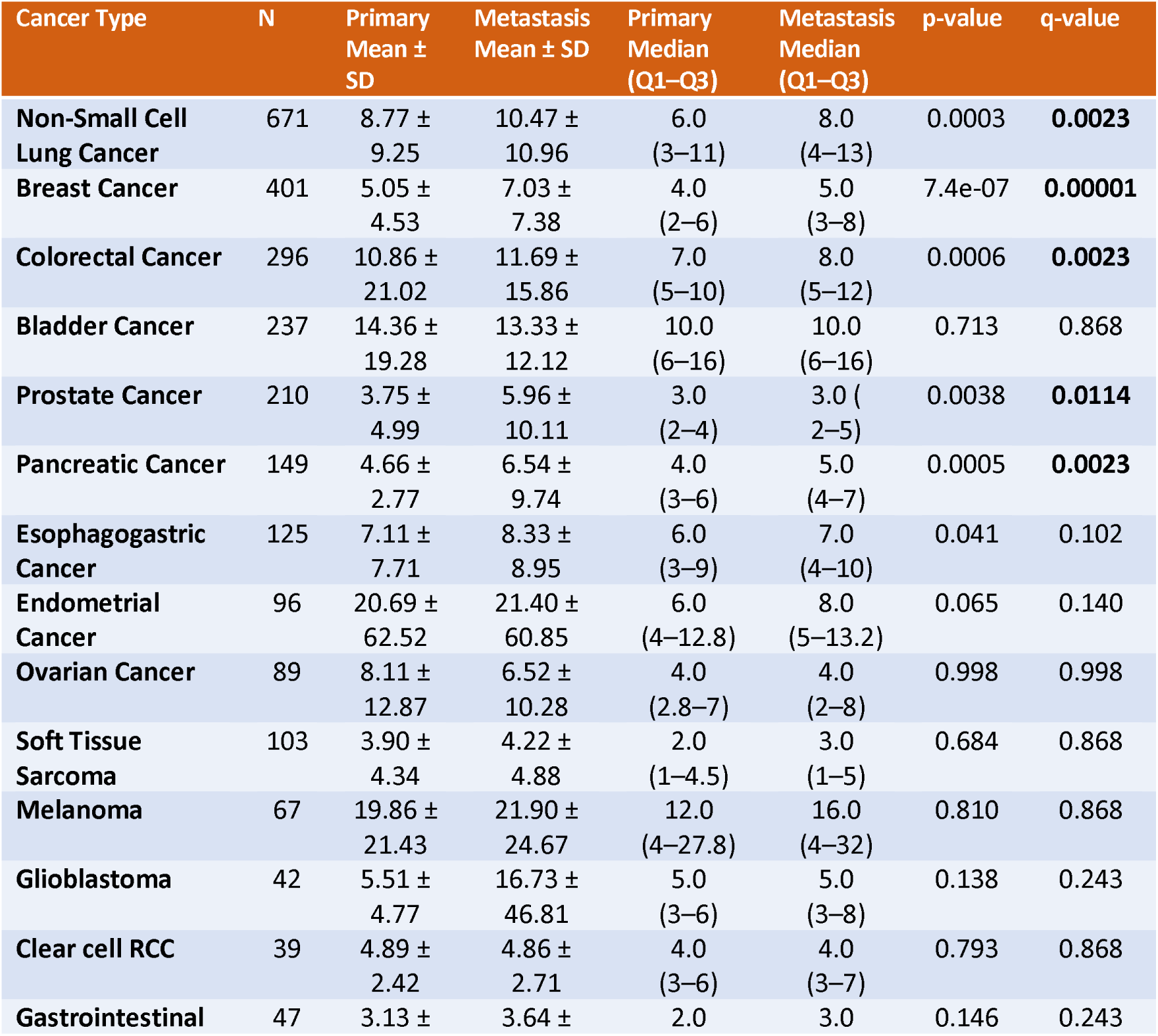
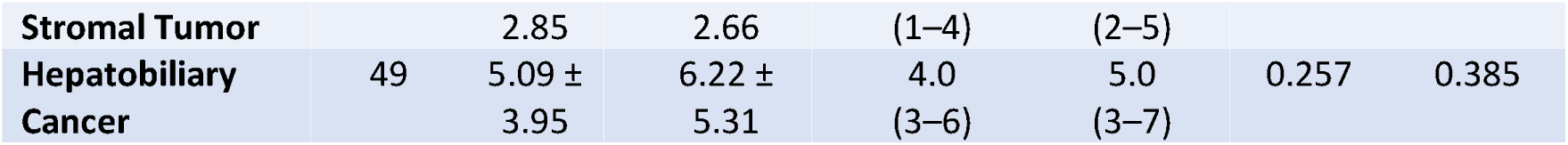
Comparison of Mutation Burden Between Primary and Metastatic Samples Across 15 Cancer Types.

In bladder, ovarian, soft tissue sarcoma, renal cell carcinoma, and melanoma, mutation burden did not differ significantly between compartments (q > 0.86 for all). Melanoma displayed the highest absolute counts in both compartments (mean >19), but large inter-sample variability precluded statistical significance. Endometrial, esophagogastric, and glioma tumors exhibited numerical increases in metastases that did not remain significant after correction (e.g., endometrial: median 6.0 vs. 8.0; q = 0.14). Collectively, these findings indicate that the overall increase in metastatic mutation burden was largely driven by NSCLC, breast, colorectal, pancreatic, and prostate cancers, whereas limited sample sizes may have reduced power in other subtypes.

### Non-Small Cell Lung Cancer

The NSCLC cohort consisted of 671 patients (1,342 samples). The median age at sequencing was 62 years, and 52% were female. Adenocarcinoma was the most common histology. Metastatic samples showed higher mutation burden (mean 10.3 vs. 8.9; median 8 vs. 6; q = 0.0023) and higher FGA (median 0.19 vs. 0.09) (Table S1, Figures S1).

Frequently altered genes included TP53 (54.7% vs. 52.8%), EGFR (32.5% vs. 30.6%), KRAS (26.8% vs. 26.2%), CDKN2A (23.6% vs. 15.0%), and STK11 (15.9% vs. 13.8%). No individual gene reached statistical significance after multiple testing (Table S2, Figures S2). Targetable alterations (ALK, ROS1, RET, HER2, MET, NTRK) were infrequent. The high prevalence of EGFR mutations likely reflects selection bias, as testing may have been performed in clinically enriched populations (e.g., non-smokers, female patients, or those negative for other drivers).

In lung adenocarcinoma (n = 489), metastatic tumors had slightly higher mutation burden (median 8 vs. 7; q = 0.10), but this did not reach significance. Gene-level analyses also showed no significant differences. Thus, the increase observed in the overall NSCLC cohort may result from modest distributed differences rather than a single driver, or from reduced statistical power in histology-specific subsets.

### Breast Cancer

A total of 401 patients (802 samples) were analyzed, of whom 95.3% were female. The median age at sequencing was 55 years (Table S3). Metastatic tumors demonstrated higher mutation counts (median 5 vs. 4; mean 7.06 vs. 5.02; p < 0.01) (Figure S3). TP53, PIK3CA, CCND1, and GATA3 were most frequently altered (Table S4, Figure S4). ESR1 was significantly enriched in metastases, consistent with endocrine resistance biology. Lack of IHC subtype data may explain the predominance of ESR1 alterations, reflecting hormone receptor–positive disease. In 27 patients with lobular carcinoma, no significant difference in mutation burden was observed.

### Colorectal Cancer

A total of 296 patients (592 samples) were included (Table S5). Median age at sequencing was 60 years, and 56.4% were male. Metastatic tumors had higher mutation burden (median 8 vs. 7; mean 10.92 vs. 9.22; p < 0.01) (Figure S5).

Frequent alterations included TP53 (78.4% vs. 74.0%), APC (72.5% vs. 69.6%), KRAS (42.9% vs. 39.5%), SMAD4 (18.7% vs. 15.3%), and PIK3CA (14.9% vs. 14.5%) (Figure S6, Table S6). No genes reached q-value significance, although nominal enrichment was noted for ASXL1 (11.8% vs. 6.3%), AGO2 (11.6% vs. 3.9%), SRC (8.4% vs. 3.6%), and NKX3-1 (7.1% vs. 2.7%). In rectal cancer (n = 54), no significant differences in mutation burden were detected.

### Prostate Cancer

The cohort comprised 210 patients (420 samples). Median age at sequencing was 67 years (IQR 61–74), and all were male. Median survival after sequencing was 2.3 years, with median age at death 73.4 years (Table S7).

Metastatic tumors had nearly double the FGA compared with primaries (0.22 vs. 0.10, IQR 0.09–0.34 vs. 0.02–0.20). Mutation burden was also higher (mean 5.7 vs. 3.74), although the median remained 3 in both groups.

Gene-level analysis (Figure S7, Table S8) revealed frequent alterations in TP53 (48.1%), AR (32.9%), TMPRSS2 (32.9%), PTEN (30.0%), and ERG (28.1%). AR alterations were markedly enriched in metastases (32.9% vs. 3.3%; q < 10¹). PTEN, MYC, RB1, ATM, FANCA, and CDKN1B also appeared more frequent in metastases, though without significance after correction. No gene was enriched in primaries. These results are consistent with AR-driven resistance and genomic instability in advanced prostate cancer.

### Other Cancer Subtypes

Among 11 additional tumor types evaluated—melanoma, endometrial, bladder, ovarian, pancreatic, gastroesophageal, soft tissue sarcoma, clear cell renal cell carcinoma, GIST, biliary tract, and glioblastoma—ten showed no significant gene-level differences after correction. The exception was pancreatic cancer, which demonstrated a significantly higher mutation burden in metastases.

In melanoma (n = 67), alterations in BRAF, TERT, CDKN2A, and NF1 were common and similarly distributed (Figure S8, Table S9). Endometrial cancer (n = 96) showed recurrent alterations in TP53, PTEN, PIK3CA, ARID1A, KRAS, KMT2D, and CTNNB1 without inter-compartment differences (Figure S9, Table S10). In bladder cancer (n = 237), TP53, TERT, ARID1A, KMT2D, KDM6A, CDKN2A, FGFR3, RB1, and PIK3CA were altered at comparable frequencies (Figure S10, Table S11).

Ovarian cancer (n = 89) was characterized by frequent TP53, KRAS, MYC, CCNE1, ARID1A, PIK3CA, BRCA1, and ERBB2 alterations, without significant differences (Figure S11, Table S12). Pancreatic cancer (n = 149) showed a significantly higher metastatic mutation burden (mean 6.79 vs. 4.68; p = 2.51 × 10[[; q = 1.42 × 10[³), although no single gene met q-value significance (Figure S12, Table S13).

Gastroesophageal cancers (n = 125) displayed frequent TP53, CDKN2A, KRAS, ERBB2, and ARID1A mutations, with minor differences in low-frequency genes (CDKN2B, MTAP, GNAS, CIC, ASXL1, CREBBP), none statistically significant (Figure S13, Table S14). Soft tissue sarcomas (n = 103) showed alterations in TP53, CDK4, MDM2, and RB1 without inter-compartment differences (Figure S14, Table S15).

Clear cell renal cell carcinoma (n = 39) demonstrated consistent frequencies of VHL, PBRM1, BAP1, and SETD2 alterations (Figure S15, Table S16). GISTs (n = 47) showed KIT mutations and CDKN2A/B deletions at similar rates (Figure S16, Table S17). Biliary tract cancers (n = 37) exhibited KRAS, TP53, ARID1A, and CDKN2A alterations with overlapping distributions (Figure S17, Table S18). Glioblastoma (n = 42) displayed consistent alteration patterns in TERT, EGFR, CDKN2A/B, PTEN, and TP53, with no significant differences (Figure S18, Table S19).

## Discussion

In this paired, pan-cancer comparison, metastatic tumors had a higher mutation burden (median 6 vs 5) and a larger fraction of the genome affected by copy-number change (FGA; median 0.186 vs 0.140) than primaries, but the size of this difference varied by cancer type rather than being uniform across all lineages. This pattern suggests that mutation burden tends to be higher in metastases across many tumor types; however, in some subtypes the difference does not reach statistical significance because of limited sample size, wide variance, and multiple-testing correction; therefore, interpretation should take tumor type and sampling context into account [2–4].

At the gene level, 11 genes were significantly more frequent in metastases: KDM5A, CDKN2A, MYC, ESR1, RAD21, AR, RECQL4, ASXL1, AGO2, BCL2, and NFKB2. Functionally, these point to established routes by which tumors acquire fitness in advanced disease, namely loss of G1/S checkpoint control (CDKN2A), enhanced oncogenic signaling or dosage (MYC), therapy-conditioned adaptation along hormone pathways (ESR1, AR), changes in chromatin regulation and genome maintenance (KDM5A, ASXL1, RAD21, RECQL4), post-transcriptional regulation (AGO2), and survival advantages via anti-apoptotic or inflammatory signaling (BCL2, NFKB2). The overall pattern is consistent with selection of context-dependent modules rather than the acquisition of entirely new, metastasis-specific drivers, aligning with prior genomic studies of metastatic disease [2,13,16–19].

Copy-number changes similarly favored metastases, with recurrent gains of MYC, CCND1, ERBB2, FGFR1, and MDM2, along with losses of CDKN2A, PTEN, and RB1. These alterations underscore proliferative advantages and loss of tumor-suppressor control as selective pressures in metastatic disease. Although targeted sequencing panels have limited resolution for complex rearrangements, these findings are concordant with whole-genome reports of copy-number signatures, chromothripsis, and extrachromosomal DNA, all of which link genome-scale instability to aggressive biology [2–4,20–22]. The higher FGA in metastases (0.186 vs. 0.140) further highlights this genomic instability. Structural variants, including rearrangements involving PAX8, SDC4, PTK2, and ABCC4, were also more common in metastases. Many of these involve genes connected to DNA-damage response and chromatin regulation, suggesting a mechanistic link between replication stress, altered repair, and metastatic progression, consistent with whole-genome descriptions of structurally unstable tumors [2–4].

Lineage-specific patterns clarify clinical implications. In breast cancer, metastatic tumors had a higher mutation burden (median 5 vs. 4), with ESR1 as the only significantly enriched gene, consistent with its established role in endocrine resistance [16,23–25]. In prostate cancer, although the median mutation burden remained 3 in both compartments, enrichment of AR-pathway alterations and extensive copy-number scarring are consistent with AR-driven evolution in advanced disease [17–19]. In colorectal cancer, the difference was modest (median 8 vs. 7), and no single-gene differences survived multiple-testing correction, fitting a model of quantitatively higher but qualitatively similar profiles shaped by early dissemination and parallel evolution [5–7]. In pancreatic cancer, metastases showed a small but reproducible increase (median 5 vs. 4) despite overall low burdens, consistent with reports of late dissemination and limited driver diversity [26–28]. In NSCLC, mutation burden (median 8 vs. 6) and FGA (median 0.19 vs. 0.09) were higher in metastases, whereas common drivers (TP53, EGFR, KRAS, CDKN2A, STK11) were shared across compartments and no single-gene differences remained after correction, supporting a model of copy-number–driven adaptation rather than acquisition of new dominant drivers [2–4]. Targetable alterations (ALK, ROS1, RET, ERBB2/HER2, MET, NTRK) were rare in both groups, and the high EGFR prevalence likely reflects clinically enriched testing populations [29,30].

Among other tumor types, melanoma displayed very high but variable burdens in both compartments, consistent with UV mutagenesis and MAPK pathway dominance (BRAF, NRAS, NF1) [3,11,20–22]. Endometrial cancer showed a higher median burden in metastases (∼8 vs. 6), although this was not significant after correction (q=0.14), consistent with hypermutation from mismatch-repair deficiency or POLE exonuclease mutations, often co-occurring with PTEN, PIK3CA, and ARID1A alterations [3,11,20–22]. In renal cell carcinoma, mutation burden was similar between primary and metastatic tumors. Metastatic progression typically occurs along constrained evolutionary routes, with truncal VHL mutations accompanied by heterogeneous branch-level alterations in PBRM1, SETD2, or BAP1. Thus, even though total burden remains comparable across compartments, underlying clonal diversity persists [31,32].

Taken together, these findings suggest that increased mutation burden in metastases is concentrated in selected lineages, while copy-number and structural changes are more universal hallmarks of metastatic adaptation. Importantly, a higher mutation burden should not be interpreted as synonymous with more actionable therapeutic opportunities. Interpretation is improved when variant function, clonality, and prior exposures are considered alongside mutation counts [2–4,13,16–19,20–22]. Moreover, panel-derived tumor mutational burden should be interpreted with caution, particularly in treated populations where chemotherapy-induced signatures (e.g., platinum, temozolomide) can inflate counts without adding proportionate neoantigens [12,14]. Notably, although metastases generally showed higher burdens, even the tumor types with statistically significant increases remained below the 10 mut/Mb threshold that underpins tumor-agnostic immunotherapy approvals. As such, analyses of this type may provide limited incremental value for identifying patients eligible for tumor-agnostic immune checkpoint inhibitors.

This study has several important limitations that should be considered when interpreting the findings. It is a retrospective, multi-center analysis built from the public AACR Project GENIE v18.0 release via cBioPortal, in which several clinical fields are masked in the public (“clinical tier”) files; as a result, prior treatments, disease stage at sampling, and exact sequencing timelines are not consistently available [33,34]. As a result, detailed clinical information, including prior lines of therapy, treatment exposures, and disease stage at the time of biopsy, was not uniformly available. These factors can strongly influence the mutational landscape, particularly the emergence of therapy-associated resistance alterations, but could not be systematically evaluated here. Second, the dataset was derived from targeted next-generation sequencing panels rather than whole-exome or whole-genome sequencing. While panel-based assays enable broad clinical coverage across centers, they differ in gene content, probe design, and analytic pipelines. Such heterogeneity limits cross-panel comparability and may underestimate structural variants, copy-number complexity, and mutational processes that extend beyond targeted footprints. Mutation burden estimates were therefore restricted to the boundaries of each panel and are not directly equivalent to exome-derived tumor mutational burden. Third, the inference of sampling order was based on available metadata, and in some cases sequencing of metastatic lesions may not have occurred strictly later than primary tumor sampling. Although discordant cases were excluded when identifiable, some uncertainty remains regarding temporal relationships between paired samples. Fourth, survival analyses were derived from age-anchored intervals in the clinical files. These summaries describe survival after sequencing but cannot disentangle prognostic implications from sampling context, clinical stage, or treatment history.Finally, the study was limited by sample size in several cancer subtypes. While significant differences in mutation burden and genomic instability were observed in common cancers such as NSCLC, breast, colorectal, pancreatic, and prostate tumors, analyses in rarer lineages were often underpowered, leaving open the possibility that additional compartment-specific differences exist but were not detected.

### Conclusion

Metastatic tumors generally preserve the truncal drivers of their primaries but exhibit lineage-dependent shifts shaped by timing, treatment exposure, and tissue context. Genome-wide instability, reflected in higher FGA, recurrent CNA, and structural alterations, emerges as a consistent hallmark of metastases, whereas increases in mutation burden are confined to a subset of lineages. Importantly, even where burden was higher, values typically remained below the threshold used for tumor-agnostic immunotherapy. Thus, while primary–metastatic comparisons provide valuable biological insights, their role in expanding the pool of patients eligible for tumor-agnostic immunotherapies may be limited. Primary sequencing remains an adequate baseline for identifying core actionable drivers, and re-biopsy at progression is most informative in lineages where context-dependent changes (e.g., ESR1, AR, focal CNA/SV) are expected.

## Data Availability

All data used in this study were obtained from the AACR Project GENIE dataset via cBioPortal (https://www.cbioportal.org/
) and are publicly available. Derived results supporting the findings are provided within the manuscript and supplementary files.

## Notes

**Funding:** None

### Competing Interest Statement

The authors have declared no competing interest.

### Funding Statement

This study did not receive any funding

