## Supplementary Material for "Genomic Divergence Between Matched Primary and Metastatic Tumors Across Cancer Types: A Pan-Cancer Analysis of 5,692 Samples"

### **Supplementary Material – Table of Contents**

### Table S1. Baseline Characteristics for NSCLC

### Figure S1. Mutation Burden Comparison in Primary and Metastatic NSCLC

### Figure S2. Distribution of the Most Frequently Mutated Genes in Primary and Metastatic NSCLC

### Table S2. Gene-Level Mutation Frequencies in Primary Versus Metastatic NSCLC Samples

### Table S3. Baseline Characteristics for Breast Cancer

### Figure S3. Mutation Burden Comparison in Primary and Metastatic Breast Cancer

### Figure S4. Distribution of the Most Frequently Mutated Genes in Primary and Metastatic Breast Cancer

### Table S4. Gene-Level Mutation Frequencies in Primary Versus Metastatic Breast Cancer Samples

### Table S5. Baseline Characteristics for Colorectal Cancer

### Figure S5. Mutation Burden Comparison in Primary and Metastatic Colorectal Cancer

### Figure S6. Distribution of the Most Frequently Mutated Genes in Primary and Metastatic Colorectal Cancer

### Table S6. Gene-Level Mutation Frequencies in Primary Versus Metastatic Colorectal Cancer Samples

### Table S7. Prostate Cancer Demographic and Survival Characteristics

### Figure S7. Distribution of the Most Frequently Mutated Genes in Primary and Metastatic Prostate Cancer

### Table S8. Gene-Level Mutation Frequencies in Primary Versus Metastatic Prostate Cancer Samples

### Figure S8. Distribution of the Most Frequently Mutated Genes in Primary and Metastatic Melanoma

### Table S9. Gene-Level Mutation Frequencies in Primary Versus Metastatic Melanoma Samples

### Figure S9. Distribution of the Most Frequently Mutated Genes in Primary and Metastatic Endometrial Cancer

### Table S10. Gene-Level Mutation Frequencies in Primary Versus Metastatic Endometrial Cancer Samples

### Figure S10. Distribution of the Most Frequently Mutated Genes in Primary and Metastatic Bladder Cancer

### Table S11. Gene-Level Mutation Frequencies in Primary Versus Metastatic Bladder Cancer Samples

### Figure S11. Distribution of the Most Frequently Mutated Genes in Primary and Metastatic Ovarian Cancer

### Table S12. Gene-Level Mutation Frequencies in Primary Versus Metastatic Ovarian Cancer Samples

### Figure S12. Distribution of the Most Frequently Mutated Genes in Primary and Metastatic Pancreatic Cancer

### Table S13. Gene-Level Mutation Frequencies in Primary Versus Metastatic Pancreatic Cancer Samples

### Figure S13. Distribution of the Most Frequently Mutated Genes in Primary and Metastatic Gastroesophageal Cancer

### Table S14. Gene-Level Mutation Frequencies in Primary Versus Metastatic Gastroesophageal Cancer Samples

### Figure S14. Distribution of the Most Frequently Mutated Genes in Primary and Metastatic Soft Tissue Sarcoma

### Table S15. Gene-Level Mutation Frequencies in Primary Versus Metastatic Soft Tissue Sarcoma Samples

### Figure S15. Distribution of the Most Frequently Mutated Genes in Primary and Metastatic Clear Cell Renal Cell Carcinoma

### Table S16. Gene-Level Mutation Frequencies in Primary Versus Metastatic Clear Cell Renal Cell Carcinoma Samples

### Figure S16. Distribution of the Most Frequently Mutated Genes in Primary and Metastatic Gastrointestinal Stromal Tumors (GIST)

### Table S17. Gene-Level Mutation Frequencies in Primary Versus Metastatic Gastrointestinal Stromal Tumors (GIST)

### Figure S17. Distribution of the Most Frequently Mutated Genes in Primary and Metastatic Biliary Tract Cancer

### Table S18. Gene-Level Mutation Frequencies in Primary Versus Metastatic Biliary Tract Cancer Samples

### Figure S18. Distribution of the Most Frequently Mutated Genes in Primary and Metastatic Glioblastoma

### Table S19. Gene-Level Mutation Frequencies in Primary Versus Metastatic Glioblastoma Samples

### Table S20. Gene-Level Mutation Frequencies in Primary Versus Metastatic Tumors Across the Entire Cohort

### **Table S1. Baseline Characteristics for NSCLC**

| Section | Characteristic | Level | N | Percent | Median |
| --- | --- | --- | --- | --- | --- |
| Overall | All patients |  | 671 | 100 |  |
| Age | Age at sequencing (years) |  |  |  | 62 |
| Sex | Sex | Female | 348 | 51.9 |  |
|  |  | Male | 310 | 46.2 |  |
|  |  | Unknown | 13 | 1.9 |  |
| Ethnicity | Ethnicity | Non-Spanish/non-Hispanic | 565 | 84.2 |  |
|  |  | Spanish/Hispanic | 31 | 4.6 |  |
|  |  | Unknown | 63 | 9.4 |  |
|  |  | Not Collected | 12 | 1.8 |  |
| Race | Race | White | 514 | 76.6 |  |
|  |  | Asian | 49 | 7.3 |  |
|  |  | Black | 41 | 6.1 |  |
|  |  | Other | 18 | 2.7 |  |
|  |  | Unknown | 39 | 5.8 |  |
|  |  | Native American | 2 | 0.3 |  |
|  |  | Pacific Islander | 1 | 0.1 |  |
| Survival | Median survival after sequencing (years) |  |  |  | 2.41 |
| Survival | Median age at death (years) |  |  |  | 65.8 |

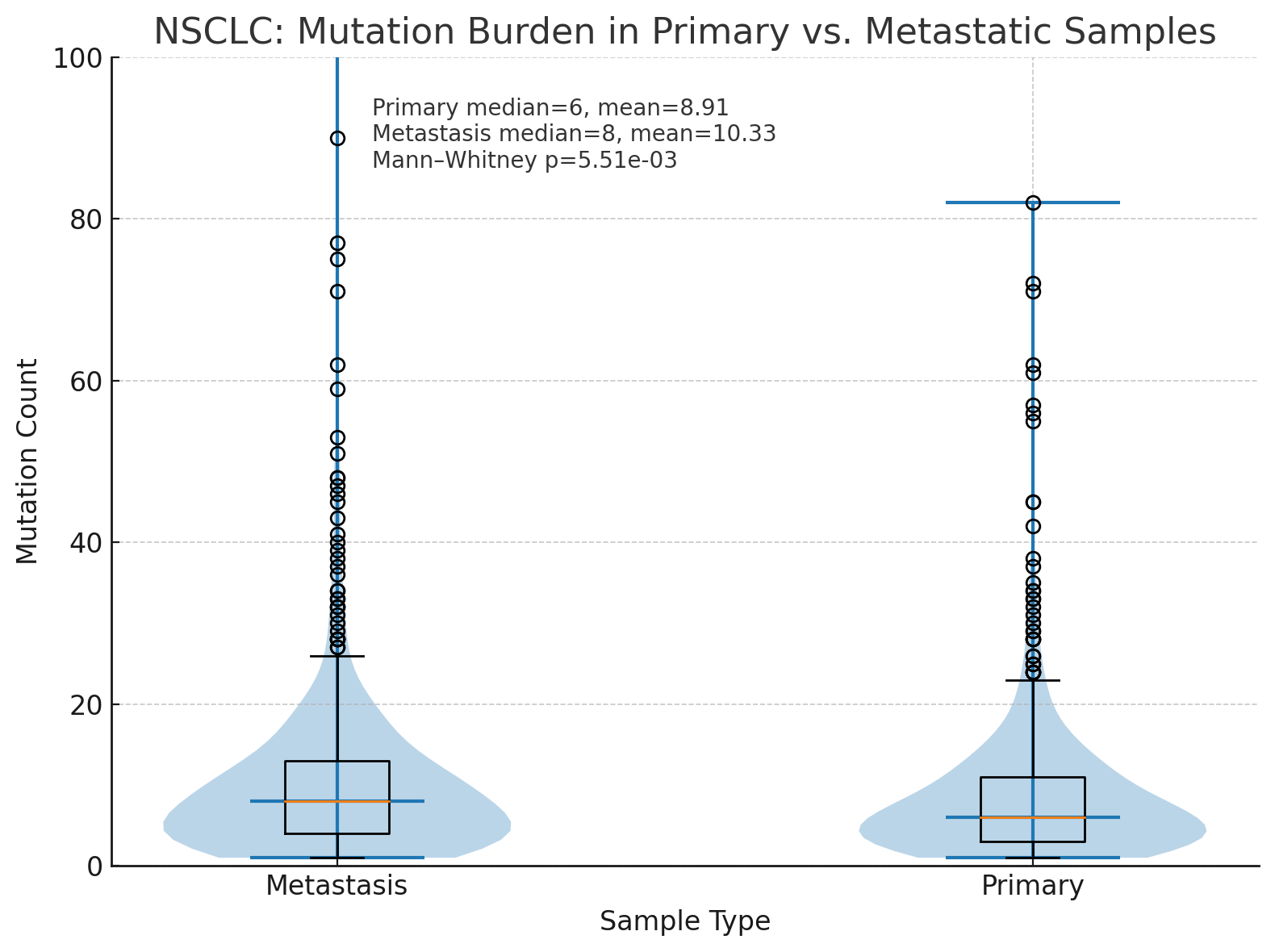

**Figure S1.** Mutation Burden Comparison in Primary and Metastatic NSCLC

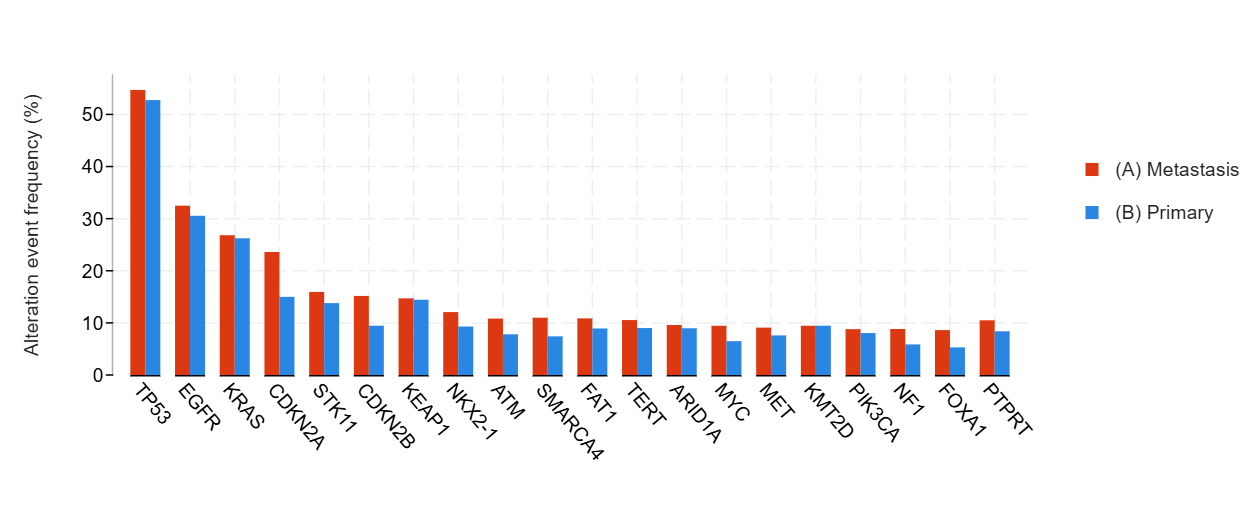

**Figure S2.** Distribution of the Most Frequently Mutated Genes in Primary and Metastatic NSCLC

**Table S2. Gene-Level Mutation Frequencies in Primary Versus Metastatic NSCLC**

| **Gene** | **Cytoband** | **(A) Metastasis** | **(B) Primary** | **Co-occurrence Pattern** | **Log2 Ratio** | **p-Value** | **q-Value** | **Enriched in** |
| --- | --- | --- | --- | --- | --- | --- | --- | --- |
| **TP53** | 17p13.1 | 367 (54.69%) | 354 (52.76%) |  | 0.05 | 0.511 | 1.00 | (A) Metastasis |
| **EGFR** | 7p11.2 | 218 (32.49%) | 205 (30.55%) |  | 0.09 | 0.481 | 1.00 | (A) Metastasis |
| **KRAS** | 12p12.1 | 180 (26.83%) | 176 (26.23%) |  | 0.03 | 0.853 | 1.00 | (A) Metastasis |
| **CDKN2A** | 9p21.3 | 157 (23.61%) | 100 (14.99%) |  | 0.66 | 7,29E-02 | 0.0556 | (A) Metastasis |
| **STK11** | 19p13.3 | 106 (15.94%) | 92 (13.79%) |  | 0.21 | 0.282 | 1.00 | (A) Metastasis |
| **CDKN2B** | 9p21.3 | 98 (15.17%) | 61 (9.46%) |  | 0.68 | 2,22E+00 | 0.0806 | (A) Metastasis |
| **KEAP1** | 19p13.2 | 95 (14.71%) | 92 (14.44%) |  | 0.03 | 0.937 | 1.00 | (A) Metastasis |
| **NKX2-1** | 14q13.3 | 78 (12.07%) | 60 (9.30%) |  | 0.38 | 0.125 | 1.00 | (A) Metastasis |
| **ATM** | 11q22.3 | 72 (10.83%) | 52 (7.81%) |  | 0.47 | 0.0599 | 1.00 | (A) Metastasis |
| **SMARCA4** | 19p13.2 | 71 (10.99%) | 48 (7.42%) |  | 0.57 | 0.0272 | 0.759 | (A) Metastasis |
| **FAT1** | 4q35.2 | 68 (10.86%) | 54 (8.93%) |  | 0.28 | 0.294 | 1.00 | (A) Metastasis |
| **TERT** | 5p15.33 | 68 (10.54%) | 58 (9.01%) |  | 0.23 | 0.399 | 1.00 | (A) Metastasis |
| **ARID1A** | 1p36.11 | 62 (9.60%) | 58 (8.96%) |  | 0.10 | 0.703 | 1.00 | (A) Metastasis |
| **MYC** | 8q24.21 | 61 (9.44%) | 42 (6.49%) |  | 0.54 | 0.0516 | 1.00 | (A) Metastasis |
| **MET** | 7q31.2 | 61 (9.09%) | 51 (7.60%) |  | 0.26 | 0.374 | 1.00 | (A) Metastasis |
| **KMT2D** | 12q13.12 | 61 (9.44%) | 61 (9.46%) |  | -0.00 | 1.00 | 1.00 | (B) Primary |
| **PIK3CA** | 3q26.32 | 59 (8.79%) | 54 (8.05%) |  | 0.13 | 0.694 | 1.00 | (A) Metastasis |
| **NF1** | 17q11.2 | 57 (8.82%) | 38 (5.87%) |  | 0.59 | 0.0434 | 1.00 | (A) Metastasis |
| **FOXA1** | 14q21.1 | 54 (8.61%) | 32 (5.30%) |  | 0.70 | 0.0251 | 0.738 | (A) Metastasis |
| **PTPRT** | 20q12-q13.11 | 53 (10.50%) | 42 (8.38%) |  | 0.32 | 0.281 | 1.00 | (A) Metastasis |
| **MTAP** | 9p21.3 | 52 (13.65%) | 27 (8.88%) |  | 0.62 | 0.0548 | 1.00 | (A) Metastasis |
| **RBM10** | Xp11.3 | 52 (8.29%) | 49 (8.09%) |  | 0.04 | 0.918 | 1.00 | (A) Metastasis |
| **NFKBIA** | 14q13.2 | 50 (7.86%) | 39 (6.19%) |  | 0.34 | 0.272 | 1.00 | (A) Metastasis |
| **ALK** | 2p23.2-p23.1 | 50 (7.45%) | 47 (7.00%) |  | 0.09 | 0.753 | 1.00 | (A) Metastasis |
| **SDHA** | 5p15.33 | 49 (7.59%) | 36 (5.67%) |  | 0.42 | 0.179 | 1.00 | (A) Metastasis |
| **APC** | 5q22.2 | 47 (7.07%) | 34 (5.11%) |  | 0.47 | 0.138 | 1.00 | (A) Metastasis |
| **MDM2** | 12q15 | 46 (7.12%) | 36 (5.58%) |  | 0.35 | 0.304 | 1.00 | (A) Metastasis |
| **SETD2** | 3p21.31 | 43 (6.66%) | 35 (5.41%) |  | 0.30 | 0.353 | 1.00 | (A) Metastasis |
| **SMAD4** | 18q21.2 | 42 (6.32%) | 30 (4.50%) |  | 0.49 | 0.148 | 1.00 | (A) Metastasis |
| **ERBB4** | 2q34 | 42 (6.32%) | 31 (4.66%) |  | 0.44 | 0.188 | 1.00 | (A) Metastasis |
| **ATRX** | Xq21.1 | 42 (6.50%) | 46 (7.13%) |  | -0.13 | 0.661 | 1.00 | (B) Primary |
| **ERBB2** | 17q12 | 40 (5.96%) | 35 (5.22%) |  | 0.19 | 0.635 | 1.00 | (A) Metastasis |
| **ROS1** | 6q22.1 | 39 (5.98%) | 28 (4.30%) |  | 0.48 | 0.209 | 1.00 | (A) Metastasis |
| **PTPRD** | 9p24.1-p23 | 39 (7.46%) | 39 (7.32%) |  | 0.03 | 1.00 | 1.00 | (A) Metastasis |
| **RET** | 10q11.21 | 39 (5.81%) | 38 (5.66%) |  | 0.04 | 1.00 | 1.00 | (A) Metastasis |
| **CREBBP** | 16p13.3 | 38 (5.88%) | 28 (4.33%) |  | 0.44 | 0.209 | 1.00 | (A) Metastasis |
| **RB1** | 13q14.2 | 38 (5.71%) | 38 (5.71%) |  | 0.00 | 1.00 | 1.00 | (A) Metastasis |
| **FLT1** | 13q12.3 | 37 (5.73%) | 24 (3.72%) |  | 0.62 | 0.115 | 1.00 | (A) Metastasis |
| **MGA** | 15q15.1 | 37 (6.02%) | 31 (5.28%) |  | 0.19 | 0.619 | 1.00 | (A) Metastasis |
| **BRAF** | 7q34 | 37 (5.51%) | 37 (5.51%) |  | - | 1.00 | 1.00 | (B) Primary |
| **IL7R** | 5p13.2 | 36 (5.74%) | 27 (4.46%) |  | 0.36 | 0.365 | 1.00 | (A) Metastasis |
| **CARD11** | 7p22.2 | 35 (5.42%) | 25 (3.86%) |  | 0.49 | 0.189 | 1.00 | (A) Metastasis |
| **NOTCH1** | 9q34.3 | 34 (5.07%) | 20 (2.99%) |  | 0.76 | 0.0700 | 1.00 | (A) Metastasis |
| **PIK3CG** | 7q22.3 | 32 (6.34%) | 22 (4.37%) |  | 0.54 | 0.208 | 1.00 | (A) Metastasis |
| **KMT2C** | 7q36.1 | 32 (6.34%) | 34 (6.75%) |  | -0.09 | 0.801 | 1.00 | (B) Primary |
| **GRIN2A** | 16p13.2 | 31 (6.14%) | 23 (4.56%) |  | 0.43 | 0.328 | 1.00 | (A) Metastasis |
| **FLT4** | 5q35.3 | 31 (4.81%) | 24 (3.73%) |  | 0.37 | 0.409 | 1.00 | (A) Metastasis |
| **NTRK3** | 15q25.3 | 31 (4.80%) | 32 (4.95%) |  | -0.04 | 1.00 | 1.00 | (B) Primary |
| **ARID2** | 12q12 | 30 (4.65%) | 29 (4.49%) |  | 0.05 | 0.895 | 1.00 | (A) Metastasis |
| **KDM5A** | 12p13.33 | 29 (4.62%) | 17 (2.81%) |  | 0.72 | 0.0999 | 1.00 | (A) Metastasis |
| **EPHA3** | 3p11.1 | 29 (5.54%) | 24 (4.41%) |  | 0.33 | 0.402 | 1.00 | (A) Metastasis |
| **ZFHX3** | 16q22.2-q22.3 | 29 (5.87%) | 23 (4.73%) |  | 0.31 | 0.477 | 1.00 | (A) Metastasis |
| **BRCA2** | 13q13.1 | 29 (4.49%) | 25 (3.86%) |  | 0.22 | 0.582 | 1.00 | (A) Metastasis |
| **FLT3** | 13q12.2 | 28 (4.21%) | 19 (2.85%) |  | 0.56 | 0.185 | 1.00 | (A) Metastasis |
| **RICTOR** | 5p13.1 | 28 (4.46%) | 19 (3.14%) |  | 0.51 | 0.237 | 1.00 | (A) Metastasis |
| **KDR** | 4q12 | 28 (4.21%) | 20 (3.01%) |  | 0.48 | 0.303 | 1.00 | (A) Metastasis |
| **TET1** | 10q21.3 | 28 (4.47%) | 20 (3.32%) |  | 0.43 | 0.307 | 1.00 | (A) Metastasis |
| **NOTCH3** | 19p13.12 | 28 (4.47%) | 22 (3.63%) |  | 0.30 | 0.474 | 1.00 | (A) Metastasis |
| **KMT2A** | 11q23.3 | 28 (4.33%) | 23 (3.57%) |  | 0.28 | 0.568 | 1.00 | (A) Metastasis |
| **POLE** | 12q24.33 | 28 (4.47%) | 24 (3.94%) |  | 0.18 | 0.673 | 1.00 | (A) Metastasis |
| **PREX2** | 8q13.2 | 28 (6.31%) | 26 (6.22%) |  | 0.02 | 1.00 | 1.00 | (A) Metastasis |
| **DDR2** | 1q23.3 | 27 (4.14%) | 21 (3.23%) |  | 0.36 | 0.462 | 1.00 | (A) Metastasis |
| **PDGFRA** | 4q12 | 27 (4.04%) | 22 (3.29%) |  | 0.30 | 0.561 | 1.00 | (A) Metastasis |
| **CDK4** | 12q14.1 | 27 (4.18%) | 25 (3.86%) |  | 0.11 | 0.779 | 1.00 | (A) Metastasis |
| **NFE2L2** | 2q31.2 | 27 (4.18%) | 25 (3.86%) |  | 0.11 | 0.779 | 1.00 | (A) Metastasis |
| **MED12** | Xq13.1 | 27 (4.30%) | 28 (4.61%) |  | -0.10 | 0.890 | 1.00 | (B) Primary |
| **BCOR** | Xp11.4 | 27 (4.18%) | 28 (4.33%) |  | -0.05 | 1.00 | 1.00 | (B) Primary |
| **EPHB1** | 3q22.2 | 26 (5.14%) | 15 (2.97%) |  | 0.79 | 0.110 | 1.00 | (A) Metastasis |
| **ASXL1** | 20q11.21 | 26 (4.02%) | 18 (2.79%) |  | 0.53 | 0.283 | 1.00 | (A) Metastasis |
| **NOTCH2** | 1p12 | 26 (4.02%) | 19 (2.95%) |  | 0.45 | 0.363 | 1.00 | (A) Metastasis |
| **RAD21** | 8q24.11 | 26 (4.09%) | 19 (3.03%) |  | 0.43 | 0.363 | 1.00 | (A) Metastasis |
| **AR** | Xq12 | 26 (4.02%) | 21 (3.25%) |  | 0.31 | 0.462 | 1.00 | (A) Metastasis |
| **ATR** | 3q23 | 26 (4.14%) | 23 (3.80%) |  | 0.13 | 0.773 | 1.00 | (A) Metastasis |
| **FGFR1** | 8p11.23 | 26 (3.87%) | 24 (3.58%) |  | 0.12 | 0.776 | 1.00 | (A) Metastasis |
| **NSD1** | 5q35.3 | 26 (4.15%) | 24 (3.97%) |  | 0.06 | 0.886 | 1.00 | (A) Metastasis |
| **GLI1** | 12q13.3 | 26 (4.10%) | 27 (4.29%) |  | -0.07 | 0.889 | 1.00 | (B) Primary |
| **NBN** | 8q21.3 | 25 (3.87%) | 13 (2.02%) |  | 0.94 | 0.0688 | 1.00 | (A) Metastasis |
| **JAK2** | 9p24.1 | 25 (3.76%) | 14 (2.11%) |  | 0.83 | 0.103 | 1.00 | (A) Metastasis |
| **NTRK1** | 1q23.1 | 25 (3.87%) | 16 (2.47%) |  | 0.65 | 0.157 | 1.00 | (A) Metastasis |
| **PMS2** | 7p22.1 | 25 (3.87%) | 17 (2.63%) |  | 0.56 | 0.214 | 1.00 | (A) Metastasis |

### **Table S3. Baseline Characteristics for Breast Cancer**

| Section | Characteristic | Level | N | Percent | Median / Q1–Q3 |
| --- | --- | --- | --- | --- | --- |
| Overall | All patients |  | 401 | 100 |  |
| Age | Age at sequencing (years) |  |  |  | 55.0 (46.0–62.0) |
| Sex | Sex | Female | 382 | 95.3 |  |
|  |  | Unknown | 16 | 4.0 |  |
|  |  | Male | 3 | 0.7 |  |
| Ethnicity | Ethnicity | Non-Spanish/non-Hispanic | 328 | 81.8 |  |
|  |  | Unknown | 45 | 11.2 |  |
|  |  | Spanish/Hispanic | 25 | 6.2 |  |
|  |  | Not Collected | 3 | 0.7 |  |
| Race | Race | White | 273 | 68.1 |  |
|  |  | Black | 41 | 10.2 |  |
|  |  | Unknown | 37 | 9.2 |  |
|  |  | Asian | 32 | 8.0 |  |
|  |  | Other | 18 | 4.5 |  |
|  |  | Unknown | 16 | 4.0 |  |
| Survival | Median age at death (years) |  |  |  | 56.0 |

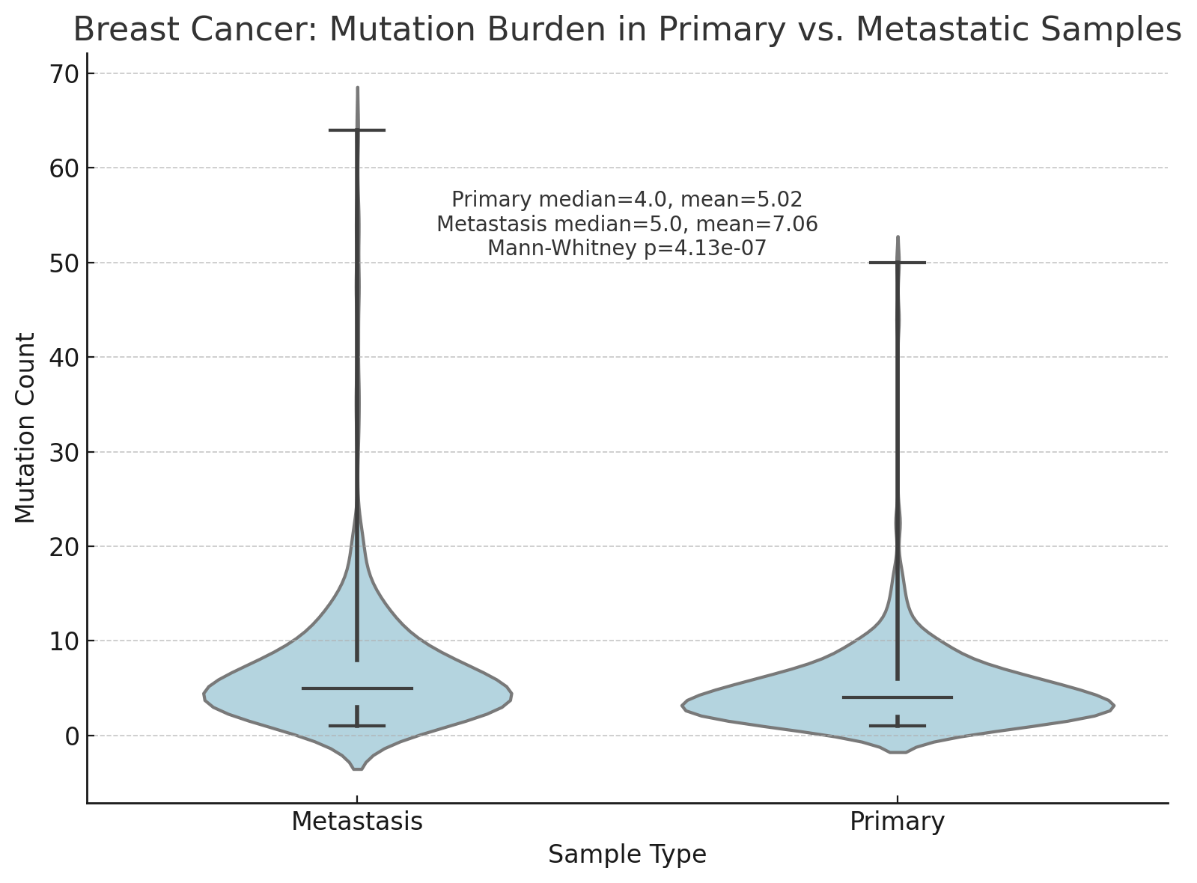

**Figure S3.** Mutation Burden Comparison in Primary and Metastatic Breast Cancer

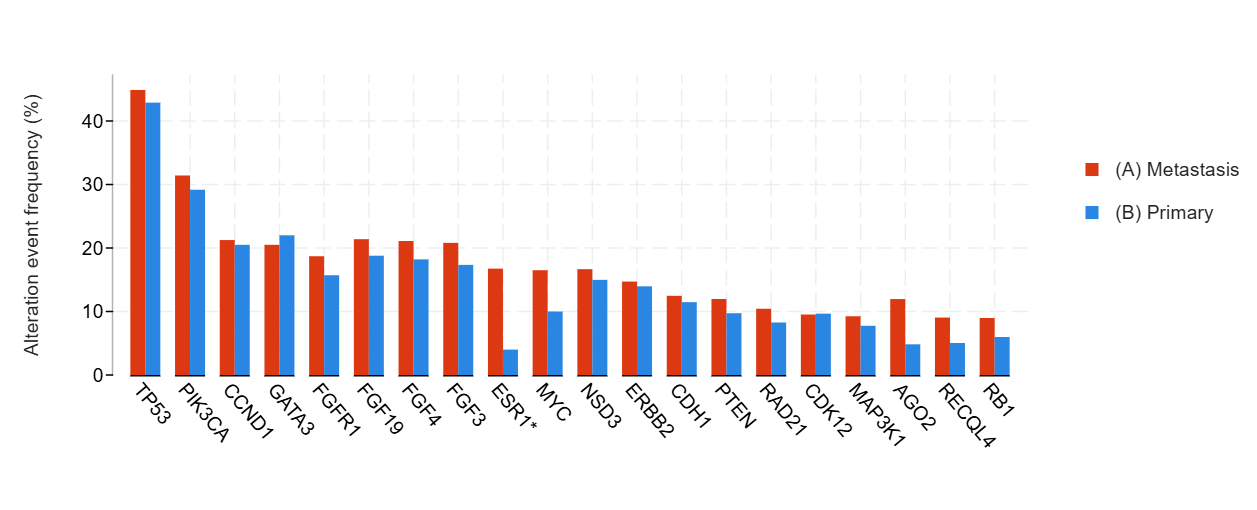

**Figure S4.** Distribution of the Most Frequently Mutated Genes in Primary and Metastatic Breast Cancer

Table S4. **Gene-Level Mutation Frequencies in Primary Versus Metastatic Breast Cancer**

| **Gene** | **Cytoband** | **(A) Metastasis** | **(B) Primary** | **Co-occurrence Pattern** | **Log2 Ratio** | **p-Value** | **q-Value** | **Enriched in** |
| --- | --- | --- | --- | --- | --- | --- | --- | --- |
| **TP53** | 17p13.1 | 180 (44.89%) | 172 (42.89%) |  | 0.07 | 0.618 | 1.00 | (A) Metastasis |
| **PIK3CA** | 3q26.32 | 126 (31.42%) | 117 (29.18%) |  | 0.11 | 0.539 | 1.00 | (A) Metastasis |
| **CCND1** | 11q13.3 | 85 (21.25%) | 82 (20.50%) |  | 0.05 | 0.862 | 1.00 | (A) Metastasis |
| **GATA3** | 10p14 | 82 (20.50%) | 88 (22.00%) |  | -0.10 | 0.666 | 1.00 | (B) Primary |
| **FGFR1** | 8p11.23 | 75 (18.70%) | 63 (15.71%) |  | 0.25 | 0.303 | 1.00 | (A) Metastasis |
| **FGF19** | 11q13.3 | 74 (21.39%) | 65 (18.79%) |  | 0.19 | 0.448 | 1.00 | (A) Metastasis |
| **FGF4** | 11q13.3 | 73 (21.10%) | 63 (18.21%) |  | 0.21 | 0.389 | 1.00 | (A) Metastasis |
| **FGF3** | 11q13.3 | 72 (20.81%) | 60 (17.34%) |  | 0.26 | 0.287 | 1.00 | (A) Metastasis |
| **ESR1** | 6q25.1-q25.2 | 67 (16.75%) | 16 (4.00%) |  | 2.07. | **2.30e-9** | **1,71E-03** | **(A) Metastasis** |
| **MYC** | 8q24.21 | 66 (16.50%) | 40 (10.00%) |  | 0.72 | 8,89E+00 | 1.00 | (A) Metastasis |
| **NSD3** | 8p11.23 | 59 (16.67%) | 46 (14.98%) |  | 0.15 | 0.594 | 1.00 | (A) Metastasis |
| **ERBB2** | 17q12 | 59 (14.71%) | 56 (13.97%) |  | 0.08 | 0.840 | 1.00 | (A) Metastasis |
| **CDH1** | 16q22.1 | 50 (12.47%) | 46 (11.47%) |  | 0.12 | 0.744 | 1.00 | (A) Metastasis |
| **PTEN** | 10q23.31 | 48 (11.97%) | 39 (9.73%) |  | 0.30 | 0.364 | 1.00 | (A) Metastasis |
| **RAD21** | 8q24.11 | 41 (10.43%) | 31 (8.27%) |  | 0.34 | 0.324 | 1.00 | (A) Metastasis |
| **CDK12** | 17q12 | 38 (9.52%) | 37 (9.66%) |  | -0.02 | 1.00 | 1.00 | (B) Primary |
| **MAP3K1** | 5q11.2 | 37 (9.25%) | 31 (7.75%) |  | 0.26 | 0.526 | 1.00 | (A) Metastasis |
| **AGO2** | 8q24.3 | 36 (11.96%) | 13 (4.83%) |  | 1.31. | 2,58E+00 | 0.956 | (A) Metastasis |
| **RECQL4** | 8q24.3 | 36 (9.05%) | 20 (5.04%) |  | 0.84 | 0.0368 | 1.00 | (A) Metastasis |
| **RB1** | 13q14.2 | 36 (8.98%) | 24 (5.99%) |  | 0.58 | 0.139 | 1.00 | (A) Metastasis |
| **PAK1** | 11q13.5-q14.1 | 34 (9.91%) | 29 (8.45%) |  | 0.23 | 0.597 | 1.00 | (A) Metastasis |
| **FOXA1** | 14q21.1 | 34 (8.56%) | 32 (8.40%) |  | 0.03 | 1.00 | 1.00 | (A) Metastasis |
| **ARID1A** | 1p36.11 | 33 (8.25%) | 25 (6.25%) |  | 0.40 | 0.340 | 1.00 | (A) Metastasis |
| **MAP2K4** | 17p12 | 32 (8.00%) | 27 (6.75%) |  | 0.25 | 0.503 | 1.00 | (A) Metastasis |
| **NBN** | 8q21.3 | 29 (7.25%) | 21 (5.25%) |  | 0.47 | 0.307 | 1.00 | (A) Metastasis |
| **PPM1D** | 17q23.2 | 29 (7.42%) | 29 (8.15%) |  | -0.14 | 0.785 | 1.00 | (B) Primary |
| **PREX2** | 8q13.2 | 28 (9.18%) | 11 (4.04%) |  | 1.18. | 0.0191 | 1.00 | (A) Metastasis |
| **PRDM14** | 8q13.3 | 27 (8.97%) | 10 (3.72%) |  | 1.27. | 0.0161 | 1.00 | (A) Metastasis |
| **KMT2C** | 7q36.1 | 27 (7.80%) | 18 (5.22%) |  | 0.58 | 0.217 | 1.00 | (A) Metastasis |
| **NF1** | 17q11.2 | 27 (6.75%) | 22 (5.50%) |  | 0.30 | 0.556 | 1.00 | (A) Metastasis |
| **MDM2** | 12q15 | 27 (6.75%) | 23 (5.75%) |  | 0.23 | 0.662 | 1.00 | (A) Metastasis |
| **MCL1** | 1q21.2 | 26 (6.50%) | 23 (5.75%) |  | 0.18 | 0.768 | 1.00 | (A) Metastasis |
| **TBX3** | 12q24.21 | 26 (7.54%) | 28 (8.12%) |  | -0.11 | 0.887 | 1.00 | (B) Primary |
| **GNAS** | 20q13.32 | 25 (6.23%) | 19 (4.74%) |  | 0.40 | 0.438 | 1.00 | (A) Metastasis |
| **AKT1** | 14q32.33 | 25 (6.23%) | 22 (5.49%) |  | 0.18 | 0.764 | 1.00 | (A) Metastasis |
| **ELOC** | 8q21.11 | 24 (6.14%) | 12 (3.37%) |  | 0.86 | 0.0882 | 1.00 | (A) Metastasis |
| **FAT1** | 4q35.2 | 24 (6.03%) | 15 (3.94%) |  | 0.62 | 0.192 | 1.00 | (A) Metastasis |
| **KMT2D** | 12q13.12 | 23 (5.75%) | 11 (2.75%) |  | 1.06. | 0.0524 | 1.00 | (A) Metastasis |
| **AXIN2** | 17q24.1 | 23 (5.81%) | 21 (5.53%) |  | 0.07 | 0.878 | 1.00 | (A) Metastasis |
| **RTEL1** | 20q13.33 | 23 (7.64%) | 20 (7.43%) |  | 0.04 | 1.00 | 1.00 | (A) Metastasis |

### **Table S6. Baseline Characteristics for Colorectal Cancer**

| Section | Characteristic | Level | N | Percent | Median | Q1–Q3 |
| --- | --- | --- | --- | --- | --- | --- |
| Overall | All patients |  | 296 | 100 |  |  |
| Age | Age at sequencing (years) |  |  |  | 60 | 46–64 |
| Sex | Sex | Female | 135 | 45.6 |  |  |
|  |  | Male | 158 | 53.4 |  |  |
|  |  | Unknown | 3 | 1.0 |  |  |
| Ethnicity | Ethnicity | Non-Spanish/non-Hispanic | 231 | 78.0 |  |  |
|  |  | Spanish/Hispanic | 16 | 5.4 |  |  |
|  |  | Unknown | 43 | 14.5 |  |  |
|  |  | Not Collected | 6 | 2.0 |  |  |
| Race | Race | White | 210 | 71.0 |  |  |
|  |  | Asian | 27 | 9.1 |  |  |
|  |  | Black | 31 | 10.5 |  |  |
|  |  | Other | 9 | 3.0 |  |  |
|  |  | Unknown | 18 | 6.1 |  |  |
|  |  | Native American | 1 | 0.3 |  |  |
|  |  | Pacific Islander | 0 | 0.0 |  |  |
| Survival | Median survival after sequencing (years) |  |  |  | 2.35 |  |
| Survival | Median age at death (years) |  |  |  | 64.2 |  |

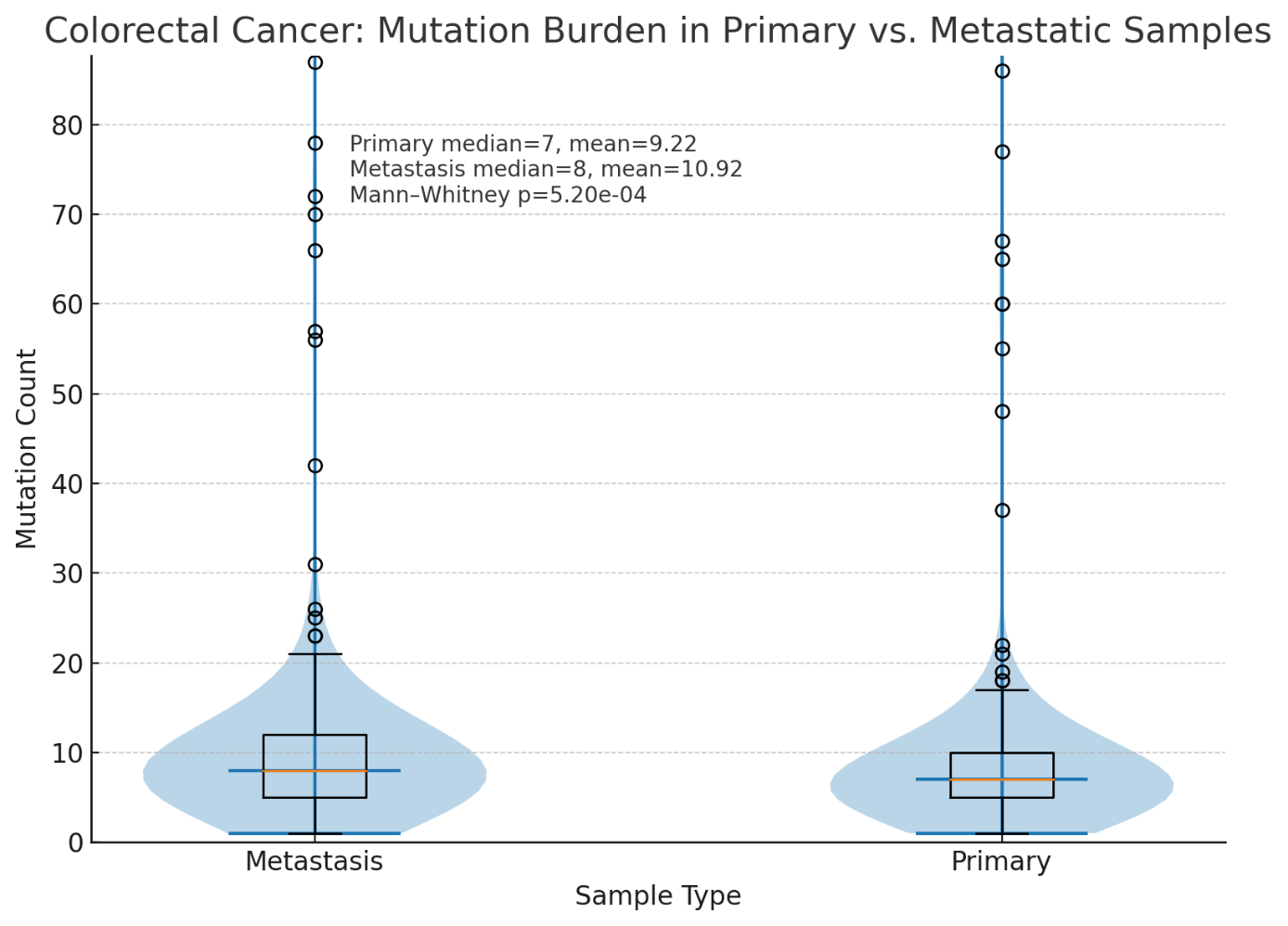

**Figure S5.** Mutation Burden Comparison in Primary and Metastatic Colorectal Cancer

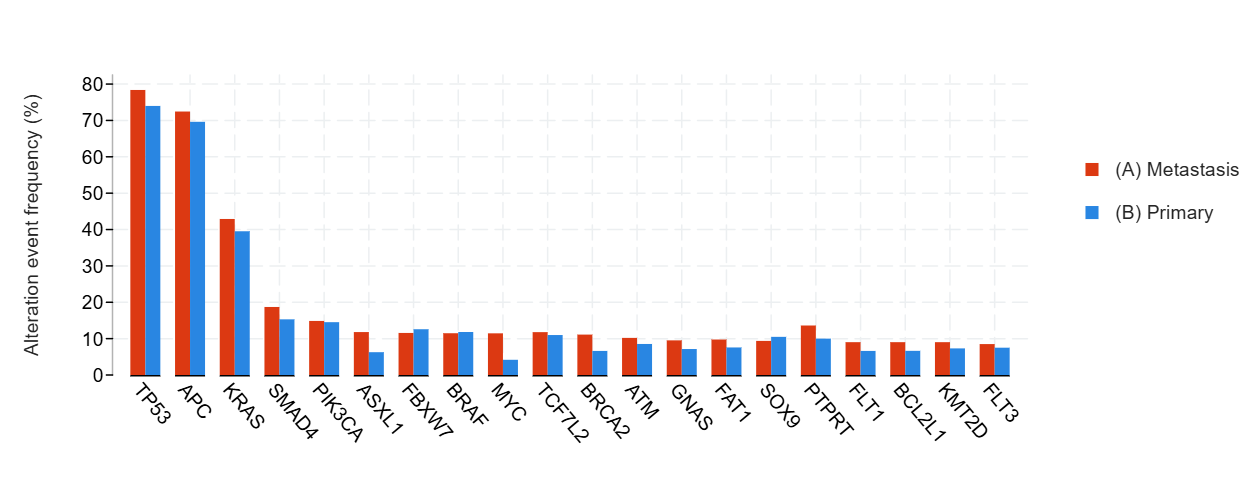

**Figure S6.** Distribution of the Most Frequently Mutated Genes in Primary and Metastatic Colorectal Cancer

Table S6. **Gene-Level Mutation Frequencies in Primary Versus Metastatic Colorectal Cancer**

| **Gene** | **Cytoband** | **(A) Metastasis** | **(B) Primary** | **Co-occurrence Pattern** | **Log2 Ratio** | **p-Value** | **q-Value** | **Enriched in** |
| --- | --- | --- | --- | --- | --- | --- | --- | --- |
| **TP53** | 17p13.1 | 232 (78.38%) | 219 (73.99%) |  | 0.08 | 0.247 | 1.00 | (A) Metastasis |
| **APC** | 5q22.2 | 213 (72.45%) | 204 (69.62%) |  | 0.06 | 0.468 | 1.00 | (A) Metastasis |
| **KRAS** | 12p12.1 | 127 (42.91%) | 117 (39.53%) |  | 0.12 | 0.452 | 1.00 | (A) Metastasis |
| **SMAD4** | 18q21.2 | 55 (18.71%) | 45 (15.31%) |  | 0.29 | 0.323 | 1.00 | (A) Metastasis |
| **PIK3CA** | 3q26.32 | 44 (14.86%) | 43 (14.53%) |  | 0.03 | 1.00 | 1.00 | (A) Metastasis |
| **ASXL1** | 20q11.21 | 34 (11.81%) | 18 (6.27%) |  | 0.91 | 0.0284 | 1.00 | (A) Metastasis |
| **FBXW7** | 4q31.3 | 34 (11.56%) | 37 (12.59%) |  | -0.12 | 0.800 | 1.00 | (B) Primary |
| **BRAF** | 7q34 | 34 (11.49%) | 35 (11.82%) |  | -0.04 | 1.00 | 1.00 | (B) Primary |
| **MYC** | 8q24.21 | 33 (11.46%) | 12 (4.18%) |  | 1.45. | 1,63E+00 | 0.352 | (A) Metastasis |
| **TCF7L2** | 10q25.2-q25.3 | 33 (11.79%) | 30 (10.99%) |  | 0.10 | 0.790 | 1.00 | (A) Metastasis |
| **BRCA2** | 13q13.1 | 32 (11.11%) | 19 (6.62%) |  | 0.75 | 0.0775 | 1.00 | (A) Metastasis |
| **ATM** | 11q22.3 | 30 (10.20%) | 25 (8.53%) |  | 0.26 | 0.571 | 1.00 | (A) Metastasis |
| **GNAS** | 20q13.32 | 28 (9.52%) | 21 (7.14%) |  | 0.42 | 0.371 | 1.00 | (A) Metastasis |
| **FAT1** | 4q35.2 | 28 (9.76%) | 20 (7.58%) |  | 0.36 | 0.450 | 1.00 | (A) Metastasis |
| **SOX9** | 17q24.3 | 27 (9.38%) | 30 (10.49%) |  | -0.16 | 0.678 | 1.00 | (B) Primary |
| **PTPRT** | 20q12-q13.11 | 26 (13.61%) | 19 (10.00%) |  | 0.44 | 0.341 | 1.00 | (A) Metastasis |
| **FLT1** | 13q12.3 | 26 (9.03%) | 19 (6.62%) |  | 0.45 | 0.352 | 1.00 | (A) Metastasis |
| **BCL2L1** | 20q11.21 | 26 (9.03%) | 19 (6.64%) |  | 0.44 | 0.352 | 1.00 | (A) Metastasis |
| **KMT2D** | 12q13.12 | 26 (9.03%) | 21 (7.32%) |  | 0.30 | 0.543 | 1.00 | (A) Metastasis |
| **FLT3** | 13q12.2 | 25 (8.50%) | 22 (7.51%) |  | 0.18 | 0.761 | 1.00 | (A) Metastasis |
| **RECQL4** | 8q24.3 | 24 (8.45%) | 5 (1.79%) |  | 1.24. | 4,00E-01 | 0.259 | (A) Metastasis |
| **SMAD2** | 18q21.1 | 24 (8.33%) | 15 (5.23%) |  | 0.67 | 0.184 | 1.00 | (A) Metastasis |
| **PTEN** | 10q23.31 | 24 (8.11%) | 18 (6.08%) |  | 0.42 | 0.424 | 1.00 | (A) Metastasis |
| **CDK8** | 13q12.13 | 23 (8.01%) | 16 (6.04%) |  | 0.41 | 0.408 | 1.00 | (A) Metastasis |
| **AURKA** | 20q13.2 | 21 (7.29%) | 12 (4.18%) |  | 0.80 | 0.151 | 1.00 | (A) Metastasis |
| **ERBB2** | 17q12 | 21 (7.09%) | 29 (9.80%) |  | -0.47 | 0.301 | 1.00 | (B) Primary |
| **RNF43** | 17q22 | 21 (7.32%) | 21 (7.92%) |  | -0.12 | 0.873 | 1.00 | (B) Primary |
| **AGO2** | 8q24.3 | 20 (11.63%) | 6 (3.85%) |  | 1.6. | 0.0128 | 1.00 | (A) Metastasis |
| **NKX3-1** | 8p21.2 | 20 (7.07%) | 7 (2.68%) |  | 1.4. | 0.0280 | 1.00 | (A) Metastasis |
| **PREX2** | 8q13.2 | 20 (11.36%) | 10 (6.29%) |  | 0.85 | 0.126 | 1.00 | (A) Metastasis |
| **FGFR1** | 8p11.23 | 20 (6.76%) | 15 (5.07%) |  | 0.42 | 0.486 | 1.00 | (A) Metastasis |
| **ARID1A** | 1p36.11 | 20 (6.94%) | 17 (5.92%) |  | 0.23 | 0.734 | 1.00 | (A) Metastasis |
| **DIS3** | 13q21.33 | 19 (6.60%) | 13 (4.53%) |  | 0.54 | 0.363 | 1.00 | (A) Metastasis |
| **ERBB4** | 2q34 | 19 (6.46%) | 14 (4.76%) |  | 0.44 | 0.474 | 1.00 | (A) Metastasis |
| **NRAS** | 1p13.2 | 19 (6.42%) | 19 (6.42%) |  | - | 1.00 | 1.00 | (B) Primary |
| **NSD3** | 8p11.23 | 18 (6.82%) | 12 (5.29%) |  | 0.37 | 0.572 | 1.00 | (A) Metastasis |
| **ROS1** | 6q22.1 | 18 (6.21%) | 15 (5.19%) |  | 0.26 | 0.721 | 1.00 | (A) Metastasis |
| **SRC** | 20q11.23 | 17 (8.42%) | 8 (3.60%) |  | 1.22. | 0.0401 | 1.00 | (A) Metastasis |
| **CARD11** | 7p22.2 | 17 (5.90%) | 9 (3.14%) |  | 0.91 | 0.159 | 1.00 | (A) Metastasis |
| **NOTCH3** | 19p13.12 | 17 (5.92%) | 9 (3.40%) |  | 0.80 | 0.227 | 1.00 | (A) Metastasis |

### **Table S7. Prostate Cancer Demographic and Survival Characteristics**

| Section | Characteristic | Level | N | Percent | Median / Q1–Q3 |
| --- | --- | --- | --- | --- | --- |
| Overall | All patients |  | 210 | 100 |  |
| Age | Age at sequencing (years) |  |  |  | 67.0 (61.0–74.0) |
| Sex | Sex | Male | 210 | 100 |  |
|  |  | Female | 0 | 0 |  |
|  |  | Unknown | 0 | 0 |  |
| Ethnicity | Ethnicity | Non-Spanish/non-Hispanic | 170 | 81.0 |  |
|  |  | Spanish/Hispanic | 12 | 5.7 |  |
|  |  | Unknown | 23 | 11.0 |  |
|  |  | Not Collected | 5 | 2.3 |  |
| Race | Race | White | 152 | 72.4 |  |
|  |  | Black | 18 | 8.6 |  |
|  |  | Asian | 9 | 4.3 |  |
|  |  | Other | 6 | 2.9 |  |
|  |  | Unknown | 25 | 11.9 |  |
| Survival | Median survival after sequencing (years) |  |  |  | 2.3 |
| Survival | Median age at death (years) |  |  |  | 73.4 |

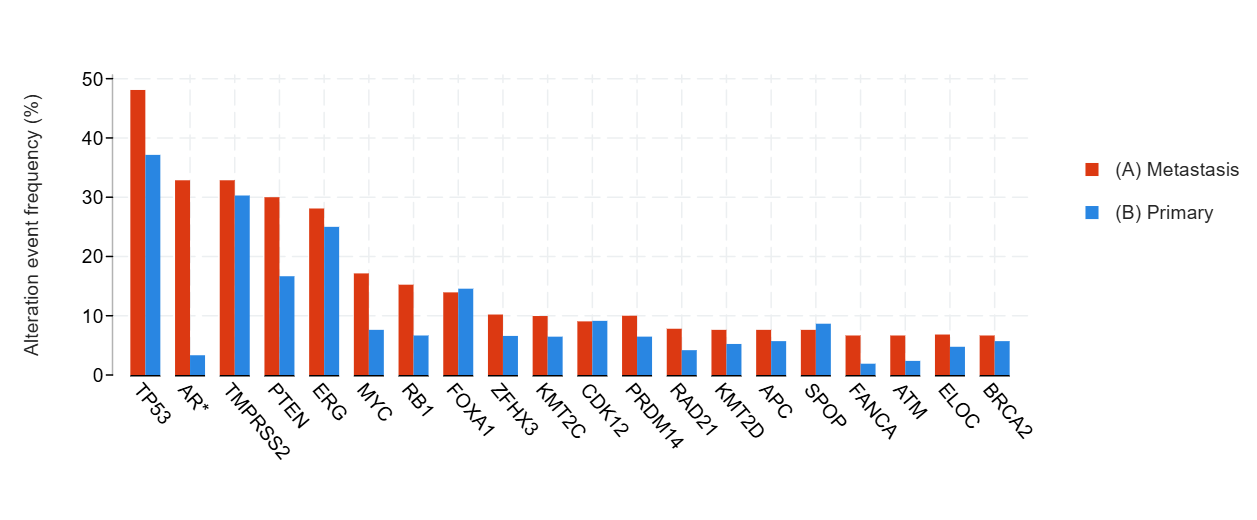
**Figure S7.** Distribution of the Most Frequently Mutated Genes in Primary and Metastatic Prostate Cancer

**Table S8.** **Gene-Level Mutation Frequencies in Primary Versus Metastatic Prostate Cancer**

| **Gene** | **Cytoband** | **(A) Metastasis** | **(B) Primary** | **Co-occurrence Pattern** | **Log2 Ratio** | **p-Value** | **q-Value** | **Enriched in** |
| --- | --- | --- | --- | --- | --- | --- | --- | --- |
| **TP53** | 17p13.1 | 101 (48.10%) | 78 (37.14%) |  | 0.37 | 0.0298 | 1.00 | (A) Metastasis |
| **AR** | Xq12 | 69 (32.86%) | 7 (3.33%) |  | Mar.30 | **< 10-10** | **< 10-10** | (A) Metastasis |
| **TMPRSS2** | 21q22.3 | 69 (32.86%) | 63 (30.29%) |  | 0.12 | 0.600 | 1.00 | (A) Metastasis |
| **PTEN** | 10q23.31 | 63 (30.00%) | 35 (16.67%) |  | 0.85 | 1,75E+00 | 0.457 | (A) Metastasis |
| **ERG** | 21q22.2 | 59 (28.10%) | 52 (25.00%) |  | 0.17 | 0.507 | 1.00 | (A) Metastasis |
| **MYC** | 8q24.21 | 36 (17.14%) | 16 (7.62%) |  | Oca.17 | 4,51E+00 | 0.783 | (A) Metastasis |
| **RB1** | 13q14.2 | 32 (15.24%) | 14 (6.67%) |  | Oca.19 | 7,35E+00 | 0.958 | (A) Metastasis |
| **FOXA1** | 14q21.1 | 29 (13.94%) | 30 (14.56%) |  | -0.06 | 0.889 | 1.00 | (B) Primary |
| **ZFHX3** | 16q22.2-q22.3 | 20 (10.20%) | 12 (6.59%) |  | 0.63 | 0.267 | 1.00 | (A) Metastasis |
| **KMT2C** | 7q36.1 | 20 (9.95%) | 13 (6.47%) |  | 0.62 | 0.275 | 1.00 | (A) Metastasis |
| **CDK12** | 17q12 | 19 (9.05%) | 19 (9.13%) |  | -0.01 | 1.00 | 1.00 | (B) Primary |
| **PRDM14** | 8q13.3 | 17 (10.00%) | 9 (6.47%) |  | 0.63 | 0.308 | 1.00 | (A) Metastasis |
| **RAD21** | 8q24.11 | 16 (7.80%) | 8 (4.19%) |  | 0.90 | 0.145 | 1.00 | (A) Metastasis |
| **KMT2D** | 12q13.12 | 16 (7.62%) | 11 (5.24%) |  | 0.54 | 0.427 | 1.00 | (A) Metastasis |
| **APC** | 5q22.2 | 16 (7.62%) | 12 (5.71%) |  | 0.42 | 0.558 | 1.00 | (A) Metastasis |
| **SPOP** | 17q21.33 | 16 (7.62%) | 18 (8.65%) |  | -0.18 | 0.724 | 1.00 | (B) Primary |
| **FANCA** | 16q24.3 | 14 (6.67%) | 4 (1.90%) |  | Oca.81 | 0.0169 | 1.00 | (A) Metastasis |
| **ATM** | 11q22.3 | 14 (6.67%) | 5 (2.38%) |  | Oca.49 | 0.0577 | 1.00 | (A) Metastasis |
| **ELOC** | 8q21.11 | 14 (6.83%) | 9 (4.76%) |  | 0.52 | 0.400 | 1.00 | (A) Metastasis |
| **BRCA2** | 13q13.1 | 14 (6.67%) | 12 (5.71%) |  | 0.22 | 0.840 | 1.00 | (A) Metastasis |
| **JAK1** | 1p31.3 | 13 (6.19%) | 5 (2.40%) |  | Oca.36 | 0.0892 | 1.00 | (A) Metastasis |
| **CIC** | 19q13.2 | 13 (6.19%) | 7 (3.37%) |  | 0.88 | 0.252 | 1.00 | (A) Metastasis |
| **MGA** | 15q15.1 | 12 (5.88%) | 5 (2.66%) |  | Oca.15 | 0.140 | 1.00 | (A) Metastasis |
| **PREX2** | 8q13.2 | 12 (6.94%) | 6 (4.23%) |  | 0.72 | 0.339 | 1.00 | (A) Metastasis |
| **ANKRD11** | 16q24.3 | 12 (6.15%) | 7 (3.87%) |  | 0.67 | 0.353 | 1.00 | (A) Metastasis |
| **PIK3CA** | 3q26.32 | 12 (5.71%) | 13 (6.19%) |  | -0.12 | 1.00 | 1.00 | (B) Primary |
| **AMER1** | Xq11.2 | 11 (5.47%) | 2 (1.00%) |  | Şub.46 | 0.0204 | 1.00 | (A) Metastasis |
| **IRS2** | 13q34 | 11 (5.47%) | 3 (1.49%) |  | Oca.87 | 0.0531 | 1.00 | (A) Metastasis |
| **NCOR1** | 17p12-p11.2 | 11 (5.47%) | 3 (1.49%) |  | Oca.87 | 0.0531 | 1.00 | (A) Metastasis |
| **NBN** | 8q21.3 | 11 (5.24%) | 8 (3.81%) |  | 0.46 | 0.640 | 1.00 | (A) Metastasis |
| **RECQL4** | 8q24.3 | 11 (5.29%) | 12 (5.80%) |  | -0.13 | 0.834 | 1.00 | (B) Primary |
| **CDKN1B** | 12p13.1 | 10 (4.76%) | 3 (1.43%) |  | Oca.74 | 0.0873 | 1.00 | (A) Metastasis |
| **INPPL1** | 11q13.4 | 10 (5.88%) | 3 (2.16%) |  | Oca.45 | 0.154 | 1.00 | (A) Metastasis |
| **CTNNB1** | 3p22.1 | 10 (4.76%) | 4 (1.90%) |  | Oca.32 | 0.172 | 1.00 | (A) Metastasis |
| **PTPRS** | 19p13.3 | 10 (5.05%) | 5 (2.53%) |  | 1.00 | 0.292 | 1.00 | (A) Metastasis |
| **SPEN** | 1p36.21-p36.13 | 10 (4.98%) | 5 (2.49%) |  | 1.00 | 0.292 | 1.00 | (A) Metastasis |
| **CDH1** | 16q22.1 | 10 (4.76%) | 6 (2.86%) |  | 0.74 | 0.446 | 1.00 | (A) Metastasis |
| **KMT2A** | 11q23.3 | 10 (4.76%) | 7 (3.33%) |  | 0.51 | 0.622 | 1.00 | (A) Metastasis |
| **FAT1** | 4q35.2 | 10 (4.76%) | 9 (4.33%) |  | 0.14 | 1.00 | 1.00 | (A) Metastasis |
| **PTPRD** | 9p24.1-p23 | 9 (4.48%) | 3 (1.49%) |  | Oca.59 | 0.0871 | 1.00 | (A) Metastasis |

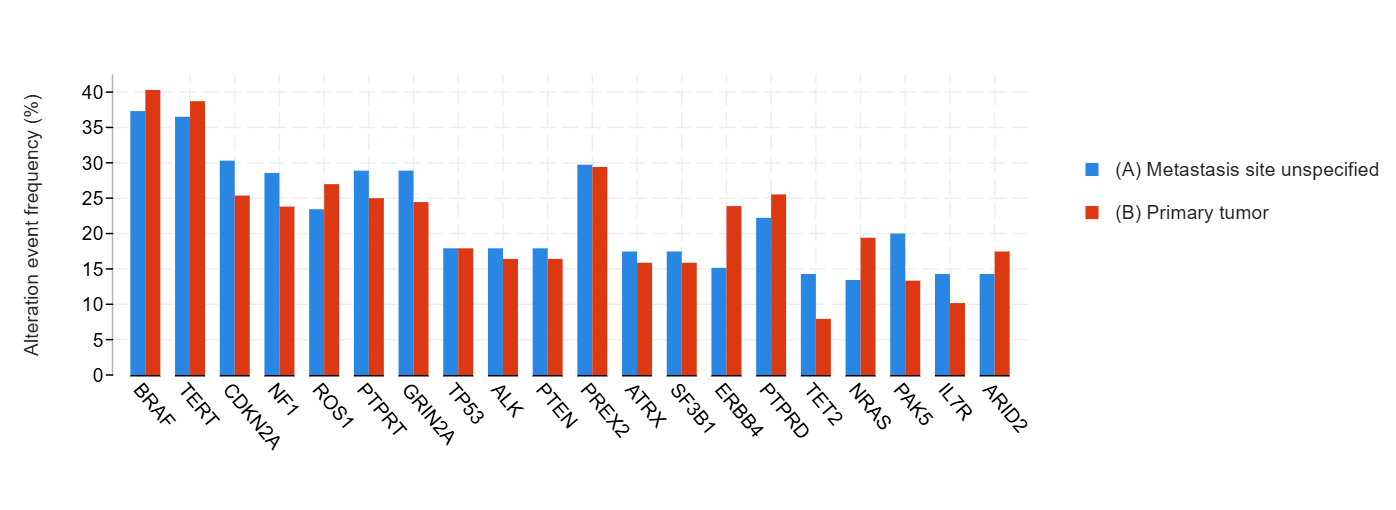

**Figure S8.** Distribution of the Most Frequently Mutated Genes in Primary and Metastatic Melanoma

**Table S9.** **Gene-Level Mutation Frequencies in Primary Versus Metastatic Melanoma**

| **Gene** | **Cyt**  **oband** | **(A) Metastasis site ...** | **(B) Primary tumor** | **Co-occurrence Pattern** | **Log2 Ratio** | **p-Value** | **q-Value** | **Enriched in** |
| --- | --- | --- | --- | --- | --- | --- | --- | --- |
| **BRAF** | 7q34 | 25 (37.31%) | 27 (40.30%) |  | -0.11 | 0.859 | 1.00 | (B) Primary tumor |
| **TERT** | 5p15.33 | 23 (36.51%) | 24 (38.71%) |  | -0.08 | 0.855 | 1.00 | (B) Primary tumor |
| **CDKN2A** | 9p21.3 | 20 (30.30%) | 17 (25.37%) |  | 0.26 | 0.566 | 1.00 | (A) Metastasis site ... |
| **NF1** | 17q11.2 | 18 (28.57%) | 15 (23.81%) |  | 0.26 | 0.686 | 1.00 | (A) Metastasis site ... |
| **ROS1** | 6q22.1 | 15 (23.44%) | 17 (26.98%) |  | -0.20 | 0.687 | 1.00 | (B) Primary tumor |
| **PTPRT** | 20q12-q13.11 | 13 (28.89%) | 11 (25.00%) |  | 0.21 | 0.812 | 1.00 | (A) Metastasis site ... |
| **GRIN2A** | 16p13.2 | 13 (28.89%) | 11 (24.44%) |  | 0.24 | 0.812 | 1.00 | (A) Metastasis site ... |
| **TP53** | 17p13.1 | 12 (17.91%) | 12 (17.91%) |  | - | 1.00 | 1.00 | (B) Primary tumor |
| **ALK** | 2p23.2-p23.1 | 12 (17.91%) | 11 (16.42%) |  | 0.13 | 1.00 | 1.00 | (A) Metastasis site ... |
| **PTEN** | 10q23.31 | 12 (17.91%) | 11 (16.42%) |  | 0.13 | 1.00 | 1.00 | (A) Metastasis site ... |
| **PREX2** | 8q13.2 | 11 (29.73%) | 10 (29.41%) |  | 0.02 | 1.00 | 1.00 | (A) Metastasis site ... |
| **ATRX** | Xq21.1 | 11 (17.46%) | 10 (15.87%) |  | 0.14 | 1.00 | 1.00 | (A) Metastasis site ... |
| **SF3B1** | 2q33.1 | 11 (17.46%) | 10 (15.87%) |  | 0.14 | 1.00 | 1.00 | (A) Metastasis site ... |
| **ERBB4** | 2q34 | 10 (15.15%) | 16 (23.88%) |  | -0.66 | 0.275 | 1.00 | (B) Primary tumor |
| **PTPRD** | 9p24.1-p23 | 10 (22.22%) | 12 (25.53%) |  | -0.20 | 0.809 | 1.00 | (B) Primary tumor |
| **TET2** | 4q24 | 9 (14.29%) | 5 (7.94%) |  | 0.85 | 0.396 | 1.00 | (A) Metastasis site ... |
| **NRAS** | 1p13.2 | 9 (13.43%) | 13 (19.40%) |  | -0.53 | 0.485 | 1.00 | (B) Primary tumor |
| **PAK5** | 20p12.2 | 9 (20.00%) | 6 (13.33%) |  | 0.58 | 0.573 | 1.00 | (A) Metastasis site ... |
| **IL7R** | 5p13.2 | 9 (14.29%) | 6 (10.17%) |  | 0.49 | 0.586 | 1.00 | (A) Metastasis site ... |
| **ARID2** | 12q12 | 9 (14.29%) | 11 (17.46%) |  | -0.29 | 0.808 | 1.00 | (B) Primary tumor |
| **FGFR2** | 10q26.13 | 9 (13.43%) | 9 (13.43%) |  | - | 1.00 | 1.00 | (B) Primary tumor |
| **MET** | 7q31.2 | 9 (13.43%) | 9 (13.43%) |  | - | 1.00 | 1.00 | (B) Primary tumor |
| **RAD21** | 8q24.11 | 8 (12.70%) | 2 (3.39%) |  | Oca.91 | 0.0968 | 1.00 | (A) Metastasis site ... |
| **NOTCH2** | 1p12 | 8 (12.70%) | 3 (4.76%) |  | Oca.42 | 0.205 | 1.00 | (A) Metastasis site ... |
| **TP63** | 3q28 | 8 (17.78%) | 4 (9.09%) |  | 0.97 | 0.353 | 1.00 | (A) Metastasis site ... |
| **FAT1** | 4q35.2 | 8 (12.70%) | 10 (16.95%) |  | -0.42 | 0.612 | 1.00 | (B) Primary tumor |
| **KMT2A** | 11q23.3 | 8 (12.70%) | 6 (9.52%) |  | 0.42 | 0.778 | 1.00 | (A) Metastasis site ... |
| **ATM** | 11q22.3 | 8 (12.12%) | 8 (11.94%) |  | 0.02 | 1.00 | 1.00 | (A) Metastasis site ... |
| **KDR** | 4q12 | 8 (12.12%) | 8 (11.94%) |  | 0.02 | 1.00 | 1.00 | (A) Metastasis site ... |
| **CDKN2B** | 9p21.3 | 8 (12.70%) | 9 (14.29%) |  | -0.17 | 1.00 | 1.00 | (B) Primary tumor |
| **CARD11** | 7p22.2 | 8 (12.70%) | 8 (12.70%) |  | - | 1.00 | 1.00 | (B) Primary tumor |
| **KMT2D** | 12q13.12 | 7 (11.11%) | 4 (6.35%) |  | 0.81 | 0.530 | 1.00 | (A) Metastasis site ... |
| **FLT1** | 13q12.3 | 7 (11.11%) | 9 (14.29%) |  | -0.36 | 0.606 | 1.00 | (B) Primary tumor |
| **PDGFRA** | 4q12 | 7 (10.61%) | 5 (7.69%) |  | 0.46 | 0.763 | 1.00 | (A) Metastasis site ... |
| **EPHA7** | 6q16.1 | 7 (15.91%) | 6 (13.04%) |  | 0.29 | 0.770 | 1.00 | (A) Metastasis site ... |
| **ATR** | 3q23 | 7 (11.11%) | 7 (11.86%) |  | -0.09 | 1.00 | 1.00 | (B) Primary tumor |
| **MYC** | 8q24.21 | 6 (9.52%) | 2 (3.17%) |  | Oca.58 | 0.273 | 1.00 | (A) Metastasis site ... |
| **BRCA1** | 17q21.31 | 6 (9.52%) | 3 (4.76%) |  | 1.00 | 0.491 | 1.00 | (A) Metastasis site ... |
| **NOTCH3** | 19p13.12 | 6 (9.52%) | 8 (13.56%) |  | -0.51 | 0.575 | 1.00 | (B) Primary tumor |
| **FLT3** | 13q12.2 | 6 (9.09%) | 6 (8.96%) |  | 0.02 | 1.00 | 1.00 | (A) Metastasis site ... |
| **PIK3C2G** | 12p12.3 | 6 (13.33%) | 5 (11.11%) |  | 0.26 | 1.00 | 1.00 | (A) Metastasis site ... |
| **ARID1A** | 1p36.11 | 6 (9.52%) | 6 (9.52%) |  | - | 1.00 | 1.00 | (B) Primary tumor |
| **DNMT3A** | 2p23.3 | 6 (9.52%) | 6 (9.52%) |  | - | 1.00 | 1.00 | (B) Primary tumor |
| **NOTCH1** | 9q34.3 | 6 (8.96%) | 5 (7.46%) |  | 0.26 | 1.00 | 1.00 | (A) Metastasis site ... |
| **GLI1** | 12q13.3 | 6 (9.68%) | 5 (8.33%) |  | 0.22 | 1.00 | 1.00 | (A) Metastasis site ... |
| **NOTCH4** | 6p21.32 | 6 (13.33%) | 6 (13.33%) |  | - | 1.00 | 1.00 | (B) Primary tumor |
| **MGA** | 15q15.1 | 6 (9.68%) | 5 (9.09%) |  | 0.09 | 1.00 | 1.00 | (A) Metastasis site ... |
| **POLD1** | 19q13.33 | 5 (8.06%) | 1 (1.79%) |  | Şub.18 | 0.210 | 1.00 | (A) Metastasis site ... |
| **NSD1** | 5q35.3 | 5 (8.06%) | 8 (13.56%) |  | -0.75 | 0.388 | 1.00 | (B) Primary tumor |
| **KDM5A** | 12p13.33 | 5 (8.06%) | 2 (3.39%) |  | Oca.25 | 0.440 | 1.00 | (A) Metastasis site ... |
| **APC** | 5q22.2 | 5 (7.58%) | 8 (11.94%) |  | -0.66 | 0.561 | 1.00 | (B) Primary tumor |
| **ERG** | 21q22.2 | 5 (7.94%) | 3 (5.08%) |  | 0.64 | 0.718 | 1.00 | (A) Metastasis site ... |
| **SMO** | 7q32.1 | 5 (7.58%) | 4 (5.97%) |  | 0.34 | 0.744 | 1.00 | (A) Metastasis site ... |
| **EGFR** | 7p11.2 | 5 (7.46%) | 7 (10.45%) |  | -0.49 | 0.764 | 1.00 | (B) Primary tumor |
| **EZH2** | 7q36.1 | 5 (7.69%) | 4 (6.15%) |  | 0.32 | 1.00 | 1.00 | (A) Metastasis site ... |
| **ASXL1** | 20q11.21 | 5 (7.94%) | 5 (7.94%) |  | - | 1.00 | 1.00 | (B) Primary tumor |
| **BRCA2** | 13q13.1 | 5 (7.94%) | 5 (7.94%) |  | - | 1.00 | 1.00 | (B) Primary tumor |
| **GNAQ** | 9q21.2 | 5 (7.46%) | 5 (7.46%) |  | - | 1.00 | 1.00 | (B) Primary tumor |
| **ZFHX3** | 16q22.2-q22.3 | 5 (11.36%) | 5 (12.20%) |  | -0.10 | 1.00 | 1.00 | (B) Primary tumor |
| **MST1R** | 3p21.31 | 5 (11.36%) | 4 (9.52%) |  | 0.25 | 1.00 | 1.00 | (A) Metastasis site ... |

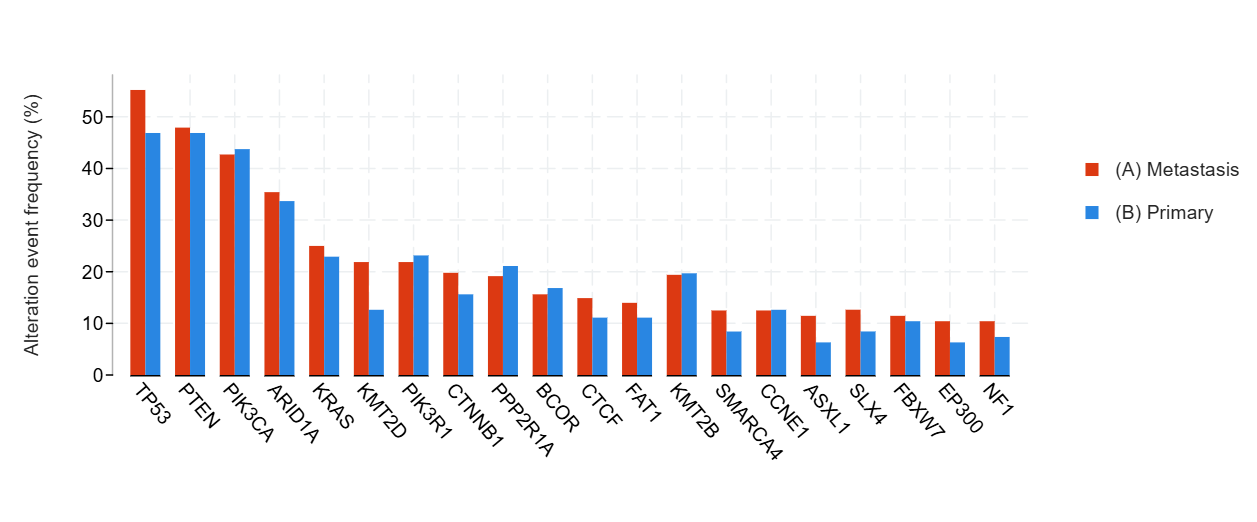

**Figure S9.** Distribution of the Most Frequently Mutated Genes in Primary and Metastatic Endometrial Cancer

**Table S10.** **Gene-Level Mutation Frequencies in Primary Versus Metastatic Endometrial Cancer**

| **Gene** | **Cytoband** | **(A) Metastasis** | **(B) Primary** | **Co-occurrence Pattern** | **Log2 Ratio** | **p-Value** | **q-Value** | **Enriched in** |
| --- | --- | --- | --- | --- | --- | --- | --- | --- |
| **TP53** | 17p13.1 | 53 (55.21%) | 45 (46.88%) |  | 0.24 | 0.312 | 1.00 | (A) Metastasis |
| **PTEN** | 10q23.31 | 46 (47.92%) | 45 (46.88%) |  | 0.03 | 1.00 | 1.00 | (A) Metastasis |
| **PIK3CA** | 3q26.32 | 41 (42.71%) | 42 (43.75%) |  | -0.03 | 1.00 | 1.00 | (B) Primary |
| **ARID1A** | 1p36.11 | 34 (35.42%) | 32 (33.68%) |  | 0.07 | 0.879 | 1.00 | (A) Metastasis |
| **KRAS** | 12p12.1 | 24 (25.00%) | 22 (22.92%) |  | 0.13 | 0.866 | 1.00 | (A) Metastasis |
| **KMT2D** | 12q13.12 | 21 (21.88%) | 12 (12.63%) |  | 0.79 | 0.125 | 1.00 | (A) Metastasis |
| **PIK3R1** | 5q13.1 | 21 (21.88%) | 22 (23.16%) |  | -0.08 | 0.864 | 1.00 | (B) Primary |
| **CTNNB1** | 3p22.1 | 19 (19.79%) | 15 (15.63%) |  | 0.34 | 0.571 | 1.00 | (A) Metastasis |
| **PPP2R1A** | 19q13.41 | 18 (19.15%) | 19 (21.11%) |  | -0.14 | 0.854 | 1.00 | (B) Primary |
| **BCOR** | Xp11.4 | 15 (15.63%) | 16 (16.84%) |  | -0.11 | 0.847 | 1.00 | (B) Primary |
| **CTCF** | 16q22.1 | 14 (14.89%) | 10 (11.11%) |  | 0.42 | 0.515 | 1.00 | (A) Metastasis |
| **FAT1** | 4q35.2 | 13 (13.98%) | 10 (11.11%) |  | 0.33 | 0.658 | 1.00 | (A) Metastasis |
| **KMT2B** | 19q13.12 | 13 (19.40%) | 13 (19.70%) |  | -0.02 | 1.00 | 1.00 | (B) Primary |
| **SMARCA4** | 19p13.2 | 12 (12.50%) | 8 (8.42%) |  | 0.57 | 0.479 | 1.00 | (A) Metastasis |
| **CCNE1** | 19q12 | 12 (12.50%) | 12 (12.63%) |  | -0.02 | 1.00 | 1.00 | (B) Primary |
| **ASXL1** | 20q11.21 | 11 (11.46%) | 6 (6.32%) |  | 0.86 | 0.310 | 1.00 | (A) Metastasis |
| **SLX4** | 16p13.3 | 11 (12.64%) | 7 (8.43%) |  | 0.58 | 0.458 | 1.00 | (A) Metastasis |
| **FBXW7** | 4q31.3 | 11 (11.46%) | 10 (10.42%) |  | 0.14 | 1.00 | 1.00 | (A) Metastasis |
| **EP300** | 22q13.2 | 10 (10.42%) | 6 (6.32%) |  | 0.72 | 0.434 | 1.00 | (A) Metastasis |
| **NF1** | 17q11.2 | 10 (10.42%) | 7 (7.37%) |  | 0.50 | 0.613 | 1.00 | (A) Metastasis |
| **TERT** | 5p15.33 | 10 (10.42%) | 7 (7.37%) |  | 0.50 | 0.613 | 1.00 | (A) Metastasis |
| **NOTCH1** | 9q34.3 | 10 (10.42%) | 7 (7.29%) |  | 0.51 | 0.613 | 1.00 | (A) Metastasis |
| **MAP3K1** | 5q11.2 | 9 (9.38%) | 5 (5.26%) |  | 0.83 | 0.406 | 1.00 | (A) Metastasis |
| **NSD2** | 4p16.3 | 9 (10.23%) | 5 (6.02%) |  | 0.76 | 0.407 | 1.00 | (A) Metastasis |
| **JAK1** | 1p31.3 | 9 (9.57%) | 5 (5.56%) |  | 0.79 | 0.407 | 1.00 | (A) Metastasis |
| **ZFHX3** | 16q22.2-q22.3 | 9 (12.50%) | 8 (10.96%) |  | 0.19 | 0.802 | 1.00 | (A) Metastasis |
| **ARID1B** | 6q25.3 | 9 (9.47%) | 10 (10.53%) |  | -0.15 | 1.00 | 1.00 | (B) Primary |
| **BRCA2** | 13q13.1 | 9 (9.38%) | 8 (8.42%) |  | 0.15 | 1.00 | 1.00 | (A) Metastasis |
| **APC** | 5q22.2 | 9 (9.38%) | 8 (8.33%) |  | 0.17 | 1.00 | 1.00 | (A) Metastasis |
| **ATM** | 11q22.3 | 9 (9.38%) | 8 (8.33%) |  | 0.17 | 1.00 | 1.00 | (A) Metastasis |
| **ERBB2** | 17q12 | 9 (9.38%) | 8 (8.33%) |  | 0.17 | 1.00 | 1.00 | (A) Metastasis |
| **STK11** | 19p13.3 | 8 (8.33%) | 3 (3.13%) |  | Oca.42 | 0.213 | 1.00 | (A) Metastasis |
| **CREBBP** | 16p13.3 | 8 (8.33%) | 4 (4.21%) |  | 0.98 | 0.372 | 1.00 | (A) Metastasis |
| **CDK12** | 17q12 | 8 (8.51%) | 4 (4.44%) |  | 0.94 | 0.373 | 1.00 | (A) Metastasis |
| **SOX17** | 8q11.23 | 8 (10.96%) | 5 (6.85%) |  | 0.68 | 0.563 | 1.00 | (A) Metastasis |
| **KMT2A** | 11q23.3 | 8 (8.33%) | 5 (5.26%) |  | 0.66 | 0.567 | 1.00 | (A) Metastasis |
| **AKT1** | 14q32.33 | 8 (8.33%) | 6 (6.25%) |  | 0.42 | 0.782 | 1.00 | (A) Metastasis |
| **MED12** | Xq13.1 | 8 (8.51%) | 9 (10.00%) |  | -0.23 | 0.802 | 1.00 | (B) Primary |
| **ERBB3** | 12q13.2 | 8 (8.33%) | 9 (9.47%) |  | -0.19 | 0.805 | 1.00 | (B) Primary |
| **INPPL1** | 11q13.4 | 8 (11.94%) | 8 (12.12%) |  | -0.02 | 1.00 | 1.00 | (B) Primary |
| **PTPRS** | 19p13.3 | 7 (9.59%) | 3 (4.11%) |  | Oca.22 | 0.326 | 1.00 | (A) Metastasis |
| **PTPRT** | 20q12-q13.11 | 7 (9.46%) | 3 (4.11%) |  | Oca.20 | 0.327 | 1.00 | (A) Metastasis |
| **FGFR1** | 8p11.23 | 7 (7.29%) | 3 (3.13%) |  | Oca.22 | 0.331 | 1.00 | (A) Metastasis |
| **MYC** | 8q24.21 | 7 (7.29%) | 4 (4.21%) |  | 0.79 | 0.537 | 1.00 | (A) Metastasis |
| **TSC2** | 16p13.3 | 7 (7.29%) | 4 (4.21%) |  | 0.79 | 0.537 | 1.00 | (A) Metastasis |
| **ERBB4** | 2q34 | 7 (7.29%) | 4 (4.17%) |  | 0.81 | 0.537 | 1.00 | (A) Metastasis |
| **AXIN2** | 17q24.1 | 7 (7.53%) | 4 (4.44%) |  | 0.76 | 0.537 | 1.00 | (A) Metastasis |
| **SPEN** | 1p36.21-p36.13 | 7 (9.46%) | 5 (6.85%) |  | 0.47 | 0.765 | 1.00 | (A) Metastasis |
| **MTOR** | 1p36.22 | 7 (7.29%) | 5 (5.26%) |  | 0.47 | 0.767 | 1.00 | (A) Metastasis |
| **NTRK1** | 1q23.1 | 7 (7.29%) | 5 (5.26%) |  | 0.47 | 0.767 | 1.00 | (A) Metastasis |
| **PTCH1** | 9q22.32 | 7 (7.29%) | 5 (5.26%) |  | 0.47 | 0.767 | 1.00 | (A) Metastasis |
| **ATR** | 3q23 | 7 (7.45%) | 5 (5.56%) |  | 0.42 | 0.767 | 1.00 | (A) Metastasis |
| **FGFR2** | 10q26.13 | 7 (7.29%) | 9 (9.38%) |  | -0.36 | 0.795 | 1.00 | (B) Primary |
| **RB1** | 13q14.2 | 7 (7.29%) | 9 (9.38%) |  | -0.36 | 0.795 | 1.00 | (B) Primary |
| **BRD4** | 19p13.12 | 7 (7.37%) | 6 (6.32%) |  | 0.22 | 1.00 | 1.00 | (A) Metastasis |
| **IRS2** | 13q34 | 7 (9.46%) | 6 (8.22%) |  | 0.20 | 1.00 | 1.00 | (A) Metastasis |
| **SPOP** | 17q21.33 | 7 (7.45%) | 6 (6.67%) |  | 0.16 | 1.00 | 1.00 | (A) Metastasis |
| **CCND1** | 11q13.3 | 7 (7.29%) | 7 (7.37%) |  | -0.02 | 1.00 | 1.00 | (B) Primary |
| **DICER1** | 14q32.13 | 7 (7.29%) | 7 (7.37%) |  | -0.02 | 1.00 | 1.00 | (B) Primary |
| **CEBPA** | 19q13.11 | 6 (6.32%) | 2 (2.11%) |  | Oca.58 | 0.279 | 1.00 | (A) Metastasis |
| **KDM6A** | Xp11.3 | 6 (6.25%) | 2 (2.11%) |  | Oca.57 | 0.279 | 1.00 | (A) Metastasis |
| **KEAP1** | 19p13.2 | 6 (6.32%) | 3 (3.19%) |  | 0.98 | 0.497 | 1.00 | (A) Metastasis |
| **ZRSR2** | Xp22.2 | 6 (6.32%) | 3 (3.16%) |  | 1.00 | 0.497 | 1.00 | (A) Metastasis |
| **AXL** | 19q13.2 | 6 (6.25%) | 3 (3.16%) |  | 0.98 | 0.497 | 1.00 | (A) Metastasis |
| **ESR1** | 6q25.1-q25.2 | 6 (6.25%) | 3 (3.16%) |  | 0.98 | 0.497 | 1.00 | (A) Metastasis |
| **NF2** | 22q12.2 | 6 (6.25%) | 3 (3.16%) |  | 0.98 | 0.497 | 1.00 | (A) Metastasis |
| **PRDM1** | 6q21 | 6 (6.25%) | 3 (3.16%) |  | 0.98 | 0.497 | 1.00 | (A) Metastasis |
| **RET** | 10q11.21 | 6 (6.25%) | 3 (3.13%) |  | 1.00 | 0.497 | 1.00 | (A) Metastasis |
| **RNF43** | 17q22 | 6 (6.38%) | 8 (8.89%) |  | -0.48 | 0.586 | 1.00 | (B) Primary |
| **ARID5B** | 10q21.2 | 6 (8.22%) | 4 (5.48%) |  | 0.58 | 0.745 | 1.00 | (A) Metastasis |
| **MDC1** | 6p21.33 | 6 (8.22%) | 4 (5.48%) |  | 0.58 | 0.745 | 1.00 | (A) Metastasis |
| **NOTCH2** | 1p12 | 6 (6.25%) | 4 (4.21%) |  | 0.57 | 0.747 | 1.00 | (A) Metastasis |
| **TET2** | 4q24 | 6 (6.25%) | 4 (4.21%) |  | 0.57 | 0.747 | 1.00 | (A) Metastasis |
| **KDM5A** | 12p13.33 | 6 (6.38%) | 4 (4.44%) |  | 0.52 | 0.748 | 1.00 | (A) Metastasis |
| **NSD1** | 5q35.3 | 6 (6.38%) | 7 (7.78%) |  | -0.29 | 0.779 | 1.00 | (B) Primary |
| **NFE2L2** | 2q31.2 | 6 (6.25%) | 7 (7.37%) |  | -0.24 | 0.782 | 1.00 | (B) Primary |
| **ARHGAP35** | 19q13.32 | 6 (10.34%) | 5 (11.11%) |  | -0.10 | 1.00 | 1.00 | (B) Primary |
| **SH2B3** | 12q24.12 | 6 (6.32%) | 5 (5.26%) |  | 0.26 | 1.00 | 1.00 | (A) Metastasis |
| **KDM5C** | Xp11.22 | 6 (6.32%) | 5 (5.32%) |  | 0.25 | 1.00 | 1.00 | (A) Metastasis |
| **AMER1** | Xq11.2 | 6 (8.11%) | 5 (6.85%) |  | 0.24 | 1.00 | 1.00 | (A) Metastasis |
| **NOTCH4** | 6p21.32 | 6 (8.11%) | 5 (6.85%) |  | 0.24 | 1.00 | 1.00 | (A) Metastasis |
| **ATRX** | Xq21.1 | 6 (6.25%) | 6 (6.32%) |  | -0.02 | 1.00 | 1.00 | (B) Primary |
| **AGO2** | 8q24.3 | 6 (8.96%) | 5 (7.58%) |  | 0.24 | 1.00 | 1.00 | (A) Metastasis |
| **MGA** | 15q15.1 | 6 (6.52%) | 5 (5.56%) |  | 0.23 | 1.00 | 1.00 | (A) Metastasis |
| **FLT4** | 5q35.3 | 6 (6.25%) | 5 (5.26%) |  | 0.25 | 1.00 | 1.00 | (A) Metastasis |
| **COL7A1** | 3p21.31 | 5 (25.00%) | 1 (5.88%) |  | 2.Eyl | 0.189 | 1.00 | (A) Metastasis |
| **EPHA5** | 4q13.1-q13.2 | 5 (6.58%) | 2 (2.56%) |  | Oca.36 | 0.273 | 1.00 | (A) Metastasis |
| **POLE** | 12q24.33 | 5 (5.38%) | 7 (7.78%) |  | -0.53 | 0.563 | 1.00 | (B) Primary |
| **MSH2** | 2p21-p16.3 | 5 (5.21%) | 7 (7.37%) |  | -0.50 | 0.567 | 1.00 | (B) Primary |
| **POLD1** | 19q13.33 | 5 (5.43%) | 3 (3.33%) |  | 0.71 | 0.721 | 1.00 | (A) Metastasis |
| **DNMT3A** | 2p23.3 | 5 (5.21%) | 3 (3.16%) |  | 0.72 | 0.721 | 1.00 | (A) Metastasis |
| **PALB2** | 16p12.2 | 5 (5.21%) | 3 (3.16%) |  | 0.72 | 0.721 | 1.00 | (A) Metastasis |
| **SMO** | 7q32.1 | 5 (5.21%) | 3 (3.13%) |  | 0.74 | 0.721 | 1.00 | (A) Metastasis |
| **GLI1** | 12q13.3 | 5 (5.32%) | 7 (7.37%) |  | -0.47 | 0.767 | 1.00 | (B) Primary |
| **MRE11** | 11q21 | 5 (5.32%) | 4 (4.44%) |  | 0.26 | 1.00 | 1.00 | (A) Metastasis |
| **UPF1** | 19p13.11 | 5 (7.46%) | 4 (6.06%) |  | 0.30 | 1.00 | 1.00 | (A) Metastasis |
| **KMT2C** | 7q36.1 | 5 (6.76%) | 5 (6.85%) |  | -0.02 | 1.00 | 1.00 | (B) Primary |
| **BLM** | 15q26.1 | 5 (5.21%) | 4 (4.21%) |  | 0.31 | 1.00 | 1.00 | (A) Metastasis |
| **DIS3** | 13q21.33 | 5 (5.21%) | 4 (4.21%) |  | 0.31 | 1.00 | 1.00 | (A) Metastasis |
| **GRIN2A** | 16p13.2 | 5 (6.76%) | 4 (5.48%) |  | 0.30 | 1.00 | 1.00 | (A) Metastasis |

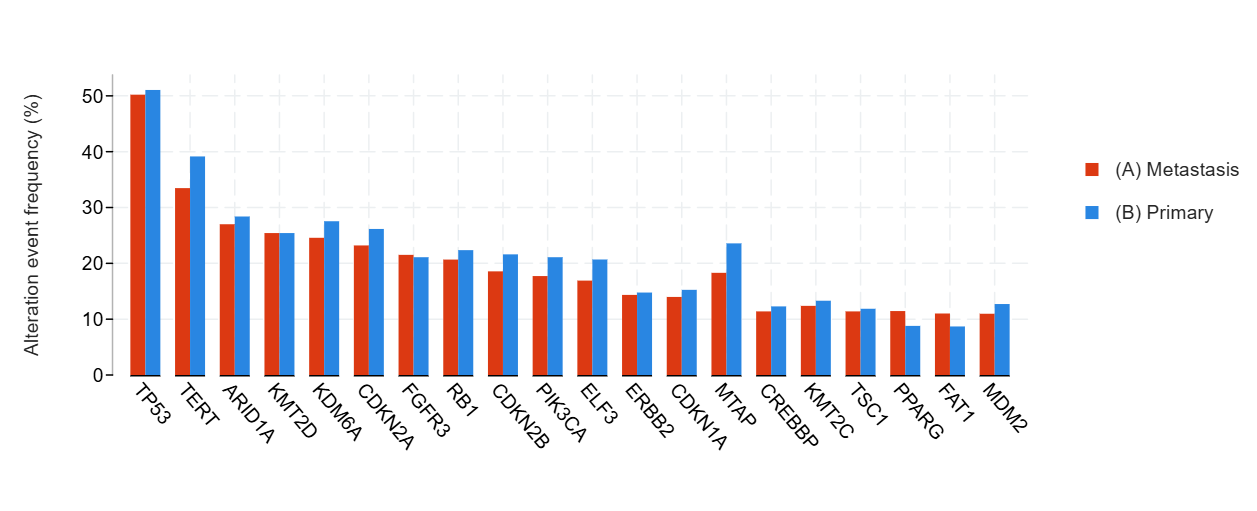

**Figure S10.** Distribution of the Most Frequently Mutated Genes in Primary and Metastatic Bladder Cancer

**Table S11.** **Gene-Level Mutation Frequencies in Primary Versus Metastatic Bladder Cancer**

| **Gene** | **Cytoband** | **(A) Metastasis** | **(B) Primary** | **Co-occurrence Pattern** | **Log2 Ratio** | **p-Value** | **q-Value** | **Enriched in** |
| --- | --- | --- | --- | --- | --- | --- | --- | --- |
| **TP53** | 17p13.1 | 119 (50.21%) | 121 (51.05%) |  | -0.02 | 0.927 | 1.00 | (B) Primary |
| **TERT** | 5p15.33 | 79 (33.47%) | 92 (39.15%) |  | -0.23 | 0.214 | 1.00 | (B) Primary |
| **ARID1A** | 1p36.11 | 64 (27.00%) | 67 (28.39%) |  | -0.07 | 0.759 | 1.00 | (B) Primary |
| **KMT2D** | 12q13.12 | 60 (25.42%) | 60 (25.42%) |  | - | 1.00 | 1.00 | (B) Primary |
| **KDM6A** | Xp11.3 | 58 (24.58%) | 65 (27.54%) |  | -0.16 | 0.529 | 1.00 | (B) Primary |
| **CDKN2A** | 9p21.3 | 55 (23.21%) | 62 (26.16%) |  | -0.17 | 0.523 | 1.00 | (B) Primary |
| **FGFR3** | 4p16.3 | 51 (21.52%) | 50 (21.10%) |  | 0.03 | 1.00 | 1.00 | (A) Metastasis |
| **RB1** | 13q14.2 | 49 (20.68%) | 53 (22.36%) |  | -0.11 | 0.738 | 1.00 | (B) Primary |
| **CDKN2B** | 9p21.3 | 44 (18.57%) | 51 (21.61%) |  | -0.22 | 0.424 | 1.00 | (B) Primary |
| **PIK3CA** | 3q26.32 | 42 (17.72%) | 50 (21.10%) |  | -0.25 | 0.416 | 1.00 | (B) Primary |
| **ELF3** | 1q32.1 | 35 (16.91%) | 42 (20.69%) |  | -0.29 | 0.376 | 1.00 | (B) Primary |
| **ERBB2** | 17q12 | 34 (14.35%) | 35 (14.77%) |  | -0.04 | 1.00 | 1.00 | (B) Primary |
| **CDKN1A** | 6p21.2 | 33 (13.98%) | 36 (15.25%) |  | -0.13 | 0.795 | 1.00 | (B) Primary |
| **MTAP** | 9p21.3 | 28 (18.30%) | 29 (23.58%) |  | -0.37 | 0.298 | 1.00 | (B) Primary |
| **CREBBP** | 16p13.3 | 27 (11.39%) | 29 (12.29%) |  | -0.11 | 0.778 | 1.00 | (B) Primary |
| **KMT2C** | 7q36.1 | 27 (12.39%) | 29 (13.30%) |  | -0.10 | 0.886 | 1.00 | (B) Primary |
| **TSC1** | 9q34.13 | 27 (11.39%) | 28 (11.86%) |  | -0.06 | 0.887 | 1.00 | (B) Primary |
| **PPARG** | 3p25.2 | 26 (11.45%) | 19 (8.80%) |  | 0.38 | 0.432 | 1.00 | (A) Metastasis |
| **FAT1** | 4q35.2 | 26 (11.02%) | 20 (8.70%) |  | 0.34 | 0.440 | 1.00 | (A) Metastasis |
| **MDM2** | 12q15 | 26 (10.97%) | 30 (12.71%) |  | -0.21 | 0.572 | 1.00 | (B) Primary |
| **ERBB3** | 12q13.2 | 24 (10.13%) | 21 (8.90%) |  | 0.19 | 0.754 | 1.00 | (A) Metastasis |
| **STAG2** | Xq25 | 23 (9.75%) | 30 (12.71%) |  | -0.38 | 0.382 | 1.00 | (B) Primary |
| **ATM** | 11q22.3 | 23 (9.70%) | 24 (10.13%) |  | -0.06 | 1.00 | 1.00 | (B) Primary |
| **CDK12** | 17q12 | 23 (9.70%) | 23 (10.00%) |  | -0.04 | 1.00 | 1.00 | (B) Primary |
| **CCND1** | 11q13.3 | 21 (8.86%) | 23 (9.75%) |  | -0.14 | 0.754 | 1.00 | (B) Primary |
| **PTPRT** | 20q12-q13.11 | 21 (9.68%) | 20 (9.22%) |  | 0.07 | 1.00 | 1.00 | (A) Metastasis |
| **SMARCA4** | 19p13.2 | 20 (8.44%) | 18 (7.63%) |  | 0.15 | 0.866 | 1.00 | (A) Metastasis |
| **E2F3** | 6p22.3 | 19 (8.76%) | 26 (11.98%) |  | -0.45 | 0.345 | 1.00 | (B) Primary |
| **RAF1** | 3p25.2 | 19 (8.02%) | 15 (6.36%) |  | 0.33 | 0.594 | 1.00 | (A) Metastasis |
| **EP300** | 22q13.2 | 19 (8.05%) | 23 (9.75%) |  | -0.28 | 0.628 | 1.00 | (B) Primary |
| **NF1** | 17q11.2 | 19 (8.02%) | 16 (6.78%) |  | 0.24 | 0.726 | 1.00 | (A) Metastasis |
| **ATR** | 3q23 | 19 (8.02%) | 20 (8.70%) |  | -0.12 | 0.868 | 1.00 | (B) Primary |
| **NOTCH4** | 6p21.32 | 18 (8.26%) | 15 (6.88%) |  | 0.26 | 0.718 | 1.00 | (A) Metastasis |
| **KMT2A** | 11q23.3 | 18 (7.63%) | 21 (8.90%) |  | -0.22 | 0.739 | 1.00 | (B) Primary |
| **NSD1** | 5q35.3 | 18 (7.63%) | 16 (6.96%) |  | 0.13 | 0.859 | 1.00 | (A) Metastasis |
| **RBM10** | Xp11.3 | 18 (7.63%) | 18 (7.83%) |  | -0.04 | 1.00 | 1.00 | (B) Primary |
| **FGF4** | 11q13.3 | 18 (8.26%) | 18 (8.26%) |  | - | 1.00 | 1.00 | (B) Primary |
| **FGF3** | 11q13.3 | 18 (8.22%) | 18 (8.26%) |  | -0.01 | 1.00 | 1.00 | (B) Primary |
| **TSC2** | 16p13.3 | 17 (7.17%) | 11 (4.66%) |  | 0.62 | 0.330 | 1.00 | (A) Metastasis |
| **FOXA1** | 14q21.1 | 17 (7.23%) | 12 (5.24%) |  | 0.47 | 0.445 | 1.00 | (A) Metastasis |
| **ATRX** | Xq21.1 | 17 (7.17%) | 13 (5.51%) |  | 0.38 | 0.572 | 1.00 | (A) Metastasis |
| **POLE** | 12q24.33 | 17 (7.17%) | 14 (6.09%) |  | 0.24 | 0.712 | 1.00 | (A) Metastasis |
| **SLX4** | 16p13.3 | 17 (7.52%) | 14 (6.51%) |  | 0.21 | 0.713 | 1.00 | (A) Metastasis |
| **SPEN** | 1p36.21-p36.13 | 17 (7.80%) | 20 (9.17%) |  | -0.23 | 0.732 | 1.00 | (B) Primary |
| **FGF19** | 11q13.3 | 17 (7.76%) | 19 (8.72%) |  | -0.17 | 0.732 | 1.00 | (B) Primary |
| **BRCA2** | 13q13.1 | 17 (7.17%) | 15 (6.36%) |  | 0.17 | 0.855 | 1.00 | (A) Metastasis |
| **MGA** | 15q15.1 | 16 (6.90%) | 21 (9.29%) |  | -0.43 | 0.393 | 1.00 | (B) Primary |
| **MCL1** | 1q21.2 | 16 (6.78%) | 20 (8.47%) |  | -0.32 | 0.603 | 1.00 | (B) Primary |
| **BAP1** | 3p21.1 | 16 (6.75%) | 13 (5.51%) |  | 0.29 | 0.702 | 1.00 | (A) Metastasis |
| **NSD3** | 8p11.23 | 16 (7.11%) | 18 (8.37%) |  | -0.24 | 0.722 | 1.00 | (B) Primary |
| **TBX3** | 12q24.21 | 16 (7.34%) | 15 (6.88%) |  | 0.09 | 1.00 | 1.00 | (A) Metastasis |
| **TET2** | 4q24 | 15 (6.36%) | 10 (4.24%) |  | 0.58 | 0.412 | 1.00 | (A) Metastasis |
| **EPHA5** | 4q13.1-q13.2 | 15 (6.88%) | 11 (4.91%) |  | 0.49 | 0.423 | 1.00 | (A) Metastasis |
| **ERCC2** | 19q13.32 | 15 (6.33%) | 19 (8.05%) |  | -0.35 | 0.483 | 1.00 | (B) Primary |
| **PTPRD** | 9p24.1-p23 | 15 (6.88%) | 13 (5.86%) |  | 0.23 | 0.700 | 1.00 | (A) Metastasis |
| **EGFR** | 7p11.2 | 15 (6.33%) | 13 (5.49%) |  | 0.21 | 0.846 | 1.00 | (A) Metastasis |
| **NOTCH3** | 19p13.12 | 15 (6.33%) | 13 (5.65%) |  | 0.16 | 0.846 | 1.00 | (A) Metastasis |
| **ARID2** | 12q12 | 15 (6.36%) | 15 (6.36%) |  | - | 1.00 | 1.00 | (B) Primary |
| **NOTCH1** | 9q34.3 | 14 (5.91%) | 11 (4.64%) |  | 0.35 | 0.547 | 1.00 | (A) Metastasis |
| **ARID1B** | 6q25.3 | 14 (5.93%) | 17 (7.20%) |  | -0.28 | 0.711 | 1.00 | (B) Primary |
| **APC** | 5q22.2 | 14 (5.93%) | 13 (5.49%) |  | 0.11 | 0.846 | 1.00 | (A) Metastasis |
| **FGFR1** | 8p11.23 | 14 (5.91%) | 14 (5.91%) |  | - | 1.00 | 1.00 | (B) Primary |
| **FLT4** | 5q35.3 | 14 (5.93%) | 15 (6.36%) |  | -0.10 | 1.00 | 1.00 | (B) Primary |
| **SETD2** | 3p21.31 | 14 (5.91%) | 13 (5.51%) |  | 0.10 | 1.00 | 1.00 | (A) Metastasis |
| **SDHA** | 5p15.33 | 13 (5.51%) | 6 (2.56%) |  | 1.Eki | 0.158 | 1.00 | (A) Metastasis |
| **KMT2B** | 19q13.12 | 13 (6.28%) | 20 (9.85%) |  | -0.65 | 0.207 | 1.00 | (B) Primary |
| **PTPRS** | 19p13.3 | 13 (5.99%) | 8 (3.69%) |  | 0.70 | 0.371 | 1.00 | (A) Metastasis |
| **PIK3C2G** | 12p12.3 | 13 (5.96%) | 11 (5.05%) |  | 0.24 | 0.834 | 1.00 | (A) Metastasis |
| **NCOR1** | 17p12-p11.2 | 13 (5.96%) | 15 (6.88%) |  | -0.21 | 0.846 | 1.00 | (B) Primary |
| **CDH1** | 16q22.1 | 13 (5.51%) | 13 (5.49%) |  | 0.01 | 1.00 | 1.00 | (A) Metastasis |
| **GRIN2A** | 16p13.2 | 12 (5.50%) | 19 (8.72%) |  | -0.66 | 0.263 | 1.00 | (B) Primary |
| **GATA3** | 10p14 | 12 (5.08%) | 19 (8.05%) |  | -0.66 | 0.265 | 1.00 | (B) Primary |
| **IGF1R** | 15q26.3 | 12 (5.06%) | 9 (3.81%) |  | 0.41 | 0.656 | 1.00 | (A) Metastasis |
| **NF2** | 22q12.2 | 12 (5.06%) | 14 (5.93%) |  | -0.23 | 0.693 | 1.00 | (B) Primary |
| **PREX2** | 8q13.2 | 12 (5.77%) | 10 (4.90%) |  | 0.24 | 0.827 | 1.00 | (A) Metastasis |
| **FANCA** | 16q24.3 | 12 (5.06%) | 10 (4.24%) |  | 0.26 | 0.828 | 1.00 | (A) Metastasis |
| **ZFHX3** | 16q22.2-q22.3 | 12 (5.61%) | 12 (5.61%) |  | - | 1.00 | 1.00 | (B) Primary |
| **SF3B1** | 2q33.1 | 12 (5.06%) | 11 (4.66%) |  | 0.12 | 1.00 | 1.00 | (A) Metastasis |
| **CIC** | 19q13.2 | 11 (4.66%) | 6 (2.61%) |  | 0.84 | 0.324 | 1.00 | (A) Metastasis |
| **LATS1** | 6q25.1 | 11 (5.07%) | 7 (3.23%) |  | 0.65 | 0.471 | 1.00 | (A) Metastasis |
| **CARD11** | 7p22.2 | 11 (4.66%) | 7 (2.97%) |  | 0.65 | 0.472 | 1.00 | (A) Metastasis |
| **SETDB1** | 1q21.3 | 11 (8.15%) | 12 (10.81%) |  | -0.41 | 0.515 | 1.00 | (B) Primary |
| **ANKRD11** | 16q24.3 | 11 (5.14%) | 15 (7.01%) |  | -0.45 | 0.545 | 1.00 | (B) Primary |
| **RET** | 10q11.21 | 11 (4.64%) | 8 (3.38%) |  | 0.46 | 0.641 | 1.00 | (A) Metastasis |
| **JAK1** | 1p31.3 | 11 (4.64%) | 8 (3.48%) |  | 0.42 | 0.641 | 1.00 | (A) Metastasis |
| **FBXW7** | 4q31.3 | 11 (4.64%) | 14 (5.91%) |  | -0.35 | 0.682 | 1.00 | (B) Primary |
| **MYC** | 8q24.21 | 11 (4.64%) | 12 (5.08%) |  | -0.13 | 0.835 | 1.00 | (B) Primary |
| **SH2B3** | 12q24.12 | 11 (4.72%) | 12 (5.15%) |  | -0.13 | 1.00 | 1.00 | (B) Primary |
| **ROS1** | 6q22.1 | 11 (4.64%) | 11 (4.66%) |  | -0.01 | 1.00 | 1.00 | (B) Primary |
| **DNMT1** | 19p13.2 | 11 (5.07%) | 11 (5.07%) |  | - | 1.00 | 1.00 | (B) Primary |
| **KRAS** | 12p12.1 | 11 (4.64%) | 10 (4.22%) |  | 0.14 | 1.00 | 1.00 | (A) Metastasis |
| **TGFBR1** | 9q22.33 | 11 (5.07%) | 12 (5.53%) |  | -0.13 | 1.00 | 1.00 | (B) Primary |
| **VHL** | 3p25.3 | 10 (4.24%) | 4 (1.69%) |  | Oca.33 | 0.113 | 1.00 | (A) Metastasis |
| **SDHC** | 1q23.3 | 10 (4.24%) | 15 (6.36%) |  | -0.58 | 0.412 | 1.00 | (B) Primary |
| **FLT1** | 13q12.3 | 10 (4.24%) | 7 (2.97%) |  | 0.51 | 0.623 | 1.00 | (A) Metastasis |
| **RICTOR** | 5p13.1 | 10 (4.22%) | 12 (5.22%) |  | -0.31 | 0.666 | 1.00 | (B) Primary |
| **PBRM1** | 3p21.1 | 10 (4.24%) | 8 (3.42%) |  | 0.31 | 0.811 | 1.00 | (A) Metastasis |
| **TP53BP1** | 15q15.3 | 10 (4.42%) | 11 (5.09%) |  | -0.20 | 0.825 | 1.00 | (B) Primary |
| **DDR2** | 1q23.3 | 10 (4.22%) | 11 (4.66%) |  | -0.14 | 0.828 | 1.00 | (B) Primary |
| **MYCL** | 1p34.2 | 10 (4.22%) | 11 (4.66%) |  | -0.14 | 0.828 | 1.00 | (B) Primary |

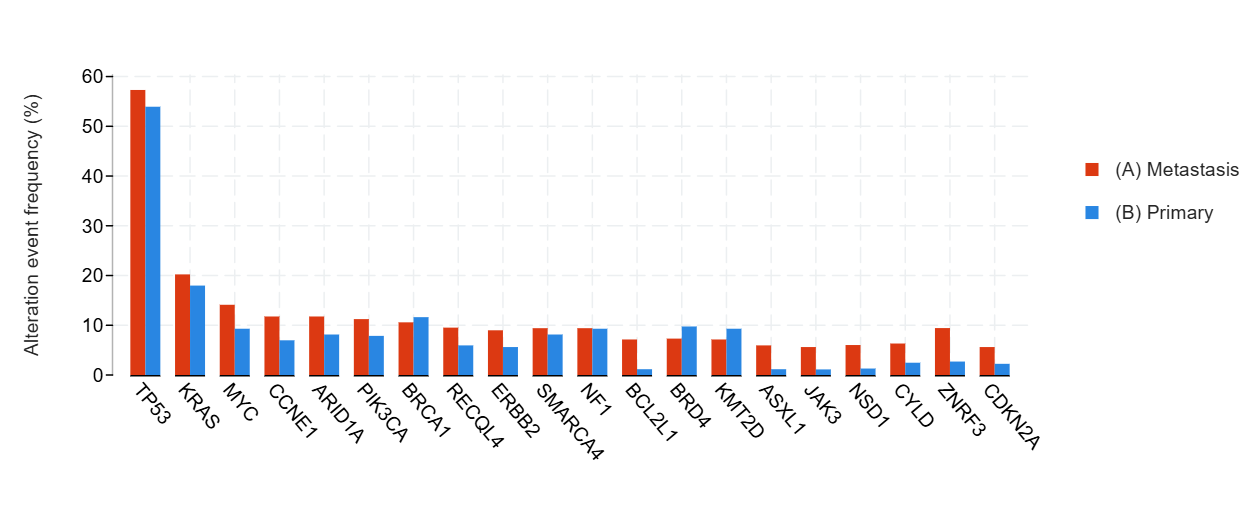

**Figure S11.** Distribution of the Most Frequently Mutated Genes in Primary and Metastatic Ovarian Cancer

**Table S12.** **Gene-Level Mutation Frequencies in Primary Versus Metastatic Ovarian Cancer**

| **Gene** | **Cytoband** | **(A) Metastasis** | **(B) Primary** | **Co-occurrence Pattern** | **Log2 Ratio** | **p-Value** | **q-Value** | **Enriched in** |
| --- | --- | --- | --- | --- | --- | --- | --- | --- |
| **TP53** | 17p13.1 | 51 (57.30%) | 48 (53.93%) |  | 0.09 | 0.763 | 1.00 | (A) Metastasis |
| **KRAS** | 12p12.1 | 18 (20.22%) | 16 (17.98%) |  | 0.17 | 0.708 | 1.00 | (A) Metastasis |
| **MYC** | 8q24.21 | 12 (14.12%) | 8 (9.30%) |  | 0.60 | 0.352 | 1.00 | (A) Metastasis |
| **CCNE1** | 19q12 | 10 (11.76%) | 6 (6.98%) |  | 0.75 | 0.307 | 1.00 | (A) Metastasis |
| **ARID1A** | 1p36.11 | 10 (11.76%) | 7 (8.14%) |  | 0.53 | 0.456 | 1.00 | (A) Metastasis |
| **PIK3CA** | 3q26.32 | 10 (11.24%) | 7 (7.87%) |  | 0.51 | 0.611 | 1.00 | (A) Metastasis |
| **BRCA1** | 17q21.31 | 9 (10.59%) | 10 (11.63%) |  | -0.14 | 1.00 | 1.00 | (B) Primary |
| **RECQL4** | 8q24.3 | 8 (9.52%) | 5 (5.95%) |  | 0.68 | 0.565 | 1.00 | (A) Metastasis |
| **ERBB2** | 17q12 | 8 (8.99%) | 5 (5.62%) |  | 0.68 | 0.566 | 1.00 | (A) Metastasis |
| **SMARCA4** | 19p13.2 | 8 (9.41%) | 7 (8.14%) |  | 0.21 | 0.794 | 1.00 | (A) Metastasis |
| **NF1** | 17q11.2 | 8 (9.41%) | 8 (9.30%) |  | 0.02 | 1.00 | 1.00 | (A) Metastasis |
| **BCL2L1** | 20q11.21 | 6 (7.14%) | 1 (1.16%) |  | Şub.62 | 0.0624 | 1.00 | (A) Metastasis |
| **BRD4** | 19p13.12 | 6 (7.32%) | 8 (9.76%) |  | -0.42 | 0.781 | 1.00 | (B) Primary |
| **KMT2D** | 12q13.12 | 6 (7.14%) | 8 (9.30%) |  | -0.38 | 0.782 | 1.00 | (B) Primary |
| **ASXL1** | 20q11.21 | 5 (5.95%) | 1 (1.16%) |  | Şub.36 | 0.115 | 1.00 | (A) Metastasis |
| **JAK3** | 19p13.11 | 5 (5.62%) | 1 (1.12%) |  | Şub.32 | 0.211 | 1.00 | (A) Metastasis |
| **NSD1** | 5q35.3 | 5 (6.02%) | 1 (1.30%) |  | Şub.21 | 0.212 | 1.00 | (A) Metastasis |
| **CYLD** | 16q12.1 | 5 (6.33%) | 2 (2.47%) |  | Oca.36 | 0.274 | 1.00 | (A) Metastasis |
| **ZNRF3** | 22q12.1 | 5 (9.43%) | 1 (2.70%) |  | Oca.80 | 0.394 | 1.00 | (A) Metastasis |
| **CDKN2A** | 9p21.3 | 5 (5.62%) | 2 (2.25%) |  | Oca.32 | 0.444 | 1.00 | (A) Metastasis |
| **KDM5C** | Xp11.22 | 5 (5.95%) | 3 (3.57%) |  | 0.74 | 0.720 | 1.00 | (A) Metastasis |
| **BRAF** | 7q34 | 5 (5.62%) | 3 (3.37%) |  | 0.74 | 0.720 | 1.00 | (A) Metastasis |
| **MET** | 7q31.2 | 5 (5.62%) | 3 (3.37%) |  | 0.74 | 0.720 | 1.00 | (A) Metastasis |
| **NOTCH3** | 19p13.12 | 5 (6.10%) | 5 (6.85%) |  | -0.17 | 1.00 | 1.00 | (B) Primary |
| **CDK12** | 17q12 | 5 (5.95%) | 4 (5.19%) |  | 0.20 | 1.00 | 1.00 | (A) Metastasis |
| **PRKCI** | 3q26.2 | 5 (6.58%) | 4 (5.26%) |  | 0.32 | 1.00 | 1.00 | (A) Metastasis |
| **SH2B3** | 12q24.12 | 5 (6.10%) | 4 (4.88%) |  | 0.32 | 1.00 | 1.00 | (A) Metastasis |
| **FGFR1** | 8p11.23 | 5 (5.62%) | 4 (4.49%) |  | 0.32 | 1.00 | 1.00 | (A) Metastasis |
| **CEBPA** | 19q13.11 | 4 (4.88%) | 0 (0.00%) |  | >10 | 0.120 | 1.00 | (A) Metastasis |
| **BABAM1** | 19p13.11 | 4 (5.33%) | 0 (0.00%) |  | >10 | 0.122 | 1.00 | (A) Metastasis |
| **ROS1** | 6q22.1 | 4 (4.71%) | 1 (1.16%) |  | 2.Şub | 0.210 | 1.00 | (A) Metastasis |
| **TFE3** | Xp11.23 | 4 (12.50%) | 1 (3.85%) |  | Oca.70 | 0.367 | 1.00 | (A) Metastasis |
| **NSD3** | 8p11.23 | 4 (5.33%) | 1 (1.49%) |  | Oca.84 | 0.370 | 1.00 | (A) Metastasis |
| **KMT2C** | 7q36.1 | 4 (7.41%) | 2 (3.57%) |  | 1.May | 0.434 | 1.00 | (A) Metastasis |
| **CDKN2B** | 9p21.3 | 4 (4.71%) | 2 (2.33%) |  | 1.Şub | 0.443 | 1.00 | (A) Metastasis |
| **CREBBP** | 16p13.3 | 4 (4.71%) | 2 (2.33%) |  | 1.Şub | 0.443 | 1.00 | (A) Metastasis |
| **CDH1** | 16q22.1 | 4 (4.55%) | 2 (2.25%) |  | 1.Şub | 0.444 | 1.00 | (A) Metastasis |
| **RAD21** | 8q24.11 | 4 (4.88%) | 2 (2.44%) |  | 1.00 | 0.682 | 1.00 | (A) Metastasis |
| **FAT1** | 4q35.2 | 4 (4.94%) | 2 (2.74%) |  | 0.85 | 0.684 | 1.00 | (A) Metastasis |
| **ACVR1** | 2q24.1 | 4 (5.06%) | 2 (2.90%) |  | 0.80 | 0.685 | 1.00 | (A) Metastasis |
| **PTPRD** | 9p24.1-p23 | 4 (7.27%) | 3 (4.76%) |  | 0.61 | 0.704 | 1.00 | (A) Metastasis |
| **MECOM** | 3q26.2 | 4 (12.50%) | 3 (8.82%) |  | 0.50 | 0.705 | 1.00 | (A) Metastasis |
| **GATA1** | Xp11.23 | 4 (7.41%) | 3 (5.36%) |  | 0.47 | 0.714 | 1.00 | (A) Metastasis |
| **CARD11** | 7p22.2 | 4 (4.76%) | 3 (3.49%) |  | 0.45 | 0.718 | 1.00 | (A) Metastasis |
| **EIF1AX** | Xp22.12 | 4 (7.69%) | 5 (9.62%) |  | -0.32 | 1.00 | 1.00 | (B) Primary |
| **TET1** | 10q21.3 | 4 (4.88%) | 4 (5.26%) |  | -0.11 | 1.00 | 1.00 | (B) Primary |
| **KDM6A** | Xp11.3 | 4 (4.76%) | 4 (4.65%) |  | 0.03 | 1.00 | 1.00 | (A) Metastasis |
| **COL7A1** | 3p21.31 | 4 (13.79%) | 3 (14.29%) |  | -0.05 | 1.00 | 1.00 | (B) Primary |
| **GNA11** | 19p13.3 | 4 (4.49%) | 3 (3.37%) |  | 0.42 | 1.00 | 1.00 | (A) Metastasis |
| **RET** | 10q11.21 | 4 (4.49%) | 3 (3.37%) |  | 0.42 | 1.00 | 1.00 | (A) Metastasis |
| **AR** | Xq12 | 4 (4.71%) | 5 (5.81%) |  | -0.31 | 1.00 | 1.00 | (B) Primary |
| **NTRK1** | 1q23.1 | 4 (4.71%) | 5 (5.81%) |  | -0.31 | 1.00 | 1.00 | (B) Primary |
| **AGO2** | 8q24.3 | 4 (8.70%) | 3 (6.52%) |  | 0.42 | 1.00 | 1.00 | (A) Metastasis |
| **ZFHX3** | 16q22.2-q22.3 | 3 (6.00%) | 0 (0.00%) |  | >10 | 0.243 | 1.00 | (A) Metastasis |
| **GEN1** | 2p24.2 | 3 (10.00%) | 0 (0.00%) |  | >10 | 0.253 | 1.00 | (A) Metastasis |
| **MTAP** | 9p21.3 | 3 (5.66%) | 0 (0.00%) |  | >10 | 0.266 | 1.00 | (A) Metastasis |
| **AURKB** | 17p13.1 | 3 (3.57%) | 1 (1.16%) |  | Oca.62 | 0.365 | 1.00 | (A) Metastasis |
| **ETV6** | 12p13.2 | 3 (3.57%) | 1 (1.16%) |  | Oca.62 | 0.365 | 1.00 | (A) Metastasis |
| **WT1** | 11p13 | 3 (3.57%) | 1 (1.16%) |  | Oca.62 | 0.365 | 1.00 | (A) Metastasis |
| **PMS2** | 7p22.1 | 3 (3.53%) | 1 (1.16%) |  | Oca.60 | 0.368 | 1.00 | (A) Metastasis |

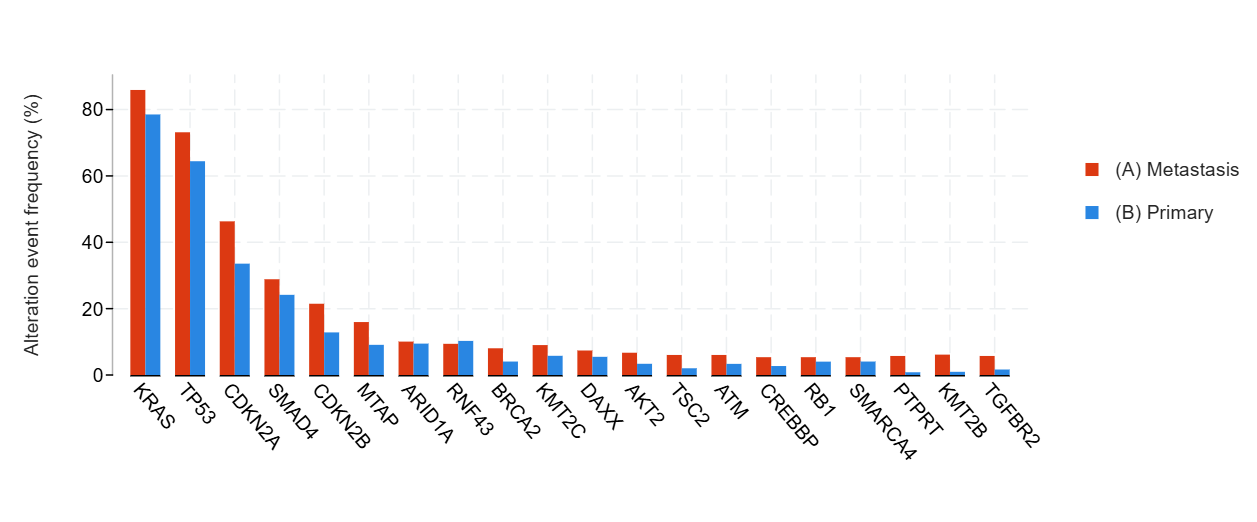

**Figure S12.** Distribution of the Most Frequently Mutated Genes in Primary and Metastatic Pancreatic Cancer

**Table S13**. **Gene-Level Mutation Frequencies in Primary Versus Metastatic Pancreatic Cancer**

| **Gene** | **Cytoband** | **(A) Metastasis** | **(B) Primary** | **Co-occurrence Pattern** | **Log2 Ratio** | **p-Value** | **q-Value** | **Enriched in** |
| --- | --- | --- | --- | --- | --- | --- | --- | --- |
| **KRAS** | 12p12.1 | 128 (85.91%) | 117 (78.52%) |  | 0.13 | 0.129 | 1.00 | (A) Metastasis |
| **TP53** | 17p13.1 | 109 (73.15%) | 96 (64.43%) |  | 0.18 | 0.133 | 1.00 | (A) Metastasis |
| **CDKN2A** | 9p21.3 | 69 (46.31%) | 50 (33.56%) |  | 0.46 | 0.0330 | 0.739 | (A) Metastasis |
| **SMAD4** | 18q21.2 | 43 (28.86%) | 36 (24.16%) |  | 0.26 | 0.431 | 1.00 | (A) Metastasis |
| **CDKN2B** | 9p21.3 | 32 (21.48%) | 19 (12.84%) |  | 0.74 | 0.0642 | 1.00 | (A) Metastasis |
| **MTAP** | 9p21.3 | 15 (15.96%) | 6 (9.09%) |  | 0.81 | 0.241 | 1.00 | (A) Metastasis |
| **ARID1A** | 1p36.11 | 15 (10.07%) | 14 (9.46%) |  | 0.09 | 1.00 | 1.00 | (A) Metastasis |
| **RNF43** | 17q22 | 14 (9.40%) | 15 (10.27%) |  | -0.13 | 0.847 | 1.00 | (B) Primary |
| **BRCA2** | 13q13.1 | 12 (8.05%) | 6 (4.05%) |  | 0.99 | 0.223 | 1.00 | (A) Metastasis |
| **KMT2C** | 7q36.1 | 11 (9.02%) | 7 (5.79%) |  | 0.64 | 0.463 | 1.00 | (A) Metastasis |
| **DAXX** | 6p21.32 | 11 (7.38%) | 8 (5.48%) |  | 0.43 | 0.637 | 1.00 | (A) Metastasis |
| **AKT2** | 19q13.2 | 10 (6.71%) | 5 (3.38%) |  | 0.99 | 0.289 | 1.00 | (A) Metastasis |
| **TSC2** | 16p13.3 | 9 (6.04%) | 3 (2.03%) |  | Oca.58 | 0.138 | 1.00 | (A) Metastasis |
| **ATM** | 11q22.3 | 9 (6.04%) | 5 (3.36%) |  | 0.85 | 0.291 | 1.00 | (A) Metastasis |
| **CREBBP** | 16p13.3 | 8 (5.37%) | 4 (2.70%) |  | 0.99 | 0.378 | 1.00 | (A) Metastasis |
| **RB1** | 13q14.2 | 8 (5.37%) | 6 (4.03%) |  | 0.42 | 0.785 | 1.00 | (A) Metastasis |
| **SMARCA4** | 19p13.2 | 8 (5.37%) | 6 (4.05%) |  | 0.41 | 0.785 | 1.00 | (A) Metastasis |
| **PTPRT** | 20q12-q13.11 | 7 (5.74%) | 1 (0.83%) |  | Şub.80 | 0.0658 | 1.00 | (A) Metastasis |
| **KMT2B** | 19q13.12 | 7 (6.14%) | 1 (0.96%) |  | Şub.67 | 0.0676 | 1.00 | (A) Metastasis |
| **TGFBR2** | 3p24.1 | 7 (5.74%) | 2 (1.65%) |  | Oca.80 | 0.172 | 1.00 | (A) Metastasis |
| **FANCA** | 16q24.3 | 7 (4.70%) | 2 (1.35%) |  | Oca.80 | 0.173 | 1.00 | (A) Metastasis |
| **SETD2** | 3p21.31 | 7 (4.70%) | 2 (1.35%) |  | Oca.80 | 0.173 | 1.00 | (A) Metastasis |
| **MEN1** | 11q13.1 | 7 (4.70%) | 7 (4.73%) |  | -0.01 | 1.00 | 1.00 | (B) Primary |
| **CCNE1** | 19q12 | 6 (4.03%) | 2 (1.35%) |  | Oca.58 | 0.282 | 1.00 | (A) Metastasis |
| **ATR** | 3q23 | 6 (4.03%) | 2 (1.37%) |  | Oca.56 | 0.283 | 1.00 | (A) Metastasis |
| **PALB2** | 16p12.2 | 6 (4.03%) | 3 (2.03%) |  | 0.99 | 0.501 | 1.00 | (A) Metastasis |
| **U2AF1** | 21q22.3 | 6 (4.03%) | 3 (2.03%) |  | 0.99 | 0.501 | 1.00 | (A) Metastasis |
| **SMAD3** | 15q22.33 | 6 (4.96%) | 4 (3.31%) |  | 0.58 | 0.749 | 1.00 | (A) Metastasis |
| **PREX2** | 8q13.2 | 6 (5.22%) | 4 (3.85%) |  | 0.44 | 0.751 | 1.00 | (A) Metastasis |
| **MYC** | 8q24.21 | 6 (4.03%) | 5 (3.38%) |  | 0.25 | 1.00 | 1.00 | (A) Metastasis |
| **NTRK3** | 15q25.3 | 5 (3.36%) | 1 (0.68%) |  | Şub.31 | 0.214 | 1.00 | (A) Metastasis |
| **PAX5** | 9p13.2 | 5 (3.36%) | 1 (0.68%) |  | Şub.31 | 0.214 | 1.00 | (A) Metastasis |
| **BRAF** | 7q34 | 5 (3.36%) | 2 (1.34%) |  | Oca.32 | 0.448 | 1.00 | (A) Metastasis |
| **ANKRD11** | 16q24.3 | 5 (4.13%) | 3 (2.48%) |  | 0.74 | 0.722 | 1.00 | (A) Metastasis |
| **KMT2D** | 12q13.12 | 5 (3.36%) | 5 (3.38%) |  | -0.01 | 1.00 | 1.00 | (B) Primary |
| **KDM6A** | Xp11.3 | 5 (3.36%) | 4 (2.70%) |  | 0.31 | 1.00 | 1.00 | (A) Metastasis |
| **EP300** | 22q13.2 | 4 (2.68%) | 0 (0.00%) |  | >10 | 0.122 | 1.00 | (A) Metastasis |
| **FAT1** | 4q35.2 | 4 (2.70%) | 0 (0.00%) |  | >10 | 0.122 | 1.00 | (A) Metastasis |
| **MET** | 7q31.2 | 4 (2.68%) | 1 (0.67%) |  | 2.00 | 0.371 | 1.00 | (A) Metastasis |
| **TEK** | 9p21.2 | 4 (3.51%) | 1 (0.96%) |  | Oca.87 | 0.372 | 1.00 | (A) Metastasis |
| **PTPRD** | 9p24.1-p23 | 4 (3.28%) | 2 (1.63%) |  | 1.Oca | 0.446 | 1.00 | (A) Metastasis |
| **POLQ** | 3q13.33 | 4 (14.81%) | 2 (8.00%) |  | 0.89 | 0.670 | 1.00 | (A) Metastasis |
| **GLI1** | 12q13.3 | 4 (2.70%) | 2 (1.35%) |  | 1.00 | 0.684 | 1.00 | (A) Metastasis |
| **CDK6** | 7q21.2 | 4 (2.68%) | 2 (1.35%) |  | 0.99 | 0.684 | 1.00 | (A) Metastasis |
| **ERCC2** | 19q13.32 | 4 (2.68%) | 2 (1.35%) |  | 0.99 | 0.684 | 1.00 | (A) Metastasis |
| **TERT** | 5p15.33 | 4 (2.68%) | 2 (1.35%) |  | 0.99 | 0.684 | 1.00 | (A) Metastasis |
| **AKT1** | 14q32.33 | 4 (2.68%) | 2 (1.34%) |  | 1.00 | 0.684 | 1.00 | (A) Metastasis |
| **ERBB2** | 17q12 | 4 (2.68%) | 2 (1.34%) |  | 1.00 | 0.684 | 1.00 | (A) Metastasis |
| **JAK2** | 9p24.1 | 4 (2.68%) | 2 (1.34%) |  | 1.00 | 0.684 | 1.00 | (A) Metastasis |
| **NOTCH1** | 9q34.3 | 4 (2.68%) | 2 (1.34%) |  | 1.00 | 0.684 | 1.00 | (A) Metastasis |
| **RBM10** | Xp11.3 | 4 (2.70%) | 2 (1.37%) |  | 0.98 | 0.684 | 1.00 | (A) Metastasis |
| **MED12** | Xq13.1 | 4 (2.68%) | 2 (1.37%) |  | 0.97 | 0.684 | 1.00 | (A) Metastasis |
| **RPTOR** | 17q25.3 | 4 (2.68%) | 2 (1.37%) |  | 0.97 | 0.684 | 1.00 | (A) Metastasis |
| **INSR** | 19p13.2 | 4 (3.31%) | 4 (3.31%) |  | - | 1.00 | 1.00 | (B) Primary |
| **PTPRS** | 19p13.3 | 4 (3.31%) | 3 (2.48%) |  | 0.42 | 1.00 | 1.00 | (A) Metastasis |
| **GNAS** | 20q13.32 | 4 (2.68%) | 3 (2.01%) |  | 0.42 | 1.00 | 1.00 | (A) Metastasis |
| **CEBPA** | 19q13.11 | 4 (2.68%) | 3 (2.03%) |  | 0.41 | 1.00 | 1.00 | (A) Metastasis |
| **COL7A1** | 3p21.31 | 4 (14.81%) | 3 (12.00%) |  | 0.30 | 1.00 | 1.00 | (A) Metastasis |
| **NSD3** | 8p11.23 | 4 (2.84%) | 3 (2.33%) |  | 0.29 | 1.00 | 1.00 | (A) Metastasis |
| **FGFR1** | 8p11.23 | 4 (2.68%) | 4 (2.68%) |  | - | 1.00 | 1.00 | (B) Primary |

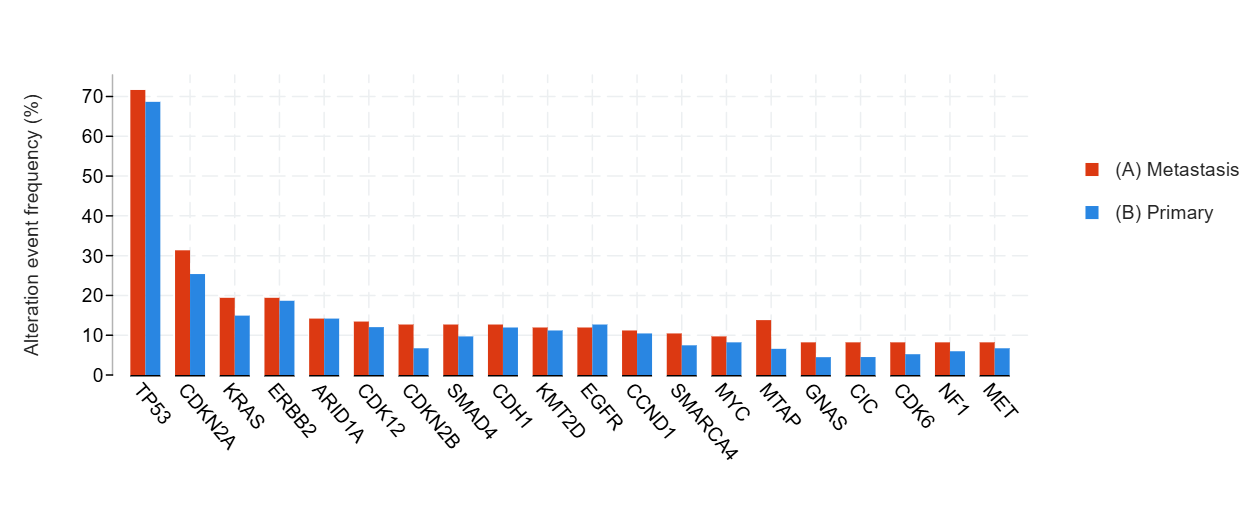

**Figure S13.** Distribution of the Most Frequently Mutated Genes in Primary and Metastatic Esophagogastric Cancer

**Table S14.** **Gene-Level Mutation Frequencies in Primary Versus Metastatic Esophagogastric Cancer**

| **Gene** | **Cytoband** | **(A) Metastasis** | **(B) Primary** | **Co-occurrence Pattern** | **Log2 Ratio** | **p-Value** | **q-Value** | **Enriched in** |
| --- | --- | --- | --- | --- | --- | --- | --- | --- |
| **TP53** | 17p13.1 | 96 (71.64%) | 92 (68.66%) |  | 0.06 | 0.689 | 1.00 | (A) Metastasis |
| **CDKN2A** | 9p21.3 | 42 (31.34%) | 34 (25.37%) |  | 0.30 | 0.343 | 1.00 | (A) Metastasis |
| **KRAS** | 12p12.1 | 26 (19.40%) | 20 (14.93%) |  | 0.38 | 0.418 | 1.00 | (A) Metastasis |
| **ERBB2** | 17q12 | 26 (19.40%) | 25 (18.66%) |  | 0.06 | 1.00 | 1.00 | (A) Metastasis |
| **ARID1A** | 1p36.11 | 19 (14.18%) | 19 (14.18%) |  | - | 1.00 | 1.00 | (B) Primary |
| **CDK12** | 17q12 | 18 (13.43%) | 16 (12.03%) |  | 0.16 | 0.855 | 1.00 | (A) Metastasis |
| **CDKN2B** | 9p21.3 | 17 (12.69%) | 9 (6.72%) |  | 0.92 | 0.147 | 1.00 | (A) Metastasis |
| **SMAD4** | 18q21.2 | 17 (12.69%) | 13 (9.70%) |  | 0.39 | 0.562 | 1.00 | (A) Metastasis |
| **CDH1** | 16q22.1 | 17 (12.69%) | 16 (11.94%) |  | 0.09 | 1.00 | 1.00 | (A) Metastasis |
| **KMT2D** | 12q13.12 | 16 (11.94%) | 15 (11.19%) |  | 0.09 | 1.00 | 1.00 | (A) Metastasis |
| **EGFR** | 7p11.2 | 16 (11.94%) | 17 (12.69%) |  | -0.09 | 1.00 | 1.00 | (B) Primary |
| **CCND1** | 11q13.3 | 15 (11.19%) | 14 (10.45%) |  | 0.10 | 1.00 | 1.00 | (A) Metastasis |
| **SMARCA4** | 19p13.2 | 14 (10.45%) | 10 (7.46%) |  | 0.49 | 0.522 | 1.00 | (A) Metastasis |
| **MYC** | 8q24.21 | 13 (9.70%) | 11 (8.21%) |  | 0.24 | 0.831 | 1.00 | (A) Metastasis |
| **MTAP** | 9p21.3 | 12 (13.79%) | 5 (6.58%) |  | 1.Tem | 0.198 | 1.00 | (A) Metastasis |
| **GNAS** | 20q13.32 | 11 (8.21%) | 6 (4.48%) |  | 0.87 | 0.316 | 1.00 | (A) Metastasis |
| **CIC** | 19q13.2 | 11 (8.21%) | 6 (4.51%) |  | 0.86 | 0.316 | 1.00 | (A) Metastasis |
| **CDK6** | 7q21.2 | 11 (8.21%) | 7 (5.22%) |  | 0.65 | 0.343 | 1.00 | (A) Metastasis |
| **NF1** | 17q11.2 | 11 (8.21%) | 8 (5.97%) |  | 0.46 | 0.635 | 1.00 | (A) Metastasis |
| **MET** | 7q31.2 | 11 (8.21%) | 9 (6.72%) |  | 0.29 | 0.817 | 1.00 | (A) Metastasis |
| **PIK3CA** | 3q26.32 | 11 (8.21%) | 9 (6.72%) |  | 0.29 | 0.817 | 1.00 | (A) Metastasis |
| **RARA** | 17q21.2 | 11 (8.21%) | 10 (7.46%) |  | 0.14 | 1.00 | 1.00 | (A) Metastasis |
| **ARID1B** | 6q25.3 | 10 (7.52%) | 6 (4.51%) |  | 0.74 | 0.440 | 1.00 | (A) Metastasis |
| **ZFHX3** | 16q22.2-q22.3 | 10 (10.64%) | 9 (9.78%) |  | 0.12 | 1.00 | 1.00 | (A) Metastasis |
| **AURKA** | 20q13.2 | 9 (6.72%) | 3 (2.24%) |  | Oca.58 | 0.137 | 1.00 | (A) Metastasis |
| **APC** | 5q22.2 | 9 (6.72%) | 5 (3.73%) |  | 0.85 | 0.411 | 1.00 | (A) Metastasis |
| **VEGFA** | 6p21.1 | 9 (6.98%) | 6 (4.76%) |  | 0.55 | 0.596 | 1.00 | (A) Metastasis |
| **CCNE1** | 19q12 | 9 (6.72%) | 7 (5.22%) |  | 0.36 | 0.797 | 1.00 | (A) Metastasis |
| **BRCA2** | 13q13.1 | 9 (6.72%) | 8 (5.97%) |  | 0.17 | 1.00 | 1.00 | (A) Metastasis |
| **FGFR2** | 10q26.13 | 9 (6.72%) | 9 (6.72%) |  | - | 1.00 | 1.00 | (B) Primary |
| **ASXL1** | 20q11.21 | 8 (5.97%) | 2 (1.49%) |  | 2.00 | 0.103 | 1.00 | (A) Metastasis |
| **KDM6A** | Xp11.3 | 8 (5.97%) | 3 (2.24%) |  | Oca.42 | 0.138 | 1.00 | (A) Metastasis |
| **KDM5A** | 12p13.33 | 8 (5.97%) | 3 (2.26%) |  | Oca.40 | 0.217 | 1.00 | (A) Metastasis |
| **PGR** | 11q22.1 | 8 (8.51%) | 4 (4.35%) |  | 0.97 | 0.372 | 1.00 | (A) Metastasis |
| **FAT1** | 4q35.2 | 8 (6.02%) | 4 (3.03%) |  | 0.99 | 0.377 | 1.00 | (A) Metastasis |
| **NKX2-1** | 14q13.3 | 8 (5.97%) | 4 (2.99%) |  | 1.00 | 0.377 | 1.00 | (A) Metastasis |
| **PTPRT** | 20q12-q13.11 | 8 (8.08%) | 5 (5.05%) |  | 0.68 | 0.568 | 1.00 | (A) Metastasis |
| **ATM** | 11q22.3 | 8 (5.97%) | 6 (4.48%) |  | 0.42 | 0.785 | 1.00 | (A) Metastasis |
| **PTPRD** | 9p24.1-p23 | 8 (8.08%) | 7 (7.00%) |  | 0.21 | 0.795 | 1.00 | (A) Metastasis |
| **FGF19** | 11q13.3 | 8 (8.00%) | 9 (9.00%) |  | -0.17 | 0.807 | 1.00 | (B) Primary |
| **FGF3** | 11q13.3 | 8 (8.00%) | 9 (9.00%) |  | -0.17 | 0.807 | 1.00 | (B) Primary |
| **FGF4** | 11q13.3 | 8 (8.00%) | 9 (9.00%) |  | -0.17 | 0.807 | 1.00 | (B) Primary |
| **ERBB3** | 12q13.2 | 8 (5.97%) | 7 (5.22%) |  | 0.19 | 1.00 | 1.00 | (A) Metastasis |
| **MDM2** | 12q15 | 8 (5.97%) | 7 (5.22%) |  | 0.19 | 1.00 | 1.00 | (A) Metastasis |
| **NOTCH3** | 19p13.12 | 8 (5.97%) | 7 (5.26%) |  | 0.18 | 1.00 | 1.00 | (A) Metastasis |
| **IL7R** | 5p13.2 | 7 (5.26%) | 1 (0.76%) |  | Şub.80 | 0.0662 | 1.00 | (A) Metastasis |
| **CREBBP** | 16p13.3 | 7 (5.22%) | 3 (2.24%) |  | Oca.22 | 0.334 | 1.00 | (A) Metastasis |
| **FBXW7** | 4q31.3 | 7 (5.22%) | 3 (2.24%) |  | Oca.22 | 0.334 | 1.00 | (A) Metastasis |
| **BAP1** | 3p21.1 | 7 (5.22%) | 4 (2.99%) |  | 0.81 | 0.540 | 1.00 | (A) Metastasis |
| **BRD4** | 19p13.12 | 7 (5.22%) | 4 (2.99%) |  | 0.81 | 0.540 | 1.00 | (A) Metastasis |
| **PTEN** | 10q23.31 | 7 (5.22%) | 4 (2.99%) |  | 0.81 | 0.540 | 1.00 | (A) Metastasis |
| **PREX2** | 8q13.2 | 7 (8.33%) | 6 (7.41%) |  | 0.17 | 1.00 | 1.00 | (A) Metastasis |
| **FLT1** | 13q12.3 | 7 (5.22%) | 6 (4.48%) |  | 0.22 | 1.00 | 1.00 | (A) Metastasis |
| **RNF43** | 17q22 | 7 (5.22%) | 7 (5.26%) |  | -0.01 | 1.00 | 1.00 | (B) Primary |
| **CRLF2** | Xp22.33 and Yp11.2 | 6 (4.51%) | 0 (0.00%) |  | >10 | 0.0295 | 1.00 | (A) Metastasis |
| **BCOR** | Xp11.4 | 6 (4.48%) | 3 (2.24%) |  | 1.00 | 0.500 | 1.00 | (A) Metastasis |
| **BRCA1** | 17q21.31 | 6 (4.48%) | 3 (2.24%) |  | 1.00 | 0.500 | 1.00 | (A) Metastasis |
| **IDH2** | 15q26.1 | 6 (4.48%) | 3 (2.24%) |  | 1.00 | 0.500 | 1.00 | (A) Metastasis |
| **NCOA3** | 20q13.12 | 6 (6.38%) | 4 (4.35%) |  | 0.55 | 0.747 | 1.00 | (A) Metastasis |
| **TERT** | 5p15.33 | 6 (4.51%) | 4 (3.01%) |  | 0.58 | 0.749 | 1.00 | (A) Metastasis |
| **NOTCH1** | 9q34.3 | 6 (4.48%) | 4 (2.99%) |  | 0.58 | 0.749 | 1.00 | (A) Metastasis |
| **SMAD2** | 18q21.1 | 6 (4.48%) | 4 (2.99%) |  | 0.58 | 0.749 | 1.00 | (A) Metastasis |
| **BRAF** | 7q34 | 6 (4.48%) | 8 (5.97%) |  | -0.42 | 0.785 | 1.00 | (B) Primary |
| **KMT2A** | 11q23.3 | 6 (4.48%) | 5 (3.73%) |  | 0.26 | 1.00 | 1.00 | (A) Metastasis |
| **NSD3** | 8p11.23 | 6 (5.04%) | 5 (4.35%) |  | 0.21 | 1.00 | 1.00 | (A) Metastasis |
| **ERBB4** | 2q34 | 6 (4.48%) | 7 (5.22%) |  | -0.22 | 1.00 | 1.00 | (B) Primary |
| **AR** | Xq12 | 6 (4.48%) | 6 (4.48%) |  | - | 1.00 | 1.00 | (B) Primary |
| **CCND3** | 6p21.1 | 6 (4.48%) | 6 (4.48%) |  | - | 1.00 | 1.00 | (B) Primary |
| **PTPRS** | 19p13.3 | 6 (6.06%) | 6 (6.06%) |  | - | 1.00 | 1.00 | (B) Primary |
| **AGO2** | 8q24.3 | 5 (5.95%) | 1 (1.23%) |  | Şub.27 | 0.211 | 1.00 | (A) Metastasis |
| **INSR** | 19p13.2 | 5 (5.05%) | 1 (1.01%) |  | Şub.32 | 0.212 | 1.00 | (A) Metastasis |
| **BCL2L1** | 20q11.21 | 5 (3.73%) | 1 (0.75%) |  | Şub.32 | 0.213 | 1.00 | (A) Metastasis |
| **IKZF1** | 7p12.2 | 5 (3.73%) | 1 (0.75%) |  | Şub.32 | 0.213 | 1.00 | (A) Metastasis |
| **KMT2C** | 7q36.1 | 5 (5.05%) | 2 (2.02%) |  | Oca.32 | 0.282 | 1.00 | (A) Metastasis |
| **MAP3K13** | 3q27.2 | 5 (5.00%) | 2 (2.00%) |  | Oca.32 | 0.445 | 1.00 | (A) Metastasis |
| **MGA** | 15q15.1 | 5 (3.91%) | 2 (1.60%) |  | Oca.29 | 0.447 | 1.00 | (A) Metastasis |
| **FOXL2** | 3q22.3 | 5 (3.73%) | 2 (1.50%) |  | Oca.31 | 0.447 | 1.00 | (A) Metastasis |
| **RAD52** | 12p13.33 | 5 (3.73%) | 2 (1.50%) |  | Oca.31 | 0.447 | 1.00 | (A) Metastasis |
| **CDKN1B** | 12p13.1 | 5 (3.73%) | 2 (1.49%) |  | Oca.32 | 0.447 | 1.00 | (A) Metastasis |
| **SDHA** | 5p15.33 | 5 (3.73%) | 2 (1.49%) |  | Oca.32 | 0.447 | 1.00 | (A) Metastasis |
| **EPHA5** | 4q13.1-q13.2 | 5 (5.05%) | 3 (3.00%) |  | 0.75 | 0.498 | 1.00 | (A) Metastasis |
| **DOT1L** | 19p13.3 | 5 (5.00%) | 3 (3.00%) |  | 0.74 | 0.500 | 1.00 | (A) Metastasis |
| **KMT2B** | 19q13.12 | 5 (5.95%) | 3 (3.70%) |  | 0.68 | 0.720 | 1.00 | (A) Metastasis |
| **DNMT1** | 19p13.2 | 5 (5.05%) | 3 (3.03%) |  | 0.74 | 0.721 | 1.00 | (A) Metastasis |
| **FOXA1** | 14q21.1 | 5 (3.76%) | 3 (2.27%) |  | 0.73 | 0.722 | 1.00 | (A) Metastasis |
| **PIM1** | 6p21.2 | 5 (3.73%) | 3 (2.24%) |  | 0.74 | 0.722 | 1.00 | (A) Metastasis |
| **BRIP1** | 17q23.2 | 5 (3.73%) | 7 (5.22%) |  | -0.49 | 0.769 | 1.00 | (B) Primary |
| **ALK** | 2p23.2-p23.1 | 5 (3.73%) | 5 (3.73%) |  | - | 1.00 | 1.00 | (B) Primary |
| **NOTCH2** | 1p12 | 5 (3.73%) | 5 (3.73%) |  | - | 1.00 | 1.00 | (B) Primary |
| **NTRK3** | 15q25.3 | 5 (3.73%) | 5 (3.73%) |  | - | 1.00 | 1.00 | (B) Primary |
| **PBRM1** | 3p21.1 | 5 (3.73%) | 5 (3.73%) |  | - | 1.00 | 1.00 | (B) Primary |
| **RET** | 10q11.21 | 5 (3.73%) | 5 (3.73%) |  | - | 1.00 | 1.00 | (B) Primary |
| **ROS1** | 6q22.1 | 5 (3.73%) | 5 (3.73%) |  | - | 1.00 | 1.00 | (B) Primary |
| **CTNNB1** | 3p22.1 | 5 (3.73%) | 6 (4.48%) |  | -0.26 | 1.00 | 1.00 | (B) Primary |
| **ATR** | 3q23 | 5 (3.73%) | 4 (3.01%) |  | 0.31 | 1.00 | 1.00 | (A) Metastasis |
| **RICTOR** | 5p13.1 | 5 (3.73%) | 5 (3.76%) |  | -0.01 | 1.00 | 1.00 | (B) Primary |
| **FLT4** | 5q35.3 | 5 (3.76%) | 6 (4.51%) |  | -0.26 | 1.00 | 1.00 | (B) Primary |
| **PAK5** | 20p12.2 | 4 (4.04%) | 0 (0.00%) |  | >10 | 0.121 | 1.00 | (A) Metastasis |
| **ATRX** | Xq21.1 | 4 (2.99%) | 0 (0.00%) |  | >10 | 0.122 | 1.00 | (A) Metastasis |

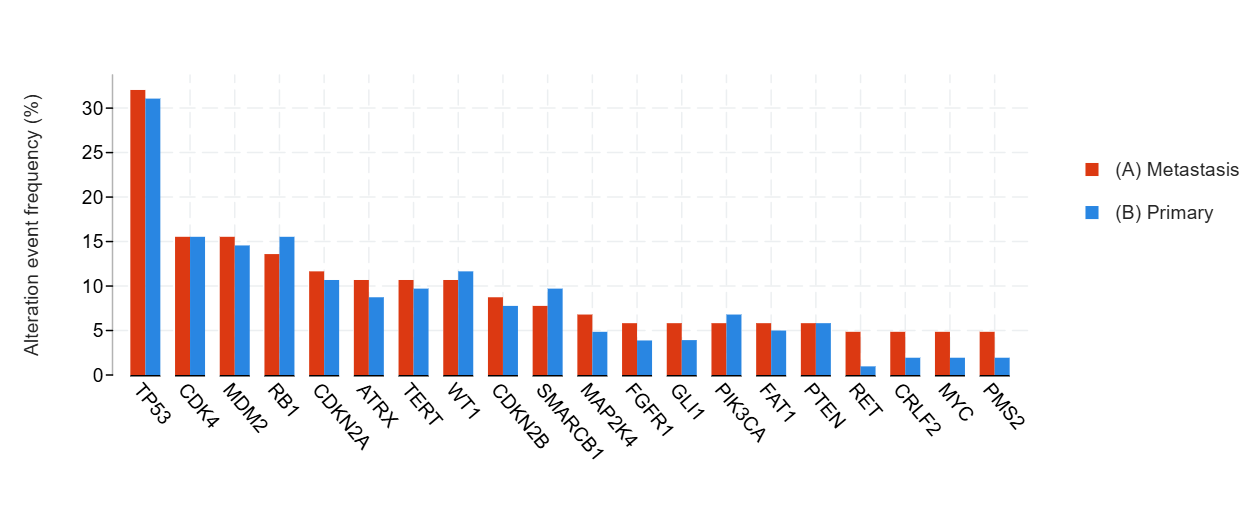

Figure S14. Distribution of the Most Frequently Mutated Genes in Primary and Metastatic Soft Tissue Sarcoma

**Table S15.** **Gene-Level Mutation Frequencies in Primary Versus Metastatic Soft Tissue Sarcoma**

| **Gene** | **Cytoband** | **(A) Metastasis** | **(B) Primary** | **Co-occurrence Pattern** | **Log2 Ratio** | **p-Value** | **q-Value** | **Enriched in** |
| --- | --- | --- | --- | --- | --- | --- | --- | --- |
| **TP53** | 17p13.1 | 33 (32.04%) | 32 (31.07%) |  | 0.04 | 1.00 | 1.00 | (A) Metastasis |
| **CDK4** | 12q14.1 | 16 (15.53%) | 16 (15.53%) |  | - | 1.00 | 1.00 | (B) Primary |
| **MDM2** | 12q15 | 16 (15.53%) | 15 (14.56%) |  | 0.09 | 1.00 | 1.00 | (A) Metastasis |
| **RB1** | 13q14.2 | 14 (13.59%) | 16 (15.53%) |  | -0.19 | 0.844 | 1.00 | (B) Primary |
| **CDKN2A** | 9p21.3 | 12 (11.65%) | 11 (10.68%) |  | 0.13 | 1.00 | 1.00 | (A) Metastasis |
| **ATRX** | Xq21.1 | 11 (10.68%) | 9 (8.74%) |  | 0.29 | 0.815 | 1.00 | (A) Metastasis |
| **TERT** | 5p15.33 | 11 (10.68%) | 10 (9.71%) |  | 0.14 | 1.00 | 1.00 | (A) Metastasis |
| **WT1** | 11p13 | 11 (10.68%) | 12 (11.65%) |  | -0.13 | 1.00 | 1.00 | (B) Primary |
| **CDKN2B** | 9p21.3 | 9 (8.74%) | 8 (7.77%) |  | 0.17 | 1.00 | 1.00 | (A) Metastasis |
| **SMARCB1** | 22q11.23\|22q11 | 8 (7.77%) | 10 (9.71%) |  | -0.32 | 0.806 | 1.00 | (B) Primary |
| **MAP2K4** | 17p12 | 7 (6.80%) | 5 (4.85%) |  | 0.49 | 0.570 | 1.00 | (A) Metastasis |
| **FGFR1** | 8p11.23 | 6 (5.83%) | 4 (3.88%) |  | 0.58 | 0.540 | 1.00 | (A) Metastasis |
| **GLI1** | 12q13.3 | 6 (5.83%) | 4 (3.92%) |  | 0.57 | 0.748 | 1.00 | (A) Metastasis |
| **PIK3CA** | 3q26.32 | 6 (5.83%) | 7 (6.80%) |  | -0.22 | 0.784 | 1.00 | (B) Primary |
| **FAT1** | 4q35.2 | 6 (5.83%) | 5 (5.00%) |  | 0.22 | 1.00 | 1.00 | (A) Metastasis |
| **PTEN** | 10q23.31 | 6 (5.83%) | 6 (5.83%) |  | - | 1.00 | 1.00 | (B) Primary |
| **RET** | 10q11.21 | 5 (4.85%) | 1 (0.97%) |  | Şub.32 | 0.212 | 1.00 | (A) Metastasis |
| **CRLF2** | Xp22.33 and Yp11.2 | 5 (4.85%) | 2 (1.94%) |  | Oca.32 | 0.445 | 1.00 | (A) Metastasis |
| **MYC** | 8q24.21 | 5 (4.85%) | 2 (1.94%) |  | Oca.32 | 0.445 | 1.00 | (A) Metastasis |
| **PMS2** | 7p22.1 | 5 (4.85%) | 2 (1.94%) |  | Oca.32 | 0.445 | 1.00 | (A) Metastasis |
| **CARD11** | 7p22.2 | 5 (4.85%) | 3 (2.91%) |  | 0.74 | 0.721 | 1.00 | (A) Metastasis |
| **IGF1R** | 15q26.3 | 5 (4.85%) | 3 (2.91%) |  | 0.74 | 0.721 | 1.00 | (A) Metastasis |
| **TSC2** | 16p13.3 | 5 (4.85%) | 3 (2.91%) |  | 0.74 | 0.721 | 1.00 | (A) Metastasis |
| **NAB2** | 12q13.3 | 5 (4.85%) | 4 (3.88%) |  | 0.32 | 1.00 | 1.00 | (A) Metastasis |
| **MED12** | Xq13.1 | 5 (4.85%) | 4 (4.00%) |  | 0.28 | 1.00 | 1.00 | (A) Metastasis |
| **LATS1** | 6q25.1 | 4 (4.08%) | 1 (1.02%) |  | 2.00 | 0.369 | 1.00 | (A) Metastasis |
| **ESR1** | 6q25.1-q25.2 | 4 (3.88%) | 1 (0.97%) |  | 2.00 | 0.369 | 1.00 | (A) Metastasis |
| **SMARCA4** | 19p13.2 | 4 (3.88%) | 1 (0.97%) |  | 2.00 | 0.369 | 1.00 | (A) Metastasis |
| **ZRSR2** | Xp22.2 | 4 (3.88%) | 1 (0.98%) |  | Oca.99 | 0.369 | 1.00 | (A) Metastasis |
| **ROS1** | 6q22.1 | 4 (3.88%) | 7 (6.80%) |  | -0.81 | 0.537 | 1.00 | (B) Primary |
| **RAD21** | 8q24.11 | 4 (3.88%) | 2 (1.96%) |  | 0.99 | 0.683 | 1.00 | (A) Metastasis |
| **KDM5A** | 12p13.33 | 4 (3.88%) | 2 (2.00%) |  | 0.96 | 0.683 | 1.00 | (A) Metastasis |
| **RAC1** | 7p22.1 | 4 (3.88%) | 2 (2.00%) |  | 0.96 | 0.683 | 1.00 | (A) Metastasis |
| **RBM10** | Xp11.3 | 4 (3.88%) | 2 (2.00%) |  | 0.96 | 0.683 | 1.00 | (A) Metastasis |
| **EPHA5** | 4q13.1-q13.2 | 4 (4.08%) | 3 (2.97%) |  | 0.46 | 0.718 | 1.00 | (A) Metastasis |
| **ARID1B** | 6q25.3 | 4 (3.88%) | 3 (2.91%) |  | 0.42 | 1.00 | 1.00 | (A) Metastasis |
| **CCND2** | 12p13.32 | 4 (3.88%) | 3 (2.91%) |  | 0.42 | 1.00 | 1.00 | (A) Metastasis |
| **KMT2D** | 12q13.12 | 4 (3.88%) | 3 (2.91%) |  | 0.42 | 1.00 | 1.00 | (A) Metastasis |
| **NOTCH1** | 9q34.3 | 4 (3.88%) | 3 (2.91%) |  | 0.42 | 1.00 | 1.00 | (A) Metastasis |
| **AGO2** | 8q24.3 | 4 (4.17%) | 3 (3.49%) |  | 0.26 | 1.00 | 1.00 | (A) Metastasis |
| **CYSLTR2** | 13q14.2 | 4 (4.17%) | 3 (3.49%) |  | 0.26 | 1.00 | 1.00 | (A) Metastasis |
| **PREX2** | 8q13.2 | 4 (4.17%) | 3 (3.49%) |  | 0.26 | 1.00 | 1.00 | (A) Metastasis |
| **NSD3** | 8p11.23 | 4 (3.96%) | 4 (4.55%) |  | -0.20 | 1.00 | 1.00 | (B) Primary |
| **CRKL** | 22q11.21 | 4 (3.88%) | 4 (3.88%) |  | - | 1.00 | 1.00 | (B) Primary |
| **DICER1** | 14q32.13 | 4 (3.88%) | 4 (3.88%) |  | - | 1.00 | 1.00 | (B) Primary |
| **MAPK1** | 22q11.22 | 4 (3.88%) | 4 (3.88%) |  | - | 1.00 | 1.00 | (B) Primary |
| **FLCN** | 17p11.2 | 4 (3.88%) | 5 (4.85%) |  | -0.32 | 1.00 | 1.00 | (B) Primary |
| **NF1** | 17q11.2 | 4 (3.88%) | 5 (4.85%) |  | -0.32 | 1.00 | 1.00 | (B) Primary |
| **PDCD1** | 2q37.3 | 4 (4.08%) | 4 (4.08%) |  | - | 1.00 | 1.00 | (B) Primary |
| **CARM1** | 19p13.2 | 3 (3.13%) | 0 (0.00%) |  | >10 | 0.248 | 1.00 | (A) Metastasis |
| **ZFHX3** | 16q22.2-q22.3 | 3 (3.06%) | 1 (1.03%) |  | Oca.57 | 0.621 | 1.00 | (A) Metastasis |
| **CD276** | 15q24.1 | 3 (3.06%) | 1 (1.02%) |  | Oca.58 | 0.621 | 1.00 | (A) Metastasis |
| **DDR2** | 1q23.3 | 3 (2.91%) | 1 (0.97%) |  | Oca.58 | 0.621 | 1.00 | (A) Metastasis |
| **ERBB4** | 2q34 | 3 (2.91%) | 1 (0.97%) |  | Oca.58 | 0.621 | 1.00 | (A) Metastasis |
| **KDM6A** | Xp11.3 | 3 (2.91%) | 1 (0.97%) |  | Oca.58 | 0.621 | 1.00 | (A) Metastasis |
| **SDHA** | 5p15.33 | 3 (2.91%) | 1 (0.98%) |  | Oca.57 | 0.621 | 1.00 | (A) Metastasis |
| **ASXL1** | 20q11.21 | 3 (2.91%) | 2 (1.94%) |  | 0.58 | 0.684 | 1.00 | (A) Metastasis |
| **BRAF** | 7q34 | 3 (2.91%) | 2 (1.94%) |  | 0.58 | 0.684 | 1.00 | (A) Metastasis |
| **CDKN2C** | 1p32.3 | 3 (2.91%) | 2 (1.94%) |  | 0.58 | 0.684 | 1.00 | (A) Metastasis |
| **ERBB3** | 12q13.2 | 3 (2.91%) | 2 (1.94%) |  | 0.58 | 0.684 | 1.00 | (A) Metastasis |

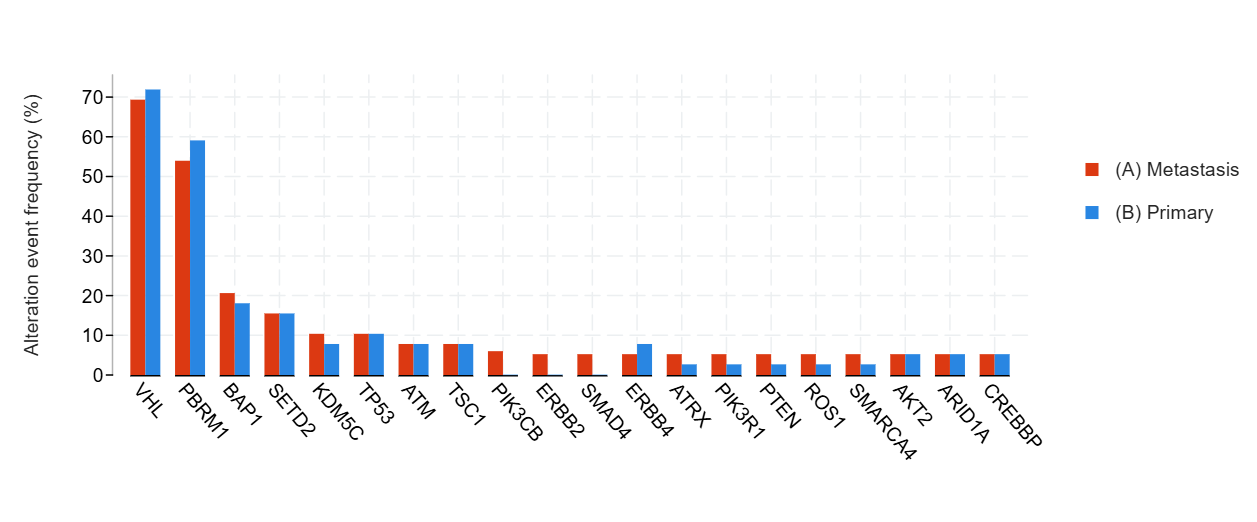

**Figure S15.** Distribution of the Most Frequently Mutated Genes in Primary and Metastatic Clear Cell RCC

**Table S16.** **Gene-Level Mutation Frequencies in Primary Versus Metastatic Clear Cell RCC**

| **ene** | **Cytoband** | **(A) Metastasis** | **(B) Primary** | **Co-occurrence Pattern** | **Log2 Ratio** | **p-Value** | **q-Value** | **Enriched in** |
| --- | --- | --- | --- | --- | --- | --- | --- | --- |
| **VHL** | 3p25.3 | 27 (69.23%) | 28 (71.79%) |  | -0.05 | 1.00 | 1.00 | (B) Primary |
| **PBRM1** | 3p21.1 | 21 (53.85%) | 23 (58.97%) |  | -0.13 | 0.820 | 1.00 | (B) Primary |
| **BAP1** | 3p21.1 | 8 (20.51%) | 7 (17.95%) |  | 0.19 | 1.00 | 1.00 | (A) Metastasis |
| **SETD2** | 3p21.31 | 6 (15.38%) | 6 (15.38%) |  | - | 1.00 | 1.00 | (B) Primary |
| **KDM5C** | Xp11.22 | 4 (10.26%) | 3 (7.69%) |  | 0.42 | 1.00 | 1.00 | (A) Metastasis |
| **TP53** | 17p13.1 | 4 (10.26%) | 4 (10.26%) |  | - | 1.00 | 1.00 | (B) Primary |
| **ATM** | 11q22.3 | 3 (7.69%) | 3 (7.69%) |  | - | 1.00 | 1.00 | (B) Primary |
| **TSC1** | 9q34.13 | 3 (7.69%) | 3 (7.69%) |  | - | 1.00 | 1.00 | (B) Primary |
| **PIK3CB** | 3q22.3 | 2 (5.88%) | 0 (0.00%) |  | >10 | 0.493 | 1.00 | (A) Metastasis |
| **ERBB2** | 17q12 | 2 (5.13%) | 0 (0.00%) |  | >10 | 0.494 | 1.00 | (A) Metastasis |
| **SMAD4** | 18q21.2 | 2 (5.13%) | 0 (0.00%) |  | >10 | 0.494 | 1.00 | (A) Metastasis |
| **ERBB4** | 2q34 | 2 (5.13%) | 3 (7.69%) |  | -0.58 | 1.00 | 1.00 | (B) Primary |
| **ATRX** | Xq21.1 | 2 (5.13%) | 1 (2.56%) |  | 1.00 | 1.00 | 1.00 | (A) Metastasis |
| **PIK3R1** | 5q13.1 | 2 (5.13%) | 1 (2.56%) |  | 1.00 | 1.00 | 1.00 | (A) Metastasis |
| **PTEN** | 10q23.31 | 2 (5.13%) | 1 (2.56%) |  | 1.00 | 1.00 | 1.00 | (A) Metastasis |
| **ROS1** | 6q22.1 | 2 (5.13%) | 1 (2.56%) |  | 1.00 | 1.00 | 1.00 | (A) Metastasis |
| **SMARCA4** | 19p13.2 | 2 (5.13%) | 1 (2.56%) |  | 1.00 | 1.00 | 1.00 | (A) Metastasis |
| **AKT2** | 19q13.2 | 2 (5.13%) | 2 (5.13%) |  | - | 1.00 | 1.00 | (B) Primary |
| **ARID1A** | 1p36.11 | 2 (5.13%) | 2 (5.13%) |  | - | 1.00 | 1.00 | (B) Primary |
| **CREBBP** | 16p13.3 | 2 (5.13%) | 2 (5.13%) |  | - | 1.00 | 1.00 | (B) Primary |
| **MTOR** | 1p36.22 | 2 (5.13%) | 2 (5.13%) |  | - | 1.00 | 1.00 | (B) Primary |
| **NF2** | 22q12.2 | 2 (5.13%) | 2 (5.13%) |  | - | 1.00 | 1.00 | (B) Primary |
| **SF3B1** | 2q33.1 | 2 (5.13%) | 2 (5.13%) |  | - | 1.00 | 1.00 | (B) Primary |
| **TERT** | 5p15.33 | 2 (5.26%) | 2 (5.26%) |  | - | 1.00 | 1.00 | (B) Primary |
| **FAT1** | 4q35.2 | 2 (5.26%) | 2 (5.56%) |  | -0.08 | 1.00 | 1.00 | (B) Primary |
| **NSD1** | 5q35.3 | 2 (5.26%) | 2 (5.56%) |  | -0.08 | 1.00 | 1.00 | (B) Primary |
| **CUL3** | 2q36.2 | 2 (5.88%) | 2 (5.88%) |  | - | 1.00 | 1.00 | (B) Primary |
| **RASA1** | 5q14.3 | 1 (2.63%) | 2 (5.56%) |  | -1.08 | 0.610 | 1.00 | (B) Primary |
| **CDKN2A** | 9p21.3 | 1 (2.56%) | 3 (7.69%) |  | -1.58 | 0.615 | 1.00 | (B) Primary |
| **SOS1** | 2p22.1 | 1 (3.45%) | 1 (4.00%) |  | -0.21 | 1.00 | 1.00 | (B) Primary |
| **BRCA2** | 13q13.1 | 1 (2.56%) | 0 (0.00%) |  | >10 | 1.00 | 1.00 | (A) Metastasis |
| **FANCA** | 16q24.3 | 1 (2.56%) | 0 (0.00%) |  | >10 | 1.00 | 1.00 | (A) Metastasis |
| **PAX8** | 2q14.1 | 1 (2.56%) | 0 (0.00%) |  | >10 | 1.00 | 1.00 | (A) Metastasis |
| **SDHA** | 5p15.33 | 1 (2.56%) | 0 (0.00%) |  | >10 | 1.00 | 1.00 | (A) Metastasis |
| **FLT3** | 13q12.2 | 1 (2.56%) | 2 (5.13%) |  | -1.00 | 1.00 | 1.00 | (B) Primary |
| **POLD1** | 19q13.33 | 1 (2.56%) | 0 (0.00%) |  | >10 | 1.00 | 1.00 | (A) Metastasis |
| **HGF** | 7q21.11 | 1 (2.94%) | 1 (2.94%) |  | - | 1.00 | 1.00 | (B) Primary |
| **PDCD1** | 2q37.3 | 1 (2.94%) | 1 (2.94%) |  | - | 1.00 | 1.00 | (B) Primary |
| **RAD54L** | 1p34.1 | 1 (2.94%) | 1 (2.94%) |  | - | 1.00 | 1.00 | (B) Primary |
| **B2M** | 15q21.1 | 1 (2.63%) | 1 (2.63%) |  | - | 1.00 | 1.00 | (B) Primary |

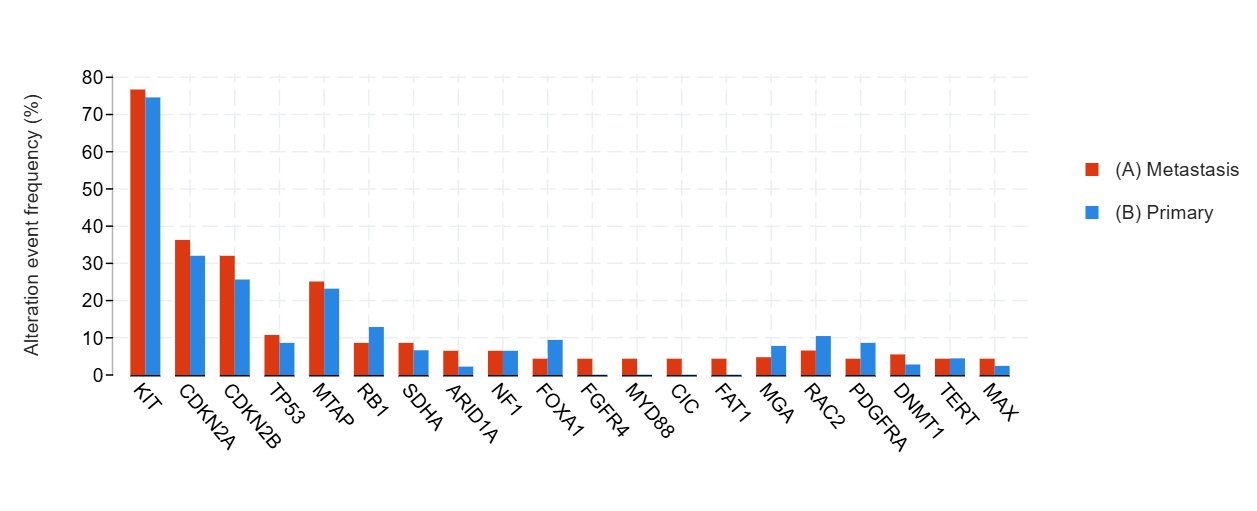

**Figure S16.** Distribution of the Most Frequently Mutated Genes in Primary and Metastatic Gastrointestinal Stromal Tumor

**Table S17.** **Gene-Level Mutation Frequencies in Primary Versus Metastatic Gastrointestinal Stromal Tumor**

| **Gene** | **Cytoband** | **(A) Metastasis** | **(B) Primary** | **Co-occurrence Pattern** | **Log2 Ratio** | **p-Value** | **q-Value** | **Enriched in** |
| --- | --- | --- | --- | --- | --- | --- | --- | --- |
| **KIT** | 4q12 | 36 (76.60%) | 35 (74.47%) |  | 0.04 | 1.00 | 1.00 | (A) Metastasis |
| **CDKN2A** | 9p21.3 | 17 (36.17%) | 15 (31.91%) |  | 0.18 | 0.828 | 1.00 | (A) Metastasis |
| **CDKN2B** | 9p21.3 | 15 (31.91%) | 12 (25.53%) |  | 0.32 | 0.649 | 1.00 | (A) Metastasis |
| **TP53** | 17p13.1 | 5 (10.64%) | 4 (8.51%) |  | 0.32 | 1.00 | 1.00 | (A) Metastasis |
| **MTAP** | 9p21.3 | 5 (25.00%) | 3 (23.08%) |  | 0.12 | 1.00 | 1.00 | (A) Metastasis |
| **RB1** | 13q14.2 | 4 (8.51%) | 6 (12.77%) |  | -0.58 | 0.740 | 1.00 | (B) Primary |
| **SDHA** | 5p15.33 | 4 (8.51%) | 3 (6.52%) |  | 0.38 | 1.00 | 1.00 | (A) Metastasis |
| **ARID1A** | 1p36.11 | 3 (6.38%) | 1 (2.13%) |  | Oca.58 | 0.617 | 1.00 | (A) Metastasis |
| **NF1** | 17q11.2 | 3 (6.38%) | 3 (6.38%) |  | - | 1.00 | 1.00 | (B) Primary |
| **FOXA1** | 14q21.1 | 2 (4.26%) | 4 (9.30%) |  | -1.13 | 0.420 | 1.00 | (B) Primary |
| **FGFR4** | 5q35.2 | 2 (4.26%) | 0 (0.00%) |  | >10 | 0.495 | 1.00 | (A) Metastasis |
| **MYD88** | 3p22.2 | 2 (4.26%) | 0 (0.00%) |  | >10 | 0.495 | 1.00 | (A) Metastasis |
| **CIC** | 19q13.2 | 2 (4.26%) | 0 (0.00%) |  | >10 | 0.495 | 1.00 | (A) Metastasis |
| **FAT1** | 4q35.2 | 2 (4.26%) | 0 (0.00%) |  | >10 | 0.495 | 1.00 | (A) Metastasis |
| **MGA** | 15q15.1 | 2 (4.65%) | 3 (7.69%) |  | -0.73 | 0.665 | 1.00 | (B) Primary |
| **RAC2** | 22q13.1 | 2 (6.45%) | 3 (10.34%) |  | -0.68 | 0.666 | 1.00 | (B) Primary |
| **PDGFRA** | 4q12 | 2 (4.26%) | 4 (8.51%) |  | -1.00 | 0.677 | 1.00 | (B) Primary |
| **DNMT1** | 19p13.2 | 2 (5.41%) | 1 (2.70%) |  | 1.00 | 1.00 | 1.00 | (A) Metastasis |
| **TERT** | 5p15.33 | 2 (4.26%) | 2 (4.35%) |  | -0.03 | 1.00 | 1.00 | (B) Primary |
| **MAX** | 14q23.3 | 2 (4.26%) | 1 (2.33%) |  | 0.87 | 1.00 | 1.00 | (A) Metastasis |
| **RNF43** | 17q22 | 2 (4.26%) | 2 (4.55%) |  | -0.10 | 1.00 | 1.00 | (B) Primary |
| **NOTCH3** | 19p13.12 | 2 (4.26%) | 1 (2.27%) |  | 0.90 | 1.00 | 1.00 | (A) Metastasis |
| **FLT1** | 13q12.3 | 2 (4.26%) | 2 (4.26%) |  | - | 1.00 | 1.00 | (B) Primary |
| **KMT2D** | 12q13.12 | 2 (4.26%) | 2 (4.26%) |  | - | 1.00 | 1.00 | (B) Primary |
| **MTOR** | 1p36.22 | 2 (4.26%) | 2 (4.26%) |  | - | 1.00 | 1.00 | (B) Primary |
| **NTRK3** | 15q25.3 | 2 (4.26%) | 2 (4.26%) |  | - | 1.00 | 1.00 | (B) Primary |
| **ANKRD11** | 16q24.3 | 2 (5.88%) | 1 (3.03%) |  | 0.96 | 1.00 | 1.00 | (A) Metastasis |
| **CREBBP** | 16p13.3 | 2 (4.26%) | 3 (6.38%) |  | -0.58 | 1.00 | 1.00 | (B) Primary |
| **PTEN** | 10q23.31 | 2 (4.26%) | 3 (6.38%) |  | -0.58 | 1.00 | 1.00 | (B) Primary |
| **BRD4** | 19p13.12 | 2 (4.26%) | 1 (2.13%) |  | 1.00 | 1.00 | 1.00 | (A) Metastasis |
| **FANCA** | 16q24.3 | 2 (4.26%) | 1 (2.13%) |  | 1.00 | 1.00 | 1.00 | (A) Metastasis |
| **NOTCH1** | 9q34.3 | 2 (4.26%) | 1 (2.13%) |  | 1.00 | 1.00 | 1.00 | (A) Metastasis |
| **SMC1A** | Xp11.22 | 1 (100.00%) | 0 (0.00%) |  | >10 | 0.200 | 1.00 | (A) Metastasis |
| **KLF4** | 9q31.2 | 1 (2.13%) | 2 (4.65%) |  | -1.13 | 0.604 | 1.00 | (B) Primary |
| **ARID2** | 12q12 | 1 (2.13%) | 3 (6.38%) |  | -1.58 | 0.617 | 1.00 | (B) Primary |
| **NKX2-1** | 14q13.3 | 1 (2.13%) | 3 (6.38%) |  | -1.58 | 0.617 | 1.00 | (B) Primary |
| **ERCC3** | 2q14.3 | 1 (2.13%) | 1 (2.17%) |  | -0.03 | 1.00 | 1.00 | (B) Primary |
| **KEAP1** | 19p13.2 | 1 (2.13%) | 1 (2.17%) |  | -0.03 | 1.00 | 1.00 | (B) Primary |
| **PBRM1** | 3p21.1 | 1 (2.13%) | 1 (2.17%) |  | -0.03 | 1.00 | 1.00 | (B) Primary |
| **SOX9** | 17q24.3 | 1 (2.13%) | 1 (2.17%) |  | -0.03 | 1.00 | 1.00 | (B) Primary |

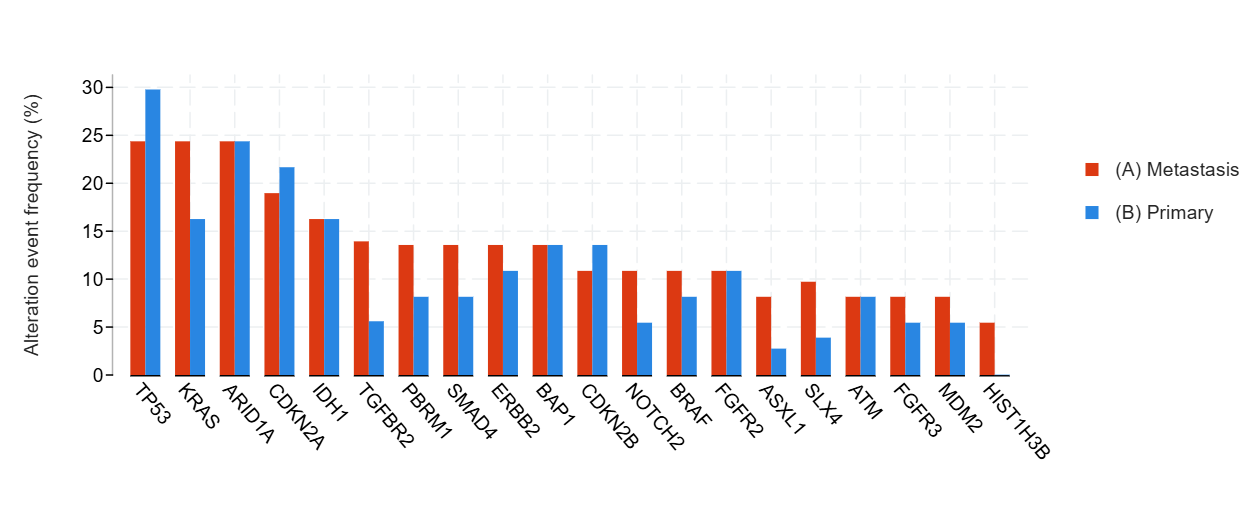

**Figure S17.** Distribution of the Most Frequently Mutated Genes in Primary and Metastatic Biliary Cancer

**Table S 18.** **Gene-Level Mutation Frequencies in Primary Versus Metastatic Biliary Cancer**

| **Gene** | **Cytoband** | **(A) Metastasis** | **(B) Primary** | **Co-occurrence Pattern** | **Log2 Ratio** | **p-Value** | **q-Value** | **Enriched in** |
| --- | --- | --- | --- | --- | --- | --- | --- | --- |
| **KRAS** | 12p12.1 | 9 (24.32%) | 6 (16.22%) |  | 0.58 | 0.564 | 1.00 | (A) Metastasis |
| **TP53** | 17p13.1 | 9 (24.32%) | 11 (29.73%) |  | -0.29 | 0.794 | 1.00 | (B) Primary |
| **ARID1A** | 1p36.11 | 9 (24.32%) | 9 (24.32%) |  | - | 1.00 | 1.00 | (B) Primary |
| **CDKN2A** | 9p21.3 | 7 (18.92%) | 8 (21.62%) |  | -0.19 | 1.00 | 1.00 | (B) Primary |
| **IDH1** | 2q34 | 6 (16.22%) | 6 (16.22%) |  | - | 1.00 | 1.00 | (B) Primary |
| **TGFBR2** | 3p24.1 | 5 (13.89%) | 2 (5.56%) |  | Oca.32 | 0.429 | 1.00 | (A) Metastasis |
| **PBRM1** | 3p21.1 | 5 (13.51%) | 3 (8.11%) |  | 0.74 | 0.711 | 1.00 | (A) Metastasis |
| **SMAD4** | 18q21.2 | 5 (13.51%) | 3 (8.11%) |  | 0.74 | 0.711 | 1.00 | (A) Metastasis |
| **ERBB2** | 17q12 | 5 (13.51%) | 4 (10.81%) |  | 0.32 | 1.00 | 1.00 | (A) Metastasis |
| **BAP1** | 3p21.1 | 5 (13.51%) | 5 (13.51%) |  | - | 1.00 | 1.00 | (B) Primary |
| **NOTCH2** | 1p12 | 4 (10.81%) | 2 (5.41%) |  | 1.00 | 0.674 | 1.00 | (A) Metastasis |
| **BRAF** | 7q34 | 4 (10.81%) | 3 (8.11%) |  | 0.42 | 1.00 | 1.00 | (A) Metastasis |
| **FGFR2** | 10q26.13 | 4 (10.81%) | 4 (10.81%) |  | - | 1.00 | 1.00 | (B) Primary |
| **CDKN2B** | 9p21.3 | 4 (10.81%) | 5 (13.51%) |  | -0.32 | 1.00 | 1.00 | (B) Primary |
| **ASXL1** | 20q11.21 | 3 (8.11%) | 1 (2.70%) |  | Oca.58 | 0.615 | 1.00 | (A) Metastasis |
| **SLX4** | 16p13.3 | 3 (9.68%) | 1 (3.85%) |  | Oca.33 | 0.617 | 1.00 | (A) Metastasis |
| **ATM** | 11q22.3 | 3 (8.11%) | 3 (8.11%) |  | - | 1.00 | 1.00 | (B) Primary |
| **FGFR3** | 4p16.3 | 3 (8.11%) | 2 (5.41%) |  | 0.58 | 1.00 | 1.00 | (A) Metastasis |
| **MDM2** | 12q15 | 3 (8.11%) | 2 (5.41%) |  | 0.58 | 1.00 | 1.00 | (A) Metastasis |
| **HIST1H3B** | 6p22.2 | 2 (5.41%) | 0 (0.00%) |  | >10 | 0.493 | 1.00 | (A) Metastasis |
| **MED12** | Xq13.1 | 2 (5.41%) | 0 (0.00%) |  | >10 | 0.493 | 1.00 | (A) Metastasis |
| **ARAF** | Xp11.3 | 2 (5.41%) | 0 (0.00%) |  | >10 | 0.493 | 1.00 | (A) Metastasis |
| **ERBB4** | 2q34 | 2 (5.41%) | 0 (0.00%) |  | >10 | 0.493 | 1.00 | (A) Metastasis |
| **GNAS** | 20q13.32 | 2 (5.41%) | 0 (0.00%) |  | >10 | 0.493 | 1.00 | (A) Metastasis |
| **STAG2** | Xq25 | 2 (5.41%) | 0 (0.00%) |  | >10 | 0.493 | 1.00 | (A) Metastasis |
| **SRC** | 20q11.23 | 2 (5.56%) | 1 (2.70%) |  | 1.Nis | 0.615 | 1.00 | (A) Metastasis |
| **AXL** | 19q13.2 | 2 (5.41%) | 1 (2.70%) |  | 1.00 | 1.00 | 1.00 | (A) Metastasis |
| **ETV6** | 12p13.2 | 2 (5.41%) | 1 (2.70%) |  | 1.00 | 1.00 | 1.00 | (A) Metastasis |
| **IDH2** | 15q26.1 | 2 (5.41%) | 1 (2.70%) |  | 1.00 | 1.00 | 1.00 | (A) Metastasis |
| **KEAP1** | 19p13.2 | 2 (5.41%) | 1 (2.70%) |  | 1.00 | 1.00 | 1.00 | (A) Metastasis |
| **MCL1** | 1q21.2 | 2 (5.41%) | 1 (2.70%) |  | 1.00 | 1.00 | 1.00 | (A) Metastasis |
| **PTEN** | 10q23.31 | 2 (5.41%) | 1 (2.70%) |  | 1.00 | 1.00 | 1.00 | (A) Metastasis |
| **AXIN1** | 16p13.3 | 2 (5.56%) | 2 (5.56%) |  | - | 1.00 | 1.00 | (B) Primary |
| **CTNNB1** | 3p22.1 | 2 (5.41%) | 3 (8.11%) |  | -0.58 | 1.00 | 1.00 | (B) Primary |
| **JAK1** | 1p31.3 | 2 (5.41%) | 1 (2.78%) |  | 0.96 | 1.00 | 1.00 | (A) Metastasis |
| **FGF4** | 11q13.3 | 2 (5.56%) | 1 (2.78%) |  | 1.00 | 1.00 | 1.00 | (A) Metastasis |
| **TOP1** | 20q12 | 2 (5.56%) | 1 (2.78%) |  | 1.00 | 1.00 | 1.00 | (A) Metastasis |
| **SETDB1** | 1q21.3 | 2 (13.33%) | 1 (9.09%) |  | 0.55 | 1.00 | 1.00 | (A) Metastasis |
| **ZFHX3** | 16q22.2-q22.3 | 2 (5.88%) | 2 (5.88%) |  | - | 1.00 | 1.00 | (B) Primary |
| **EPHA7** | 6q16.1 | 2 (5.88%) | 3 (8.57%) |  | -0.54 | 1.00 | 1.00 | (B) Primary |
| **APC** | 5q22.2 | 2 (5.41%) | 2 (5.41%) |  | - | 1.00 | 1.00 | (B) Primary |
| **EP300** | 22q13.2 | 2 (5.41%) | 2 (5.41%) |  | - | 1.00 | 1.00 | (B) Primary |
| **ERBB3** | 12q13.2 | 2 (5.41%) | 2 (5.41%) |  | - | 1.00 | 1.00 | (B) Primary |
| **KMT2D** | 12q13.12 | 2 (5.41%) | 2 (5.41%) |  | - | 1.00 | 1.00 | (B) Primary |
| **NTRK2** | 9q21.33 | 2 (5.41%) | 2 (5.41%) |  | - | 1.00 | 1.00 | (B) Primary |
| **SF3B1** | 2q33.1 | 2 (5.41%) | 2 (5.41%) |  | - | 1.00 | 1.00 | (B) Primary |
| **XPO1** | 2p15 | 2 (5.41%) | 2 (5.41%) |  | - | 1.00 | 1.00 | (B) Primary |
| **ELANE** | 19p13.3 | 1 (100.00%) | 0 (0.00%) |  | >10 | 0.0263 | 0.797 | (A) Metastasis |
| **HELQ** | 4q21.23 | 1 (100.00%) | 0 (0.00%) |  | >10 | 0.0263 | 0.797 | (A) Metastasis |
| **MCM8** | 20p12.3 | 1 (100.00%) | 0 (0.00%) |  | >10 | 0.0263 | 0.797 | (A) Metastasis |
| **POLH** | 6p21.1 | 1 (100.00%) | 0 (0.00%) |  | >10 | 0.0263 | 0.797 | (A) Metastasis |
| **TLX3** | 5q35.1 | 1 (100.00%) | 0 (0.00%) |  | >10 | 0.0263 | 0.797 | (A) Metastasis |
| **ALOX12B** | 17p13.1 | 1 (2.78%) | 0 (0.00%) |  | >10 | 0.493 | 1.00 | (A) Metastasis |
| **EPHA3** | 3p11.1 | 1 (2.78%) | 0 (0.00%) |  | >10 | 0.493 | 1.00 | (A) Metastasis |
| **AGO2** | 8q24.3 | 1 (3.33%) | 2 (7.69%) |  | -1.21 | 0.592 | 1.00 | (B) Primary |
| **KMT2C** | 7q36.1 | 1 (2.78%) | 3 (8.33%) |  | -1.58 | 0.614 | 1.00 | (B) Primary |
| **CDK12** | 17q12 | 1 (2.70%) | 2 (5.56%) |  | -1.04 | 0.615 | 1.00 | (B) Primary |
| **RIT1** | 1q22 | 1 (2.70%) | 2 (5.56%) |  | -1.04 | 0.615 | 1.00 | (B) Primary |
| **ARID2** | 12q12 | 1 (2.70%) | 3 (8.11%) |  | -1.58 | 0.615 | 1.00 | (B) Primary |
| **BRCA2** | 13q13.1 | 1 (2.70%) | 2 (5.41%) |  | -1.00 | 1.00 | 1.00 | (B) Primary |
| **MDM4** | 1q32.1 | 1 (2.70%) | 2 (5.41%) |  | -1.00 | 1.00 | 1.00 | (B) Primary |
| **SETD2** | 3p21.31 | 1 (2.70%) | 2 (5.41%) |  | -1.00 | 1.00 | 1.00 | (B) Primary |
| **HIST1H3D** | 6p22.2 | 1 (2.94%) | 1 (2.94%) |  | - | 1.00 | 1.00 | (B) Primary |
| **HIST1H3E** | 6p22.2 | 1 (2.94%) | 1 (2.94%) |  | - | 1.00 | 1.00 | (B) Primary |
| **HIST2H3C** | 1q21.2 | 1 (2.94%) | 1 (2.94%) |  | - | 1.00 | 1.00 | (B) Primary |
| **CEBPA** | 19q13.11 | 1 (2.86%) | 1 (2.86%) |  | - | 1.00 | 1.00 | (B) Primary |
| **TCF7L2** | 10q25.2-q25.3 | 1 (2.86%) | 1 (2.86%) |  | - | 1.00 | 1.00 | (B) Primary |
| **MALT1** | 18q21.32 | 1 (2.94%) | 2 (5.88%) |  | -1.00 | 1.00 | 1.00 | (B) Primary |
| **FGF19** | 11q13.3 | 1 (2.78%) | 1 (2.78%) |  | - | 1.00 | 1.00 | (B) Primary |
| **FGF3** | 11q13.3 | 1 (2.78%) | 1 (2.78%) |  | - | 1.00 | 1.00 | (B) Primary |
| **GRIN2A** | 16p13.2 | 1 (2.78%) | 1 (2.78%) |  | - | 1.00 | 1.00 | (B) Primary |
| **HIST1H2BD** | 6p22.2 | 1 (2.78%) | 1 (2.78%) |  | - | 1.00 | 1.00 | (B) Primary |
| **LATS2** | 13q12.11 | 1 (2.78%) | 1 (2.78%) |  | - | 1.00 | 1.00 | (B) Primary |
| **PIK3CD** | 1p36.22 | 1 (2.78%) | 1 (2.78%) |  | - | 1.00 | 1.00 | (B) Primary |
| **PTPRT** | 20q12-q13.11 | 1 (2.78%) | 1 (2.78%) |  | - | 1.00 | 1.00 | (B) Primary |
| **RYBP** | 3p13 | 1 (2.78%) | 1 (2.78%) |  | - | 1.00 | 1.00 | (B) Primary |
| **TBX3** | 12q24.21 | 1 (2.78%) | 1 (2.78%) |  | - | 1.00 | 1.00 | (B) Primary |
| **FAT1** | 4q35.2 | 1 (2.70%) | 0 (0.00%) |  | >10 | 1.00 | 1.00 | (A) Metastasis |
| **KLF4** | 9q31.2 | 1 (2.70%) | 0 (0.00%) |  | >10 | 1.00 | 1.00 | (A) Metastasis |
| **NOTCH3** | 19p13.12 | 1 (2.70%) | 0 (0.00%) |  | >10 | 1.00 | 1.00 | (A) Metastasis |

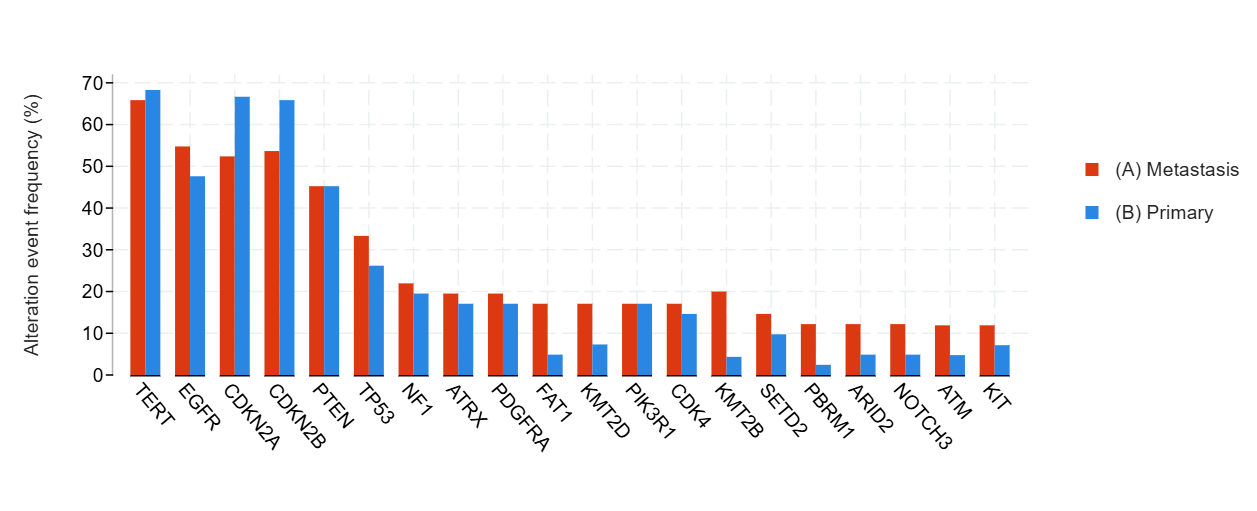

**Figure S18.** Distribution of the Most Frequently Mutated Genes in Primary and Metastatic Glioblastoma

**Table S19.** **Gene-Level Mutation Frequencies in Primary Versus Metastatic Glioblastoma**

| **Gene** | **Cytoband** | **(A) Metastasis** | **(B) Primary** | **Co-occurrence Pattern** | **Log2 Ratio** | **p-Value** | **q-Value** | **Enriched in** |
| --- | --- | --- | --- | --- | --- | --- | --- | --- |
| **TERT** | 5p15.33 | 27 (65.85%) | 28 (68.29%) |  | -0.05 | 1.00 | 1.00 | (B) Primary |
| **EGFR** | 7p11.2 | 23 (54.76%) | 20 (47.62%) |  | 0.20 | 0.663 | 1.00 | (A) Metastasis |
| **CDKN2A** | 9p21.3 | 22 (52.38%) | 28 (66.67%) |  | -0.35 | 0.266 | 1.00 | (B) Primary |
| **CDKN2B** | 9p21.3 | 22 (53.66%) | 27 (65.85%) |  | -0.30 | 0.368 | 1.00 | (B) Primary |
| **PTEN** | 10q23.31 | 19 (45.24%) | 19 (45.24%) |  | - | 1.00 | 1.00 | (B) Primary |
| **TP53** | 17p13.1 | 14 (33.33%) | 11 (26.19%) |  | 0.35 | 0.634 | 1.00 | (A) Metastasis |
| **NF1** | 17q11.2 | 9 (21.95%) | 8 (19.51%) |  | 0.17 | 1.00 | 1.00 | (A) Metastasis |
| **ATRX** | Xq21.1 | 8 (19.51%) | 7 (17.07%) |  | 0.19 | 1.00 | 1.00 | (A) Metastasis |
| **PDGFRA** | 4q12 | 8 (19.51%) | 7 (17.07%) |  | 0.19 | 1.00 | 1.00 | (A) Metastasis |
| **FAT1** | 4q35.2 | 7 (17.07%) | 2 (4.88%) |  | Oca.81 | 0.155 | 1.00 | (A) Metastasis |
| **KMT2D** | 12q13.12 | 7 (17.07%) | 3 (7.32%) |  | Oca.22 | 0.312 | 1.00 | (A) Metastasis |
| **PIK3R1** | 5q13.1 | 7 (17.07%) | 7 (17.07%) |  | - | 1.00 | 1.00 | (B) Primary |
| **CDK4** | 12q14.1 | 7 (17.07%) | 6 (14.63%) |  | 0.22 | 1.00 | 1.00 | (A) Metastasis |
| **KMT2B** | 19q13.12 | 6 (20.00%) | 1 (4.35%) |  | Şub.20 | 0.123 | 1.00 | (A) Metastasis |
| **SETD2** | 3p21.31 | 6 (14.63%) | 4 (9.76%) |  | 0.58 | 0.737 | 1.00 | (A) Metastasis |
| **PBRM1** | 3p21.1 | 5 (12.20%) | 1 (2.44%) |  | Şub.32 | 0.201 | 1.00 | (A) Metastasis |
| **ARID2** | 12q12 | 5 (12.20%) | 2 (4.88%) |  | Oca.32 | 0.432 | 1.00 | (A) Metastasis |
| **NOTCH3** | 19p13.12 | 5 (12.20%) | 2 (4.88%) |  | Oca.32 | 0.432 | 1.00 | (A) Metastasis |
| **ATM** | 11q22.3 | 5 (11.90%) | 2 (4.76%) |  | Oca.32 | 0.433 | 1.00 | (A) Metastasis |
| **KIT** | 4q12 | 5 (11.90%) | 3 (7.14%) |  | 0.74 | 0.713 | 1.00 | (A) Metastasis |
| **BRCA2** | 13q13.1 | 4 (9.76%) | 0 (0.00%) |  | >10 | 0.116 | 1.00 | (A) Metastasis |
| **MED12** | Xq13.1 | 4 (9.76%) | 0 (0.00%) |  | >10 | 0.116 | 1.00 | (A) Metastasis |
| **TSC2** | 16p13.3 | 4 (9.76%) | 0 (0.00%) |  | >10 | 0.116 | 1.00 | (A) Metastasis |
| **FBXW7** | 4q31.3 | 4 (9.52%) | 0 (0.00%) |  | >10 | 0.116 | 1.00 | (A) Metastasis |
| **SLX4** | 16p13.3 | 4 (10.81%) | 0 (0.00%) |  | >10 | 0.122 | 1.00 | (A) Metastasis |
| **DROSHA** | 5p13.3 | 4 (13.33%) | 0 (0.00%) |  | >10 | 0.124 | 1.00 | (A) Metastasis |
| **ARID1A** | 1p36.11 | 4 (9.76%) | 1 (2.44%) |  | 2.00 | 0.359 | 1.00 | (A) Metastasis |
| **KDM5C** | Xp11.22 | 4 (9.76%) | 1 (2.44%) |  | 2.00 | 0.359 | 1.00 | (A) Metastasis |
| **PMS2** | 7p22.1 | 4 (9.76%) | 1 (2.44%) |  | 2.00 | 0.359 | 1.00 | (A) Metastasis |
| **PIK3CB** | 3q22.3 | 4 (11.76%) | 2 (5.88%) |  | 1.00 | 0.673 | 1.00 | (A) Metastasis |
| **CREBBP** | 16p13.3 | 4 (9.76%) | 2 (4.88%) |  | 1.00 | 0.675 | 1.00 | (A) Metastasis |
| **FGFR4** | 5q35.2 | 4 (9.76%) | 2 (4.88%) |  | 1.00 | 0.675 | 1.00 | (A) Metastasis |
| **MDM4** | 1q32.1 | 4 (9.76%) | 2 (4.88%) |  | 1.00 | 0.675 | 1.00 | (A) Metastasis |
| **MSH2** | 2p21-p16.3 | 4 (9.76%) | 2 (4.88%) |  | 1.00 | 0.675 | 1.00 | (A) Metastasis |
| **POLE** | 12q24.33 | 4 (9.76%) | 2 (4.88%) |  | 1.00 | 0.675 | 1.00 | (A) Metastasis |
| **KDR** | 4q12 | 4 (9.52%) | 2 (4.76%) |  | 1.00 | 0.676 | 1.00 | (A) Metastasis |
| **NOTCH1** | 9q34.3 | 4 (9.52%) | 2 (4.76%) |  | 1.00 | 0.676 | 1.00 | (A) Metastasis |
| **ERBB3** | 12q13.2 | 4 (9.76%) | 4 (9.76%) |  | - | 1.00 | 1.00 | (B) Primary |
| **PIK3CA** | 3q26.32 | 4 (9.52%) | 4 (9.52%) |  | - | 1.00 | 1.00 | (B) Primary |
| **PTPN11** | 12q24.13 | 4 (9.52%) | 3 (7.14%) |  | 0.42 | 1.00 | 1.00 | (A) Metastasis |
| **RB1** | 13q14.2 | 4 (9.52%) | 3 (7.14%) |  | 0.42 | 1.00 | 1.00 | (A) Metastasis |
| **DNMT3B** | 20q11.21 | 3 (8.82%) | 0 (0.00%) |  | >10 | 0.239 | 1.00 | (A) Metastasis |
| **IRS2** | 13q34 | 3 (8.82%) | 0 (0.00%) |  | >10 | 0.239 | 1.00 | (A) Metastasis |
| **PIK3C2G** | 12p12.3 | 3 (8.82%) | 0 (0.00%) |  | >10 | 0.239 | 1.00 | (A) Metastasis |
| **RPS6KB2** | 11q13.2 | 3 (8.82%) | 0 (0.00%) |  | >10 | 0.239 | 1.00 | (A) Metastasis |
| **ARID1B** | 6q25.3 | 3 (7.32%) | 0 (0.00%) |  | >10 | 0.241 | 1.00 | (A) Metastasis |
| **ASXL1** | 20q11.21 | 3 (7.32%) | 0 (0.00%) |  | >10 | 0.241 | 1.00 | (A) Metastasis |
| **BRIP1** | 17q23.2 | 3 (7.32%) | 0 (0.00%) |  | >10 | 0.241 | 1.00 | (A) Metastasis |
| **CTCF** | 16q22.1 | 3 (7.32%) | 0 (0.00%) |  | >10 | 0.241 | 1.00 | (A) Metastasis |
| **ERCC4** | 16p13.12 | 3 (7.32%) | 0 (0.00%) |  | >10 | 0.241 | 1.00 | (A) Metastasis |
| **FLT1** | 13q12.3 | 3 (7.32%) | 0 (0.00%) |  | >10 | 0.241 | 1.00 | (A) Metastasis |
| **IGF1R** | 15q26.3 | 3 (7.32%) | 0 (0.00%) |  | >10 | 0.241 | 1.00 | (A) Metastasis |
| **SMARCA4** | 19p13.2 | 3 (7.32%) | 0 (0.00%) |  | >10 | 0.241 | 1.00 | (A) Metastasis |
| **CSF3R** | 1p34.3 | 3 (7.32%) | 0 (0.00%) |  | >10 | 0.241 | 1.00 | (A) Metastasis |
| **AKT1** | 14q32.33 | 3 (7.14%) | 0 (0.00%) |  | >10 | 0.241 | 1.00 | (A) Metastasis |
| **APC** | 5q22.2 | 3 (7.14%) | 0 (0.00%) |  | >10 | 0.241 | 1.00 | (A) Metastasis |
| **ERBB2** | 17q12 | 3 (7.14%) | 0 (0.00%) |  | >10 | 0.241 | 1.00 | (A) Metastasis |
| **HNF1A** | 12q24.31 | 3 (7.14%) | 0 (0.00%) |  | >10 | 0.241 | 1.00 | (A) Metastasis |
| **JAK3** | 19p13.11 | 3 (7.14%) | 0 (0.00%) |  | >10 | 0.241 | 1.00 | (A) Metastasis |
| **CSDE1** | 1p13.2 | 3 (10.00%) | 0 (0.00%) |  | >10 | 0.249 | 1.00 | (A) Metastasis |
| **PRKD1** | 14q12 | 3 (10.00%) | 0 (0.00%) |  | >10 | 0.249 | 1.00 | (A) Metastasis |
| **EPHA5** | 4q13.1-q13.2 | 3 (8.82%) | 1 (2.94%) |  | Oca.58 | 0.614 | 1.00 | (A) Metastasis |
| **TBX3** | 12q24.21 | 3 (8.82%) | 1 (2.94%) |  | Oca.58 | 0.614 | 1.00 | (A) Metastasis |
| **MST1R** | 3p21.31 | 3 (8.82%) | 1 (3.03%) |  | Oca.54 | 0.614 | 1.00 | (A) Metastasis |
| **NCOA3** | 20q13.12 | 3 (8.82%) | 1 (3.03%) |  | Oca.54 | 0.614 | 1.00 | (A) Metastasis |
| **ATR** | 3q23 | 3 (7.32%) | 1 (2.44%) |  | Oca.58 | 0.616 | 1.00 | (A) Metastasis |
| **BRD4** | 19p13.12 | 3 (7.32%) | 1 (2.44%) |  | Oca.58 | 0.616 | 1.00 | (A) Metastasis |
| **CCND2** | 12p13.32 | 3 (7.32%) | 1 (2.44%) |  | Oca.58 | 0.616 | 1.00 | (A) Metastasis |
| **FANCA** | 16q24.3 | 3 (7.32%) | 1 (2.44%) |  | Oca.58 | 0.616 | 1.00 | (A) Metastasis |
| **KMT2A** | 11q23.3 | 3 (7.32%) | 1 (2.44%) |  | Oca.58 | 0.616 | 1.00 | (A) Metastasis |
| **PMS1** | 2q32.2 | 3 (7.32%) | 1 (2.44%) |  | Oca.58 | 0.616 | 1.00 | (A) Metastasis |
| **ROS1** | 6q22.1 | 3 (7.32%) | 1 (2.44%) |  | Oca.58 | 0.616 | 1.00 | (A) Metastasis |
| **SOX2** | 3q26.33 | 3 (7.32%) | 1 (2.44%) |  | Oca.58 | 0.616 | 1.00 | (A) Metastasis |
| **SUFU** | 10q24.32 | 3 (7.32%) | 1 (2.44%) |  | Oca.58 | 0.616 | 1.00 | (A) Metastasis |
| **MET** | 7q31.2 | 3 (7.14%) | 1 (2.38%) |  | Oca.58 | 0.616 | 1.00 | (A) Metastasis |
| **NSD3** | 8p11.23 | 3 (8.11%) | 1 (3.33%) |  | Oca.28 | 0.622 | 1.00 | (A) Metastasis |
| **SOS1** | 2p22.1 | 3 (8.11%) | 1 (3.33%) |  | Oca.28 | 0.622 | 1.00 | (A) Metastasis |
| **GLI1** | 12q13.3 | 3 (7.32%) | 4 (10.00%) |  | -0.45 | 0.712 | 1.00 | (B) Primary |
| **CBL** | 11q23.3 | 3 (7.32%) | 2 (4.88%) |  | 0.58 | 1.00 | 1.00 | (A) Metastasis |
| **KDM5A** | 12p13.33 | 3 (7.32%) | 2 (4.88%) |  | 0.58 | 1.00 | 1.00 | (A) Metastasis |
| **MDM2** | 12q15 | 3 (7.32%) | 4 (9.76%) |  | -0.42 | 1.00 | 1.00 | (B) Primary |
| **H3F3A** | 1q42.12 | 3 (7.32%) | 2 (5.00%) |  | 0.55 | 1.00 | 1.00 | (A) Metastasis |
| **BRAF** | 7q34 | 3 (7.14%) | 4 (9.52%) |  | -0.42 | 1.00 | 1.00 | (B) Primary |
| **ARID5B** | 10q21.2 | 3 (8.82%) | 2 (5.88%) |  | 0.58 | 1.00 | 1.00 | (A) Metastasis |
| **SPEN** | 1p36.21-p36.13 | 3 (8.82%) | 2 (5.88%) |  | 0.58 | 1.00 | 1.00 | (A) Metastasis |
| **TEK** | 9p21.2 | 2 (6.67%) | 5 (21.74%) |  | -1.71 | 0.218 | 1.00 | (B) Primary |
| **MTAP** | 9p21.3 | 2 (22.22%) | 5 (55.56%) |  | -1.32 | 0.335 | 1.00 | (B) Primary |
| **PAX5** | 9p13.2 | 2 (4.88%) | 5 (12.20%) |  | -1.32 | 0.432 | 1.00 | (B) Primary |
| **COP1** | 1q25.1-q25.2 | 2 (5.88%) | 0 (0.00%) |  | >10 | 0.493 | 1.00 | (A) Metastasis |
| **GRIN2A** | 16p13.2 | 2 (5.88%) | 0 (0.00%) |  | >10 | 0.493 | 1.00 | (A) Metastasis |
| **HGF** | 7q21.11 | 2 (5.88%) | 0 (0.00%) |  | >10 | 0.493 | 1.00 | (A) Metastasis |
| **JUN** | 1p32.1 | 2 (5.88%) | 0 (0.00%) |  | >10 | 0.493 | 1.00 | (A) Metastasis |
| **NOTCH4** | 6p21.32 | 2 (5.88%) | 0 (0.00%) |  | >10 | 0.493 | 1.00 | (A) Metastasis |
| **PIK3C3** | 18q12.3 | 2 (5.88%) | 0 (0.00%) |  | >10 | 0.493 | 1.00 | (A) Metastasis |
| **TGFBR2** | 3p24.1 | 2 (5.88%) | 0 (0.00%) |  | >10 | 0.493 | 1.00 | (A) Metastasis |
| **TP63** | 3q28 | 2 (5.88%) | 0 (0.00%) |  | >10 | 0.493 | 1.00 | (A) Metastasis |
| **ANKRD11** | 16q24.3 | 2 (5.88%) | 0 (0.00%) |  | >10 | 0.493 | 1.00 | (A) Metastasis |
| **PGR** | 11q22.1 | 2 (5.88%) | 0 (0.00%) |  | >10 | 0.493 | 1.00 | (A) Metastasis |

**Table S20. Gene-Level Mutation Frequencies in Primary Versus Metastatic Tumors Across the Entire Cohort (n = 1216 Genes)**

| **Gene** | **Cytoband** | **(A) Metastasis** | **(B) Primary** | **Log2 Ratio** | **p-Value** | **q-Value** | **Enriched in** |
| --- | --- | --- | --- | --- | --- | --- | --- |
| TP53 | 17p13.1 | 1504 (52.85%) | 1380 (48.49%) | 0.12 | 1.109e-3 | 0.0585 | (A) Metastasis |
| KRAS | 12p12.1 | 590 (20.73%) | 535 (18.80%) | 0.14 | 0.0722 | 0.731 | (A) Metastasis |
| CDKN2A | 9p21.3 | 491 (17.31%) | 391 (13.77%) | 0.33 | 2.461e-4 | 0.0305 | (A) Metastasis |
| PIK3CA | 3q26.32 | 381 (13.39%) | 389 (13.67%) | -0.03 | 0.786 | 1.00 | (B) Primary |
| APC | 5q22.2 | 361 (12.74%) | 319 (11.24%) | 0.18 | 0.0861 | 0.770 | (A) Metastasis |
| EGFR | 7p11.2 | 325 (11.42%) | 299 (10.51%) | 0.12 | 0.289 | 1.00 | (A) Metastasis |
| ARID1A | 1p36.11 | 308 (10.99%) | 298 (10.64%) | 0.05 | 0.699 | 1.00 | (A) Metastasis |
| CDKN2B | 9p21.3 | 305 (10.89%) | 246 (8.79%) | 0.31 | 9.223e-3 | 0.224 | (A) Metastasis |
| TERT | 5p15.33 | 300 (10.76%) | 290 (10.42%) | 0.05 | 0.695 | 1.00 | (A) Metastasis |
| PTEN | 10q23.31 | 291 (10.22%) | 247 (8.68%) | 0.24 | 0.0513 | 0.617 | (A) Metastasis |
| KMT2D | 12q13.12 | 281 (10.04%) | 246 (8.79%) | 0.19 | 0.120 | 0.859 | (A) Metastasis |
| MYC | 8q24.21 | 273 (9.74%) | 160 (5.71%) | 0.77 | 1.78e-8 | 2.166e-5 | (A) Metastasis |
| ERBB2 | 17q12 | 230 (8.08%) | 219 (7.70%) | 0.07 | 0.623 | 1.00 | (A) Metastasis |
| RB1 | 13q14.2 | 229 (8.07%) | 200 (7.05%) | 0.20 | 0.146 | 0.956 | (A) Metastasis |
| FAT1 | 4q35.2 | 228 (8.25%) | 172 (6.49%) | 0.35 | 0.0145 | 0.299 | (A) Metastasis |
| ATM | 11q22.3 | 226 (7.97%) | 167 (5.89%) | 0.44 | 2.028e-3 | 0.0849 | (A) Metastasis |
| NF1 | 17q11.2 | 209 (7.46%) | 162 (5.79%) | 0.37 | 0.0134 | 0.292 | (A) Metastasis |
| SMAD4 | 18q21.2 | 204 (7.19%) | 149 (5.25%) | 0.45 | 2.486e-3 | 0.0974 | (A) Metastasis |
| SMARCA4 | 19p13.2 | 182 (6.50%) | 133 (4.75%) | 0.45 | 5.299e-3 | 0.157 | (A) Metastasis |
| FGFR1 | 8p11.23 | 182 (6.39%) | 151 (5.31%) | 0.27 | 0.0901 | 0.770 | (A) Metastasis |
| BRCA2 | 13q13.1 | 172 (6.14%) | 143 (5.11%) | 0.27 | 0.104 | 0.828 | (A) Metastasis |
| CCND1 | 11q13.3 | 171 (6.11%) | 166 (5.93%) | 0.04 | 0.822 | 1.00 | (A) Metastasis |
| CDK12 | 17q12 | 167 (6.02%) | 151 (5.67%) | 0.09 | 0.603 | 1.00 | (A) Metastasis |
| KMT2C | 7q36.1 | 166 (7.30%) | 145 (6.39%) | 0.19 | 0.240 | 1.00 | (A) Metastasis |
| PTPRT | 20q12-q13.11 | 163 (7.20%) | 124 (5.50%) | 0.39 | 0.0204 | 0.375 | (A) Metastasis |
| AR | Xq12 | 159 (5.67%) | 88 (3.14%) | 0.85 | 4.560e-6 | 1.845e-3 | (A) Metastasis |
| BRAF | 7q34 | 157 (5.52%) | 149 (5.24%) | 0.08 | 0.681 | 1.00 | (A) Metastasis |
| CREBBP | 16p13.3 | 156 (5.57%) | 123 (4.39%) | 0.34 | 0.0492 | 0.617 | (A) Metastasis |
| FOXA1 | 14q21.1 | 156 (5.65%) | 126 (4.76%) | 0.25 | 0.143 | 0.951 | (A) Metastasis |
| NSD3 | 8p11.23 | 154 (6.16%) | 115 (5.16%) | 0.25 | 0.148 | 0.962 | (A) Metastasis |
| STK11 | 19p13.3 | 152 (5.36%) | 126 (4.44%) | 0.27 | 0.110 | 0.835 | (A) Metastasis |
| ATRX | Xq21.1 | 150 (5.35%) | 138 (4.93%) | 0.12 | 0.506 | 1.00 | (A) Metastasis |
| KEAP1 | 19p13.2 | 149 (5.32%) | 129 (4.66%) | 0.19 | 0.268 | 1.00 | (A) Metastasis |
| MDM2 | 12q15 | 146 (5.21%) | 135 (4.82%) | 0.11 | 0.541 | 1.00 | (A) Metastasis |
| MTAP | 9p21.3 | 145 (9.35%) | 95 (8.16%) | 0.20 | 0.306 | 1.00 | (A) Metastasis |
| KDM6A | Xp11.3 | 143 (5.11%) | 128 (4.57%) | 0.16 | 0.383 | 1.00 | (A) Metastasis |
| PREX2 | 8q13.2 | 142 (7.02%) | 98 (5.28%) | 0.41 | 0.0276 | 0.439 | (A) Metastasis |
| FGF3 | 11q13.3 | 142 (6.23%) | 130 (5.71%) | 0.12 | 0.492 | 1.00 | (A) Metastasis |
| ASXL1 | 20q11.21 | 139 (4.96%) | 80 (2.86%) | 0.80 | 5.824e-5 | 0.0141 | (A) Metastasis |
| RAD21 | 8q24.11 | 139 (5.06%) | 86 (3.21%) | 0.66 | 6.395e-4 | 0.0406 | (A) Metastasis |
| GATA3 | 10p14 | 139 (4.96%) | 143 (5.11%) | -0.04 | 0.855 | 1.00 | (B) Primary |
| NOTCH1 | 9q34.3 | 137 (4.81%) | 105 (3.69%) | 0.38 | 0.0415 | 0.542 | (A) Metastasis |
| MET | 7q31.2 | 137 (4.81%) | 111 (3.90%) | 0.30 | 0.104 | 0.828 | (A) Metastasis |
| PTPRD | 9p24.1-p23 | 135 (5.87%) | 114 (4.81%) | 0.29 | 0.118 | 0.856 | (A) Metastasis |
| ROS1 | 6q22.1 | 135 (4.80%) | 112 (3.99%) | 0.27 | 0.152 | 0.970 | (A) Metastasis |
| NOTCH3 | 19p13.12 | 134 (4.84%) | 102 (3.84%) | 0.33 | 0.0725 | 0.731 | (A) Metastasis |
| ZFHX3 | 16q22.2-q22.3 | 133 (6.01%) | 98 (4.56%) | 0.40 | 0.0359 | 0.490 | (A) Metastasis |
| FGF4 | 11q13.3 | 132 (5.80%) | 125 (5.49%) | 0.08 | 0.700 | 1.00 | (A) Metastasis |
| FGF19 | 11q13.3 | 130 (5.71%) | 125 (5.50%) | 0.06 | 0.797 | 1.00 | (A) Metastasis |
| RECQL4 | 8q24.3 | 128 (4.60%) | 66 (2.40%) | 0.94 | 9.985e-6 | 3.031e-3 | (A) Metastasis |
| ERBB4 | 2q34 | 128 (4.51%) | 102 (3.60%) | 0.33 | 0.0805 | 0.749 | (A) Metastasis |
| CDH1 | 16q22.1 | 128 (4.52%) | 115 (4.05%) | 0.16 | 0.395 | 1.00 | (A) Metastasis |
| KMT2A | 11q23.3 | 127 (4.54%) | 109 (3.90%) | 0.22 | 0.258 | 1.00 | (A) Metastasis |
| MGA | 15q15.1 | 125 (4.62%) | 99 (3.91%) | 0.24 | 0.219 | 1.00 | (A) Metastasis |
| SETD2 | 3p21.31 | 125 (4.46%) | 106 (3.79%) | 0.24 | 0.226 | 1.00 | (A) Metastasis |
| ALK | 2p23.2-p23.1 | 125 (4.39%) | 112 (3.94%) | 0.16 | 0.389 | 1.00 | (A) Metastasis |
| AGO2 | 8q24.3 | 124 (6.20%) | 64 (3.49%) | 0.83 | 9.393e-5 | 0.0182 | (A) Metastasis |
| ESR1 | 6q25.1-q25.2 | 123 (4.37%) | 61 (2.17%) | 1.01 | 3.982e-6 | 1.845e-3 | (A) Metastasis |
| RBM10 | Xp11.3 | 123 (4.45%) | 101 (3.80%) | 0.22 | 0.246 | 1.00 | (A) Metastasis |
| ARID1B | 6q25.3 | 122 (4.37%) | 89 (3.19%) | 0.45 | 0.0245 | 0.410 | (A) Metastasis |
| KDM5A | 12p13.33 | 121 (4.37%) | 66 (2.48%) | 0.82 | 1.374e-4 | 0.0208 | (A) Metastasis |
| ATR | 3q23 | 121 (4.36%) | 95 (3.57%) | 0.29 | 0.145 | 0.955 | (A) Metastasis |
| ARID2 | 12q12 | 119 (4.25%) | 103 (3.68%) | 0.21 | 0.304 | 1.00 | (A) Metastasis |
| TSC2 | 16p13.3 | 118 (4.21%) | 84 (3.00%) | 0.49 | 0.0178 | 0.333 | (A) Metastasis |
| POLE | 12q24.33 | 116 (4.19%) | 91 (3.42%) | 0.29 | 0.156 | 0.985 | (A) Metastasis |
| KMT2B | 19q13.12 | 116 (5.78%) | 103 (5.60%) | 0.05 | 0.835 | 1.00 | (A) Metastasis |
| GNAS | 20q13.32 | 115 (4.06%) | 78 (2.75%) | 0.56 | 6.711e-3 | 0.181 | (A) Metastasis |
| SDHA | 5p15.33 | 113 (4.04%) | 72 (2.60%) | 0.63 | 3.439e-3 | 0.116 | (A) Metastasis |
| CIC | 19q13.2 | 113 (4.07%) | 90 (3.38%) | 0.27 | 0.198 | 1.00 | (A) Metastasis |
| FLT1 | 13q12.3 | 112 (4.00%) | 85 (3.04%) | 0.40 | 0.0591 | 0.674 | (A) Metastasis |
| NKX2-1 | 14q13.3 | 112 (4.00%) | 89 (3.18%) | 0.33 | 0.114 | 0.837 | (A) Metastasis |
| CTNNB1 | 3p22.1 | 112 (3.94%) | 103 (3.62%) | 0.12 | 0.578 | 1.00 | (A) Metastasis |
| CARD11 | 7p22.2 | 111 (3.96%) | 78 (2.79%) | 0.51 | 0.0177 | 0.333 | (A) Metastasis |
| MED12 | Xq13.1 | 111 (4.00%) | 90 (3.38%) | 0.24 | 0.250 | 1.00 | (A) Metastasis |
| PBRM1 | 3p21.1 | 109 (3.89%) | 96 (3.47%) | 0.17 | 0.434 | 1.00 | (A) Metastasis |
| FBXW7 | 4q31.3 | 109 (3.84%) | 103 (3.63%) | 0.08 | 0.675 | 1.00 | (A) Metastasis |
| EP300 | 22q13.2 | 108 (3.86%) | 96 (3.43%) | 0.17 | 0.433 | 1.00 | (A) Metastasis |
| PIK3R1 | 5q13.1 | 108 (3.85%) | 109 (3.89%) | -0.01 | 0.945 | 1.00 | (B) Primary |
| NOTCH2 | 1p12 | 107 (3.82%) | 77 (2.75%) | 0.47 | 0.0295 | 0.439 | (A) Metastasis |
| RET | 10q11.21 | 107 (3.76%) | 99 (3.48%) | 0.11 | 0.619 | 1.00 | (A) Metastasis |
| MAP3K1 | 5q11.2 | 106 (3.79%) | 76 (2.72%) | 0.48 | 0.0286 | 0.439 | (A) Metastasis |
| GRIN2A | 16p13.2 | 106 (4.66%) | 95 (4.18%) | 0.16 | 0.471 | 1.00 | (A) Metastasis |
| FGFR3 | 4p16.3 | 106 (3.72%) | 96 (3.37%) | 0.14 | 0.519 | 1.00 | (A) Metastasis |
| NBN | 8q21.3 | 104 (3.71%) | 69 (2.47%) | 0.59 | 8.468e-3 | 0.214 | (A) Metastasis |
| KIT | 4q12 | 104 (3.65%) | 95 (3.34%) | 0.13 | 0.564 | 1.00 | (A) Metastasis |
| BCOR | Xp11.4 | 103 (3.68%) | 96 (3.43%) | 0.10 | 0.665 | 1.00 | (A) Metastasis |
| ERBB3 | 12q13.2 | 102 (3.64%) | 86 (3.07%) | 0.25 | 0.266 | 1.00 | (A) Metastasis |
| TMPRSS2 | 21q22.3 | 101 (3.64%) | 94 (3.53%) | 0.04 | 0.827 | 1.00 | (A) Metastasis |
| RNF43 | 17q22 | 100 (3.60%) | 112 (4.21%) | -0.22 | 0.263 | 1.00 | (B) Primary |
| TBX3 | 12q24.21 | 100 (4.40%) | 95 (4.19%) | 0.07 | 0.770 | 1.00 | (A) Metastasis |
| NSD1 | 5q35.3 | 99 (3.58%) | 84 (3.16%) | 0.18 | 0.409 | 1.00 | (A) Metastasis |
| BRCA1 | 17q21.31 | 98 (3.50%) | 68 (2.43%) | 0.53 | 0.0220 | 0.394 | (A) Metastasis |
| FANCA | 16q24.3 | 98 (3.50%) | 73 (2.61%) | 0.42 | 0.0620 | 0.695 | (A) Metastasis |
| CCNE1 | 19q12 | 97 (3.46%) | 74 (2.64%) | 0.39 | 0.0872 | 0.770 | (A) Metastasis |
| FLT4 | 5q35.3 | 97 (3.47%) | 90 (3.22%) | 0.11 | 0.656 | 1.00 | (A) Metastasis |
| TET2 | 4q24 | 96 (3.43%) | 77 (2.75%) | 0.32 | 0.164 | 1.00 | (A) Metastasis |
| PDGFRA | 4q12 | 96 (3.38%) | 77 (2.71%) | 0.32 | 0.164 | 1.00 | (A) Metastasis |
| ERG | 21q22.2 | 95 (3.42%) | 81 (3.04%) | 0.17 | 0.444 | 1.00 | (A) Metastasis |
| BAP1 | 3p21.1 | 94 (3.35%) | 66 (2.36%) | 0.51 | 0.0300 | 0.439 | (A) Metastasis |
| SLX4 | 16p13.3 | 92 (3.67%) | 68 (3.05%) | 0.27 | 0.260 | 1.00 | (A) Metastasis |
| SPEN | 1p36.21-p36.13 | 92 (4.04%) | 82 (3.60%) | 0.16 | 0.487 | 1.00 | (A) Metastasis |
| PIK3C2G | 12p12.3 | 91 (4.01%) | 58 (2.56%) | 0.65 | 5.968e-3 | 0.168 | (A) Metastasis |
| MCL1 | 1q21.2 | 91 (3.25%) | 72 (2.57%) | 0.34 | 0.152 | 0.970 | (A) Metastasis |
| MTOR | 1p36.22 | 91 (3.24%) | 78 (2.78%) | 0.22 | 0.349 | 1.00 | (A) Metastasis |
| TSC1 | 9q34.13 | 91 (3.25%) | 91 (3.25%) | -0.00 | 1.00 | 1.00 | (B) Primary |
| PRDM14 | 8q13.3 | 89 (4.45%) | 50 (2.72%) | 0.71 | 4.284e-3 | 0.141 | (A) Metastasis |
| DNMT3A | 2p23.3 | 89 (3.18%) | 58 (2.07%) | 0.62 | 9.598e-3 | 0.228 | (A) Metastasis |
| FLT3 | 13q12.2 | 89 (3.14%) | 74 (2.61%) | 0.27 | 0.235 | 1.00 | (A) Metastasis |
| KDR | 4q12 | 88 (3.10%) | 73 (2.57%) | 0.27 | 0.263 | 1.00 | (A) Metastasis |
| BRD4 | 19p13.12 | 87 (3.11%) | 67 (2.40%) | 0.38 | 0.120 | 0.859 | (A) Metastasis |
| NTRK1 | 1q23.1 | 85 (3.03%) | 68 (2.43%) | 0.32 | 0.190 | 1.00 | (A) Metastasis |
| RICTOR | 5p13.1 | 85 (3.06%) | 70 (2.63%) | 0.22 | 0.370 | 1.00 | (A) Metastasis |
| SF3B1 | 2q33.1 | 85 (3.03%) | 74 (2.64%) | 0.20 | 0.421 | 1.00 | (A) Metastasis |
| AKT1 | 14q32.33 | 85 (2.99%) | 76 (2.67%) | 0.16 | 0.523 | 1.00 | (A) Metastasis |
| FGFR2 | 10q26.13 | 84 (2.95%) | 71 (2.49%) | 0.24 | 0.328 | 1.00 | (A) Metastasis |
| RPTOR | 17q25.3 | 83 (2.99%) | 59 (2.22%) | 0.43 | 0.0746 | 0.734 | (A) Metastasis |
| NCOR1 | 17p12-p11.2 | 83 (3.66%) | 61 (2.69%) | 0.44 | 0.0750 | 0.734 | (A) Metastasis |
| KDM5C | Xp11.22 | 83 (2.97%) | 68 (2.46%) | 0.27 | 0.249 | 1.00 | (A) Metastasis |
| PTPRS | 19p13.3 | 83 (3.68%) | 71 (3.16%) | 0.22 | 0.367 | 1.00 | (A) Metastasis |
| EPHA5 | 4q13.1-q13.2 | 83 (3.61%) | 75 (3.12%) | 0.21 | 0.373 | 1.00 | (A) Metastasis |
| ELF3 | 1q32.1 | 83 (4.14%) | 81 (4.41%) | -0.09 | 0.690 | 1.00 | (B) Primary |
| IL7R | 5p13.2 | 81 (2.92%) | 53 (1.99%) | 0.55 | 0.0287 | 0.439 | (A) Metastasis |
| DDR2 | 1q23.3 | 81 (2.88%) | 59 (2.10%) | 0.45 | 0.0719 | 0.731 | (A) Metastasis |
| MAP2K4 | 17p12 | 81 (2.89%) | 60 (2.14%) | 0.43 | 0.0877 | 0.770 | (A) Metastasis |
| TET1 | 10q21.3 | 81 (2.94%) | 62 (2.35%) | 0.32 | 0.203 | 1.00 | (A) Metastasis |
| GLI1 | 12q13.3 | 81 (2.95%) | 79 (2.94%) | 0.00 | 1.00 | 1.00 | (A) Metastasis |
| NFKBIA | 14q13.2 | 80 (2.90%) | 60 (2.23%) | 0.38 | 0.123 | 0.865 | (A) Metastasis |
| EPHA3 | 3p11.1 | 80 (3.48%) | 65 (2.70%) | 0.37 | 0.129 | 0.885 | (A) Metastasis |
| IGF1R | 15q26.3 | 79 (2.82%) | 57 (2.04%) | 0.47 | 0.0679 | 0.731 | (A) Metastasis |
| ANKRD11 | 16q24.3 | 79 (3.58%) | 72 (3.35%) | 0.09 | 0.741 | 1.00 | (A) Metastasis |
| NTRK3 | 15q25.3 | 78 (2.78%) | 65 (2.32%) | 0.26 | 0.309 | 1.00 | (A) Metastasis |
| STAG2 | Xq25 | 78 (2.79%) | 81 (2.89%) | -0.06 | 0.810 | 1.00 | (B) Primary |
| JAK2 | 9p24.1 | 77 (2.72%) | 44 (1.55%) | 0.81 | 3.118e-3 | 0.115 | (A) Metastasis |
| PIK3CG | 7q22.3 | 77 (3.39%) | 62 (2.73%) | 0.31 | 0.228 | 1.00 | (A) Metastasis |
| AKT2 | 19q13.2 | 77 (2.75%) | 64 (2.29%) | 0.26 | 0.306 | 1.00 | (A) Metastasis |
| AXIN2 | 17q24.1 | 77 (2.80%) | 74 (2.80%) | -0.00 | 1.00 | 1.00 | (B) Primary |
| IRS2 | 13q34 | 76 (3.34%) | 56 (2.46%) | 0.44 | 0.0929 | 0.784 | (A) Metastasis |
| POLD1 | 19q13.33 | 76 (2.79%) | 57 (2.23%) | 0.32 | 0.219 | 1.00 | (A) Metastasis |
| DICER1 | 14q32.13 | 76 (2.72%) | 65 (2.33%) | 0.22 | 0.394 | 1.00 | (A) Metastasis |
| SOX9 | 17q24.3 | 76 (2.72%) | 68 (2.44%) | 0.16 | 0.555 | 1.00 | (A) Metastasis |
| SOX17 | 8q11.23 | 75 (3.33%) | 49 (2.18%) | 0.61 | 0.0225 | 0.395 | (A) Metastasis |
| NOTCH4 | 6p21.32 | 75 (3.30%) | 67 (2.95%) | 0.16 | 0.551 | 1.00 | (A) Metastasis |
| IKZF1 | 7p12.2 | 74 (2.64%) | 62 (2.21%) | 0.26 | 0.299 | 1.00 | (A) Metastasis |
| MSH6 | 2p16.3 | 73 (2.61%) | 53 (1.89%) | 0.46 | 0.0865 | 0.770 | (A) Metastasis |
| JAK1 | 1p31.3 | 73 (2.63%) | 52 (1.95%) | 0.43 | 0.103 | 0.828 | (A) Metastasis |
| BRIP1 | 17q23.2 | 73 (2.61%) | 73 (2.61%) | -0.00 | 1.00 | 1.00 | (B) Primary |
| CDK4 | 12q14.1 | 72 (2.57%) | 67 (2.39%) | 0.10 | 0.731 | 1.00 | (A) Metastasis |
| AURKA | 20q13.2 | 71 (2.54%) | 44 (1.57%) | 0.69 | 0.0140 | 0.292 | (A) Metastasis |
| PALB2 | 16p12.2 | 71 (2.53%) | 47 (1.68%) | 0.59 | 0.0319 | 0.453 | (A) Metastasis |
| DOT1L | 19p13.3 | 71 (3.12%) | 51 (2.24%) | 0.48 | 0.0808 | 0.749 | (A) Metastasis |
| PMS2 | 7p22.1 | 70 (2.50%) | 47 (1.68%) | 0.57 | 0.0394 | 0.526 | (A) Metastasis |
| JAK3 | 19p13.11 | 70 (2.47%) | 48 (1.69%) | 0.54 | 0.0503 | 0.617 | (A) Metastasis |
| EPHB1 | 3q22.2 | 70 (3.07%) | 61 (2.68%) | 0.20 | 0.478 | 1.00 | (A) Metastasis |
| PTCH1 | 9q22.32 | 70 (2.50%) | 63 (2.25%) | 0.15 | 0.599 | 1.00 | (A) Metastasis |
| RARA | 17q21.2 | 70 (2.50%) | 64 (2.29%) | 0.13 | 0.662 | 1.00 | (A) Metastasis |
| BCL2L1 | 20q11.21 | 69 (2.47%) | 38 (1.36%) | 0.86 | 3.226e-3 | 0.115 | (A) Metastasis |
| PRKN | 6q26 | 69 (2.47%) | 45 (1.61%) | 0.62 | 0.0291 | 0.439 | (A) Metastasis |
| WT1 | 11p13 | 69 (2.46%) | 61 (2.18%) | 0.18 | 0.535 | 1.00 | (A) Metastasis |
| AMER1 | Xq11.2 | 68 (2.99%) | 48 (2.11%) | 0.50 | 0.0735 | 0.731 | (A) Metastasis |
| RTEL1 | 20q13.33 | 68 (3.40%) | 50 (2.72%) | 0.32 | 0.261 | 1.00 | (A) Metastasis |
| ELOC | 8q21.11 | 67 (2.47%) | 33 (1.30%) | 0.93 | 2.285e-3 | 0.0925 | (A) Metastasis |
| NKX3-1 | 8p21.2 | 66 (2.40%) | 29 (1.10%) | 1.13 | 2.667e-4 | 0.0305 | (A) Metastasis |
| DIS3 | 13q21.33 | 66 (2.36%) | 45 (1.61%) | 0.55 | 0.0547 | 0.639 | (A) Metastasis |
| ERCC4 | 16p13.12 | 66 (2.36%) | 48 (1.72%) | 0.46 | 0.107 | 0.828 | (A) Metastasis |
| TCF7L2 | 10q25.2-q25.3 | 66 (2.41%) | 55 (2.05%) | 0.23 | 0.408 | 1.00 | (A) Metastasis |
| INPPL1 | 11q13.4 | 66 (3.30%) | 57 (3.10%) | 0.09 | 0.783 | 1.00 | (A) Metastasis |
| TP53BP1 | 15q15.3 | 65 (2.59%) | 47 (2.10%) | 0.30 | 0.293 | 1.00 | (A) Metastasis |
| ARAF | Xp11.3 | 64 (2.28%) | 43 (1.53%) | 0.57 | 0.0504 | 0.617 | (A) Metastasis |
| NCOA3 | 20q13.12 | 64 (2.90%) | 45 (2.10%) | 0.47 | 0.0991 | 0.802 | (A) Metastasis |
| RAD52 | 12p13.33 | 63 (2.28%) | 37 (1.39%) | 0.71 | 0.0155 | 0.313 | (A) Metastasis |
| PAK5 | 20p12.2 | 63 (2.78%) | 40 (1.77%) | 0.66 | 0.0220 | 0.394 | (A) Metastasis |
| ERCC5 | 13q33.1 | 63 (2.26%) | 50 (1.80%) | 0.33 | 0.254 | 1.00 | (A) Metastasis |
| PPM1D | 17q23.2 | 63 (2.33%) | 54 (2.13%) | 0.13 | 0.641 | 1.00 | (A) Metastasis |
| NRAS | 1p13.2 | 63 (2.21%) | 62 (2.18%) | 0.02 | 1.00 | 1.00 | (A) Metastasis |
| FGFR4 | 5q35.2 | 62 (2.21%) | 44 (1.57%) | 0.49 | 0.0951 | 0.796 | (A) Metastasis |
| ETV1 | 7p21.2 | 61 (2.18%) | 42 (1.50%) | 0.54 | 0.0729 | 0.731 | (A) Metastasis |
| DROSHA | 5p13.3 | 61 (3.05%) | 39 (2.12%) | 0.52 | 0.0843 | 0.770 | (A) Metastasis |
| ETV6 | 12p13.2 | 61 (2.18%) | 45 (1.61%) | 0.44 | 0.141 | 0.945 | (A) Metastasis |
| RUNX1 | 21q22.12 | 61 (2.18%) | 60 (2.15%) | 0.02 | 1.00 | 1.00 | (A) Metastasis |
| CDK6 | 7q21.2 | 60 (2.14%) | 30 (1.07%) | 1.00 | 1.896e-3 | 0.0822 | (A) Metastasis |
| TGFBR2 | 3p24.1 | 60 (2.64%) | 33 (1.45%) | 0.86 | 4.648e-3 | 0.148 | (A) Metastasis |
| CEBPA | 19q13.11 | 60 (2.18%) | 40 (1.48%) | 0.55 | 0.0687 | 0.731 | (A) Metastasis |
| NSD2 | 4p16.3 | 60 (2.39%) | 42 (1.88%) | 0.35 | 0.231 | 1.00 | (A) Metastasis |
| AXL | 19q13.2 | 60 (2.14%) | 47 (1.68%) | 0.35 | 0.241 | 1.00 | (A) Metastasis |
| PDGFRB | 5q32 | 60 (2.14%) | 57 (2.04%) | 0.07 | 0.852 | 1.00 | (A) Metastasis |
| LATS1 | 6q25.1 | 59 (2.61%) | 37 (1.64%) | 0.67 | 0.0298 | 0.439 | (A) Metastasis |
| MSH2 | 2p21-p16.3 | 59 (2.11%) | 41 (1.46%) | 0.52 | 0.0858 | 0.770 | (A) Metastasis |
| PRKDC | 8q11.21 | 58 (10.64%) | 38 (6.96%) | 0.61 | 0.0329 | 0.460 | (A) Metastasis |
| CCND2 | 12p13.32 | 58 (2.07%) | 38 (1.36%) | 0.61 | 0.0499 | 0.617 | (A) Metastasis |
| PLCG2 | 16q23.3 | 58 (2.60%) | 42 (1.94%) | 0.43 | 0.157 | 0.985 | (A) Metastasis |
| PGR | 11q22.1 | 58 (2.63%) | 45 (2.10%) | 0.33 | 0.273 | 1.00 | (A) Metastasis |
| TP63 | 3q28 | 58 (2.57%) | 54 (2.40%) | 0.10 | 0.774 | 1.00 | (A) Metastasis |
| NFE2L2 | 2q31.2 | 58 (2.07%) | 56 (2.00%) | 0.05 | 0.925 | 1.00 | (A) Metastasis |
| MDC1 | 6p21.33 | 57 (2.53%) | 32 (1.42%) | 0.83 | 9.803e-3 | 0.228 | (A) Metastasis |
| SMAD2 | 18q21.1 | 57 (2.04%) | 41 (1.47%) | 0.47 | 0.126 | 0.879 | (A) Metastasis |
| PAK1 | 11q13.5-q14.1 | 57 (2.53%) | 47 (2.09%) | 0.27 | 0.372 | 1.00 | (A) Metastasis |
| SPOP | 17q21.33 | 57 (2.05%) | 60 (2.25%) | -0.13 | 0.641 | 1.00 | (B) Primary |
| COL7A1 | 3p21.31 | 57 (11.47%) | 41 (10.57%) | 0.12 | 0.746 | 1.00 | (A) Metastasis |
| TCF3 | 19p13.3 | 56 (2.04%) | 33 (1.23%) | 0.73 | 0.0243 | 0.410 | (A) Metastasis |
| LYN | 8q12.1 | 56 (2.77%) | 35 (1.89%) | 0.56 | 0.0717 | 0.731 | (A) Metastasis |
| SOS1 | 2p22.1 | 56 (2.24%) | 38 (1.71%) | 0.39 | 0.211 | 1.00 | (A) Metastasis |
| CTCF | 16q22.1 | 56 (2.02%) | 50 (1.88%) | 0.11 | 0.769 | 1.00 | (A) Metastasis |
| ARID5B | 10q21.2 | 55 (2.44%) | 30 (1.33%) | 0.87 | 8.219e-3 | 0.212 | (A) Metastasis |
| TEK | 9p21.2 | 55 (2.73%) | 38 (2.05%) | 0.41 | 0.174 | 1.00 | (A) Metastasis |
| PPARG | 3p25.2 | 55 (2.18%) | 37 (1.65%) | 0.40 | 0.206 | 1.00 | (A) Metastasis |
| IDH1 | 2q34 | 55 (1.93%) | 46 (1.62%) | 0.26 | 0.422 | 1.00 | (A) Metastasis |
| NF2 | 22q12.2 | 55 (1.96%) | 49 (1.75%) | 0.16 | 0.621 | 1.00 | (A) Metastasis |
| EPHA7 | 6q16.1 | 55 (2.44%) | 53 (2.30%) | 0.08 | 0.771 | 1.00 | (A) Metastasis |
| PRDM1 | 6q21 | 54 (1.93%) | 41 (1.47%) | 0.40 | 0.214 | 1.00 | (A) Metastasis |
| CDKN1B | 12p13.1 | 54 (1.93%) | 45 (1.61%) | 0.26 | 0.417 | 1.00 | (A) Metastasis |
| ERCC2 | 19q13.32 | 54 (1.93%) | 56 (2.00%) | -0.05 | 0.848 | 1.00 | (B) Primary |
| LATS2 | 13q12.11 | 53 (2.35%) | 32 (1.42%) | 0.72 | 0.0280 | 0.439 | (A) Metastasis |
| RIT1 | 1q22 | 53 (1.92%) | 33 (1.25%) | 0.62 | 0.0506 | 0.617 | (A) Metastasis |
| TOP1 | 20q12 | 53 (2.33%) | 37 (1.63%) | 0.52 | 0.110 | 0.835 | (A) Metastasis |
| VHL | 3p25.3 | 53 (1.87%) | 49 (1.73%) | 0.11 | 0.691 | 1.00 | (A) Metastasis |
| HGF | 7q21.11 | 53 (2.33%) | 51 (2.24%) | 0.05 | 0.921 | 1.00 | (A) Metastasis |
| DUSP4 | 8p12 | 52 (2.60%) | 24 (1.31%) | 0.99 | 5.063e-3 | 0.157 | (A) Metastasis |
| FOXL2 | 3q22.3 | 52 (1.87%) | 27 (1.01%) | 0.89 | 8.864e-3 | 0.220 | (A) Metastasis |
| MDM4 | 1q32.1 | 52 (1.86%) | 39 (1.39%) | 0.41 | 0.204 | 1.00 | (A) Metastasis |
| EZH2 | 7q36.1 | 52 (1.84%) | 41 (1.45%) | 0.34 | 0.296 | 1.00 | (A) Metastasis |
| DNMT3B | 20q11.21 | 52 (2.31%) | 42 (1.87%) | 0.31 | 0.348 | 1.00 | (A) Metastasis |
| RASA1 | 5q14.3 | 52 (1.89%) | 41 (1.55%) | 0.28 | 0.349 | 1.00 | (A) Metastasis |
| CDKN1A | 6p21.2 | 52 (1.86%) | 46 (1.65%) | 0.18 | 0.611 | 1.00 | (A) Metastasis |
| DAXX | 6p21.32 | 51 (1.84%) | 36 (1.35%) | 0.44 | 0.161 | 1.00 | (A) Metastasis |
| NTRK2 | 9q21.33 | 51 (1.82%) | 44 (1.57%) | 0.21 | 0.535 | 1.00 | (A) Metastasis |
| ARHGAP35 | 19q13.32 | 51 (3.30%) | 40 (3.45%) | -0.06 | 0.830 | 1.00 | (B) Primary |
| CRLF2 | Xp22.33 and Yp11.2 | 50 (1.79%) | 22 (0.79%) | 1.18 | 1.204e-3 | 0.0609 | (A) Metastasis |
| TSHR | 14q31.1 | 50 (1.80%) | 29 (1.09%) | 0.73 | 0.0310 | 0.448 | (A) Metastasis |
| CYLD | 16q12.1 | 50 (1.96%) | 29 (1.22%) | 0.69 | 0.0410 | 0.541 | (A) Metastasis |
| CCND3 | 6p21.1 | 50 (1.78%) | 34 (1.22%) | 0.55 | 0.0986 | 0.802 | (A) Metastasis |
| PPP2R1A | 19q13.41 | 50 (1.80%) | 62 (2.33%) | -0.37 | 0.182 | 1.00 | (B) Primary |
| RAD50 | 5q31.1 | 50 (1.80%) | 38 (1.43%) | 0.34 | 0.284 | 1.00 | (A) Metastasis |
| PIK3CB | 3q22.3 | 50 (2.19%) | 46 (2.02%) | 0.12 | 0.757 | 1.00 | (A) Metastasis |
| MEN1 | 11q13.1 | 50 (1.79%) | 46 (1.64%) | 0.12 | 0.758 | 1.00 | (A) Metastasis |
| YES1 | 18p11.32 | 49 (2.17%) | 26 (1.16%) | 0.91 | 9.953e-3 | 0.228 | (A) Metastasis |
| ASXL2 | 2p23.3 | 49 (2.17%) | 32 (1.42%) | 0.61 | 0.0722 | 0.731 | (A) Metastasis |
| SRC | 20q11.23 | 49 (2.09%) | 34 (1.39%) | 0.59 | 0.0758 | 0.736 | (A) Metastasis |
| PRKD1 | 14q12 | 49 (2.45%) | 48 (2.61%) | -0.09 | 0.758 | 1.00 | (B) Primary |
| HNF1A | 12q24.31 | 48 (1.69%) | 33 (1.16%) | 0.54 | 0.117 | 0.853 | (A) Metastasis |
| SMO | 7q32.1 | 47 (1.66%) | 34 (1.20%) | 0.47 | 0.179 | 1.00 | (A) Metastasis |
| BLM | 15q26.1 | 47 (1.68%) | 46 (1.65%) | 0.03 | 1.00 | 1.00 | (A) Metastasis |
| HIST2H3C | 1q21.2 | 46 (2.08%) | 24 (1.12%) | 0.90 | 0.0114 | 0.257 | (A) Metastasis |
| CSF3R | 1p34.3 | 46 (1.68%) | 30 (1.13%) | 0.57 | 0.105 | 0.828 | (A) Metastasis |
| CDK8 | 13q12.13 | 46 (1.66%) | 30 (1.13%) | 0.56 | 0.106 | 0.828 | (A) Metastasis |
| MPL | 1p34.2 | 46 (1.62%) | 32 (1.13%) | 0.52 | 0.112 | 0.835 | (A) Metastasis |
| CASP8 | 2q33.1 | 46 (1.66%) | 37 (1.39%) | 0.26 | 0.440 | 1.00 | (A) Metastasis |
| POLQ | 3q13.33 | 46 (9.18%) | 40 (10.26%) | -0.16 | 0.648 | 1.00 | (B) Primary |
| CBL | 11q23.3 | 46 (1.64%) | 41 (1.47%) | 0.16 | 0.666 | 1.00 | (A) Metastasis |
| PRKCI | 3q26.2 | 46 (1.81%) | 42 (1.77%) | 0.04 | 0.915 | 1.00 | (A) Metastasis |
| FLCN | 17p11.2 | 45 (1.61%) | 32 (1.14%) | 0.49 | 0.168 | 1.00 | (A) Metastasis |
| RAD51C | 17q22 | 45 (1.62%) | 38 (1.43%) | 0.18 | 0.582 | 1.00 | (A) Metastasis |
| RAF1 | 3p25.2 | 45 (1.61%) | 39 (1.39%) | 0.21 | 0.583 | 1.00 | (A) Metastasis |
| FOXO1 | 13q14.11 | 44 (1.98%) | 21 (0.97%) | 1.03 | 5.881e-3 | 0.168 | (A) Metastasis |
| PIK3C3 | 18q12.3 | 44 (1.94%) | 25 (1.10%) | 0.81 | 0.0283 | 0.439 | (A) Metastasis |
| PHOX2B | 4p13 | 44 (1.58%) | 30 (1.08%) | 0.55 | 0.128 | 0.880 | (A) Metastasis |
| AKT3 | 1q43-q44 | 44 (1.57%) | 30 (1.07%) | 0.55 | 0.128 | 0.880 | (A) Metastasis |
| PMS1 | 2q32.2 | 44 (1.58%) | 32 (1.15%) | 0.46 | 0.204 | 1.00 | (A) Metastasis |
| DNMT1 | 19p13.2 | 44 (1.95%) | 47 (2.09%) | -0.10 | 0.752 | 1.00 | (B) Primary |
| YAP1 | 11q22.1 | 44 (1.60%) | 40 (1.52%) | 0.08 | 0.827 | 1.00 | (A) Metastasis |
| TRAF7 | 16p13.3 | 43 (1.56%) | 32 (1.21%) | 0.36 | 0.296 | 1.00 | (A) Metastasis |
| ERF | 19q13.2 | 43 (2.15%) | 33 (1.80%) | 0.26 | 0.487 | 1.00 | (A) Metastasis |
| BCL6 | 3q27.3 | 43 (1.54%) | 37 (1.32%) | 0.22 | 0.574 | 1.00 | (A) Metastasis |
| GATA1 | Xp11.23 | 42 (1.85%) | 26 (1.14%) | 0.69 | 0.0661 | 0.723 | (A) Metastasis |
| VEGFA | 6p21.1 | 42 (1.54%) | 28 (1.10%) | 0.49 | 0.185 | 1.00 | (A) Metastasis |
| STAT3 | 17q21.2 | 42 (1.52%) | 32 (1.19%) | 0.36 | 0.294 | 1.00 | (A) Metastasis |
| PARP1 | 1q42.12 | 42 (1.85%) | 36 (1.58%) | 0.22 | 0.568 | 1.00 | (A) Metastasis |
| ABL1 | 9q34.12 | 42 (1.48%) | 47 (1.66%) | -0.16 | 0.669 | 1.00 | (B) Primary |
| BCL2 | 18q21.33 | 41 (1.47%) | 13 (0.46%) | 1.66 | 1.051e-4 | 0.0182 | (A) Metastasis |
| XPO1 | 2p15 | 41 (1.46%) | 32 (1.14%) | 0.36 | 0.346 | 1.00 | (A) Metastasis |
| U2AF1 | 21q22.3 | 41 (1.46%) | 35 (1.25%) | 0.23 | 0.564 | 1.00 | (A) Metastasis |
| SMARCB1 | 22q11.23\|22q11 | 41 (1.45%) | 38 (1.34%) | 0.11 | 0.736 | 1.00 | (A) Metastasis |
| PAX5 | 9p13.2 | 41 (1.46%) | 41 (1.47%) | -0.00 | 1.00 | 1.00 | (B) Primary |
| RAC1 | 7p22.1 | 40 (1.44%) | 23 (0.86%) | 0.74 | 0.0566 | 0.655 | (A) Metastasis |
| MAPK1 | 22q11.22 | 40 (1.43%) | 28 (1.01%) | 0.51 | 0.179 | 1.00 | (A) Metastasis |
| KAT6A | 8p11.21 | 40 (7.68%) | 22 (5.35%) | 0.52 | 0.186 | 1.00 | (A) Metastasis |
| RPS6KB2 | 11q13.2 | 40 (1.78%) | 30 (1.33%) | 0.41 | 0.278 | 1.00 | (A) Metastasis |
| INSR | 19p13.2 | 40 (1.77%) | 31 (1.37%) | 0.37 | 0.339 | 1.00 | (A) Metastasis |
| CHEK2 | 22q12.1 | 40 (1.43%) | 32 (1.14%) | 0.32 | 0.407 | 1.00 | (A) Metastasis |
| TNFAIP3 | 6q23.3 | 40 (1.43%) | 33 (1.18%) | 0.28 | 0.480 | 1.00 | (A) Metastasis |
| PRKAR1A | 17q24.2 | 40 (1.43%) | 34 (1.22%) | 0.23 | 0.559 | 1.00 | (A) Metastasis |
| SETDB1 | 1q21.3 | 40 (3.82%) | 32 (4.16%) | -0.12 | 0.717 | 1.00 | (B) Primary |
| IRS1 | 2q36.3 | 40 (1.78%) | 37 (1.65%) | 0.11 | 0.818 | 1.00 | (A) Metastasis |
| TRIP13 | 5p15.33 | 39 (3.73%) | 11 (1.43%) | 1.38 | 3.327e-3 | 0.115 | (A) Metastasis |
| ZRSR2 | Xp22.2 | 39 (1.42%) | 24 (0.90%) | 0.67 | 0.0765 | 0.738 | (A) Metastasis |
| CD274 | 9p24.1 | 39 (1.40%) | 27 (0.97%) | 0.53 | 0.173 | 1.00 | (A) Metastasis |
| MECOM | 3q26.2 | 39 (7.34%) | 28 (5.30%) | 0.47 | 0.207 | 1.00 | (A) Metastasis |
| CRKL | 22q11.21 | 39 (1.39%) | 28 (1.00%) | 0.48 | 0.219 | 1.00 | (A) Metastasis |
| ZNRF3 | 22q12.1 | 39 (2.50%) | 24 (2.05%) | 0.29 | 0.444 | 1.00 | (A) Metastasis |
| MYCL | 1p34.2 | 39 (1.39%) | 33 (1.18%) | 0.24 | 0.553 | 1.00 | (A) Metastasis |
| BARD1 | 2q35 | 39 (1.41%) | 34 (1.28%) | 0.14 | 0.724 | 1.00 | (A) Metastasis |
| CD79B | 17q23.3 | 39 (1.39%) | 37 (1.32%) | 0.07 | 0.908 | 1.00 | (A) Metastasis |
| IDH2 | 15q26.1 | 38 (1.34%) | 22 (0.78%) | 0.79 | 0.0507 | 0.617 | (A) Metastasis |
| CDH4 | 20q13.33 | 38 (7.65%) | 18 (4.64%) | 0.72 | 0.0718 | 0.731 | (A) Metastasis |
| SOX2 | 3q26.33 | 38 (1.36%) | 30 (1.07%) | 0.34 | 0.393 | 1.00 | (A) Metastasis |
| PTPN11 | 12q24.13 | 38 (1.34%) | 31 (1.09%) | 0.30 | 0.400 | 1.00 | (A) Metastasis |
| E2F3 | 6p22.3 | 38 (1.69%) | 42 (1.87%) | -0.15 | 0.654 | 1.00 | (B) Primary |
| HOXB13 | 17q21.32 | 38 (1.39%) | 34 (1.33%) | 0.07 | 0.906 | 1.00 | (A) Metastasis |
| CYSLTR2 | 13q14.2 | 37 (1.85%) | 18 (0.98%) | 0.92 | 0.0289 | 0.439 | (A) Metastasis |
| PIK3CD | 1p36.22 | 37 (1.63%) | 20 (0.89%) | 0.88 | 0.0321 | 0.453 | (A) Metastasis |
| AXIN1 | 16p13.3 | 37 (1.63%) | 22 (0.97%) | 0.75 | 0.0657 | 0.723 | (A) Metastasis |
| GNA11 | 19p13.3 | 37 (1.30%) | 24 (0.84%) | 0.62 | 0.122 | 0.859 | (A) Metastasis |
| SMAD3 | 15q22.33 | 37 (1.63%) | 26 (1.15%) | 0.51 | 0.204 | 1.00 | (A) Metastasis |
| MAP3K13 | 3q27.2 | 37 (1.63%) | 32 (1.41%) | 0.21 | 0.547 | 1.00 | (A) Metastasis |
| MRE11 | 11q21 | 37 (1.33%) | 41 (1.54%) | -0.21 | 0.569 | 1.00 | (B) Primary |
| BTK | Xq22.1 | 37 (1.62%) | 33 (1.45%) | 0.16 | 0.718 | 1.00 | (A) Metastasis |
| MSH3 | 5q14.1 | 37 (1.84%) | 36 (1.95%) | -0.08 | 0.814 | 1.00 | (B) Primary |
| FANCM | 14q21.2 | 36 (7.03%) | 18 (4.44%) | 0.66 | 0.120 | 0.859 | (A) Metastasis |
| KLF4 | 9q31.2 | 36 (1.31%) | 25 (0.95%) | 0.46 | 0.247 | 1.00 | (A) Metastasis |
| FOXP1 | 3p13 | 36 (1.59%) | 26 (1.15%) | 0.47 | 0.250 | 1.00 | (A) Metastasis |
| CDC73 | 1q31.2 | 36 (1.29%) | 26 (0.93%) | 0.47 | 0.250 | 1.00 | (A) Metastasis |
| ZNF217 | 20q13.2 | 36 (6.57%) | 30 (5.45%) | 0.27 | 0.449 | 1.00 | (A) Metastasis |
| INPP4B | 4q31.21 | 36 (1.58%) | 31 (1.36%) | 0.21 | 0.623 | 1.00 | (A) Metastasis |
| SOCS1 | 16p13.13 | 36 (1.29%) | 37 (1.32%) | -0.04 | 0.907 | 1.00 | (B) Primary |
| IKBKE | 1q32.1 | 35 (1.54%) | 21 (0.92%) | 0.74 | 0.0796 | 0.749 | (A) Metastasis |
| SUFU | 10q24.32 | 35 (1.25%) | 27 (0.96%) | 0.37 | 0.371 | 1.00 | (A) Metastasis |
| MST1R | 3p21.31 | 35 (1.58%) | 27 (1.25%) | 0.34 | 0.373 | 1.00 | (A) Metastasis |
| MLH1 | 3p22.2 | 35 (1.23%) | 31 (1.09%) | 0.18 | 0.623 | 1.00 | (A) Metastasis |
| MAP2K2 | 19p13.3 | 35 (1.26%) | 30 (1.13%) | 0.16 | 0.709 | 1.00 | (A) Metastasis |
| SDHC | 1q23.3 | 35 (1.25%) | 32 (1.14%) | 0.13 | 0.806 | 1.00 | (A) Metastasis |
| RUNX1T1 | 8q21.3 | 34 (6.55%) | 10 (2.43%) | 1.43 | 3.009e-3 | 0.114 | (A) Metastasis |
| RAD54B | 8q22.1 | 34 (6.84%) | 13 (3.35%) | 1.03 | 0.0233 | 0.404 | (A) Metastasis |
| PIK3R2 | 19p13.11 | 34 (1.50%) | 19 (0.84%) | 0.84 | 0.0521 | 0.617 | (A) Metastasis |
| MUTYH | 1p34.1 | 34 (1.21%) | 19 (0.68%) | 0.84 | 0.0523 | 0.617 | (A) Metastasis |
| EIF1AX | Xp22.12 | 34 (1.51%) | 23 (1.02%) | 0.56 | 0.182 | 1.00 | (A) Metastasis |
| MYCN | 2p24.3 | 34 (1.21%) | 23 (0.82%) | 0.56 | 0.183 | 1.00 | (A) Metastasis |
| SH2B3 | 12q24.12 | 34 (1.23%) | 42 (1.56%) | -0.34 | 0.356 | 1.00 | (B) Primary |
| SETBP1 | 18q12.3 | 34 (6.34%) | 27 (5.08%) | 0.32 | 0.429 | 1.00 | (A) Metastasis |
| UPF1 | 19p13.11 | 34 (1.70%) | 25 (1.36%) | 0.32 | 0.432 | 1.00 | (A) Metastasis |
| CALR | 19p13.13 | 34 (1.25%) | 26 (1.02%) | 0.29 | 0.516 | 1.00 | (A) Metastasis |
| INPP4A | 2q11.2 | 34 (1.51%) | 28 (1.24%) | 0.28 | 0.523 | 1.00 | (A) Metastasis |
| MITF | 3p13 | 34 (1.21%) | 29 (1.04%) | 0.23 | 0.613 | 1.00 | (A) Metastasis |
| CSF1R | 5q32 | 34 (1.45%) | 40 (1.66%) | -0.19 | 0.640 | 1.00 | (B) Primary |
| EZH1 | 17q21.2 | 33 (1.65%) | 10 (0.54%) | 1.60 | 1.100e-3 | 0.0585 | (A) Metastasis |
| REL | 2p16.1 | 33 (1.18%) | 19 (0.68%) | 0.80 | 0.0524 | 0.617 | (A) Metastasis |
| GAB2 | 11q14.1 | 33 (3.15%) | 16 (2.08%) | 0.60 | 0.188 | 1.00 | (A) Metastasis |
| TGFBR1 | 9q22.33 | 33 (1.46%) | 29 (1.29%) | 0.18 | 0.702 | 1.00 | (A) Metastasis |
| MALT1 | 18q21.32 | 32 (1.44%) | 13 (0.60%) | 1.26 | 6.582e-3 | 0.181 | (A) Metastasis |
| IRF4 | 6p25.3 | 32 (1.41%) | 19 (0.84%) | 0.75 | 0.0901 | 0.770 | (A) Metastasis |
| PIK3C2B | 1q32.1 | 32 (5.85%) | 22 (4.02%) | 0.54 | 0.209 | 1.00 | (A) Metastasis |
| INHBA | 7p14.1 | 32 (1.44%) | 23 (1.06%) | 0.44 | 0.280 | 1.00 | (A) Metastasis |
| BIRC3 | 11q22.2 | 32 (1.44%) | 28 (1.30%) | 0.15 | 0.699 | 1.00 | (A) Metastasis |
| GATA2 | 3q21.3 | 32 (1.15%) | 28 (1.05%) | 0.13 | 0.795 | 1.00 | (A) Metastasis |
| RECQL | 12p12.1 | 31 (1.55%) | 18 (0.98%) | 0.66 | 0.149 | 0.965 | (A) Metastasis |
| NEGR1 | 1p31.1 | 31 (1.39%) | 24 (1.07%) | 0.38 | 0.345 | 1.00 | (A) Metastasis |
| HRAS | 11p15.5 | 31 (1.09%) | 29 (1.02%) | 0.10 | 0.897 | 1.00 | (A) Metastasis |
| PLK2 | 5q11.2 | 30 (1.33%) | 17 (0.76%) | 0.82 | 0.0774 | 0.740 | (A) Metastasis |
| COP1 | 1q25.1-q25.2 | 30 (1.32%) | 19 (0.80%) | 0.72 | 0.0863 | 0.770 | (A) Metastasis |
| ACVR1 | 2q24.1 | 30 (1.11%) | 17 (0.67%) | 0.72 | 0.107 | 0.828 | (A) Metastasis |
| ERRFI1 | 1p36.23 | 30 (1.35%) | 20 (0.92%) | 0.55 | 0.202 | 1.00 | (A) Metastasis |
| H3F3C | 12p11.21 | 30 (1.33%) | 21 (0.93%) | 0.51 | 0.260 | 1.00 | (A) Metastasis |
| WRN | 8p12 | 30 (5.61%) | 23 (4.34%) | 0.37 | 0.398 | 1.00 | (A) Metastasis |
| BMPR1A | 10q23.2 | 30 (1.07%) | 23 (0.82%) | 0.38 | 0.408 | 1.00 | (A) Metastasis |
| CUL3 | 2q36.2 | 30 (1.32%) | 26 (1.14%) | 0.20 | 0.687 | 1.00 | (A) Metastasis |
| RPS6KA4 | 11q13.1 | 30 (1.33%) | 27 (1.20%) | 0.15 | 0.790 | 1.00 | (A) Metastasis |
| STAT5B | 17q21.2 | 29 (1.31%) | 16 (0.74%) | 0.82 | 0.0724 | 0.731 | (A) Metastasis |
| SMARCA2 | 9p24.3 | 29 (2.77%) | 15 (1.95%) | 0.51 | 0.283 | 1.00 | (A) Metastasis |
| KLF5 | 13q22.1 | 29 (2.77%) | 17 (2.21%) | 0.33 | 0.546 | 1.00 | (A) Metastasis |
| HIST1H1C | 6p22.2 | 29 (1.29%) | 25 (1.11%) | 0.21 | 0.682 | 1.00 | (A) Metastasis |
| RIF1 | 2q23.3 | 29 (5.84%) | 25 (6.44%) | -0.14 | 0.778 | 1.00 | (B) Primary |
| EWSR1 | 22q12.2 | 29 (5.27%) | 26 (4.73%) | 0.16 | 0.782 | 1.00 | (A) Metastasis |
| NUP93 | 16q13 | 29 (1.30%) | 30 (1.38%) | -0.09 | 0.896 | 1.00 | (B) Primary |
| ERCC3 | 2q14.3 | 29 (1.04%) | 30 (1.08%) | -0.05 | 0.897 | 1.00 | (B) Primary |
| JUN | 1p32.1 | 28 (1.23%) | 17 (0.75%) | 0.72 | 0.133 | 0.904 | (A) Metastasis |
| PDCD1LG2 | 9p24.1 | 28 (1.10%) | 17 (0.71%) | 0.62 | 0.178 | 1.00 | (A) Metastasis |
| EED | 11q14.2 | 28 (1.23%) | 22 (0.93%) | 0.40 | 0.394 | 1.00 | (A) Metastasis |
| TENT5C | 1p12 | 28 (1.00%) | 22 (0.79%) | 0.35 | 0.478 | 1.00 | (A) Metastasis |
| DOCK8 | 9p24.3 | 28 (5.63%) | 19 (4.90%) | 0.20 | 0.654 | 1.00 | (A) Metastasis |
| MAP2K1 | 15q22.31 | 28 (1.00%) | 31 (1.10%) | -0.15 | 0.697 | 1.00 | (B) Primary |
| GPS2 | 17p13.1 | 27 (1.22%) | 15 (0.70%) | 0.81 | 0.0883 | 0.770 | (A) Metastasis |
| USP8 | 15q21.2 | 27 (1.75%) | 11 (0.95%) | 0.88 | 0.0983 | 0.802 | (A) Metastasis |
| FH | 1q43 | 27 (0.96%) | 17 (0.61%) | 0.67 | 0.172 | 1.00 | (A) Metastasis |
| FANCD2 | 3p25.3 | 27 (4.92%) | 21 (3.85%) | 0.35 | 0.461 | 1.00 | (A) Metastasis |
| FUBP1 | 1p31.1 | 27 (1.19%) | 21 (0.93%) | 0.36 | 0.469 | 1.00 | (A) Metastasis |
| ERCC6 | 10q11.23 | 27 (5.39%) | 17 (4.36%) | 0.31 | 0.535 | 1.00 | (A) Metastasis |
| ALOX12B | 17p13.1 | 27 (1.18%) | 24 (1.00%) | 0.24 | 0.576 | 1.00 | (A) Metastasis |
| NRG1 | 8p12 | 27 (5.38%) | 18 (4.60%) | 0.22 | 0.646 | 1.00 | (A) Metastasis |
| PDCD1 | 2q37.3 | 27 (1.19%) | 26 (1.15%) | 0.05 | 1.00 | 1.00 | (A) Metastasis |
| EXT1 | 8q24.11 | 26 (4.90%) | 9 (1.70%) | 1.52 | 5.216e-3 | 0.157 | (A) Metastasis |
| RSPO2 | 8q23.1 | 26 (5.10%) | 8 (1.98%) | 1.36 | 0.0136 | 0.292 | (A) Metastasis |
| IFNGR1 | 6q23.3 | 26 (1.15%) | 17 (0.76%) | 0.61 | 0.220 | 1.00 | (A) Metastasis |
| HLA-A | 6p22.1 | 26 (1.17%) | 18 (0.83%) | 0.49 | 0.291 | 1.00 | (A) Metastasis |
| HIST1H3B | 6p22.2 | 26 (0.94%) | 20 (0.76%) | 0.32 | 0.464 | 1.00 | (A) Metastasis |
| IGF2 | 11p15.5 | 26 (0.94%) | 21 (0.79%) | 0.25 | 0.562 | 1.00 | (A) Metastasis |
| LZTR1 | 22q11.21\|22q11.1-q11.2 | 26 (2.45%) | 16 (2.04%) | 0.27 | 0.637 | 1.00 | (A) Metastasis |
| SRSF2 | 17q25.1 | 26 (0.95%) | 29 (1.08%) | -0.19 | 0.685 | 1.00 | (B) Primary |
| CSDE1 | 1p13.2 | 26 (1.30%) | 21 (1.14%) | 0.18 | 0.769 | 1.00 | (A) Metastasis |
| SUZ12 | 17q11.2 | 26 (0.93%) | 26 (0.94%) | -0.00 | 1.00 | 1.00 | (B) Primary |
| GLI2 | 2q14.2 | 26 (4.93%) | 25 (4.76%) | 0.05 | 1.00 | 1.00 | (A) Metastasis |
| EPAS1 | 2p21 | 26 (1.30%) | 23 (1.25%) | 0.05 | 1.00 | 1.00 | (A) Metastasis |
| H3F3A | 1q42.12 | 25 (0.91%) | 17 (0.63%) | 0.52 | 0.279 | 1.00 | (A) Metastasis |
| SHQ1 | 3p13 | 25 (1.11%) | 19 (0.84%) | 0.39 | 0.449 | 1.00 | (A) Metastasis |
| POT1 | 7q31.33 | 25 (1.61%) | 15 (1.29%) | 0.32 | 0.524 | 1.00 | (A) Metastasis |
| FANCC | 9q22.32 | 25 (0.89%) | 24 (0.86%) | 0.06 | 1.00 | 1.00 | (A) Metastasis |
| PIM1 | 6p21.2 | 24 (0.86%) | 13 (0.47%) | 0.88 | 0.0978 | 0.802 | (A) Metastasis |
| MAFB | 20q12 | 24 (4.73%) | 10 (2.53%) | 0.91 | 0.112 | 0.835 | (A) Metastasis |
| H3F3B | 17q25.1 | 24 (0.89%) | 13 (0.51%) | 0.79 | 0.137 | 0.921 | (A) Metastasis |
| NTHL1 | 16p13.3 | 24 (0.96%) | 13 (0.58%) | 0.72 | 0.185 | 1.00 | (A) Metastasis |
| ICOSLG | 21q22.3 | 24 (1.07%) | 17 (0.76%) | 0.49 | 0.347 | 1.00 | (A) Metastasis |
| PIK3R3 | 1p34.1 | 24 (1.07%) | 18 (0.80%) | 0.41 | 0.439 | 1.00 | (A) Metastasis |
| KAT6B | 10q22.2 | 24 (4.77%) | 24 (6.11%) | -0.36 | 0.455 | 1.00 | (B) Primary |
| B2M | 15q21.1 | 24 (0.86%) | 29 (1.04%) | -0.28 | 0.494 | 1.00 | (B) Primary |
| FYN | 6q21 | 24 (1.09%) | 19 (0.88%) | 0.30 | 0.542 | 1.00 | (A) Metastasis |
| SESN1 | 6q21 | 24 (1.20%) | 24 (1.31%) | -0.12 | 0.773 | 1.00 | (B) Primary |
| BCORL1 | Xq26.1 | 24 (4.36%) | 23 (4.18%) | 0.06 | 1.00 | 1.00 | (A) Metastasis |
| SMYD3 | 1q44 | 23 (1.15%) | 13 (0.71%) | 0.70 | 0.181 | 1.00 | (A) Metastasis |
| MTA1 | 14q32.33 | 23 (4.63%) | 11 (2.84%) | 0.71 | 0.217 | 1.00 | (A) Metastasis |
| WWTR1 | 3q25.1 | 23 (1.15%) | 16 (0.87%) | 0.40 | 0.424 | 1.00 | (A) Metastasis |
| CARM1 | 19p13.2 | 23 (1.15%) | 16 (0.87%) | 0.40 | 0.424 | 1.00 | (A) Metastasis |
| STAT5A | 17q21.2 | 23 (1.04%) | 17 (0.79%) | 0.40 | 0.429 | 1.00 | (A) Metastasis |
| NPM1 | 5q35.1 | 23 (0.81%) | 18 (0.63%) | 0.35 | 0.439 | 1.00 | (A) Metastasis |
| SMARCE1 | 17q21.2 | 23 (1.48%) | 21 (1.81%) | -0.29 | 0.541 | 1.00 | (B) Primary |
| CUX1 | 7q22.1 | 23 (4.36%) | 20 (3.80%) | 0.20 | 0.756 | 1.00 | (A) Metastasis |
| CXCR4 | 2q22.1 | 23 (0.85%) | 19 (0.75%) | 0.18 | 0.757 | 1.00 | (A) Metastasis |
| PMAIP1 | 18q21.32 | 22 (0.98%) | 7 (0.31%) | 1.65 | 7.933e-3 | 0.209 | (A) Metastasis |
| GSK3B | 3q13.33 | 22 (0.97%) | 8 (0.35%) | 1.46 | 0.0158 | 0.314 | (A) Metastasis |
| GEN1 | 2p24.2 | 22 (4.30%) | 11 (2.73%) | 0.65 | 0.218 | 1.00 | (A) Metastasis |
| AURKB | 17p13.1 | 22 (0.79%) | 16 (0.57%) | 0.46 | 0.416 | 1.00 | (A) Metastasis |
| BABAM1 | 19p13.11 | 22 (0.88%) | 17 (0.76%) | 0.20 | 0.748 | 1.00 | (A) Metastasis |
| CHEK1 | 11q24.2 | 22 (0.79%) | 19 (0.71%) | 0.15 | 0.756 | 1.00 | (A) Metastasis |
| MAX | 14q23.3 | 22 (0.80%) | 19 (0.72%) | 0.15 | 0.757 | 1.00 | (A) Metastasis |
| USP28 | 11q23.2 | 22 (4.43%) | 16 (4.12%) | 0.10 | 0.869 | 1.00 | (A) Metastasis |
| RRAGC | 1p34.3 | 21 (1.05%) | 12 (0.65%) | 0.68 | 0.221 | 1.00 | (A) Metastasis |
| STK19 | 6p21.33 | 21 (1.05%) | 14 (0.76%) | 0.46 | 0.398 | 1.00 | (A) Metastasis |
| EIF4A2 | 3q27.3 | 21 (0.95%) | 26 (1.21%) | -0.35 | 0.464 | 1.00 | (B) Primary |
| SPRED1 | 15q14 | 21 (1.05%) | 15 (0.82%) | 0.36 | 0.505 | 1.00 | (A) Metastasis |
| RYBP | 3p13 | 21 (0.93%) | 17 (0.76%) | 0.30 | 0.626 | 1.00 | (A) Metastasis |
| STAT6 | 12q13.3 | 21 (3.97%) | 18 (3.42%) | 0.21 | 0.745 | 1.00 | (A) Metastasis |
| MEF2B | 19p13.11 | 21 (0.75%) | 18 (0.64%) | 0.22 | 0.748 | 1.00 | (A) Metastasis |
| RXRA | 9q34.2 | 21 (1.05%) | 22 (1.20%) | -0.19 | 0.759 | 1.00 | (B) Primary |
| MAPK3 | 16p11.2 | 21 (0.95%) | 23 (1.07%) | -0.17 | 0.763 | 1.00 | (B) Primary |
| PTPN14 | 1q32.3-q41 | 21 (4.23%) | 18 (4.64%) | -0.13 | 0.869 | 1.00 | (B) Primary |
| MYBL1 | 8q13.1 | 20 (3.82%) | 7 (1.34%) | 1.51 | 0.0177 | 0.333 | (A) Metastasis |
| ETV5 | 3q27.2 | 20 (3.64%) | 11 (2.00%) | 0.86 | 0.144 | 0.954 | (A) Metastasis |
| TNFRSF14 | 1p36.32 | 20 (0.88%) | 11 (0.48%) | 0.86 | 0.148 | 0.962 | (A) Metastasis |
| HLA-B | 6p21.33 | 20 (0.99%) | 11 (0.59%) | 0.74 | 0.207 | 1.00 | (A) Metastasis |
| MYOD1 | 11p15.1 | 20 (0.89%) | 15 (0.67%) | 0.41 | 0.498 | 1.00 | (A) Metastasis |
| RAD51D | 17q12 | 20 (0.72%) | 16 (0.60%) | 0.26 | 0.619 | 1.00 | (A) Metastasis |
| CBLB | 3q13.11 | 20 (3.78%) | 17 (3.24%) | 0.22 | 0.738 | 1.00 | (A) Metastasis |
| NUF2 | 1q23.3 | 20 (1.00%) | 21 (1.14%) | -0.19 | 0.754 | 1.00 | (B) Primary |
| ABCB11 | 2q31.1 | 20 (4.02%) | 14 (3.61%) | 0.16 | 0.861 | 1.00 | (A) Metastasis |
| RHOA | 3p21.31 | 20 (0.73%) | 18 (0.68%) | 0.09 | 0.872 | 1.00 | (A) Metastasis |
| TAP2 | 6p21.32 | 20 (1.00%) | 20 (1.09%) | -0.13 | 0.874 | 1.00 | (B) Primary |
| DNAJB1 | 19p13.12 | 20 (0.91%) | 21 (0.98%) | -0.11 | 0.876 | 1.00 | (B) Primary |
| RAD54L | 1p34.1 | 20 (0.88%) | 20 (0.88%) | -0.00 | 1.00 | 1.00 | (B) Primary |
| PNKP | 19q13.33 | 20 (4.02%) | 15 (3.87%) | 0.06 | 1.00 | 1.00 | (A) Metastasis |
| MSI1 | 12q24.31 | 20 (1.00%) | 18 (0.98%) | 0.03 | 1.00 | 1.00 | (A) Metastasis |
| HIST2H3D | 1q21.2 | 19 (1.64%) | 13 (0.94%) | 0.79 | 0.153 | 0.970 | (A) Metastasis |
| RAD51 | 15q15.1 | 19 (0.68%) | 11 (0.41%) | 0.73 | 0.202 | 1.00 | (A) Metastasis |
| BCL11B | 14q32.2 | 19 (3.73%) | 9 (2.27%) | 0.72 | 0.248 | 1.00 | (A) Metastasis |
| ITK | 5q33.3 | 19 (3.81%) | 10 (2.58%) | 0.56 | 0.346 | 1.00 | (A) Metastasis |
| EXO1 | 1q43 | 19 (3.82%) | 12 (3.09%) | 0.31 | 0.586 | 1.00 | (A) Metastasis |
| CBFB | 16q22.1 | 19 (0.69%) | 22 (0.83%) | -0.27 | 0.639 | 1.00 | (B) Primary |
| SYK | 9q22.2 | 19 (0.83%) | 23 (0.95%) | -0.21 | 0.646 | 1.00 | (B) Primary |
| RBBP8 | 18q11.2 | 19 (3.82%) | 14 (3.61%) | 0.08 | 1.00 | 1.00 | (A) Metastasis |
| POLB | 8p11.21 | 18 (3.62%) | 5 (1.29%) | 1.49 | 0.0334 | 0.461 | (A) Metastasis |
| CD276 | 15q24.1 | 18 (0.80%) | 9 (0.40%) | 1.00 | 0.121 | 0.859 | (A) Metastasis |
| BRD3 | 9q34.2 | 18 (3.56%) | 8 (2.04%) | 0.81 | 0.229 | 1.00 | (A) Metastasis |
| RHBDF2 | 17q25.1 | 18 (3.62%) | 9 (2.32%) | 0.64 | 0.326 | 1.00 | (A) Metastasis |
| HIST3H3 | 1q42.13 | 18 (0.82%) | 12 (0.56%) | 0.54 | 0.361 | 1.00 | (A) Metastasis |
| GATA6 | 18q11.2 | 18 (3.33%) | 13 (2.40%) | 0.47 | 0.370 | 1.00 | (A) Metastasis |
| POLH | 6p21.1 | 18 (3.59%) | 11 (2.82%) | 0.35 | 0.572 | 1.00 | (A) Metastasis |
| ARHGEF12 | 11q23.3 | 18 (3.62%) | 12 (3.09%) | 0.23 | 0.712 | 1.00 | (A) Metastasis |
| SLFN11 | 17q12 | 18 (1.72%) | 14 (1.82%) | -0.08 | 0.859 | 1.00 | (B) Primary |
| CD79A | 19q13.2 | 18 (0.81%) | 19 (0.87%) | -0.12 | 0.870 | 1.00 | (B) Primary |
| KIF1B | 1p36.22 | 18 (3.59%) | 14 (3.59%) | 0.00 | 1.00 | 1.00 | (A) Metastasis |
| MSI2 | 17q22 | 18 (0.90%) | 17 (0.93%) | -0.04 | 1.00 | 1.00 | (B) Primary |
| MCM8 | 20p12.3 | 17 (3.42%) | 4 (1.03%) | 1.73 | 0.0246 | 0.410 | (A) Metastasis |
| RAC2 | 22q13.1 | 17 (0.85%) | 6 (0.33%) | 1.38 | 0.0379 | 0.512 | (A) Metastasis |
| HIST1H2BD | 6p22.2 | 17 (0.75%) | 11 (0.49%) | 0.63 | 0.343 | 1.00 | (A) Metastasis |
| XRCC2 | 7q36.1 | 17 (0.62%) | 11 (0.43%) | 0.54 | 0.351 | 1.00 | (A) Metastasis |
| CCNQ | Xq28 | 17 (0.85%) | 11 (0.60%) | 0.50 | 0.449 | 1.00 | (A) Metastasis |
| PTK2B | 8p21.2 | 17 (3.42%) | 16 (4.12%) | -0.27 | 0.596 | 1.00 | (B) Primary |
| FAAP100 | 17q25.3 | 17 (3.42%) | 11 (2.84%) | 0.27 | 0.701 | 1.00 | (A) Metastasis |
| TRAF2 | 9q34.3 | 17 (0.77%) | 15 (0.70%) | 0.14 | 0.860 | 1.00 | (A) Metastasis |
| TMEM127 | 2q11.2 | 17 (0.62%) | 16 (0.61%) | 0.03 | 1.00 | 1.00 | (A) Metastasis |
| TRIM37 | 17q22 | 17 (3.42%) | 14 (3.61%) | -0.08 | 1.00 | 1.00 | (B) Primary |
| LIG4 | 13q33.3 | 17 (3.42%) | 13 (3.35%) | 0.03 | 1.00 | 1.00 | (A) Metastasis |
| DCUN1D1 | 3q26.33 | 17 (0.75%) | 17 (0.76%) | -0.00 | 1.00 | 1.00 | (B) Primary |
| PRF1 | 10q22.1 | 16 (3.05%) | 7 (1.34%) | 1.19 | 0.0896 | 0.770 | (A) Metastasis |
| MYD88 | 3p22.2 | 16 (0.57%) | 7 (0.25%) | 1.19 | 0.0925 | 0.784 | (A) Metastasis |
| MLH3 | 14q24.3 | 16 (3.16%) | 16 (4.05%) | -0.36 | 0.475 | 1.00 | (B) Primary |
| PNRC1 | 6q15 | 16 (0.70%) | 13 (0.54%) | 0.37 | 0.578 | 1.00 | (A) Metastasis |
| XIAP | Xq25 | 16 (0.71%) | 19 (0.84%) | -0.25 | 0.616 | 1.00 | (B) Primary |
| BUB1B | 15q15.1 | 16 (2.99%) | 13 (2.45%) | 0.29 | 0.707 | 1.00 | (A) Metastasis |
| TAP1 | 6p21.32 | 16 (0.80%) | 17 (0.92%) | -0.21 | 0.728 | 1.00 | (B) Primary |
| EPCAM | 2p21 | 16 (0.58%) | 16 (0.61%) | -0.06 | 1.00 | 1.00 | (B) Primary |
| ALB | 4q13.3 | 16 (1.52%) | 11 (1.42%) | 0.10 | 1.00 | 1.00 | (A) Metastasis |
| CIITA | 16p13.13 | 16 (3.02%) | 16 (3.04%) | -0.01 | 1.00 | 1.00 | (B) Primary |
| TAL1 | 1p33 | 15 (2.98%) | 4 (1.02%) | 1.55 | 0.0594 | 0.674 | (A) Metastasis |
| ENG | 9q34.11 | 15 (2.99%) | 5 (1.28%) | 1.22 | 0.111 | 0.835 | (A) Metastasis |
| TOPBP1 | 3q22.1 | 15 (3.01%) | 8 (2.06%) | 0.54 | 0.405 | 1.00 | (A) Metastasis |
| FOXF1 | 16q24.1 | 15 (1.43%) | 8 (1.04%) | 0.46 | 0.529 | 1.00 | (A) Metastasis |
| EGFL7 | 9q34.3 | 15 (0.67%) | 11 (0.49%) | 0.44 | 0.556 | 1.00 | (A) Metastasis |
| CDKN2C | 1p32.3 | 15 (0.54%) | 12 (0.43%) | 0.32 | 0.700 | 1.00 | (A) Metastasis |
| SS18 | 18q11.2 | 15 (3.01%) | 14 (3.61%) | -0.26 | 0.704 | 1.00 | (B) Primary |
| GREM1 | 15q13.3 | 15 (0.54%) | 16 (0.60%) | -0.15 | 0.858 | 1.00 | (B) Primary |
| MAPKAP1 | 9q33.3 | 15 (0.75%) | 13 (0.71%) | 0.08 | 1.00 | 1.00 | (A) Metastasis |
| PPP4R2 | 3p13 | 15 (0.75%) | 13 (0.71%) | 0.08 | 1.00 | 1.00 | (A) Metastasis |
| SERPINB4 | 18q21.33 | 14 (1.34%) | 4 (0.52%) | 1.37 | 0.0959 | 0.797 | (A) Metastasis |
| PAXIP1 | 7q36.2 | 14 (2.82%) | 17 (4.38%) | -0.64 | 0.269 | 1.00 | (B) Primary |
| PRSS1 | 7q34 | 14 (2.74%) | 16 (3.98%) | -0.54 | 0.351 | 1.00 | (B) Primary |
| HELQ | 4q21.23 | 14 (2.82%) | 7 (1.80%) | 0.64 | 0.379 | 1.00 | (A) Metastasis |
| CTLA4 | 2q33.2 | 14 (0.51%) | 18 (0.68%) | -0.42 | 0.479 | 1.00 | (B) Primary |
| FANCI | 15q26.1 | 14 (2.72%) | 8 (1.97%) | 0.46 | 0.520 | 1.00 | (A) Metastasis |
| SESN3 | 11q21 | 14 (0.70%) | 9 (0.49%) | 0.51 | 0.531 | 1.00 | (A) Metastasis |
| IL10 | 1q32.1 | 14 (0.62%) | 10 (0.44%) | 0.48 | 0.540 | 1.00 | (A) Metastasis |
| VTCN1 | 1p13.1-p12 | 14 (0.62%) | 10 (0.44%) | 0.48 | 0.540 | 1.00 | (A) Metastasis |
| ABRAXAS1 | 4q21.23 | 14 (0.51%) | 10 (0.38%) | 0.42 | 0.543 | 1.00 | (A) Metastasis |
| CTNNA1 | 5q31.2 | 14 (2.68%) | 13 (3.13%) | -0.22 | 0.699 | 1.00 | (B) Primary |
| RRAS2 | 11p15.2 | 14 (0.70%) | 15 (0.82%) | -0.22 | 0.712 | 1.00 | (B) Primary |
| NECTIN4 | 1q23.3 | 14 (2.82%) | 12 (3.09%) | -0.13 | 0.843 | 1.00 | (B) Primary |
| INHA | 2q35 | 14 (0.63%) | 12 (0.56%) | 0.18 | 0.845 | 1.00 | (A) Metastasis |
| TFE3 | Xp11.23 | 14 (2.75%) | 11 (2.78%) | -0.02 | 1.00 | 1.00 | (B) Primary |
| REST | 4q12 | 14 (1.34%) | 11 (1.43%) | -0.09 | 1.00 | 1.00 | (B) Primary |
| KNSTRN | 15q15.1 | 14 (0.70%) | 13 (0.71%) | -0.02 | 1.00 | 1.00 | (B) Primary |
| DDB1 | 11q12.2 | 13 (2.62%) | 6 (1.55%) | 0.76 | 0.353 | 1.00 | (A) Metastasis |
| PML | 15q24.1 | 13 (2.55%) | 6 (1.52%) | 0.75 | 0.353 | 1.00 | (A) Metastasis |
| LMO1 | 11p15.4 | 13 (0.47%) | 8 (0.29%) | 0.70 | 0.382 | 1.00 | (A) Metastasis |
| SDHD | 11q23.1 | 13 (0.46%) | 8 (0.29%) | 0.70 | 0.382 | 1.00 | (A) Metastasis |
| GBA | 1q22 | 13 (2.62%) | 7 (1.80%) | 0.54 | 0.499 | 1.00 | (A) Metastasis |
| XRCC1 | 19q13.31 | 13 (2.59%) | 7 (1.79%) | 0.53 | 0.499 | 1.00 | (A) Metastasis |
| ID3 | 1p36.12 | 13 (0.47%) | 9 (0.34%) | 0.50 | 0.523 | 1.00 | (A) Metastasis |
| GATA4 | 8p23.1 | 13 (2.40%) | 10 (1.85%) | 0.38 | 0.537 | 1.00 | (A) Metastasis |
| FAN1 | 15q13.3 | 13 (2.62%) | 13 (3.35%) | -0.36 | 0.552 | 1.00 | (B) Primary |
| UBE2T | 1q32.1 | 13 (2.59%) | 8 (2.05%) | 0.34 | 0.661 | 1.00 | (A) Metastasis |
| SMC3 | 10q25.2 | 13 (2.43%) | 10 (1.88%) | 0.37 | 0.674 | 1.00 | (A) Metastasis |
| UIMC1 | 5q35.2 | 13 (2.61%) | 11 (2.84%) | -0.12 | 0.838 | 1.00 | (B) Primary |
| RRAS | 19q13.33 | 13 (0.65%) | 13 (0.71%) | -0.12 | 0.846 | 1.00 | (B) Primary |
| BBC3 | 19q13.32 | 13 (0.58%) | 14 (0.62%) | -0.11 | 0.850 | 1.00 | (B) Primary |
| RAD51B | 14q24.1 | 13 (0.57%) | 14 (0.62%) | -0.11 | 0.850 | 1.00 | (B) Primary |
| GNAQ | 9q21.2 | 13 (0.46%) | 13 (0.46%) | - | 1.00 | 1.00 | (B) Primary |
| IGF1 | 12q23.2 | 13 (0.57%) | 13 (0.57%) | - | 1.00 | 1.00 | (B) Primary |
| FUS | 16p11.2 | 13 (2.43%) | 12 (2.41%) | 0.01 | 1.00 | 1.00 | (A) Metastasis |
| HIST1H3F | 6p22.2 | 13 (0.59%) | 13 (0.61%) | -0.04 | 1.00 | 1.00 | (B) Primary |
| SESN2 | 1p35.3 | 12 (0.60%) | 2 (0.11%) | 2.46 | 0.0139 | 0.292 | (A) Metastasis |
| PGBD5 | 1q42.13 | 12 (1.15%) | 3 (0.39%) | 1.56 | 0.114 | 0.837 | (A) Metastasis |
| RSPO3 | 6q22.33 | 12 (2.41%) | 5 (1.29%) | 0.91 | 0.324 | 1.00 | (A) Metastasis |
| SLC25A13 | 7q21.3 | 12 (2.41%) | 5 (1.29%) | 0.91 | 0.324 | 1.00 | (A) Metastasis |
| TLX3 | 5q35.1 | 12 (2.41%) | 5 (1.29%) | 0.91 | 0.324 | 1.00 | (A) Metastasis |
| QKI | 6q26 | 12 (2.22%) | 7 (1.37%) | 0.70 | 0.359 | 1.00 | (A) Metastasis |
| WAS | Xp11.23 | 12 (2.38%) | 6 (1.53%) | 0.64 | 0.474 | 1.00 | (A) Metastasis |
| SHOC2 | 10q25.2 | 12 (0.60%) | 8 (0.44%) | 0.46 | 0.510 | 1.00 | (A) Metastasis |
| ATXN7 | 3p14.1 | 12 (1.15%) | 12 (1.56%) | -0.44 | 0.534 | 1.00 | (B) Primary |
| RINT1 | 7q22.3 | 12 (2.40%) | 7 (1.79%) | 0.42 | 0.643 | 1.00 | (A) Metastasis |
| EME1 | 17q21.33 | 12 (2.41%) | 11 (2.84%) | -0.23 | 0.832 | 1.00 | (B) Primary |
| BCL2L11 | 2q13 | 12 (0.53%) | 13 (0.58%) | -0.12 | 0.844 | 1.00 | (B) Primary |
| SDHB | 1p36.13 | 12 (0.43%) | 11 (0.39%) | 0.12 | 1.00 | 1.00 | (A) Metastasis |
| SH2D1A | Xq25 | 12 (0.44%) | 11 (0.42%) | 0.06 | 1.00 | 1.00 | (A) Metastasis |
| XPC | 3p25.1 | 12 (2.24%) | 11 (2.08%) | 0.11 | 1.00 | 1.00 | (A) Metastasis |
| DIS3L2 | 2q37.1 | 12 (2.40%) | 9 (2.31%) | 0.05 | 1.00 | 1.00 | (A) Metastasis |
| MUS81 | 11q13.1 | 12 (2.41%) | 9 (2.32%) | 0.06 | 1.00 | 1.00 | (A) Metastasis |
| SERPINB3 | 18q21.33 | 11 (1.05%) | 1 (0.13%) | 3.02 | 0.0175 | 0.333 | (A) Metastasis |
| MYB | 6q23.3 | 11 (2.01%) | 6 (1.10%) | 0.87 | 0.328 | 1.00 | (A) Metastasis |
| JAZF1 | 7p15.2-p15.1 | 11 (2.21%) | 5 (1.29%) | 0.78 | 0.447 | 1.00 | (A) Metastasis |
| ETAA1 | 2p14 | 11 (1.05%) | 5 (0.65%) | 0.70 | 0.451 | 1.00 | (A) Metastasis |
| STK40 | 1p34.3 | 11 (0.49%) | 7 (0.31%) | 0.65 | 0.480 | 1.00 | (A) Metastasis |
| HIST1H3J | 6p22.1 | 11 (0.50%) | 14 (0.65%) | -0.39 | 0.551 | 1.00 | (B) Primary |
| CBFA2T3 | 16q24.3 | 11 (2.21%) | 6 (1.55%) | 0.52 | 0.623 | 1.00 | (A) Metastasis |
| RELA | 11q13.1 | 11 (2.21%) | 6 (1.55%) | 0.52 | 0.623 | 1.00 | (A) Metastasis |
| CTR9 | 11p15.4 | 11 (1.05%) | 10 (1.30%) | -0.30 | 0.661 | 1.00 | (B) Primary |
| FANCL | 2p16.1 | 11 (2.11%) | 7 (1.69%) | 0.32 | 0.812 | 1.00 | (A) Metastasis |
| HABP2 | 10q25.3 | 11 (2.21%) | 7 (1.80%) | 0.29 | 0.812 | 1.00 | (A) Metastasis |
| PPP6C | 9q33.3 | 11 (0.50%) | 9 (0.42%) | 0.25 | 0.824 | 1.00 | (A) Metastasis |
| ETV4 | 17q21.31 | 11 (2.00%) | 11 (2.00%) | - | 1.00 | 1.00 | (B) Primary |
| DKC1 | Xq28 | 11 (2.20%) | 9 (2.30%) | -0.07 | 1.00 | 1.00 | (B) Primary |
| SBDS | 7q11.21 | 10 (1.88%) | 4 (0.76%) | 1.32 | 0.177 | 1.00 | (A) Metastasis |
| PTP4A1 | 6q12 | 10 (0.50%) | 4 (0.22%) | 1.20 | 0.184 | 1.00 | (A) Metastasis |
| BRCC3 | Xq28 | 10 (2.00%) | 4 (1.03%) | 0.96 | 0.289 | 1.00 | (A) Metastasis |
| FANCF | 11p14.3 | 10 (1.83%) | 5 (0.92%) | 1.00 | 0.298 | 1.00 | (A) Metastasis |
| EIF4E | 4q23 | 10 (0.45%) | 5 (0.23%) | 0.96 | 0.302 | 1.00 | (A) Metastasis |
| APLNR | 11q12.1 | 10 (0.95%) | 4 (0.52%) | 0.88 | 0.417 | 1.00 | (A) Metastasis |
| XRCC5 | 2q35 | 10 (2.01%) | 5 (1.29%) | 0.64 | 0.446 | 1.00 | (A) Metastasis |
| TACC3 | 4p16.3 | 10 (0.35%) | 6 (0.21%) | 0.74 | 0.454 | 1.00 | (A) Metastasis |
| FANCG | 9p13.3 | 10 (1.82%) | 7 (1.27%) | 0.51 | 0.626 | 1.00 | (A) Metastasis |
| RAB35 | 12q24.23 | 10 (0.45%) | 12 (0.56%) | -0.30 | 0.673 | 1.00 | (B) Primary |
| KCNQ1 | 11p15.5-p15.4 | 10 (2.01%) | 6 (1.55%) | 0.38 | 0.800 | 1.00 | (A) Metastasis |
| NEIL2 | 8p23.1 | 10 (2.01%) | 6 (1.55%) | 0.38 | 0.800 | 1.00 | (A) Metastasis |
| XRCC4 | 5q14.2 | 10 (2.01%) | 6 (1.55%) | 0.38 | 0.800 | 1.00 | (A) Metastasis |
| NFKBIZ | 3q12.3 | 10 (1.91%) | 8 (1.53%) | 0.32 | 0.813 | 1.00 | (A) Metastasis |
| RHEB | 7q36.1 | 10 (0.37%) | 8 (0.30%) | 0.27 | 0.815 | 1.00 | (A) Metastasis |
| HIST1H3E | 6p22.2 | 10 (0.45%) | 11 (0.51%) | -0.18 | 0.829 | 1.00 | (B) Primary |
| PHF6 | Xq26.2 | 10 (0.63%) | 9 (0.69%) | -0.13 | 1.00 | 1.00 | (B) Primary |
| SLC34A2 | 4p15.2 | 10 (2.00%) | 8 (2.04%) | -0.03 | 1.00 | 1.00 | (B) Primary |
| NADK | 1p36.33 | 9 (0.86%) | 2 (0.26%) | 1.73 | 0.131 | 0.895 | (A) Metastasis |
| SERPINA1 | 14q32.13 | 9 (1.81%) | 11 (2.84%) | -0.65 | 0.365 | 1.00 | (B) Primary |
| SMARCD1 | 12q13.12 | 9 (0.40%) | 14 (0.62%) | -0.64 | 0.404 | 1.00 | (B) Primary |
| MBD4 | 3q21.3 | 9 (1.81%) | 4 (1.03%) | 0.81 | 0.408 | 1.00 | (A) Metastasis |
| NFKBIE | 6p21.1 | 9 (1.81%) | 4 (1.03%) | 0.81 | 0.408 | 1.00 | (A) Metastasis |
| HIST1H3D | 6p22.2 | 9 (0.41%) | 12 (0.56%) | -0.46 | 0.517 | 1.00 | (B) Primary |
| GAB1 | 4q31.21 | 9 (0.86%) | 4 (0.52%) | 0.73 | 0.575 | 1.00 | (A) Metastasis |
| KLLN | 10q23.31 | 9 (1.80%) | 5 (1.28%) | 0.49 | 0.598 | 1.00 | (A) Metastasis |
| FANCE | 6p21.31 | 9 (1.65%) | 6 (1.10%) | 0.58 | 0.605 | 1.00 | (A) Metastasis |
| SDHAF2 | 11q12.2 | 9 (0.32%) | 11 (0.40%) | -0.29 | 0.663 | 1.00 | (B) Primary |
| BCL10 | 1p22.3 | 9 (0.41%) | 7 (0.32%) | 0.32 | 0.804 | 1.00 | (A) Metastasis |
| XRCC6 | 22q13.2 | 9 (1.81%) | 7 (1.80%) | 0.01 | 1.00 | 1.00 | (A) Metastasis |
| SPRTN | 1q42.2 | 9 (0.86%) | 6 (0.78%) | 0.14 | 1.00 | 1.00 | (A) Metastasis |
| TERC | 3q26.2 | 8 (20.51%) | 4 (3.33%) | 2.62 | 1.599e-3 | 0.0719 | (A) Metastasis |
| EXT2 | 11p11.2 | 8 (1.51%) | 2 (0.38%) | 1.99 | 0.108 | 0.828 | (A) Metastasis |
| TDG | 12q23.3 | 8 (1.61%) | 2 (0.52%) | 1.64 | 0.200 | 1.00 | (A) Metastasis |
| BABAM2 | 2p23.2 | 8 (1.61%) | 3 (0.77%) | 1.06 | 0.364 | 1.00 | (A) Metastasis |
| EZHIP | Xp11.22 | 8 (0.76%) | 3 (0.39%) | 0.97 | 0.373 | 1.00 | (A) Metastasis |
| DDB2 | 11p11.2 | 8 (1.50%) | 4 (0.75%) | 0.99 | 0.385 | 1.00 | (A) Metastasis |
| ERCC1 | 19q13.32 | 8 (1.57%) | 4 (1.01%) | 0.64 | 0.566 | 1.00 | (A) Metastasis |
| HIST1H3G | 6p22.2 | 8 (0.36%) | 5 (0.23%) | 0.64 | 0.581 | 1.00 | (A) Metastasis |
| HMBS | 11q23.3 | 8 (1.61%) | 8 (2.06%) | -0.36 | 0.622 | 1.00 | (B) Primary |
| GPC3 | Xq26.2 | 8 (1.51%) | 6 (1.14%) | 0.40 | 0.789 | 1.00 | (A) Metastasis |
| CRTC1 | 19p13.11 | 8 (1.51%) | 9 (1.70%) | -0.17 | 0.813 | 1.00 | (B) Primary |
| XRCC3 | 14q32.33 | 8 (1.56%) | 7 (1.73%) | -0.15 | 1.00 | 1.00 | (B) Primary |
| TRAF3 | 14q32.32 | 8 (1.58%) | 6 (1.52%) | 0.06 | 1.00 | 1.00 | (A) Metastasis |
| KBTBD4 | 11p11.2 | 8 (0.76%) | 6 (0.78%) | -0.03 | 1.00 | 1.00 | (B) Primary |
| HIST1H3A | 6p22.2 | 8 (0.36%) | 7 (0.33%) | 0.15 | 1.00 | 1.00 | (A) Metastasis |
| LRP1B | 2q22.1-q22.2 | 7 (31.82%) | 3 (13.04%) | 1.29 | 0.165 | 1.00 | (A) Metastasis |
| NEIL3 | 4q34.3 | 7 (1.41%) | 2 (0.52%) | 1.45 | 0.312 | 1.00 | (A) Metastasis |
| RHOT1 | 17q11.2 | 7 (1.41%) | 2 (0.52%) | 1.45 | 0.312 | 1.00 | (A) Metastasis |
| MLLT1 | 19p13.3 | 7 (0.67%) | 7 (0.90%) | -0.44 | 0.596 | 1.00 | (B) Primary |
| TAZ | Xq28 | 7 (1.41%) | 8 (2.06%) | -0.55 | 0.601 | 1.00 | (B) Primary |
| CD74 | 5q33.1 | 7 (58.33%) | 7 (70.00%) | -0.26 | 0.675 | 1.00 | (B) Primary |
| ELANE | 19p13.3 | 7 (1.41%) | 4 (1.03%) | 0.45 | 0.764 | 1.00 | (A) Metastasis |
| FANCB | Xp22.2 | 7 (1.39%) | 4 (1.03%) | 0.44 | 0.764 | 1.00 | (A) Metastasis |
| CYP19A1 | 15q21.2 | 7 (0.67%) | 4 (0.52%) | 0.37 | 0.768 | 1.00 | (A) Metastasis |
| MAD2L2 | 1p36.22 | 7 (0.67%) | 4 (0.52%) | 0.36 | 0.768 | 1.00 | (A) Metastasis |
| UROD | 1p34.1 | 7 (1.41%) | 6 (1.55%) | -0.13 | 1.00 | 1.00 | (B) Primary |
| DCLRE1C | 10p13 | 7 (1.41%) | 5 (1.29%) | 0.13 | 1.00 | 1.00 | (A) Metastasis |
| NUTM1 | 15q14 | 7 (116.67%) | 5 (100.00%) | - | - | - | - |
| PDPK1 | 16p13.3 | 6 (0.27%) | 2 (0.09%) | 1.58 | 0.289 | 1.00 | (A) Metastasis |
| GNB1 | 1p36.33 | 6 (0.57%) | 2 (0.26%) | 1.14 | 0.479 | 1.00 | (A) Metastasis |
| NEIL1 | 15q24.2 | 6 (1.21%) | 7 (1.80%) | -0.58 | 0.576 | 1.00 | (B) Primary |
| NT5C2 | 10q24.32-q24.33 | 6 (1.19%) | 7 (1.77%) | -0.58 | 0.576 | 1.00 | (B) Primary |
| GALNT12 | 9q22.33 | 6 (1.17%) | 4 (1.00%) | 0.24 | 1.00 | 1.00 | (A) Metastasis |
| KMT5A | 12q24.31 | 6 (0.30%) | 6 (0.33%) | -0.12 | 1.00 | 1.00 | (B) Primary |
| MAP3K14 | 17q21.31 | 6 (0.27%) | 5 (0.23%) | 0.22 | 1.00 | 1.00 | (A) Metastasis |
| BCL2L12 | 19q13.33 | 5 (0.96%) | 1 (0.19%) | 2.32 | 0.217 | 1.00 | (A) Metastasis |
| CDKN1C | 11p15.4 | 5 (0.95%) | 1 (0.19%) | 2.32 | 0.217 | 1.00 | (A) Metastasis |
| OGG1 | 3p25.3 | 5 (1.01%) | 2 (0.52%) | 0.96 | 0.476 | 1.00 | (A) Metastasis |
| EGLN1 | 1q42.2 | 5 (1.00%) | 6 (1.54%) | -0.62 | 0.548 | 1.00 | (B) Primary |
| HIST1H3H | 6p22.1 | 5 (0.23%) | 7 (0.33%) | -0.53 | 0.576 | 1.00 | (B) Primary |
| HIST1H3I | 6p22.1 | 5 (0.23%) | 7 (0.33%) | -0.53 | 0.576 | 1.00 | (B) Primary |
| MST1 | 3p21.31 | 5 (0.23%) | 3 (0.14%) | 0.70 | 0.727 | 1.00 | (A) Metastasis |
| MAF | 16q23.2 | 5 (0.98%) | 5 (1.25%) | -0.35 | 0.756 | 1.00 | (B) Primary |
| HIST1H3C | 6p22.2 | 5 (0.18%) | 5 (0.20%) | -0.09 | 1.00 | 1.00 | (B) Primary |
| SYNE1 | 6q25.2 | 5 (45.45%) | 4 (57.14%) | -0.33 | 1.00 | 1.00 | (B) Primary |
| FAS | 10q23.31 | 5 (0.91%) | 4 (0.73%) | 0.32 | 1.00 | 1.00 | (A) Metastasis |
| RNF8 | 6p21.2 | 5 (1.01%) | 4 (1.03%) | -0.04 | 1.00 | 1.00 | (B) Primary |
| HLA-C | 6p21.33 | 5 (0.47%) | 3 (0.38%) | 0.30 | 1.00 | 1.00 | (A) Metastasis |
| CDC42 | 1p36.12 | 5 (0.25%) | 4 (0.22%) | 0.20 | 1.00 | 1.00 | (A) Metastasis |
| MAML2 | 11q21 | 4 (66.67%) | 1 (20.00%) | 1.74 | 0.242 | 1.00 | (A) Metastasis |
| PGAP3 | 17q12 | 4 (66.67%) | 1 (20.00%) | 1.74 | 0.242 | 1.00 | (A) Metastasis |
| GLI3 | 7p14.1 | 4 (15.38%) | 12 (8.89%) | 0.79 | 0.295 | 1.00 | (A) Metastasis |
| H19 | 11p15.5 | 4 (100.00%) | 1 (50.00%) | 1.00 | 0.333 | 1.00 | (A) Metastasis |
| TAF1 | Xq13.1 | 4 (19.05%) | 1 (4.76%) | 2.00 | 0.343 | 1.00 | (A) Metastasis |
| PDE4DIP | 1q21.2 | 4 (66.67%) | 2 (40.00%) | 0.74 | 0.567 | 1.00 | (A) Metastasis |
| PGBD3 | 10q11.23 | 4 (0.14%) | 3 (0.11%) | 0.42 | 0.726 | 1.00 | (A) Metastasis |
| DMC1 | 22q13.1 | 4 (0.80%) | 4 (1.03%) | -0.36 | 0.735 | 1.00 | (B) Primary |
| LMO2 | 11p13 | 4 (0.76%) | 5 (0.96%) | -0.32 | 0.753 | 1.00 | (B) Primary |
| CENPA | 2p23.3 | 4 (0.18%) | 3 (0.14%) | 0.37 | 1.00 | 1.00 | (A) Metastasis |
| CMTR2 | 16q22.2 | 4 (0.38%) | 2 (0.26%) | 0.56 | 1.00 | 1.00 | (A) Metastasis |
| FAAP20 | 1p36.33 | 4 (0.80%) | 3 (0.77%) | 0.06 | 1.00 | 1.00 | (A) Metastasis |
| ID4 | 6p22.3 | 4 (0.80%) | 3 (0.77%) | 0.06 | 1.00 | 1.00 | (A) Metastasis |
| KLF2 | 19p13.11 | 4 (0.80%) | 3 (0.77%) | 0.06 | 1.00 | 1.00 | (A) Metastasis |
| TCF7L1 | 2p11.2 | 3 (9.38%) | 3 (2.14%) | 2.13 | 0.0789 | 0.749 | (A) Metastasis |
| AKAP9 | 7q21.2 | 3 (50.00%) | 0 (0.00%) | >10 | 0.182 | 1.00 | (A) Metastasis |
| TLR4 | 9q33.1 | 3 (9.68%) | 4 (3.70%) | 1.39 | 0.185 | 1.00 | (A) Metastasis |
| FAH | 15q25.1 | 3 (0.60%) | 6 (1.55%) | -1.36 | 0.191 | 1.00 | (B) Primary |
| SLC25A21 | 14q13.3 | 3 (0.11%) | 0 (0.00%) | >10 | 0.250 | 1.00 | (A) Metastasis |
| FIP1L1 | 4q12 | 3 (33.33%) | 0 (0.00%) | >10 | 0.258 | 1.00 | (A) Metastasis |
| TAL2 | 9q31.2 | 3 (0.60%) | 0 (0.00%) | >10 | 0.261 | 1.00 | (A) Metastasis |
| RPA1 | 17p13.3 | 3 (0.59%) | 5 (1.24%) | -1.07 | 0.313 | 1.00 | (B) Primary |
| XPA | 9q22.33 | 3 (0.56%) | 5 (0.94%) | -0.75 | 0.504 | 1.00 | (B) Primary |
| BOD1L1 | 4p15.33 | 3 (50.00%) | 1 (20.00%) | 1.32 | 0.545 | 1.00 | (A) Metastasis |
| EP400 | 12q24.33 | 3 (50.00%) | 1 (20.00%) | 1.32 | 0.545 | 1.00 | (A) Metastasis |
| PKHD1 | 6p12.3-p12.2 | 3 (50.00%) | 1 (20.00%) | 1.32 | 0.545 | 1.00 | (A) Metastasis |
| SAMD9 | 7q21.2 | 3 (50.00%) | 1 (20.00%) | 1.32 | 0.545 | 1.00 | (A) Metastasis |
| NCOR2 | 12q24.31 | 3 (30.00%) | 1 (12.50%) | 1.26 | 0.588 | 1.00 | (A) Metastasis |
| NR0B1 | Xp21.2 | 3 (0.60%) | 4 (1.03%) | -0.77 | 0.705 | 1.00 | (B) Primary |
| PAX8 | 2q14.1 | 3 (23.08%) | 1 (12.50%) | 0.88 | 1.00 | 1.00 | (A) Metastasis |
| CCDC6 | 10q21.2 | 3 (75.00%) | 4 (200.00%) | - | - | - | - |
| CRBN | 3p26.2 | 2 (11.76%) | 0 (0.00%) | >10 | 0.204 | 1.00 | (A) Metastasis |
| STAG1 | 3q22.3 | 2 (6.90%) | 3 (2.21%) | 1.64 | 0.212 | 1.00 | (A) Metastasis |
| KDM6B | 17p13.1 | 2 (7.14%) | 4 (2.96%) | 1.27 | 0.274 | 1.00 | (A) Metastasis |
| SSX1 | Xp11.23 | 2 (25.00%) | 3 (60.00%) | -1.26 | 0.293 | 1.00 | (B) Primary |
| IKZF3 | 17q12-q21.1 | 2 (7.69%) | 5 (3.68%) | 1.07 | 0.312 | 1.00 | (A) Metastasis |
| CDH23 | 10q22.1 | 2 (22.22%) | 3 (50.00%) | -1.17 | 0.329 | 1.00 | (B) Primary |
| HFE | 6p22.2 | 2 (0.40%) | 4 (1.03%) | -1.36 | 0.413 | 1.00 | (B) Primary |
| RHOH | 4p14 | 2 (0.40%) | 4 (1.02%) | -1.36 | 0.413 | 1.00 | (B) Primary |
| CDH11 | 16q21 | 2 (33.33%) | 0 (0.00%) | >10 | 0.455 | 1.00 | (A) Metastasis |
| RBM15 | 1p13.3 | 2 (33.33%) | 0 (0.00%) | >10 | 0.455 | 1.00 | (A) Metastasis |
| HLA-DRB1 | 6p21.32 | 2 (50.00%) | 0 (0.00%) | >10 | 0.467 | 1.00 | (A) Metastasis |
| PRCC | 1q23.1 | 2 (20.00%) | 0 (0.00%) | >10 | 0.485 | 1.00 | (A) Metastasis |
| PAX3 | 2q36.1 | 2 (18.18%) | 0 (0.00%) | >10 | 0.485 | 1.00 | (A) Metastasis |
| IL6ST | 5q11.2 | 2 (25.00%) | 0 (0.00%) | >10 | 0.487 | 1.00 | (A) Metastasis |
| COPE | 19p13.11 | 2 (0.07%) | 0 (0.00%) | >10 | 0.500 | 1.00 | (A) Metastasis |
| GIPR | 19q13.32 | 2 (0.07%) | 0 (0.00%) | >10 | 0.500 | 1.00 | (A) Metastasis |
| GPR37L1 | 1q32.1 | 2 (0.07%) | 0 (0.00%) | >10 | 0.500 | 1.00 | (A) Metastasis |
| MED1 | 17q12 | 2 (0.07%) | 0 (0.00%) | >10 | 0.500 | 1.00 | (A) Metastasis |
| TTC28 | 22q12.1 | 2 (0.07%) | 0 (0.00%) | >10 | 0.500 | 1.00 | (A) Metastasis |
| AGO1 | 1p34.3 | 2 (0.19%) | 0 (0.00%) | >10 | 0.511 | 1.00 | (A) Metastasis |
| DNAH9 | 17p12 | 2 (28.57%) | 3 (60.00%) | -1.07 | 0.558 | 1.00 | (B) Primary |
| ACPP | 3q22.1 | 2 (0.07%) | 1 (0.04%) | 1.00 | 1.00 | 1.00 | (A) Metastasis |
| HCN1 | 5p12 | 2 (0.07%) | 1 (0.04%) | 1.00 | 1.00 | 1.00 | (A) Metastasis |
| PHF1 | 6p21.32 | 2 (0.07%) | 1 (0.04%) | 1.00 | 1.00 | 1.00 | (A) Metastasis |
| SNTG1 | 8q11.21 | 2 (0.07%) | 1 (0.04%) | 1.00 | 1.00 | 1.00 | (A) Metastasis |
| SRCIN1 | 17q12 | 2 (0.07%) | 1 (0.04%) | 1.00 | 1.00 | 1.00 | (A) Metastasis |
| TRAPPC9 | 8q24.3 | 2 (0.07%) | 1 (0.04%) | 1.00 | 1.00 | 1.00 | (A) Metastasis |
| YWHAE | 17p13.3 | 2 (0.07%) | 1 (0.04%) | 1.00 | 1.00 | 1.00 | (A) Metastasis |
| TPR | 1q31.1 | 2 (33.33%) | 2 (40.00%) | -0.26 | 1.00 | 1.00 | (B) Primary |
| TRIM24 | 7q33-q34 | 2 (33.33%) | 2 (40.00%) | -0.26 | 1.00 | 1.00 | (B) Primary |
| NUMA1 | 11q13.4 | 2 (33.33%) | 1 (20.00%) | 0.74 | 1.00 | 1.00 | (A) Metastasis |
| PKD1L2 | 16q23.2 | 2 (33.33%) | 1 (20.00%) | 0.74 | 1.00 | 1.00 | (A) Metastasis |
| RNF213 | 17q25.3 | 2 (33.33%) | 1 (20.00%) | 0.74 | 1.00 | 1.00 | (A) Metastasis |
| ADGRA2 | 8p11.23 | 2 (11.11%) | 2 (9.52%) | 0.22 | 1.00 | 1.00 | (A) Metastasis |
| PTPRK | 6q22.33 | 2 (0.07%) | 2 (0.07%) | - | 1.00 | 1.00 | (B) Primary |
| WRAP53 | 17p13.1 | 2 (0.07%) | 2 (0.07%) | - | 1.00 | 1.00 | (B) Primary |
| EXPH5 | 11q22.3 | 1 (100.00%) | 0 (0.00%) | >10 | 3.512e-4 | 0.0305 | (A) Metastasis |
| KIF3A | 5q31.1 | 1 (100.00%) | 0 (0.00%) | >10 | 3.512e-4 | 0.0305 | (A) Metastasis |
| LETM2 | 8p11.23 | 1 (100.00%) | 0 (0.00%) | >10 | 3.512e-4 | 0.0305 | (A) Metastasis |
| MCM2 | 3q21.3 | 1 (50.00%) | 0 (0.00%) | >10 | 7.022e-4 | 0.0406 | (A) Metastasis |
| MCM4 | 8q11.21 | 1 (50.00%) | 0 (0.00%) | >10 | 7.022e-4 | 0.0406 | (A) Metastasis |
| PTPN1 | 20q13.13 | 1 (50.00%) | 0 (0.00%) | >10 | 7.022e-4 | 0.0406 | (A) Metastasis |
| USP6 | 17p13.2 | 1 (50.00%) | 0 (0.00%) | >10 | 7.022e-4 | 0.0406 | (A) Metastasis |
| EHD1 | 11q13.1 | 1 (100.00%) | 1 (0.04%) | >10 | 7.025e-4 | 0.0406 | (A) Metastasis |
| RMRP | 9p13.3 | 1 (0.04%) | 1 (100.00%) | <-10 | 7.025e-4 | 0.0406 | (B) Primary |
| CLTC | 17q23.1 | 1 (50.00%) | 1 (0.04%) | >10 | 1.404e-3 | 0.0656 | (A) Metastasis |
| SLC45A3 | 1q32.1 | 1 (50.00%) | 1 (0.04%) | >10 | 1.404e-3 | 0.0656 | (A) Metastasis |
| PSMD13 | 11p15.5 | 1 (3.85%) | 0 (0.00%) | >10 | 0.161 | 1.00 | (A) Metastasis |
| SQSTM1 | 5q35.3 | 1 (4.00%) | 0 (0.00%) | >10 | 0.195 | 1.00 | (A) Metastasis |
| NCOA2 | 8q13.3 | 1 (16.67%) | 3 (60.00%) | -1.85 | 0.242 | 1.00 | (B) Primary |
| LMO3 | 12p12.3 | 1 (3.85%) | 1 (0.74%) | 2.38 | 0.298 | 1.00 | (A) Metastasis |
| SCG5 | 15q13.3 | 1 (0.10%) | 3 (0.39%) | -2.03 | 0.317 | 1.00 | (B) Primary |
| SMC1A | Xp11.22 | 1 (2.56%) | 1 (0.69%) | 1.88 | 0.382 | 1.00 | (A) Metastasis |
| FKBP9 | 7p14.3 | 1 (3.85%) | 2 (1.48%) | 1.38 | 0.413 | 1.00 | (A) Metastasis |
| SF1 | 11q13.1 | 1 (3.57%) | 2 (1.48%) | 1.27 | 0.434 | 1.00 | (A) Metastasis |
| TNKS | 8p23.1 | 1 (10.00%) | 0 (0.00%) | >10 | 0.455 | 1.00 | (A) Metastasis |
| ARFRP1 | 20q13.33 | 1 (4.76%) | 0 (0.00%) | >10 | 0.457 | 1.00 | (A) Metastasis |
| GID4 | 17p11.2\|17p11.2 | 1 (4.76%) | 0 (0.00%) | >10 | 0.457 | 1.00 | (A) Metastasis |
| EPHB4 | 7q22.1 | 1 (5.26%) | 0 (0.00%) | >10 | 0.475 | 1.00 | (A) Metastasis |
| CADM2 | 3p12.1 | 1 (4.00%) | 2 (1.94%) | 1.04 | 0.482 | 1.00 | (A) Metastasis |
| SLITRK6 | 13q31.1 | 1 (4.00%) | 2 (1.94%) | 1.04 | 0.482 | 1.00 | (A) Metastasis |
| ACVR1B | 12q13.13 | 1 (5.56%) | 0 (0.00%) | >10 | 0.486 | 1.00 | (A) Metastasis |
| DPYD | 1p21.3 | 1 (4.76%) | 0 (0.00%) | >10 | 0.488 | 1.00 | (A) Metastasis |
| PRAME | 22q11.22 | 1 (3.85%) | 3 (2.22%) | 0.79 | 0.509 | 1.00 | (A) Metastasis |
| NUP214 | 9q34.13 | 1 (12.50%) | 2 (40.00%) | -1.68 | 0.510 | 1.00 | (B) Primary |
| ATF1 | 12q13.12 | 1 (16.67%) | 2 (40.00%) | -1.26 | 0.545 | 1.00 | (B) Primary |
| MTR | 1q43 | 1 (16.67%) | 2 (40.00%) | -1.26 | 0.545 | 1.00 | (B) Primary |
| NCOA4 | 10q11.22 | 1 (16.67%) | 2 (40.00%) | -1.26 | 0.545 | 1.00 | (B) Primary |
| SEPTIN9 | 17q25.3 | 1 (16.67%) | 2 (40.00%) | -1.26 | 0.545 | 1.00 | (B) Primary |
| TGM7 | 15q15.2-q15.3 | 1 (16.67%) | 2 (40.00%) | -1.26 | 0.545 | 1.00 | (B) Primary |
| DMD | Xp21.2-p21.1 | 1 (3.85%) | 4 (2.96%) | 0.38 | 0.591 | 1.00 | (A) Metastasis |
| PTK2 | 8q24.3 | 1 (3.85%) | 4 (2.96%) | 0.38 | 0.591 | 1.00 | (A) Metastasis |
| TYK2 | 19p13.2 | 1 (16.67%) | 2 (33.33%) | -1.00 | 0.591 | 1.00 | (B) Primary |
| ABL2 | 1q25.2 | 1 (4.76%) | 2 (10.00%) | -1.07 | 0.606 | 1.00 | (B) Primary |
| FGF14 | 13q33.1 | 1 (4.00%) | 3 (11.11%) | -1.47 | 0.611 | 1.00 | (B) Primary |
| ABHD17A | 19p13.3 | 1 (0.04%) | 1 (0.04%) | - | 1.00 | 1.00 | (B) Primary |
| ACACA | 17q12 | 1 (0.04%) | 1 (0.04%) | - | 1.00 | 1.00 | (B) Primary |
| ACADVL | 17p13.1 | 1 (0.04%) | 1 (0.04%) | - | 1.00 | 1.00 | (B) Primary |
| ADAD1 | 4q27 | 1 (0.04%) | 1 (0.04%) | - | 1.00 | 1.00 | (B) Primary |
| ARHGEF3 | 3p14.3 | 1 (0.04%) | 1 (0.04%) | - | 1.00 | 1.00 | (B) Primary |
| ATP6V1E1 | 22q11.21 | 1 (0.04%) | 1 (0.04%) | - | 1.00 | 1.00 | (B) Primary |
| BCAS3 | 17q23.2 | 1 (0.04%) | 1 (0.04%) | - | 1.00 | 1.00 | (B) Primary |
| BRMS1L | 14q13.2 | 1 (0.04%) | 1 (0.04%) | - | 1.00 | 1.00 | (B) Primary |
| CACNA1B | 9q34.3 | 1 (0.04%) | 1 (0.04%) | - | 1.00 | 1.00 | (B) Primary |
| CADM3 | 1q23.2 | 1 (0.04%) | 1 (0.04%) | - | 1.00 | 1.00 | (B) Primary |
| CASC8 | 8q24.21 | 1 (0.04%) | 1 (0.04%) | - | 1.00 | 1.00 | (B) Primary |
| CCDC149 | 4p15.2 | 1 (0.04%) | 1 (0.04%) | - | 1.00 | 1.00 | (B) Primary |
| CDKN2A-DT | 9p21.3 | 1 (0.04%) | 1 (0.04%) | - | 1.00 | 1.00 | (B) Primary |
| CELF5 | 19p13.3 | 1 (0.04%) | 1 (0.04%) | - | 1.00 | 1.00 | (B) Primary |
| CITED1 | Xq13.1 | 1 (0.04%) | 1 (0.04%) | - | 1.00 | 1.00 | (B) Primary |
| CNOT6L | 4q21.1 | 1 (0.04%) | 1 (0.04%) | - | 1.00 | 1.00 | (B) Primary |
| COL5A1 | 9q34.3 | 1 (0.04%) | 1 (0.04%) | - | 1.00 | 1.00 | (B) Primary |
| CPA6 | 8q13.2 | 1 (0.04%) | 1 (0.04%) | - | 1.00 | 1.00 | (B) Primary |
| CREB3L1 | 11p11.2 | 1 (0.04%) | 1 (0.04%) | - | 1.00 | 1.00 | (B) Primary |
| CREM | 10p11.21 | 1 (0.04%) | 1 (0.04%) | - | 1.00 | 1.00 | (B) Primary |
| CSMD1 | 8p23.2 | 1 (0.04%) | 1 (0.04%) | - | 1.00 | 1.00 | (B) Primary |
| CYHR1 | 8q24.3 | 1 (0.04%) | 1 (0.04%) | - | 1.00 | 1.00 | (B) Primary |
| DCUN1D4 | 4q12 | 1 (0.04%) | 1 (0.04%) | - | 1.00 | 1.00 | (B) Primary |
| DHH | 12q13.12 | 1 (0.04%) | 1 (0.04%) | - | 1.00 | 1.00 | (B) Primary |
| DIPK1A | 1p22.1 | 1 (0.04%) | 1 (0.04%) | - | 1.00 | 1.00 | (B) Primary |
| DLGAP5 | 14q22.3 | 1 (0.04%) | 1 (0.04%) | - | 1.00 | 1.00 | (B) Primary |
| ERN1 | 17q23.3 | 1 (0.04%) | 1 (0.04%) | - | 1.00 | 1.00 | (B) Primary |
| EYA2 | 20q13.12 | 1 (0.04%) | 1 (0.04%) | - | 1.00 | 1.00 | (B) Primary |
| FLACC1 | 2q33.1 | 1 (0.04%) | 1 (0.04%) | - | 1.00 | 1.00 | (B) Primary |
| FUBP3 | 9q34.11-q34.12 | 1 (0.04%) | 1 (0.04%) | - | 1.00 | 1.00 | (B) Primary |
| GALM | 2p22.1 | 1 (0.04%) | 1 (0.04%) | - | 1.00 | 1.00 | (B) Primary |
| GNE | 9p13.3 | 1 (0.04%) | 1 (0.04%) | - | 1.00 | 1.00 | (B) Primary |
| GPLD1 | 6p22.3 | 1 (0.04%) | 1 (0.04%) | - | 1.00 | 1.00 | (B) Primary |
| GRIPAP1 | Xp11.23 | 1 (0.04%) | 1 (0.04%) | - | 1.00 | 1.00 | (B) Primary |
| HUNK | 21q22.11 | 1 (0.04%) | 1 (0.04%) | - | 1.00 | 1.00 | (B) Primary |
| IQSEC1 | 3p25.2-p25.1 | 1 (0.04%) | 1 (0.04%) | - | 1.00 | 1.00 | (B) Primary |
| KCNK13 | 14q32.11 | 1 (0.04%) | 1 (0.04%) | - | 1.00 | 1.00 | (B) Primary |
| KCTD16 | 5q31.3 | 1 (0.04%) | 1 (0.04%) | - | 1.00 | 1.00 | (B) Primary |
| KDELR1 | 19q13.33 | 1 (0.04%) | 1 (0.04%) | - | 1.00 | 1.00 | (B) Primary |
| KIF16B | 20p12.1 | 1 (0.04%) | 1 (0.04%) | - | 1.00 | 1.00 | (B) Primary |
| LCP2 | 5q35.1 | 1 (0.04%) | 1 (0.04%) | - | 1.00 | 1.00 | (B) Primary |
| LIMA1 | 12q13.12 | 1 (0.04%) | 1 (0.04%) | - | 1.00 | 1.00 | (B) Primary |
| LIMK2 | 22q12.2 | 1 (0.04%) | 1 (0.04%) | - | 1.00 | 1.00 | (B) Primary |
| LPCAT2 | 16q12.2 | 1 (0.04%) | 1 (0.04%) | - | 1.00 | 1.00 | (B) Primary |
| LRP1 | 12q13.3 | 1 (0.04%) | 1 (0.04%) | - | 1.00 | 1.00 | (B) Primary |
| MAP3K3 | 17q23.3 | 1 (0.04%) | 1 (0.04%) | - | 1.00 | 1.00 | (B) Primary |
| MFSD4A | 1q32.1 | 1 (0.04%) | 1 (0.04%) | - | 1.00 | 1.00 | (B) Primary |
| MOK | 14q32.31 | 1 (0.04%) | 1 (0.04%) | - | 1.00 | 1.00 | (B) Primary |
| MROH8 | 20q11.23 | 1 (0.04%) | 1 (0.04%) | - | 1.00 | 1.00 | (B) Primary |
| NAV1 | 1q32.1 | 1 (0.04%) | 1 (0.04%) | - | 1.00 | 1.00 | (B) Primary |
| NELFA | 4p16.3 | 1 (0.04%) | 1 (0.04%) | - | 1.00 | 1.00 | (B) Primary |
| NEUROD2 | 17q12 | 1 (0.04%) | 1 (0.04%) | - | 1.00 | 1.00 | (B) Primary |
| NGRN | 15q26.1 | 1 (0.04%) | 1 (0.04%) | - | 1.00 | 1.00 | (B) Primary |
| NKX2-8 | 14q13.3 | 1 (0.04%) | 1 (0.04%) | - | 1.00 | 1.00 | (B) Primary |
| NOL4 | 18q12.1 | 1 (0.04%) | 1 (0.04%) | - | 1.00 | 1.00 | (B) Primary |
| NR3C2 | 4q31.23 | 1 (0.04%) | 1 (0.04%) | - | 1.00 | 1.00 | (B) Primary |
| NRBF2 | 10q21.3 | 1 (0.04%) | 1 (0.04%) | - | 1.00 | 1.00 | (B) Primary |
| NTN5 | 19q13.33 | 1 (0.04%) | 1 (0.04%) | - | 1.00 | 1.00 | (B) Primary |
| NUDC | 1p36.11 | 1 (0.04%) | 1 (0.04%) | - | 1.00 | 1.00 | (B) Primary |
| OPTN | 10p13 | 1 (0.04%) | 1 (0.04%) | - | 1.00 | 1.00 | (B) Primary |
| P3H1 | 1p34.2 | 1 (0.04%) | 1 (0.04%) | - | 1.00 | 1.00 | (B) Primary |
| PAXIP1-AS2 | 7q36.2 | 1 (0.04%) | 1 (0.04%) | - | 1.00 | 1.00 | (B) Primary |
| PLPP4 | 10q26.12 | 1 (0.04%) | 1 (0.04%) | - | 1.00 | 1.00 | (B) Primary |
| PNPLA7 | 9q34.3 | 1 (0.04%) | 1 (0.04%) | - | 1.00 | 1.00 | (B) Primary |
| POLG | 15q26.1 | 1 (0.04%) | 1 (0.04%) | - | 1.00 | 1.00 | (B) Primary |
| PTCD3 | 2p11.2 | 1 (0.04%) | 1 (0.04%) | - | 1.00 | 1.00 | (B) Primary |
| RAD23A | 19p13.13 | 1 (0.04%) | 1 (0.04%) | - | 1.00 | 1.00 | (B) Primary |
| RBMS2 | 12q13.3 | 1 (0.04%) | 1 (0.04%) | - | 1.00 | 1.00 | (B) Primary |
| RIPK2 | 8q21.3 | 1 (0.04%) | 1 (0.04%) | - | 1.00 | 1.00 | (B) Primary |
| SEC1P | 19q13.33 | 1 (0.04%) | 1 (0.04%) | - | 1.00 | 1.00 | (B) Primary |
| SEPTIN14 | 7p11.2 | 1 (0.04%) | 1 (0.04%) | - | 1.00 | 1.00 | (B) Primary |
| SHTN1 | 10q25.3 | 1 (0.04%) | 1 (0.04%) | - | 1.00 | 1.00 | (B) Primary |
| SLC4A4 | 4q13.3 | 1 (0.04%) | 1 (0.04%) | - | 1.00 | 1.00 | (B) Primary |
| SOX6 | 11p15.2 | 1 (0.04%) | 1 (0.04%) | - | 1.00 | 1.00 | (B) Primary |
| SPATS2 | 12q13.12 | 1 (0.04%) | 1 (0.04%) | - | 1.00 | 1.00 | (B) Primary |
| SPPL3 | 12q24.31 | 1 (0.04%) | 1 (0.04%) | - | 1.00 | 1.00 | (B) Primary |
| SPTBN1 | 2p16.2 | 1 (0.04%) | 1 (0.04%) | - | 1.00 | 1.00 | (B) Primary |
| ST6GALNAC3 | 1p31.1 | 1 (0.04%) | 1 (0.04%) | - | 1.00 | 1.00 | (B) Primary |
| STRN | 2p22.2 | 1 (0.04%) | 1 (0.04%) | - | 1.00 | 1.00 | (B) Primary |
| SUPT6H | 17q11.2 | 1 (0.04%) | 1 (0.04%) | - | 1.00 | 1.00 | (B) Primary |
| THSD4 | 15q23 | 1 (0.04%) | 1 (0.04%) | - | 1.00 | 1.00 | (B) Primary |
| TMEM117 | 12q12 | 1 (0.04%) | 1 (0.04%) | - | 1.00 | 1.00 | (B) Primary |
| TNIP1 | 5q33.1 | 1 (0.04%) | 1 (0.04%) | - | 1.00 | 1.00 | (B) Primary |
| TRHDE | 12q21.1 | 1 (0.04%) | 1 (0.04%) | - | 1.00 | 1.00 | (B) Primary |
| TRIO | 5p15.2 | 1 (0.04%) | 1 (0.04%) | - | 1.00 | 1.00 | (B) Primary |
| TSPAN16 | 19p13.2 | 1 (0.04%) | 1 (0.04%) | - | 1.00 | 1.00 | (B) Primary |
| TUBGCP4 | 15q15.3 | 1 (0.04%) | 1 (0.04%) | - | 1.00 | 1.00 | (B) Primary |
| XKR4 | 8q12.1 | 1 (0.04%) | 1 (0.04%) | - | 1.00 | 1.00 | (B) Primary |
| ZNF423 | 16q12.1 | 1 (0.04%) | 1 (0.04%) | - | 1.00 | 1.00 | (B) Primary |
| ZNF83 | 19q13.41 | 1 (0.04%) | 1 (0.04%) | - | 1.00 | 1.00 | (B) Primary |
| BORCS8-MEF2B | 19p13.11 | 1 (0.04%) | 2 (0.07%) | -1.00 | 1.00 | 1.00 | (B) Primary |
| PRKACA | 19p13.12 | 1 (0.04%) | 2 (0.07%) | -1.00 | 1.00 | 1.00 | (B) Primary |
| USP34 | 2p15 | 1 (0.04%) | 2 (0.07%) | -1.00 | 1.00 | 1.00 | (B) Primary |
| SRP19 | 5q22.2 | 1 (0.22%) | 0 (0.00%) | >10 | 1.00 | 1.00 | (A) Metastasis |
| KLHL6 | 3q27.1 | 1 (3.70%) | 0 (0.00%) | >10 | 1.00 | 1.00 | (A) Metastasis |
| ZNF703 | 8p11.23 | 1 (4.76%) | 1 (4.00%) | 0.25 | 1.00 | 1.00 | (A) Metastasis |
| PAK3 | Xq23 | 1 (5.56%) | 1 (4.76%) | 0.22 | 1.00 | 1.00 | (A) Metastasis |
| SLIT2 | 4p15.31 | 1 (6.67%) | 2 (13.33%) | -1.00 | 1.00 | 1.00 | (B) Primary |
| TOP2A | 17q21.2 | 1 (6.67%) | 2 (13.33%) | -1.00 | 1.00 | 1.00 | (B) Primary |
| ETS1 | 11q24.3 | 1 (9.09%) | 1 (11.11%) | -0.29 | 1.00 | 1.00 | (B) Primary |
| EZR | 6q25.3 | 1 (9.09%) | 1 (11.11%) | -0.29 | 1.00 | 1.00 | (B) Primary |
| NUP98 | 11p15.4 | 1 (8.33%) | 0 (0.00%) | >10 | 1.00 | 1.00 | (A) Metastasis |
| FLI1 | 11q24.3 | 1 (14.29%) | 1 (16.67%) | -0.22 | 1.00 | 1.00 | (B) Primary |
| TCF12 | 15q21.3 | 1 (12.50%) | 0 (0.00%) | >10 | 1.00 | 1.00 | (A) Metastasis |
| MAGI2 | 7q21.11 | 1 (6.67%) | 0 (0.00%) | >10 | 1.00 | 1.00 | (A) Metastasis |
| RANBP2 | 2q13 | 1 (6.67%) | 0 (0.00%) | >10 | 1.00 | 1.00 | (A) Metastasis |
| SDC4 | 20q13.12 | 1 (33.33%) | 1 (25.00%) | 0.42 | 1.00 | 1.00 | (A) Metastasis |
| HDAC4 | 2q37.3 | 1 (25.00%) | 0 (0.00%) | >10 | 1.00 | 1.00 | (A) Metastasis |
| ZBTB7A | 19p13.3 | 1 (100.00%) | 0 (0.00%) | >10 | 1.00 | 1.00 | (A) Metastasis |
| C3ORF70 | 3q27.2 | 1 (25.00%) | 0 (0.00%) | >10 | 1.00 | 1.00 | (A) Metastasis |
| IFNL3 | 19q13.2 | 1 (25.00%) | 0 (0.00%) | >10 | 1.00 | 1.00 | (A) Metastasis |
| PIAS4 | 19p13.3 | 1 (25.00%) | 0 (0.00%) | >10 | 1.00 | 1.00 | (A) Metastasis |
| TIGIT | 3q13.31 | 1 (25.00%) | 0 (0.00%) | >10 | 1.00 | 1.00 | (A) Metastasis |
| LCK | 1p35.2 | 1 (10.00%) | 0 (0.00%) | >10 | 1.00 | 1.00 | (A) Metastasis |
| MTRR | 5p15.31 | 1 (10.00%) | 0 (0.00%) | >10 | 1.00 | 1.00 | (A) Metastasis |
| TPM3 | 1q21.3 | 1 (12.50%) | 1 (20.00%) | -0.68 | 1.00 | 1.00 | (B) Primary |
| ADAMTS20 | 12q12 | 1 (16.67%) | 1 (20.00%) | -0.26 | 1.00 | 1.00 | (B) Primary |
| ADGRL3 | 4q13.1 | 1 (16.67%) | 1 (20.00%) | -0.26 | 1.00 | 1.00 | (B) Primary |
| ANKRD24 | 19p13.3 | 1 (16.67%) | 1 (20.00%) | -0.26 | 1.00 | 1.00 | (B) Primary |
| BRINP3 | 1q31.1 | 1 (16.67%) | 1 (20.00%) | -0.26 | 1.00 | 1.00 | (B) Primary |
| CACNA1E | 1q25.3 | 1 (16.67%) | 1 (20.00%) | -0.26 | 1.00 | 1.00 | (B) Primary |
| CREB1 | 2q33.3 | 1 (16.67%) | 1 (20.00%) | -0.26 | 1.00 | 1.00 | (B) Primary |
| DEK | 6p22.3 | 1 (16.67%) | 1 (20.00%) | -0.26 | 1.00 | 1.00 | (B) Primary |
| ITGB2 | 21q22.3 | 1 (16.67%) | 1 (20.00%) | -0.26 | 1.00 | 1.00 | (B) Primary |
| MAGEA1 | Xq28 | 1 (16.67%) | 1 (20.00%) | -0.26 | 1.00 | 1.00 | (B) Primary |
| NIN | 14q22.1 | 1 (16.67%) | 1 (20.00%) | -0.26 | 1.00 | 1.00 | (B) Primary |
| PLEKHG5 | 1p36.31 | 1 (16.67%) | 1 (20.00%) | -0.26 | 1.00 | 1.00 | (B) Primary |
| ZNF521 | 18q11.2 | 1 (16.67%) | 1 (20.00%) | -0.26 | 1.00 | 1.00 | (B) Primary |
| ACVR2A | 2q22.3-q23.1 | 1 (16.67%) | 0 (0.00%) | >10 | 1.00 | 1.00 | (A) Metastasis |
| ADGRB3 | 6q12-q13 | 1 (16.67%) | 0 (0.00%) | >10 | 1.00 | 1.00 | (A) Metastasis |
| ARNT | 1q21.3 | 1 (16.67%) | 0 (0.00%) | >10 | 1.00 | 1.00 | (A) Metastasis |
| CMPK1 | 1p33 | 1 (16.67%) | 0 (0.00%) | >10 | 1.00 | 1.00 | (A) Metastasis |
| CSMD3 | 8q23.3 | 1 (16.67%) | 0 (0.00%) | >10 | 1.00 | 1.00 | (A) Metastasis |
| CYP2C19 | 10q23.33 | 1 (16.67%) | 0 (0.00%) | >10 | 1.00 | 1.00 | (A) Metastasis |
| FOXP4 | 6p21.1 | 1 (16.67%) | 0 (0.00%) | >10 | 1.00 | 1.00 | (A) Metastasis |
| GNAI3 | 1p13.3 | 1 (16.67%) | 0 (0.00%) | >10 | 1.00 | 1.00 | (A) Metastasis |
| GUCY1A2 | 11q22.3 | 1 (16.67%) | 0 (0.00%) | >10 | 1.00 | 1.00 | (A) Metastasis |
| HSP90AB1 | 6p21.1 | 1 (16.67%) | 0 (0.00%) | >10 | 1.00 | 1.00 | (A) Metastasis |
| LPP | 3q27.3-q28 | 1 (16.67%) | 0 (0.00%) | >10 | 1.00 | 1.00 | (A) Metastasis |
| PRDM16 | 1p36.32 | 1 (16.67%) | 0 (0.00%) | >10 | 1.00 | 1.00 | (A) Metastasis |
| PTGS2 | 1q31.1 | 1 (16.67%) | 0 (0.00%) | >10 | 1.00 | 1.00 | (A) Metastasis |
| RNASEL | 1q25.3 | 1 (16.67%) | 0 (0.00%) | >10 | 1.00 | 1.00 | (A) Metastasis |
| RPN1 | 3q21.3 | 1 (16.67%) | 0 (0.00%) | >10 | 1.00 | 1.00 | (A) Metastasis |
| SP140 | 2q37.1 | 1 (16.67%) | 0 (0.00%) | >10 | 1.00 | 1.00 | (A) Metastasis |
| TLX1 | 10q24.31 | 1 (16.67%) | 0 (0.00%) | >10 | 1.00 | 1.00 | (A) Metastasis |
| TRIM33 | 1p13.2 | 1 (16.67%) | 0 (0.00%) | >10 | 1.00 | 1.00 | (A) Metastasis |
| USP9X | Xp11.4 | 1 (16.67%) | 0 (0.00%) | >10 | 1.00 | 1.00 | (A) Metastasis |
| ZMYM2 | 13q12.11 | 1 (16.67%) | 0 (0.00%) | >10 | 1.00 | 1.00 | (A) Metastasis |
| TCL1A | 14q32.13 | 1 (10.00%) | 0 (0.00%) | >10 | 1.00 | 1.00 | (A) Metastasis |
| MERTK | 2q13 | 1 (7.69%) | 1 (6.25%) | 0.30 | 1.00 | 1.00 | (A) Metastasis |
| MN1 | 22q12.1 | 1 (10.00%) | 1 (14.29%) | -0.51 | 1.00 | 1.00 | (B) Primary |
| CDH2 | 18q12.1 | 1 (6.25%) | 1 (5.88%) | 0.09 | 1.00 | 1.00 | (A) Metastasis |
| ABCC4 | 13q32.1 | 1 (9.09%) | 1 (6.67%) | 0.45 | 1.00 | 1.00 | (A) Metastasis |
| EMSY | 11q13.5 | 1 (4.00%) | 1 (3.70%) | 0.11 | 1.00 | 1.00 | (A) Metastasis |
| CYP2D6 | 22q13.2 | 1 (4.76%) | 2 (9.09%) | -0.93 | 1.00 | 1.00 | (B) Primary |
| NPRL3 | 16p13.3 | 1 (4.00%) | 4 (3.88%) | 0.04 | 1.00 | 1.00 | (A) Metastasis |
| DEPDC5 | 22q12.2-q12.3 | 1 (4.00%) | 3 (2.91%) | 0.46 | 1.00 | 1.00 | (A) Metastasis |
| SLX1B | 16p11.2 | 1 (0.20%) | 1 (0.26%) | -0.36 | 1.00 | 1.00 | (B) Primary |
| KDM4C | 9p24.1 | 1 (0.04%) | 0 (0.00%) | >10 | 1.00 | 1.00 | (A) Metastasis |
| ABHD3 | 18q11.2 | 1 (0.04%) | 0 (0.00%) | >10 | 1.00 | 1.00 | (A) Metastasis |
| ABLIM3 | 5q32 | 1 (0.04%) | 0 (0.00%) | >10 | 1.00 | 1.00 | (A) Metastasis |
| ACAN | 15q26.1 | 1 (0.04%) | 0 (0.00%) | >10 | 1.00 | 1.00 | (A) Metastasis |
| ADAM18 | 8p11.22 | 1 (0.04%) | 0 (0.00%) | >10 | 1.00 | 1.00 | (A) Metastasis |
| ADAM2 | 8p11.22 | 1 (0.04%) | 0 (0.00%) | >10 | 1.00 | 1.00 | (A) Metastasis |
| ADCY10P1 | 6p21.1 | 1 (0.04%) | 0 (0.00%) | >10 | 1.00 | 1.00 | (A) Metastasis |
| AGBL4 | 1p33 | 1 (0.04%) | 0 (0.00%) | >10 | 1.00 | 1.00 | (A) Metastasis |
| AGT | 1q42.2 | 1 (0.04%) | 0 (0.00%) | >10 | 1.00 | 1.00 | (A) Metastasis |
| AHI1 | 6q23.3 | 1 (0.04%) | 0 (0.00%) | >10 | 1.00 | 1.00 | (A) Metastasis |
| AKAP12 | 6q25.1 | 1 (0.04%) | 0 (0.00%) | >10 | 1.00 | 1.00 | (A) Metastasis |
| APOLD1 | 12p13.1 | 1 (0.04%) | 0 (0.00%) | >10 | 1.00 | 1.00 | (A) Metastasis |
| ATAD2B | 2p24.1-p23.3 | 1 (0.04%) | 0 (0.00%) | >10 | 1.00 | 1.00 | (A) Metastasis |
| ATAD5 | 17q11.2 | 1 (0.04%) | 0 (0.00%) | >10 | 1.00 | 1.00 | (A) Metastasis |
| ATRIP | 3p21.31 | 1 (0.04%) | 0 (0.00%) | >10 | 1.00 | 1.00 | (A) Metastasis |
| ATRN | 20p13 | 1 (0.04%) | 0 (0.00%) | >10 | 1.00 | 1.00 | (A) Metastasis |
| BAIAP2 | 17q25.3 | 1 (0.04%) | 0 (0.00%) | >10 | 1.00 | 1.00 | (A) Metastasis |
| BAZ2B | 2q24.2 | 1 (0.04%) | 0 (0.00%) | >10 | 1.00 | 1.00 | (A) Metastasis |
| BCAS4 | 20q13.13 | 1 (0.04%) | 0 (0.00%) | >10 | 1.00 | 1.00 | (A) Metastasis |
| BIVM | 13q33.1 | 1 (0.04%) | 0 (0.00%) | >10 | 1.00 | 1.00 | (A) Metastasis |
| BMPR2 | 2q33.1-q33.2 | 1 (0.04%) | 0 (0.00%) | >10 | 1.00 | 1.00 | (A) Metastasis |
| C15ORF41 | 15q14 | 1 (0.04%) | 0 (0.00%) | >10 | 1.00 | 1.00 | (A) Metastasis |
| C17ORF64 | 17q23.2 | 1 (0.04%) | 0 (0.00%) | >10 | 1.00 | 1.00 | (A) Metastasis |
| C1ORF147 | 1q32.1 | 1 (0.04%) | 0 (0.00%) | >10 | 1.00 | 1.00 | (A) Metastasis |
| C2ORF83 | 2q36.3 | 1 (0.04%) | 0 (0.00%) | >10 | 1.00 | 1.00 | (A) Metastasis |
| CACNA1A | 19p13.13 | 1 (0.04%) | 0 (0.00%) | >10 | 1.00 | 1.00 | (A) Metastasis |
| CAPZB | 1p36.13 | 1 (0.04%) | 0 (0.00%) | >10 | 1.00 | 1.00 | (A) Metastasis |
| CASP3 | 4q35.1 | 1 (0.04%) | 0 (0.00%) | >10 | 1.00 | 1.00 | (A) Metastasis |
| CASZ1 | 1p36.22 | 1 (0.04%) | 0 (0.00%) | >10 | 1.00 | 1.00 | (A) Metastasis |
| CCDC57 | 17q25.3 | 1 (0.04%) | 0 (0.00%) | >10 | 1.00 | 1.00 | (A) Metastasis |
| CDH13 | 16q23.3 | 1 (0.04%) | 0 (0.00%) | >10 | 1.00 | 1.00 | (A) Metastasis |
| CEP85 | 1p36.11 | 1 (0.04%) | 0 (0.00%) | >10 | 1.00 | 1.00 | (A) Metastasis |
| CERS2 | 1q21.3 | 1 (0.04%) | 0 (0.00%) | >10 | 1.00 | 1.00 | (A) Metastasis |
| CFAP300 | 11q22.1 | 1 (0.04%) | 0 (0.00%) | >10 | 1.00 | 1.00 | (A) Metastasis |
| CLIP1 | 12q24.31 | 1 (0.04%) | 0 (0.00%) | >10 | 1.00 | 1.00 | (A) Metastasis |
| CLSTN2 | 3q23 | 1 (0.04%) | 0 (0.00%) | >10 | 1.00 | 1.00 | (A) Metastasis |
| CNNM2 | 10q24.32 | 1 (0.04%) | 0 (0.00%) | >10 | 1.00 | 1.00 | (A) Metastasis |
| CPHL1P | 3q25.1 | 1 (0.04%) | 0 (0.00%) | >10 | 1.00 | 1.00 | (A) Metastasis |
| CRHR2 | 7p14.3 | 1 (0.04%) | 0 (0.00%) | >10 | 1.00 | 1.00 | (A) Metastasis |
| CRK | 17p13.3 | 1 (0.04%) | 0 (0.00%) | >10 | 1.00 | 1.00 | (A) Metastasis |
| CTNND1 | 11q12.1 | 1 (0.04%) | 0 (0.00%) | >10 | 1.00 | 1.00 | (A) Metastasis |
| DOCK6 | 19p13.2 | 1 (0.04%) | 0 (0.00%) | >10 | 1.00 | 1.00 | (A) Metastasis |
| DOCK9 | 13q32.3 | 1 (0.04%) | 0 (0.00%) | >10 | 1.00 | 1.00 | (A) Metastasis |
| DTL | 1q32.3 | 1 (0.04%) | 0 (0.00%) | >10 | 1.00 | 1.00 | (A) Metastasis |
| EGFEM1P | 3q26.2 | 1 (0.04%) | 0 (0.00%) | >10 | 1.00 | 1.00 | (A) Metastasis |
| EMILIN1 | 2p23.3 | 1 (0.04%) | 0 (0.00%) | >10 | 1.00 | 1.00 | (A) Metastasis |
| ERC1 | 12p13.33 | 1 (0.04%) | 0 (0.00%) | >10 | 1.00 | 1.00 | (A) Metastasis |
| ERP44 | 9q31.1 | 1 (0.04%) | 0 (0.00%) | >10 | 1.00 | 1.00 | (A) Metastasis |
| FAM20C | 7p22.3 | 1 (0.04%) | 0 (0.00%) | >10 | 1.00 | 1.00 | (A) Metastasis |
| FKBP5 | 6p21.31 | 1 (0.04%) | 0 (0.00%) | >10 | 1.00 | 1.00 | (A) Metastasis |
| FRMD5 | 15q15.3 | 1 (0.04%) | 0 (0.00%) | >10 | 1.00 | 1.00 | (A) Metastasis |
| FSIP1 | 15q14 | 1 (0.04%) | 0 (0.00%) | >10 | 1.00 | 1.00 | (A) Metastasis |
| FTH1 | 11q12.3 | 1 (0.04%) | 0 (0.00%) | >10 | 1.00 | 1.00 | (A) Metastasis |
| GABRG3 | 15q12 | 1 (0.04%) | 0 (0.00%) | >10 | 1.00 | 1.00 | (A) Metastasis |
| GARNL3 | 9q33.3 | 1 (0.04%) | 0 (0.00%) | >10 | 1.00 | 1.00 | (A) Metastasis |
| GKAP1 | 9q21.32 | 1 (0.04%) | 0 (0.00%) | >10 | 1.00 | 1.00 | (A) Metastasis |
| GOLT1A | 1q32.1 | 1 (0.04%) | 0 (0.00%) | >10 | 1.00 | 1.00 | (A) Metastasis |
| GSAP | 7q11.23 | 1 (0.04%) | 0 (0.00%) | >10 | 1.00 | 1.00 | (A) Metastasis |
| HIST1H2BB | 6p22.2 | 1 (0.04%) | 0 (0.00%) | >10 | 1.00 | 1.00 | (A) Metastasis |
| HNRNPA2B1 | 7p15.2 | 1 (0.04%) | 0 (0.00%) | >10 | 1.00 | 1.00 | (A) Metastasis |
| HPGD | 4q34.1 | 1 (0.04%) | 0 (0.00%) | >10 | 1.00 | 1.00 | (A) Metastasis |
| INTS4 | 11q14.1 | 1 (0.04%) | 0 (0.00%) | >10 | 1.00 | 1.00 | (A) Metastasis |
| KDF1 | 1p36.11 | 1 (0.04%) | 0 (0.00%) | >10 | 1.00 | 1.00 | (A) Metastasis |
| KITLG | 12q21.32 | 1 (0.04%) | 0 (0.00%) | >10 | 1.00 | 1.00 | (A) Metastasis |
| LINC02145 | 5p15.31 | 1 (0.04%) | 0 (0.00%) | >10 | 1.00 | 1.00 | (A) Metastasis |
| LRBA | 4q31.3 | 1 (0.04%) | 0 (0.00%) | >10 | 1.00 | 1.00 | (A) Metastasis |
| LRRC20 | 10q22.1 | 1 (0.04%) | 0 (0.00%) | >10 | 1.00 | 1.00 | (A) Metastasis |
| LRRK1 | 15q26.3 | 1 (0.04%) | 0 (0.00%) | >10 | 1.00 | 1.00 | (A) Metastasis |
| MAD1L1 | 7p22.3 | 1 (0.04%) | 0 (0.00%) | >10 | 1.00 | 1.00 | (A) Metastasis |
| MAN2A2 | 15q26.1 | 1 (0.04%) | 0 (0.00%) | >10 | 1.00 | 1.00 | (A) Metastasis |
| MARVELD3 | 16q22.2 | 1 (0.04%) | 0 (0.00%) | >10 | 1.00 | 1.00 | (A) Metastasis |
| MEIOC | 17q21.31 | 1 (0.04%) | 0 (0.00%) | >10 | 1.00 | 1.00 | (A) Metastasis |
| MIR-1273E/1273E | | 1 (0.04%) | 0 (0.00%) | >10 | 1.00 | 1.00 | (A) Metastasis |
| MSRB3 | 12q14.3 | 1 (0.04%) | 0 (0.00%) | >10 | 1.00 | 1.00 | (A) Metastasis |
| MTHFD1L | 6q25.1 | 1 (0.04%) | 0 (0.00%) | >10 | 1.00 | 1.00 | (A) Metastasis |
| MTMR3 | 22q12.2 | 1 (0.04%) | 0 (0.00%) | >10 | 1.00 | 1.00 | (A) Metastasis |
| MTREX | 5q11.2 | 1 (0.04%) | 0 (0.00%) | >10 | 1.00 | 1.00 | (A) Metastasis |
| MX1 | 21q22.3 | 1 (0.04%) | 0 (0.00%) | >10 | 1.00 | 1.00 | (A) Metastasis |
| MYT1 | 20q13.33 | 1 (0.04%) | 0 (0.00%) | >10 | 1.00 | 1.00 | (A) Metastasis |
| NBPF10 | 1q21.1 | 1 (0.04%) | 0 (0.00%) | >10 | 1.00 | 1.00 | (A) Metastasis |
| NFAT5 | 16q22.1 | 1 (0.04%) | 0 (0.00%) | >10 | 1.00 | 1.00 | (A) Metastasis |
| NPAS1 | 19q13.32 | 1 (0.04%) | 0 (0.00%) | >10 | 1.00 | 1.00 | (A) Metastasis |
| OPHN1 | Xq12 | 1 (0.04%) | 0 (0.00%) | >10 | 1.00 | 1.00 | (A) Metastasis |
| PACRG | 6q26 | 1 (0.04%) | 0 (0.00%) | >10 | 1.00 | 1.00 | (A) Metastasis |
| PAGE1 | Xp11.23 | 1 (0.04%) | 0 (0.00%) | >10 | 1.00 | 1.00 | (A) Metastasis |
| PARP8 | 5q11.1 | 1 (0.04%) | 0 (0.00%) | >10 | 1.00 | 1.00 | (A) Metastasis |
| PATJ | 1p31.3 | 1 (0.04%) | 0 (0.00%) | >10 | 1.00 | 1.00 | (A) Metastasis |
| PAWR | 12q21.2 | 1 (0.04%) | 0 (0.00%) | >10 | 1.00 | 1.00 | (A) Metastasis |
| PDE1C | 7p14.3 | 1 (0.04%) | 0 (0.00%) | >10 | 1.00 | 1.00 | (A) Metastasis |
| PEAR1 | 1q23.1 | 1 (0.04%) | 0 (0.00%) | >10 | 1.00 | 1.00 | (A) Metastasis |
| PKD1 | 16p13.3 | 1 (0.04%) | 0 (0.00%) | >10 | 1.00 | 1.00 | (A) Metastasis |
| PLAT | 8p11.21 | 1 (0.04%) | 0 (0.00%) | >10 | 1.00 | 1.00 | (A) Metastasis |
| POLN | 4p16.3 | 1 (0.04%) | 0 (0.00%) | >10 | 1.00 | 1.00 | (A) Metastasis |
| PRDM15 | 21q22.3 | 1 (0.04%) | 0 (0.00%) | >10 | 1.00 | 1.00 | (A) Metastasis |
| PSMA7 | 20q13.33 | 1 (0.04%) | 0 (0.00%) | >10 | 1.00 | 1.00 | (A) Metastasis |
| PTPRM | 18p11.23 | 1 (0.04%) | 0 (0.00%) | >10 | 1.00 | 1.00 | (A) Metastasis |
| PTPRN2 | 7q36.3 | 1 (0.04%) | 0 (0.00%) | >10 | 1.00 | 1.00 | (A) Metastasis |
| PVT1 | 8q24.21 | 1 (0.04%) | 0 (0.00%) | >10 | 1.00 | 1.00 | (A) Metastasis |
| RBMXL1 | 1p22.2 | 1 (0.04%) | 0 (0.00%) | >10 | 1.00 | 1.00 | (A) Metastasis |
| RBX1 | 22q13.2 | 1 (0.04%) | 0 (0.00%) | >10 | 1.00 | 1.00 | (A) Metastasis |
| RCAN1 | 21q22.12 | 1 (0.04%) | 0 (0.00%) | >10 | 1.00 | 1.00 | (A) Metastasis |
| RGPD2 | 2p11.2 | 1 (0.04%) | 0 (0.00%) | >10 | 1.00 | 1.00 | (A) Metastasis |
| RGS7 | 1q43\|1q23.1 | 1 (0.04%) | 0 (0.00%) | >10 | 1.00 | 1.00 | (A) Metastasis |
| RIMBP2 | 12q24.33 | 1 (0.04%) | 0 (0.00%) | >10 | 1.00 | 1.00 | (A) Metastasis |
| RPS6KC1 | 1q32.3 | 1 (0.04%) | 0 (0.00%) | >10 | 1.00 | 1.00 | (A) Metastasis |
| RSPH9 | 6p21.1 | 1 (0.04%) | 0 (0.00%) | >10 | 1.00 | 1.00 | (A) Metastasis |
| SAMD8 | 10q22.2 | 1 (0.04%) | 0 (0.00%) | >10 | 1.00 | 1.00 | (A) Metastasis |
| SCFD2 | 4q12 | 1 (0.04%) | 0 (0.00%) | >10 | 1.00 | 1.00 | (A) Metastasis |
| SDK1 | 7p22.2 | 1 (0.04%) | 0 (0.00%) | >10 | 1.00 | 1.00 | (A) Metastasis |
| SERAC1 | 6q25.3 | 1 (0.04%) | 0 (0.00%) | >10 | 1.00 | 1.00 | (A) Metastasis |
| SFRP1 | 8p11.21 | 1 (0.04%) | 0 (0.00%) | >10 | 1.00 | 1.00 | (A) Metastasis |
| SIMC1 | 5q35.2 | 1 (0.04%) | 0 (0.00%) | >10 | 1.00 | 1.00 | (A) Metastasis |
| SLC12A3 | 16q13 | 1 (0.04%) | 0 (0.00%) | >10 | 1.00 | 1.00 | (A) Metastasis |
| SLC16A7 | 12q14.1 | 1 (0.04%) | 0 (0.00%) | >10 | 1.00 | 1.00 | (A) Metastasis |
| SLC6A4 | 17q11.2 | 1 (0.04%) | 0 (0.00%) | >10 | 1.00 | 1.00 | (A) Metastasis |
| SLC9A1 | 1p36.11 | 1 (0.04%) | 0 (0.00%) | >10 | 1.00 | 1.00 | (A) Metastasis |
| SLN | 11q22.3 | 1 (0.04%) | 0 (0.00%) | >10 | 1.00 | 1.00 | (A) Metastasis |
| SNX8 | 7p22.3 | 1 (0.04%) | 0 (0.00%) | >10 | 1.00 | 1.00 | (A) Metastasis |
| SPATA6L | 9p24.2-p24.1 | 1 (0.04%) | 0 (0.00%) | >10 | 1.00 | 1.00 | (A) Metastasis |
| SPNS2 | 17p13.2 | 1 (0.04%) | 0 (0.00%) | >10 | 1.00 | 1.00 | (A) Metastasis |
| SRBD1 | 2p21 | 1 (0.04%) | 0 (0.00%) | >10 | 1.00 | 1.00 | (A) Metastasis |
| STX8 | 17p13.1 | 1 (0.04%) | 0 (0.00%) | >10 | 1.00 | 1.00 | (A) Metastasis |
| SUGCT | 7p14.1 | 1 (0.04%) | 0 (0.00%) | >10 | 1.00 | 1.00 | (A) Metastasis |
| SUGP1 | 19p13.11 | 1 (0.04%) | 0 (0.00%) | >10 | 1.00 | 1.00 | (A) Metastasis |
| SYNDIG1 | 20p11.21 | 1 (0.04%) | 0 (0.00%) | >10 | 1.00 | 1.00 | (A) Metastasis |
| SYS1 | 20q13.12 | 1 (0.04%) | 0 (0.00%) | >10 | 1.00 | 1.00 | (A) Metastasis |
| TACC2 | 10q26.13 | 1 (0.04%) | 0 (0.00%) | >10 | 1.00 | 1.00 | (A) Metastasis |
| TAF4 | 20q13.33 | 1 (0.04%) | 0 (0.00%) | >10 | 1.00 | 1.00 | (A) Metastasis |
| TAMM41 | 3p25.2 | 1 (0.04%) | 0 (0.00%) | >10 | 1.00 | 1.00 | (A) Metastasis |
| TFAP2D | 6p12.3 | 1 (0.04%) | 0 (0.00%) | >10 | 1.00 | 1.00 | (A) Metastasis |
| THBS2 | 6q27 | 1 (0.04%) | 0 (0.00%) | >10 | 1.00 | 1.00 | (A) Metastasis |
| THEMIS2 | 1p35.3 | 1 (0.04%) | 0 (0.00%) | >10 | 1.00 | 1.00 | (A) Metastasis |
| TIGD7 | 16p13.3 | 1 (0.04%) | 0 (0.00%) | >10 | 1.00 | 1.00 | (A) Metastasis |
| TNFRSF13B | 17p11.2 | 1 (0.04%) | 0 (0.00%) | >10 | 1.00 | 1.00 | (A) Metastasis |
| TNFRSF21 | 6p12.3 | 1 (0.04%) | 0 (0.00%) | >10 | 1.00 | 1.00 | (A) Metastasis |
| TRAJ9 | 14q11.2 | 1 (0.04%) | 0 (0.00%) | >10 | 1.00 | 1.00 | (A) Metastasis |
| TREX1 | 3p21.31 | 1 (0.04%) | 0 (0.00%) | >10 | 1.00 | 1.00 | (A) Metastasis |
| TSEN2 | 3p25.2 | 1 (0.04%) | 0 (0.00%) | >10 | 1.00 | 1.00 | (A) Metastasis |
| TSPAN5 | 4q23 | 1 (0.04%) | 0 (0.00%) | >10 | 1.00 | 1.00 | (A) Metastasis |
| TTC25 | 17q21.2 | 1 (0.04%) | 0 (0.00%) | >10 | 1.00 | 1.00 | (A) Metastasis |
| TTC6 | 14q21.1 | 1 (0.04%) | 0 (0.00%) | >10 | 1.00 | 1.00 | (A) Metastasis |
| TTLL9 | 20q11.21 | 1 (0.04%) | 0 (0.00%) | >10 | 1.00 | 1.00 | (A) Metastasis |
| UCKL1 | 20q13.33 | 1 (0.04%) | 0 (0.00%) | >10 | 1.00 | 1.00 | (A) Metastasis |
| UGT1A6 | 2q37.1 | 1 (0.04%) | 0 (0.00%) | >10 | 1.00 | 1.00 | (A) Metastasis |
| UPP2 | 2q24.1 | 1 (0.04%) | 0 (0.00%) | >10 | 1.00 | 1.00 | (A) Metastasis |
| URAD | 13q12.2 | 1 (0.04%) | 0 (0.00%) | >10 | 1.00 | 1.00 | (A) Metastasis |
| USH2A | 1q41 | 1 (0.04%) | 0 (0.00%) | >10 | 1.00 | 1.00 | (A) Metastasis |
| YBX3 | 12p13.2 | 1 (0.04%) | 0 (0.00%) | >10 | 1.00 | 1.00 | (A) Metastasis |
| ZFAT | 8q24.22 | 1 (0.04%) | 0 (0.00%) | >10 | 1.00 | 1.00 | (A) Metastasis |
| ZFC3H1 | 12q21.1 | 1 (0.04%) | 0 (0.00%) | >10 | 1.00 | 1.00 | (A) Metastasis |
| ZHX3 | 20q12 | 1 (0.04%) | 0 (0.00%) | >10 | 1.00 | 1.00 | (A) Metastasis |
| ZNF407 | 18q22.3 | 1 (0.04%) | 0 (0.00%) | >10 | 1.00 | 1.00 | (A) Metastasis |
| ZNF503 | 10q22.2 | 1 (0.04%) | 0 (0.00%) | >10 | 1.00 | 1.00 | (A) Metastasis |
| ZNF532 | 18q21.32 | 1 (0.04%) | 0 (0.00%) | >10 | 1.00 | 1.00 | (A) Metastasis |
| ZNF714 | 19p12 | 1 (0.04%) | 0 (0.00%) | >10 | 1.00 | 1.00 | (A) Metastasis |
| ZSCAN20 | 1p35.1 | 1 (0.04%) | 0 (0.00%) | >10 | 1.00 | 1.00 | (A) Metastasis |
| S1PR2 | 19p13.2 | 0 (0.00%) | 1 (100.00%) | <-10 | 3.512e-4 | 0.0305 | (B) Primary |
| TRBC2 | 7q34 | 0 (0.00%) | 5 (0.18%) | <-10 | 0.0624 | 0.695 | (B) Primary |
| AFDN | 6q27 | 0 (0.00%) | 2 (40.00%) | <-10 | 0.182 | 1.00 | (B) Primary |
| DCC | 18q21.2 | 0 (0.00%) | 2 (40.00%) | <-10 | 0.182 | 1.00 | (B) Primary |
| TBX22 | Xq21.1 | 0 (0.00%) | 2 (40.00%) | <-10 | 0.182 | 1.00 | (B) Primary |
| TNK2 | 3q29 | 0 (0.00%) | 2 (40.00%) | <-10 | 0.182 | 1.00 | (B) Primary |
| PHLPP2 | 16q22.2 | 0 (0.00%) | 2 (10.53%) | <-10 | 0.231 | 1.00 | (B) Primary |
| BCR | 22q11.23 | 0 (0.00%) | 3 (11.11%) | <-10 | 0.236 | 1.00 | (B) Primary |
| C8ORF34 | 8q13.2 | 0 (0.00%) | 1 (50.00%) | <-10 | 0.333 | 1.00 | (B) Primary |
| HLA-DMA | 6p21.32 | 0 (0.00%) | 1 (50.00%) | <-10 | 0.333 | 1.00 | (B) Primary |
| HNF1B | 17q12 | 0 (0.00%) | 1 (50.00%) | <-10 | 0.333 | 1.00 | (B) Primary |
| MYH11 | 16p13.11 | 0 (0.00%) | 1 (14.29%) | <-10 | 0.389 | 1.00 | (B) Primary |
| ASPSCR1 | 17q25.3 | 0 (0.00%) | 1 (14.29%) | <-10 | 0.412 | 1.00 | (B) Primary |
| HIF1A | 14q23.2 | 0 (0.00%) | 1 (14.29%) | <-10 | 0.412 | 1.00 | (B) Primary |
| TOE1 | 1p34.1 | 0 (0.00%) | 1 (0.13%) | <-10 | 0.424 | 1.00 | (B) Primary |
| SLX1A | 16p11.2 | 0 (0.00%) | 1 (0.26%) | <-10 | 0.438 | 1.00 | (B) Primary |
| HIST1H1E | 6p22.2 | 0 (0.00%) | 1 (12.50%) | <-10 | 0.444 | 1.00 | (B) Primary |
| AFF1 | 4q21.3-q22.1 | 0 (0.00%) | 1 (20.00%) | <-10 | 0.455 | 1.00 | (B) Primary |
| BCL11A | 2p16.1 | 0 (0.00%) | 1 (20.00%) | <-10 | 0.455 | 1.00 | (B) Primary |
| IKBKB | 8p11.21 | 0 (0.00%) | 1 (20.00%) | <-10 | 0.455 | 1.00 | (B) Primary |
| ITGA9 | 3p22.2 | 0 (0.00%) | 1 (20.00%) | <-10 | 0.455 | 1.00 | (B) Primary |
| LIFR | 5p13.1 | 0 (0.00%) | 1 (20.00%) | <-10 | 0.455 | 1.00 | (B) Primary |
| LTF | 3p21.31 | 0 (0.00%) | 1 (20.00%) | <-10 | 0.455 | 1.00 | (B) Primary |
| MYH9 | 22q12.3 | 0 (0.00%) | 1 (20.00%) | <-10 | 0.455 | 1.00 | (B) Primary |
| PER1 | 17p13.1 | 0 (0.00%) | 1 (20.00%) | <-10 | 0.455 | 1.00 | (B) Primary |
| POU5F1 | 6p21.33 | 0 (0.00%) | 1 (20.00%) | <-10 | 0.455 | 1.00 | (B) Primary |
| RALGDS | 9q34.13-q34.2 | 0 (0.00%) | 1 (20.00%) | <-10 | 0.455 | 1.00 | (B) Primary |
| TGFB1 | 19q13.2 | 0 (0.00%) | 1 (20.00%) | <-10 | 0.455 | 1.00 | (B) Primary |
| TLR2 | 4q31.3 | 0 (0.00%) | 1 (20.00%) | <-10 | 0.455 | 1.00 | (B) Primary |
| TRIP11 | 14q32.12 | 0 (0.00%) | 1 (20.00%) | <-10 | 0.455 | 1.00 | (B) Primary |
| UMODL1 | 21q22.3 | 0 (0.00%) | 1 (20.00%) | <-10 | 0.455 | 1.00 | (B) Primary |
| ZSWIM4 | 19p13.13-p13.12 | 0 (0.00%) | 1 (20.00%) | <-10 | 0.455 | 1.00 | (B) Primary |
| EPHB6 | 7q34 | 0 (0.00%) | 2 (11.76%) | <-10 | 0.485 | 1.00 | (B) Primary |
| HSP90AA1 | 14q32.31 | 0 (0.00%) | 2 (9.52%) | <-10 | 0.488 | 1.00 | (B) Primary |
| LTK | 15q15.1 | 0 (0.00%) | 2 (9.52%) | <-10 | 0.488 | 1.00 | (B) Primary |
| DLG1 | 3q29 | 0 (0.00%) | 2 (0.07%) | <-10 | 0.500 | 1.00 | (B) Primary |
| SKIL | 3q26.2 | 0 (0.00%) | 2 (0.07%) | <-10 | 0.500 | 1.00 | (B) Primary |
| RHPN2 | 19q13.11 | 0 (0.00%) | 6 (4.44%) | <-10 | 0.591 | 1.00 | (B) Primary |
| RBL2 | 16q12.2 | 0 (0.00%) | 3 (2.22%) | <-10 | 1.00 | 1.00 | (B) Primary |
| GOPC | 6q22.1 | 0 (0.00%) | 1 (0.04%) | <-10 | 1.00 | 1.00 | (B) Primary |
| LAMB4 | 7q31.1 | 0 (0.00%) | 1 (0.04%) | <-10 | 1.00 | 1.00 | (B) Primary |
| CD58 | 1p13.1 | 0 (0.00%) | 1 (0.74%) | <-10 | 1.00 | 1.00 | (B) Primary |
| PRPF8 | 17p13.3 | 0 (0.00%) | 4 (2.96%) | <-10 | 1.00 | 1.00 | (B) Primary |
| LINC00894 | Xq28 | 0 (0.00%) | 1 (0.97%) | <-10 | 1.00 | 1.00 | (B) Primary |
| NPRL2 | 3p21.31 | 0 (0.00%) | 1 (0.97%) | <-10 | 1.00 | 1.00 | (B) Primary |
| INSIG1 | 7q36.3 | 0 (0.00%) | 2 (1.94%) | <-10 | 1.00 | 1.00 | (B) Primary |
| KCNIP1 | 5q35.1 | 0 (0.00%) | 2 (1.94%) | <-10 | 1.00 | 1.00 | (B) Primary |
| PRPF40B | 12q13.12 | 0 (0.00%) | 2 (1.48%) | <-10 | 1.00 | 1.00 | (B) Primary |
| CDH5 | 16q21 | 0 (0.00%) | 1 (5.88%) | <-10 | 1.00 | 1.00 | (B) Primary |
| GRM3 | 7q21.11-q21.12 | 0 (0.00%) | 1 (5.26%) | <-10 | 1.00 | 1.00 | (B) Primary |
| IRF2 | 4q35.1 | 0 (0.00%) | 1 (5.26%) | <-10 | 1.00 | 1.00 | (B) Primary |
| CCN6 | 6q21 | 0 (0.00%) | 1 (4.76%) | <-10 | 1.00 | 1.00 | (B) Primary |
| CUL4A | 13q34 | 0 (0.00%) | 1 (4.76%) | <-10 | 1.00 | 1.00 | (B) Primary |
| PTCH2 | 1p34.1 | 0 (0.00%) | 1 (7.14%) | <-10 | 1.00 | 1.00 | (B) Primary |
| CUL4B | Xq24 | 0 (0.00%) | 1 (5.88%) | <-10 | 1.00 | 1.00 | (B) Primary |
| BCL2L14 | 12p13.2 | 0 (0.00%) | 1 (100.00%) | <-10 | 1.00 | 1.00 | (B) Primary |
| CDK1 | 10q21.2 | 0 (0.00%) | 1 (0.74%) | <-10 | 1.00 | 1.00 | (B) Primary |
| CRTC2 | 1q21.3 | 0 (0.00%) | 1 (0.74%) | <-10 | 1.00 | 1.00 | (B) Primary |
| SPTA1 | 1q23.1 | 0 (0.00%) | 1 (7.69%) | <-10 | 1.00 | 1.00 | (B) Primary |
| PDK1 | 2q31.1 | 0 (0.00%) | 1 (3.70%) | <-10 | 1.00 | 1.00 | (B) Primary |
| STAT4 | 2q32.2-q32.3 | 0 (0.00%) | 1 (4.35%) | <-10 | 1.00 | 1.00 | (B) Primary |
| ATP1B2 | 17p13.1 | 0 (0.00%) | 1 (0.04%) | <-10 | 1.00 | 1.00 | (B) Primary |
| DNAH2 | 17p13.1 | 0 (0.00%) | 1 (0.04%) | <-10 | 1.00 | 1.00 | (B) Primary |
| PEX26 | 22q11.21 | 0 (0.00%) | 1 (0.04%) | <-10 | 1.00 | 1.00 | (B) Primary |
| ADAM8 | 10q26.3 | 0 (0.00%) | 1 (0.04%) | <-10 | 1.00 | 1.00 | (B) Primary |
| AIMP1 | 4q24 | 0 (0.00%) | 1 (0.04%) | <-10 | 1.00 | 1.00 | (B) Primary |
| AK9 | 6q21 | 0 (0.00%) | 1 (0.04%) | <-10 | 1.00 | 1.00 | (B) Primary |
| ALG9 | 11q23.1 | 0 (0.00%) | 1 (0.04%) | <-10 | 1.00 | 1.00 | (B) Primary |
| ANKFN1 | 17q22 | 0 (0.00%) | 1 (0.04%) | <-10 | 1.00 | 1.00 | (B) Primary |
| ANKS3 | 16p13.3 | 0 (0.00%) | 1 (0.04%) | <-10 | 1.00 | 1.00 | (B) Primary |
| ATAD1 | 10q23.31 | 0 (0.00%) | 1 (0.04%) | <-10 | 1.00 | 1.00 | (B) Primary |
| ATP8A2 | 13q12.13 | 0 (0.00%) | 1 (0.04%) | <-10 | 1.00 | 1.00 | (B) Primary |
| BCAN | 1q23.1 | 0 (0.00%) | 1 (0.04%) | <-10 | 1.00 | 1.00 | (B) Primary |
| BICC1 | 10q21.1 | 0 (0.00%) | 1 (0.04%) | <-10 | 1.00 | 1.00 | (B) Primary |
| C10ORF90 | 10q26.2 | 0 (0.00%) | 1 (0.04%) | <-10 | 1.00 | 1.00 | (B) Primary |
| CALM3 | 19q13.32 | 0 (0.00%) | 1 (0.04%) | <-10 | 1.00 | 1.00 | (B) Primary |
| CAMK2A | 5q32 | 0 (0.00%) | 1 (0.04%) | <-10 | 1.00 | 1.00 | (B) Primary |
| CCDC9 | 19q13.32 | 0 (0.00%) | 1 (0.04%) | <-10 | 1.00 | 1.00 | (B) Primary |
| CD55 | 1q32.2 | 0 (0.00%) | 1 (0.04%) | <-10 | 1.00 | 1.00 | (B) Primary |
| CHML | 1q43 | 0 (0.00%) | 1 (0.04%) | <-10 | 1.00 | 1.00 | (B) Primary |
| CLMN | 14q32.13 | 0 (0.00%) | 1 (0.04%) | <-10 | 1.00 | 1.00 | (B) Primary |
| CNOT2 | 12q15 | 0 (0.00%) | 1 (0.04%) | <-10 | 1.00 | 1.00 | (B) Primary |
| COL5A3 | 19p13.2 | 0 (0.00%) | 1 (0.04%) | <-10 | 1.00 | 1.00 | (B) Primary |
| CTDSP2 | 12q14.1 | 0 (0.00%) | 1 (0.04%) | <-10 | 1.00 | 1.00 | (B) Primary |
| CYP2S1 | 19q13.2 | 0 (0.00%) | 1 (0.04%) | <-10 | 1.00 | 1.00 | (B) Primary |
| DCLK1 | 13q13.3 | 0 (0.00%) | 1 (0.04%) | <-10 | 1.00 | 1.00 | (B) Primary |
| DIAPH3 | 13q21.2 | 0 (0.00%) | 1 (0.04%) | <-10 | 1.00 | 1.00 | (B) Primary |
| DNAJC24 | 11p13 | 0 (0.00%) | 1 (0.04%) | <-10 | 1.00 | 1.00 | (B) Primary |
| DNER | 2q36.3 | 0 (0.00%) | 1 (0.04%) | <-10 | 1.00 | 1.00 | (B) Primary |
| DOCK2 | 5q35.1 | 0 (0.00%) | 1 (0.04%) | <-10 | 1.00 | 1.00 | (B) Primary |
| DSCAML1 | 11q23.3 | 0 (0.00%) | 1 (0.04%) | <-10 | 1.00 | 1.00 | (B) Primary |
| DUX4 | 4q35.2 | 0 (0.00%) | 1 (0.04%) | <-10 | 1.00 | 1.00 | (B) Primary |
| EIF2S1 | 14q23.3 | 0 (0.00%) | 1 (0.04%) | <-10 | 1.00 | 1.00 | (B) Primary |
| EYS | 6q12 | 0 (0.00%) | 1 (0.04%) | <-10 | 1.00 | 1.00 | (B) Primary |
| FBXL20 | 17q12 | 0 (0.00%) | 1 (0.04%) | <-10 | 1.00 | 1.00 | (B) Primary |
| FBXO42 | 1p36.13 | 0 (0.00%) | 1 (0.04%) | <-10 | 1.00 | 1.00 | (B) Primary |
| GMDS | 6p25.3 | 0 (0.00%) | 1 (0.04%) | <-10 | 1.00 | 1.00 | (B) Primary |
| GMEB1 | 1p35.3 | 0 (0.00%) | 1 (0.04%) | <-10 | 1.00 | 1.00 | (B) Primary |
| GOT1L1 | 8p11.23 | 0 (0.00%) | 1 (0.04%) | <-10 | 1.00 | 1.00 | (B) Primary |
| GPATCH8 | 17q21.31 | 0 (0.00%) | 1 (0.04%) | <-10 | 1.00 | 1.00 | (B) Primary |
| GPR158 | 10p12.1 | 0 (0.00%) | 1 (0.04%) | <-10 | 1.00 | 1.00 | (B) Primary |
| GRIK4 | 11q23.3 | 0 (0.00%) | 1 (0.04%) | <-10 | 1.00 | 1.00 | (B) Primary |
| GSDMB | 17q21.1 | 0 (0.00%) | 1 (0.04%) | <-10 | 1.00 | 1.00 | (B) Primary |
| GYS2 | 12p12.1 | 0 (0.00%) | 1 (0.04%) | <-10 | 1.00 | 1.00 | (B) Primary |
| INSRR | 1q23.1 | 0 (0.00%) | 1 (0.04%) | <-10 | 1.00 | 1.00 | (B) Primary |
| KRT222 | 17q21.2 | 0 (0.00%) | 1 (0.04%) | <-10 | 1.00 | 1.00 | (B) Primary |
| KRT25 | 17q21.2 | 0 (0.00%) | 1 (0.04%) | <-10 | 1.00 | 1.00 | (B) Primary |
| LGR6 | 1q32.1 | 0 (0.00%) | 1 (0.04%) | <-10 | 1.00 | 1.00 | (B) Primary |
| LIMK1 | 7q11.23 | 0 (0.00%) | 1 (0.04%) | <-10 | 1.00 | 1.00 | (B) Primary |
| LOC105374464 | 2p22.2 | 0 (0.00%) | 1 (0.04%) | <-10 | 1.00 | 1.00 | (B) Primary |
| MAOA | Xp11.3 | 0 (0.00%) | 1 (0.04%) | <-10 | 1.00 | 1.00 | (B) Primary |
| MBIP | 14q13.3 | 0 (0.00%) | 1 (0.04%) | <-10 | 1.00 | 1.00 | (B) Primary |
| MLST8 | 16p13.3 | 0 (0.00%) | 1 (0.04%) | <-10 | 1.00 | 1.00 | (B) Primary |
| NR1H2 | 19q13.33 | 0 (0.00%) | 1 (0.04%) | <-10 | 1.00 | 1.00 | (B) Primary |
| NRF1 | 7q32.2 | 0 (0.00%) | 1 (0.04%) | <-10 | 1.00 | 1.00 | (B) Primary |
| OXR1 | 8q23.1 | 0 (0.00%) | 1 (0.04%) | <-10 | 1.00 | 1.00 | (B) Primary |
| PARG | 10q11.23 | 0 (0.00%) | 1 (0.04%) | <-10 | 1.00 | 1.00 | (B) Primary |
| PEPD | 19q13.11 | 0 (0.00%) | 1 (0.04%) | <-10 | 1.00 | 1.00 | (B) Primary |
| PLGRKT | 9p24.1 | 0 (0.00%) | 1 (0.04%) | <-10 | 1.00 | 1.00 | (B) Primary |
| PPOX | 1q23.3 | 0 (0.00%) | 1 (0.04%) | <-10 | 1.00 | 1.00 | (B) Primary |
| PXN | 12q24.23 | 0 (0.00%) | 1 (0.04%) | <-10 | 1.00 | 1.00 | (B) Primary |
| RAB3GAP2 | 1q41 | 0 (0.00%) | 1 (0.04%) | <-10 | 1.00 | 1.00 | (B) Primary |
| RAVER1 | 19p13.2 | 0 (0.00%) | 1 (0.04%) | <-10 | 1.00 | 1.00 | (B) Primary |
| RUFY2 | 10q21.3 | 0 (0.00%) | 1 (0.04%) | <-10 | 1.00 | 1.00 | (B) Primary |
| SCN4A | 17q23.3 | 0 (0.00%) | 1 (0.04%) | <-10 | 1.00 | 1.00 | (B) Primary |
| SCN8A | 12q13.13 | 0 (0.00%) | 1 (0.04%) | <-10 | 1.00 | 1.00 | (B) Primary |
| SEC16A | 9q34.3 | 0 (0.00%) | 1 (0.04%) | <-10 | 1.00 | 1.00 | (B) Primary |
| SERPINA12 | 14q32.13 | 0 (0.00%) | 1 (0.04%) | <-10 | 1.00 | 1.00 | (B) Primary |
| SLC13A2 | 17q11.2 | 0 (0.00%) | 1 (0.04%) | <-10 | 1.00 | 1.00 | (B) Primary |
| SRRM1 | 1p36.11 | 0 (0.00%) | 1 (0.04%) | <-10 | 1.00 | 1.00 | (B) Primary |
| ST8SIA4 | 5q21.1 | 0 (0.00%) | 1 (0.04%) | <-10 | 1.00 | 1.00 | (B) Primary |
| STX17 | 9q31.1 | 0 (0.00%) | 1 (0.04%) | <-10 | 1.00 | 1.00 | (B) Primary |
| SYN1 | Xp11.3-p11.23 | 0 (0.00%) | 1 (0.04%) | <-10 | 1.00 | 1.00 | (B) Primary |
| TBK1 | 12q14.2 | 0 (0.00%) | 1 (0.04%) | <-10 | 1.00 | 1.00 | (B) Primary |
| TIMM17A | 1q32.1 | 0 (0.00%) | 1 (0.04%) | <-10 | 1.00 | 1.00 | (B) Primary |
| TIMM23B | 10q11.23 | 0 (0.00%) | 1 (0.04%) | <-10 | 1.00 | 1.00 | (B) Primary |
| TMEM141 | 9q34.3 | 0 (0.00%) | 1 (0.04%) | <-10 | 1.00 | 1.00 | (B) Primary |
| TMEM189-UBE2V1 | 20q13.13 | 0 (0.00%) | 1 (0.04%) | <-10 | 1.00 | 1.00 | (B) Primary |
| TMEM72-AS1 | 10q11.21 | 0 (0.00%) | 1 (0.04%) | <-10 | 1.00 | 1.00 | (B) Primary |
| TRAJ20 | 14q11.2 | 0 (0.00%) | 1 (0.04%) | <-10 | 1.00 | 1.00 | (B) Primary |
| TRAJ46 | 14q11.2 | 0 (0.00%) | 1 (0.04%) | <-10 | 1.00 | 1.00 | (B) Primary |
| TRAJ53 | 14q11.2 | 0 (0.00%) | 1 (0.04%) | <-10 | 1.00 | 1.00 | (B) Primary |
| TRAJ54 | 14q11.2 | 0 (0.00%) | 1 (0.04%) | <-10 | 1.00 | 1.00 | (B) Primary |
| TSHZ3 | 19q12 | 0 (0.00%) | 1 (0.04%) | <-10 | 1.00 | 1.00 | (B) Primary |
| VOPP1 | 7p11.2 | 0 (0.00%) | 1 (0.04%) | <-10 | 1.00 | 1.00 | (B) Primary |
| VTI1A | 10q25.2 | 0 (0.00%) | 1 (0.04%) | <-10 | 1.00 | 1.00 | (B) Primary |
| WASF2 | 1p36.11 | 0 (0.00%) | 1 (0.04%) | <-10 | 1.00 | 1.00 | (B) Primary |
| WIPF2 | 17q21.2 | 0 (0.00%) | 1 (0.04%) | <-10 | 1.00 | 1.00 | (B) Primary |
| ZC3HAV1 | 7q34 | 0 (0.00%) | 1 (0.04%) | <-10 | 1.00 | 1.00 | (B) Primary |
| ZFP14 | 19q13.12 | 0 (0.00%) | 1 (0.04%) | <-10 | 1.00 | 1.00 | (B) Primary |
| ZNF777 | 7q36.1 | 0 (0.00%) | 1 (0.04%) | <-10 | 1.00 | 1.00 | (B) Primary |
